# Association of Covert Cerebrovascular Disease Identified Using Natural Language Processing and Future Dementia

**DOI:** 10.1101/2022.02.09.22270682

**Authors:** David M Kent, Lester Y Leung, Yichen Zhou, Patrick H Luetmer, David F Kallmes, Jason Nelson, Sunyang Fu, Chengyi Zheng, Hongfang Liu, Wansu Chen

## Abstract

**Objective:** To estimate the risk of dementia associated with incidentally-discovered covert cerebrovascular disease (CCD), including both covert brain infarction (CBI) and white matter disease (WMD).

**Patients and Methods:** We included individuals aged ≥ 50 years enrolled in the Kaiser Permanente Southern California health system receiving a head CT or MRI for a non-stroke indication from January 1, 2009 and December 31, 2019, without prior ischemic stroke, transient ischemic attack, hemipelegia, hemiparesis, dementia/Alzheimer’s disease or a visit reason / scan indication suggestive of cognitive decline. Using natural language processing (NLP), we identified the presence of CBI and WMD on the neuroimage report; WMD was characterized as mild, moderate, severe, or undetermined.

**Results:** Among 241,050 qualified individuals, NLP identified 69,931 (29.0%) with WMD and 11,328 (4.7%) with CBI. The dementia incidence rates (per 1,000 person-years) were 23.5 (95% CI 22.90 to 24.0) for patients with WMD; 29.4 (95% CI 27.9 to 31.0) with CBI and 6.0 (5.8 to 6.2) without CCD. The effect of WMD on dementia risk was stronger in younger versus older patients and for CT-versus MRI-discovered lesions. For patients with versus without WMD on CT scan, the adjusted HR (aHR) was 2.87 (2.58 to 3.19) for those < age 70 and 1.87 (1.79 to 1.95) for those ≥ age 70. For patients with versus without WMD on MRI, the aHR for dementia risk was 2.28 (1.99 to 2.62) for patients < age 70 and 1.48 (1.32 to 1.66) for those ≥ age 70. The aHR associated with CBI was 2.02 (1.70 to 2.41) for patients age <70 and 1.22 (1.15 to 1.30) for patients age ≥70 for either modality. Dementia risk with WMD was strongly correlated with WMD severity.

**Conclusion:** Incidentally-discovered CCD is common and identifies patients at high risk of dementia, representing an opportunity for prevention.

## INTRODUCTION

Cardiovascular risk factors contribute to the development of several common forms of dementia and are a focus of efforts to prevent these diseases.^1,2^ Identifying high-risk individuals before the onset of severe cognitive decline is a central goal in dementia prevention research. One potentially appealing strategy is to identify individuals with covert cerebrovascular diseases (CCD), comprising both covert brain infarction (CBI) and white matter disease (WMD), which are known to be associated with an increased risk for dementia in cohort studies with participants undergoing screening by protocol-driven neuroimaging.^3-10^

However, building upon insights from screened cohort studies and translating their findings into real-world clinical practice has unique challenges. Findings from population-based research with protocol-driven magnetic resonance imaging (MRI) screening may not be generalizable to real-world cohorts which include only patients selected for clinically-indicated neuroimaging, where imaging may be dominated by computer tomography (CT) scans and where neuroimaging interpretation and reporting are heterogeneous and poorly standardized.^11,12^ Yet studying incidentally-discovered CCD is impeded by the poor clinical documentation of these lesions; there are no ICD-9 codes for CBI or WMD and no ICD-10 codes for CBI. Additionally, these lesions are rarely documented in a patient’s problem list even when they are reported on neuroimaging reports. Given these barriers to identifying patients with CCD, it is unsurprising that no current standard of care for the management of these conditions or evidence-based strategies for prevention of dementia following their discovery exists, despite the fact that they are commonly encountered in routine care.^13^

To facilitate identification of individuals with incidentally discovered CCD, we previously developed a Natural Language Processing (NLP) algorithm identifying these conditions with a high degree of accuracy.^14,15^ In this study, we port the NLP algorithm into Kaiser Permanente Southern California (KPSC), a large integrated health care system, to examine the prognostic significance of NLP-identified, incidentally-discovered CCD on risk of dementia.

## METHODS

### Environment

In this retrospective cohort study, we utilized health plan enrollees of KPSC, an integrated health care organization that serves 4.8 million individuals (approximately 19% of the region’s population) at 15 hospitals and 230+ medical offices, broadly representative of the residents in the region.^16^ Data was extracted, via a research data warehouse, from KPSC’s electronic health record (EHR). This system integrates all aspects of care, including inpatient, emergency department, outpatient, pharmacy, and lab services, as well as billing and claims. The study protocol was approved by Tufts Medical Center’s and KPSC’s Institutional Review Board, which waived the need for informed consent.

### Population

We included individuals age ≥ 50 years enrolled in the KPSC health system who received head neuroimaging (CT, MRI) for a non-stroke indication between 2009-2019 and who had no history of ischemic stroke or dementia/Alzheimer’s. Patients with transient ischemic attack (TIA), hemiplegia and hemiparesis and other medication conditions were also excluded to ensure a stroke- and dementia-free cohort. **Supplement A1** includes the complete ICD-9/ICD-10/CPT codes used to exclude patients. For the current analysis, we additionally excluded patients with a visit reason or scan indication suggestive of cognitive symptoms or decline (e.g. confusion, disorientation, altered mental status, or dementia evaluation).

If there were multiple neuroimaging studies, the first was considered the index scan. For neuroimaging evidence of cerebral infarction to be considered “covert,” individuals were only included in the study if they did not acquire a new ICD code for a diagnosis of cerebral infarction or dementia within 60 days after the index scan. Patients who were not actively enrolled in the KPSC health plan on the index date or not continuously enrolled in the prior 12 months were also excluded (a gap of ≤45 days was allowed).

### Identification of patients with CCD

An NLP algorithm developed at Mayo Clinic and Tufts Medical Center was applied to neuroimaging reports associated with these index scans to identify individuals with documented CBI or WMD.^14^ As described in prior work, these algorithms adopted the open-source NLP pipeline MedTagger for generic NLP processing and task-specific knowledge engineering (coding of specific words and phrases referencing CBI and WMD) to yield identification of CBI and WMD that was on par with human readers of the neuroimaging reports.^14^ After two rounds of training, this NLP algorithm ultimately achieved F-scores of 0.93 and 0.92 in CBI and WMD.

### Follow-up

Follow-up started 60 days after the index scan and ended with the earliest of the following events: disenrollment from the health plan, end of the study (December 31, 2019), death, or dementia (outcome). The 60-day window after the index scan was selected to further exclude patients with symptoms of cognitive decline that might have prompted the neuroimaging scan.

### Outcome definition

The primary outcome of this study was dementia. The occurrence of dementia was defined by the following ICD-9 diagnosis codes: 290.x, 291.2, 292.82, 294.1x, 294.2x, or 331.0. Only a single code was required if accompanied by a cognitive enhancement medication dispensation, or any two codes on different dates (with the second as the event date) if cognitive enhancement medication was not dispensed. Equivalent ICD-9 and ICD-10 codes used and qualifying cognitive enhancement medications are shown in **Supplement A2**.

### Statistical Analysis

Kaplan-Meier plots were used to present dementia-free survival in patients with and without CBI and with and without WMD, with differences assessed by the log-rank test. The overall and risk-factor stratified crude incidence rates and the 95% confidence intervals (CI) were calculated using Poisson regression and reported as per 1,000 person-years of follow-up time. We examined the crude and adjusted associations of CBI and of WMD with dementia, using Cox proportional hazards regression models. For adjusted effects, we included known cardiovascular risk factors for stroke based on prediction models in the literature, including the following covariates: age, sex, race (non-Hispanic white; Asian/Pacific Islander; African American; Hispanic; Multiple/other/unknown), diabetes, hypercholesterolemia, history of smoking, mean systolic blood pressure (averaged over prior year, excluding extreme values <70 or > 200 to avoid including measurements from periods of critical illness), atrial fibrillation, carotid disease, congestive heart failure, peripheral arterial disease, and use of antiplatelet or statin therapy.^17,18^ We also included dementia risk factors, including presence of depression, body mass index, and exercise.^19-22^

For each risk factor, the proportional hazards assumption was examined by the Schoenfeld residuals test. Interaction terms were selected based on clinical judgement. In particular, we hypothesized that the effects of WMD and CBI would vary based on imaging modality. We thus compared the effect of the presence versus the absence of CBI or WMD separately in those examined by CT and by MRI. We also anticipated that these lesions might have greater prognostic importance in younger versus older patients and so included interactions with age (< age 70 versus ≥ age 70).

### Sensitivity Analysis

Three sensitivity analyses were performed: 1) defining the outcome based on a single dementia diagnostic code [to examine the stability of results when a more sensitive outcome definition is used]; 2) excluding patients who were either on an antithrombotic at baseline or who had a clinical indication for antithrombotic therapy [to examine effects among an antithrombotic free population who might be considered appropriate for a trial testing “secondary prevention” with an antiplatelet agent]; 3) starting follow up 1-year after the index scan (instead of 60-days after the scan) [to rule out with greater certainty that the index scan was not ordered because of a suspected diagnosis of cognitive decline or dementia].

### Analysis of WMD grade

To further stratify risk of dementia, we classified patients according to WMD severity using NLP, as discussed in **Supplement A3**. Using the description in neuroradiology reports, patients with WMD were classified into three severity grades: mild, moderate, or severe. Scan reports with insufficient information on severity were classified as “undetermined.” Analyses were performed in a similar fashion to the main analysis described above. However, WMD severity was interacted with imaging modality and thus treated as 10 different classes (i.e., no WMD, mild WMD, moderate WMD, severe WMD, and undetermined WMD for CT and for MRI). The reference class for all hazards was those patients who underwent MRI imaging and were found to be free of WMD (i.e., the lowest risk group). We underscore that this approach, using a common reference class, is distinct from the main analysis which contrasted dementia hazards for patients with and without CCD for each modality separately (e.g., comparing hazards for dementia among patients with versus without WMD among those undergoing CT and separately among those undergoing MRI, using different reference classes for these contrasts).

Analyses were performed using SAS (Version 9.4 for Unix; SAS Institute, Cary, NC) and R Version 3.6.0 (R Foundation, Vienna, Austria).

## RESULTS

A total of 241,050 individuals receiving brain neuroimaging, with a total of 1,049,777 person-years of follow up time, were included in our analysis cohort (**Supplement B, eFigure 1)**.

The median follow-up time was 3.73 years (range of 61 days to 10.83 years; interquartile range [IQR] 1.61 to 6.85 years). 62,479 (25.9%) patients received MRI and 178,572 (74.1%) received CT scan. CCD was identified in 74,975 (31.1%) including 11,328 (4.7%) with CBI and 69,931 (29.0%) with WMD. There were 11,554 cases of dementia identified in follow-up, with a median time to event among those with dementia of 2.95 (IQR: 1.40 to 5.16) years. **Table 1** describes patient characteristics in the total cohort and in those with CBI (regardless of WMD) and those with WMD (regardless of CBI).

**Table 1.**
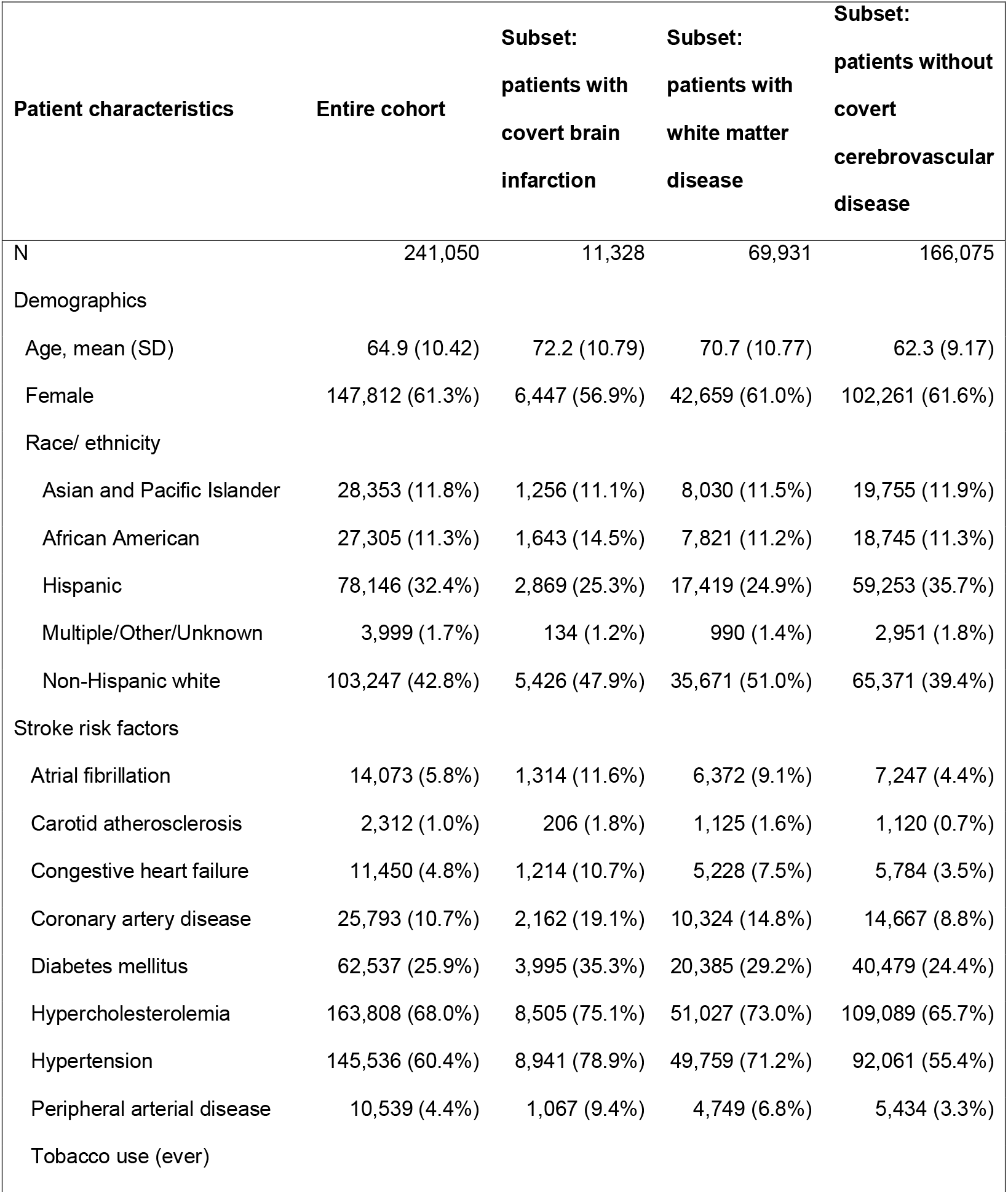
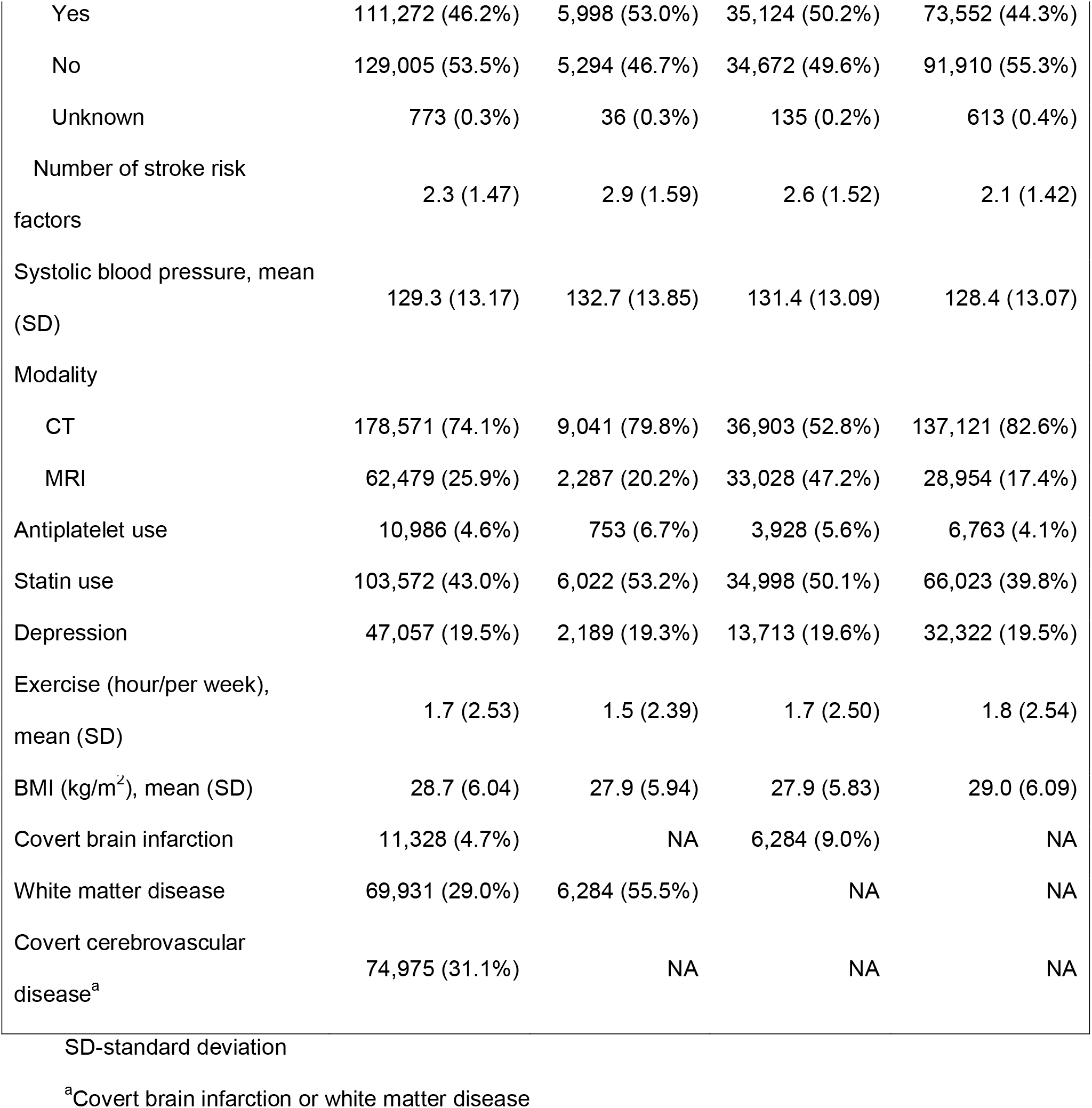
Patient demographic and clinical characteristics at baseline.

| Patient characteristics | Entire cohort | Subset:<br>patients with<br>covert brain<br>infarction | Subset:<br>patients with<br>white matter<br>disease | Subset:<br>patients without<br>covert<br>cerebrovascular<br>disease |
| --- | --- | --- | --- | --- |
| N | 241,050 | 11,328 | 69,931 | 166,075 |
| Demographics |  |  |  |  |
| Age, mean (SD) | 64.9 (10.42) | 72.2 (10.79) | 70.7 (10.77) | 62.3 (9.17) |
| Female | 147,812 (61.3%) | 6,447 (56.9%) | 42,659 (61.0%) | 102,261 (61.6%) |
| Race/ ethnicity |  |  |  |  |
| Asian and Pacific Islander | 28,353 (11.8%) | 1,256 (11.1%) | 8,030 (11.5%) | 19,755 (11.9%) |
| African American | 27,305 (11.3%) | 1,643 (14.5%) | 7,821 (11.2%) | 18,745 (11.3%) |
| Hispanic | 78,146 (32.4%) | 2,869 (25.3%) | 17,419 (24.9%) | 59,253 (35.7%) |
| Multiple/Other/Unknown | 3,999 (1.7%) | 134 (1.2%) | 990 (1.4%) | 2,951 (1.8%) |
| Non-Hispanic white | 103,247 (42.8%) | 5,426 (47.9%) | 35,671 (51.0%) | 65,371 (39.4%) |
| Stroke risk factors |  |  |  |  |
| Atrial fibrillation | 14,073 (5.8%) | 1,314 (11.6%) | 6,372 (9.1%) | 7,247 (4.4%) |
| Carotid atherosclerosis | 2,312 (1.0%) | 206 (1.8%) | 1,125 (1.6%) | 1,120 (0.7%) |
| Congestive heart failure | 11,450 (4.8%) | 1,214 (10.7%) | 5,228 (7.5%) | 5,784 (3.5%) |
| Coronary artery disease | 25,793 (10.7%) | 2,162 (19.1%) | 10,324 (14.8%) | 14,667 (8.8%) |
| Diabetes mellitus | 62,537 (25.9%) | 3,995 (35.3%) | 20,385 (29.2%) | 40,479 (24.4%) |
| Hypercholesterolemia | 163,808 (68.0%) | 8,505 (75.1%) | 51,027 (73.0%) | 109,089 (65.7%) |
| Hypertension | 145,536 (60.4%) | 8,941 (78.9%) | 49,759 (71.2%) | 92,061 (55.4%) |
| Peripheral arterial disease | 10,539 (4.4%) | 1,067 (9.4%) | 4,749 (6.8%) | 5,434 (3.3%) |
| Tobacco use (ever) |  |  |  |  |
| Yes | 111,272 (46.2%) | 5,998 (53.0%) | 35,124 (50.2%) | 73,552 (44.3%) |
| No | 129,005 (53.5%) | 5,294 (46.7%) | 34,672 (49.6%) | 91,910 (55.3%) |
| Unknown | 773 (0.3%) | 36 (0.3%) | 135 (0.2%) | 613 (0.4%) |
| Number of stroke risk factors | 2.3 (1.47) | 2.9 (1.59) | 2.6 (1.52) | 2.1 (1.42) |
| Systolic blood pressure, mean (SD) | 129.3 (13.17) | 132.7 (13.85) | 131.4 (13.09) | 128.4 (13.07) |
| Modality |  |  |  |  |
| CT | 178,571 (74.1%) | 9,041 (79.8%) | 36,903 (52.8%) | 137,121 (82.6%) |
| MRI | 62,479 (25.9%) | 2,287 (20.2%) | 33,028 (47.2%) | 28,954 (17.4%) |
| Antiplatelet use | 10,986 (4.6%) | 753 (6.7%) | 3,928 (5.6%) | 6,763 (4.1%) |
| Statin use | 103,572 (43.0%) | 6,022 (53.2%) | 34,998 (50.1%) | 66,023 (39.8%) |
| Depression | 47,057 (19.5%) | 2,189 (19.3%) | 13,713 (19.6%) | 32,322 (19.5%) |
| Exercise (hour/per week), mean (SD) | 1.7 (2.53) | 1.5 (2.39) | 1.7 (2.50) | 1.8 (2.54) |
| BMI (kg/m<sup>2</sup>), mean (SD) | 28.7 (6.04) | 27.9 (5.94) | 27.9 (5.83) | 29.0 (6.09) |
| Covert brain infarction | 11,328 (4.7%) | NA | 6,284 (9.0%) | NA |
| White matter disease | 69,931 (29.0%) | 6,284 (55.5%) | NA | NA |
| Covert cerebrovascular disease<sup>a</sup> | 74,975 (31.1%) | NA | NA | NA |

SD-standard deviation
<sup>a</sup>Covert brain infarction or white matter disease

The crude dementia incidence rates (per 1,000 person-years) were 29.4 (95% CI 27.9 to 31.0) in patients with incidentally-discovered CBI and 23.5 (22.90 to 24.0) for patients with WMD. This compares to an incidence rate of only 6.0 (5.8 to 6.2) in patients free of cerebrovascular disease (**Table 2**).

**Table 2.**
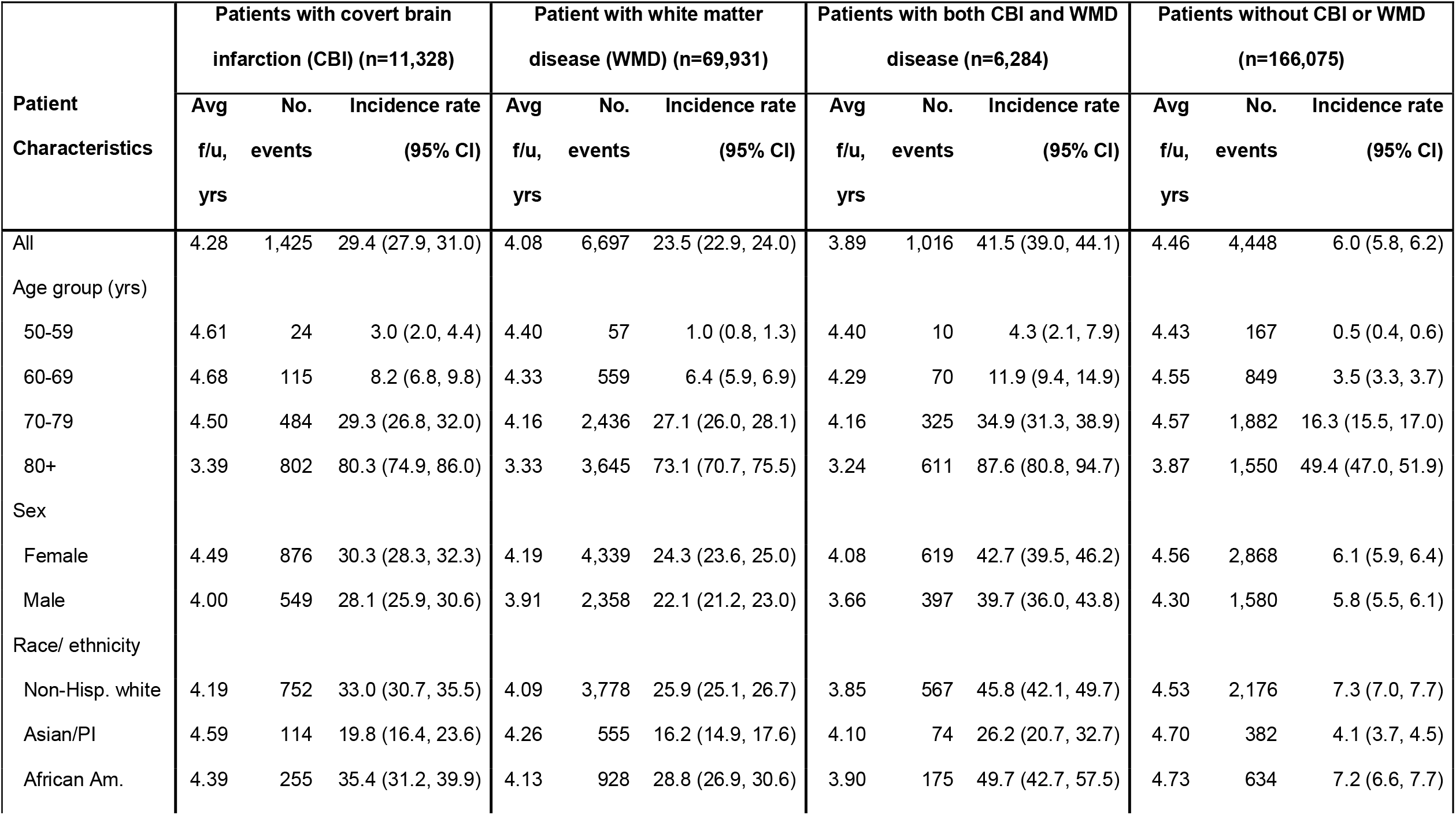
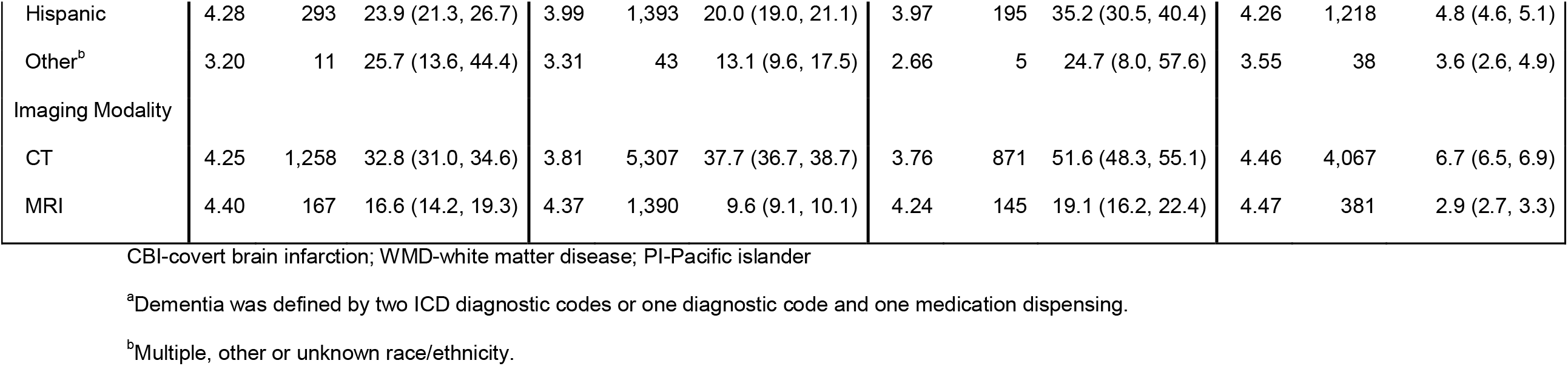
Dementia^a^ incidence rate overall and in subgroups (n=241,050).

| Patient Characteristics | Patients with covert brain infarction (CBI) (n=11,328) |  |  | Patient with white matter disease (WMD) (n=69,931) |  |  | Patients with both CBI and WMD disease (n=6,284) |  |  | Patients without CBI or WMD (n=166,075) |  |  |
| --- | --- | --- | --- | --- | --- | --- | --- | --- | --- | --- | --- | --- |
|  | Avg f/u, yrs | No. events | Incidence rate (95% CI) | Avg f/u, yrs | No. events | Incidence rate (95% CI) | Avg f/u, yrs | No. events | Incidence rate (95% CI) | Avg f/u, yrs | No. events | Incidence rate (95% CI) |
| All | 4.28 | 1,425 | 29.4 (27.9, 31.0) | 4.08 | 6,697 | 23.5 (22.9, 24.0) | 3.89 | 1,016 | 41.5 (39.0, 44.1) | 4.46 | 4,448 | 6.0 (5.8, 6.2) |
| Age group (yrs) |  |  |  |  |  |  |  |  |  |  |  |  |
| 50-59 | 4.61 | 24 | 3.0 (2.0, 4.4) | 4.40 | 57 | 1.0 (0.8, 1.3) | 4.40 | 10 | 4.3 (2.1, 7.9) | 4.43 | 167 | 0.5 (0.4, 0.6) |
| 60-69 | 4.68 | 115 | 8.2 (6.8, 9.8) | 4.33 | 559 | 6.4 (5.9, 6.9) | 4.29 | 70 | 11.9 (9.4, 14.9) | 4.55 | 849 | 3.5 (3.3, 3.7) |
| 70-79 | 4.50 | 484 | 29.3 (26.8, 32.0) | 4.16 | 2,436 | 27.1 (26.0, 28.1) | 4.16 | 325 | 34.9 (31.3, 38.9) | 4.57 | 1,882 | 16.3 (15.5, 17.0) |
| 80+ | 3.39 | 802 | 80.3 (74.9, 86.0) | 3.33 | 3,645 | 73.1 (70.7, 75.5) | 3.24 | 611 | 87.6 (80.8, 94.7) | 3.87 | 1,550 | 49.4 (47.0, 51.9) |
| Sex |  |  |  |  |  |  |  |  |  |  |  |  |
| Female | 4.49 | 876 | 30.3 (28.3, 32.3) | 4.19 | 4,339 | 24.3 (23.6, 25.0) | 4.08 | 619 | 42.7 (39.5, 46.2) | 4.56 | 2,868 | 6.1 (5.9, 6.4) |
| Male | 4.00 | 549 | 28.1 (25.9, 30.6) | 3.91 | 2,358 | 22.1 (21.2, 23.0) | 3.66 | 397 | 39.7 (36.0, 43.8) | 4.30 | 1,580 | 5.8 (5.5, 6.1) |
| Race/ ethnicity |  |  |  |  |  |  |  |  |  |  |  |  |
| Non-Hisp. white | 4.19 | 752 | 33.0 (30.7, 35.5) | 4.09 | 3,778 | 25.9 (25.1, 26.7) | 3.85 | 567 | 45.8 (42.1, 49.7) | 4.53 | 2,176 | 7.3 (7.0, 7.7) |
| Asian/PI | 4.59 | 114 | 19.8 (16.4, 23.6) | 4.26 | 555 | 16.2 (14.9, 17.6) | 4.10 | 74 | 26.2 (20.7, 32.7) | 4.70 | 382 | 4.1 (3.7, 4.5) |
| African Am. | 4.39 | 255 | 35.4 (31.2, 39.9) | 4.13 | 928 | 28.8 (26.9, 30.6) | 3.90 | 175 | 49.7 (42.7, 57.5) | 4.73 | 634 | 7.2 (6.6, 7.7) |
| Hispanic | 4.28 | 293 | 23.9 (21.3, 26.7) | 3.99 | 1,393 | 20.0 (19.0, 21.1) | 3.97 | 195 | 35.2 (30.5, 40.4) | 4.26 | 1,218 | 4.8 (4.6, 5.1) |
| Other<sup>b</sup> | 3.20 | 11 | 25.7 (13.6, 44.4) | 3.31 | 43 | 13.1 (9.6, 17.5) | 2.66 | 5 | 24.7 (8.0, 57.6) | 3.55 | 38 | 3.6 (2.6, 4.9) |
| Imaging Modality |  |  |  |  |  |  |  |  |  |  |  |  |
| CT | 4.25 | 1,258 | 32.8 (31.0, 34.6) | 3.81 | 5,307 | 37.7 (36.7, 38.7) | 3.76 | 871 | 51.6 (48.3, 55.1) | 4.46 | 4,067 | 6.7 (6.5, 6.9) |
| MRI | 4.40 | 167 | 16.6 (14.2, 19.3) | 4.37 | 1,390 | 9.6 (9.1, 10.1) | 4.24 | 145 | 19.1 (16.2, 22.4) | 4.47 | 381 | 2.9 (2.7, 3.3) |

CBI-covert brain infarction; WMD-white matter disease; PI-Pacific islander
<sup>a</sup>Dementia was defined by two ICD diagnostic codes or one diagnostic code and one medication dispensing.
<sup>b</sup>Multiple, other or unknown race/ethnicity.

Dementia-free survival in those with and without CBI with and without WMD, stratified by modality (CT versus MRI), are shown in **Figure 1a** and **1b**, respectively. These graphs display clearly the higher incidence rates in patients with CCD detected by CT compared to MRI. In patients with CBI, when detected by CT, the annualized incidence rate was 32.8 (31.0 to 34.5), whereas with MRI the incidence was 16.6 (14.2 to 19.3). In patients with WMD, when detected by CT, the annualized incidence rate was 37.7 (36.7 to 38.7), whereas with MRI the incidence was 9.6 (9.1 to 10.1). Overall, the crude hazard ratio (HR) for dementia associated with WMD was 3.72 (3.58 to 3.86); and with CBI was 2.91 (2.75 to 3.07) (**Table 3**).

**Table 3.** **Crude and adjusted hazard ratios (HRs) for dementia by covert brain infarction (CBI) and white matter disease (WMD)**

|  | HR | 95% CI |
| --- | --- | --- |
| ***Crude*** |  |  |
| CBI | 2.91 | (2.75, 3.07) |
| WMD | 3.72 | (3.58, 3.86) |
| ***Adjusted (varied by age and modality)*** |  |  |
| CBI |  |  |
| Age<70 years | 2.02 | (1.70, 2.41) |
| Age≥70 years | 1.22 | (1.15, 1.30) |
| WMD |  |  |
| MRI, age<70 | 2.28 | (1.99, 2.62) |
| MRI, age≥70 | 1.48 | (1.32, 1.66) |
| CT, age<70 | 2.87 | (2.58, 3.19) |
| CT, age≥70 | 1.87 | (1.79, 1.95) |

HR-hazard ratio; CBI-covert brain infarction; WMD-white matter disease; MRI-magnetic resonance imaging; CT-computerized tomography
The multivariable model included age, sex, race/ethnicity, atrial fibrillation, carotid atherosclerosis, congestive heart failure, coronary artery disease, diabetes mellitus, hypercholesterolemia, hypertension, peripheral arterial disease, ever tobacco use, mean systolic blood pressure in one year, antiplatelet use, statin use, depression, exercise (hour/per week), BMI (kg/m<sup>2</sup>), modality, age\*CBI, age\*WMD, and modality\*WMD. Refer to Supplement B, eTable 1 for HRs of all the covariates.

**Figure 1:**
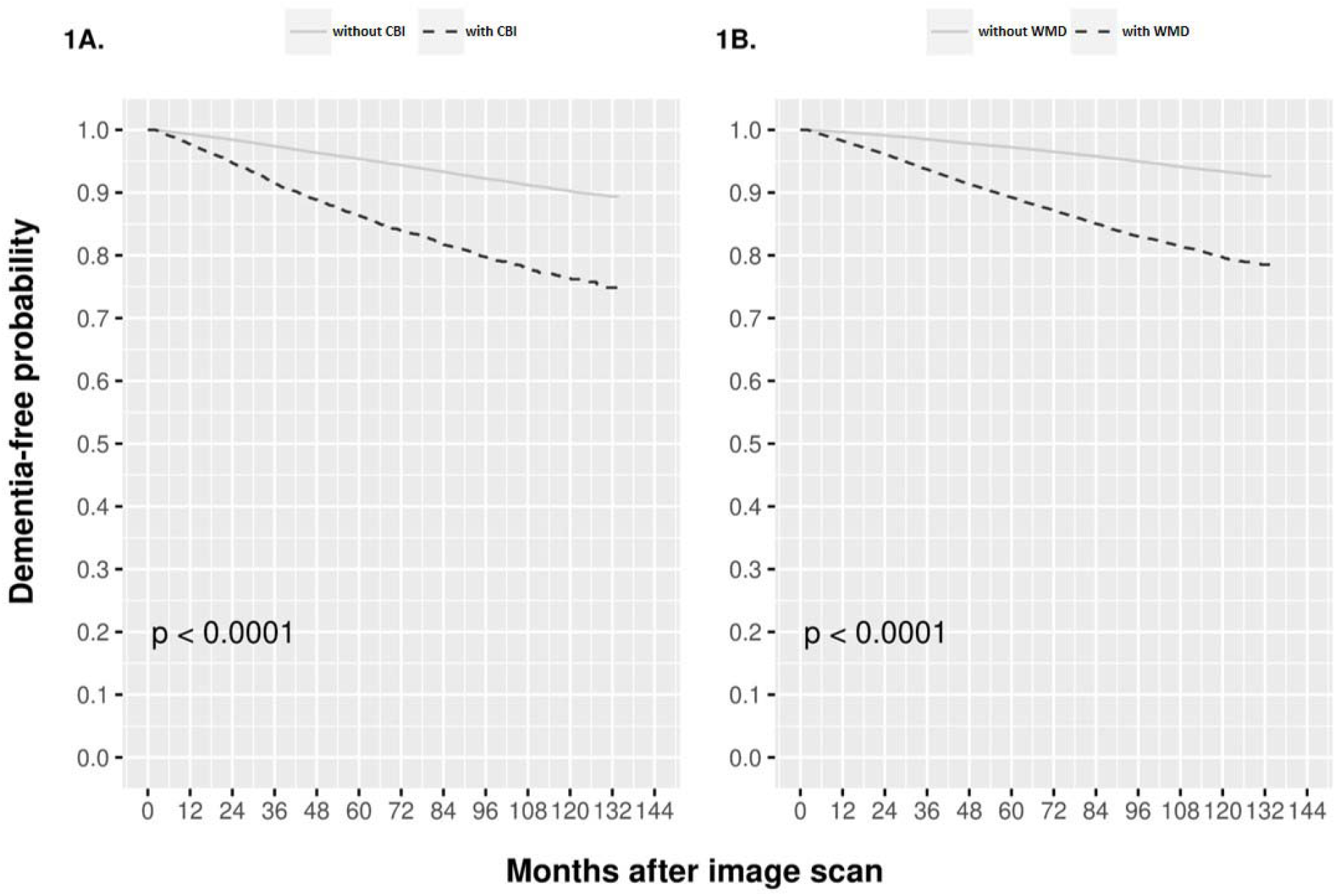
Kaplan-Meier plot of dementia-free survival with and without covert brain infarct (A) and with and without white matter disease (B) stratified by imaging modality (CT versus MRI) Differences in dementia free survival were assessed by the log rank test. CBI: Covert brain infarct; WMD: White matter disease; CT: Computed tomography; MR: Magnetic resonance

In a multivariable model controlling for major cardiovascular and dementia risk factors, we examined the effect of WMD on dementia risk. Among patients undergoing CT scan, the adjusted HR for the presence versus the absence of WMD was 2.87 (2.58 to 3.19) for those < age 70 and 1.87 (1.79 to 1.95) for those ≥ age 70. Among patients undergoing MRI, the adjusted HR for the presence versus the absence of WMD was 2.28 (1.99 to 2.62) for those < age 70 and 1.48 (1.32 to 1.66) for those ≥ age 70. The adjusted HR associated with CBI was 2.02 (1.70 to 2.41) for patients age <70 and 1.22 (1.15 to 1.30) for patients age ≥ 70 regardless of modality.

**Supplement B, eTable 1** shows the crude and adjusted effects of other risk factors included in our model. In general, the predictive effects of all cardiovascular and dementia risk factors were in the expected direction in the multivariable model. Patients on statins or antiplatelet agents at the time of the index scan had crudely higher risks of dementia, presumably due to confounding by indication, but these effects were nullified after full adjustment.

### Sensitivity Analyses

Dementia incidence patterns were largely the same across sensitivity analyses with a roughly uniform relative increase in outcome rates when dementia was established with only a single diagnostic code (**Supplement B, eTable 2**): 35.2 (95% CI: 33.5 to 36.9) per 1,000 person-years for patients with CBI; 27.9 (27.3 to 28.6) for patients with WMD and 7.3 (7.1 to 7.5) for patients without CBI or WMD. Patients without any indications for antithrombotic therapy had overall lower dementia incidence than the full cohort: 23.8 (22.1 to 25.5) per 1,000 person-years in those with CBI; 18.4 (17.8 to 19.0) for those with WMD; and 4.6 (4.5 to 4.8) for those without CCD (**Supplement B, eTable 3**). Neither incidence rates (**Supplement B, eTable 4**) nor HRs (**Supplement B, eTable 5**) changed substantially when follow-up started from 1 year post index scan rather than 60 days.

### Analysis of WMD grade

As shown in **Supplement B, eTable 6**, among patients imaged by CT scan, 21% (36903/178571) were found to have WMD. Of these, 55% had mild disease, 10% had moderate disease, and 5% had severe disease; severity was “undetermined” for the remaining 30% of patients. Among patients imaged by MRI, 53% (33028/62479) were found to have WMD. Of these, 62% had mild disease, 15% had moderate disease and 5% had severe disease; severity was underdetermined in the remaining 17% of patients. Within each imaging modality, the incidence of dementia increased monotonically in the expected direction among those with no, mild, moderate and severe disease. However, as shown in **Figure 2** and **Supplement B, eTable 6** the incidence was substantially higher among those imaged by CT. Incidence ranged from 3.1 (95% CI 2.8 to 3.4) per 1,000 person years among those who were WMD negative by MRI to 64.1 (58.3 to 70.4) per 1,000 person years for those with severe WMD as detected by CT scan. Patients with mild disease by CT scan had a similar incidence of dementia to those with severe disease on MRI. Adjusted HRs compared to patients who were negative for WMD by MRI ranged from 1.41 (1.25 to 1.60) for those with mild disease on MRI to 4.11 (3.58 to 4.72) for those with severe disease on CT scan (**Supplement B, eTable 6**).

**Figure 2:**
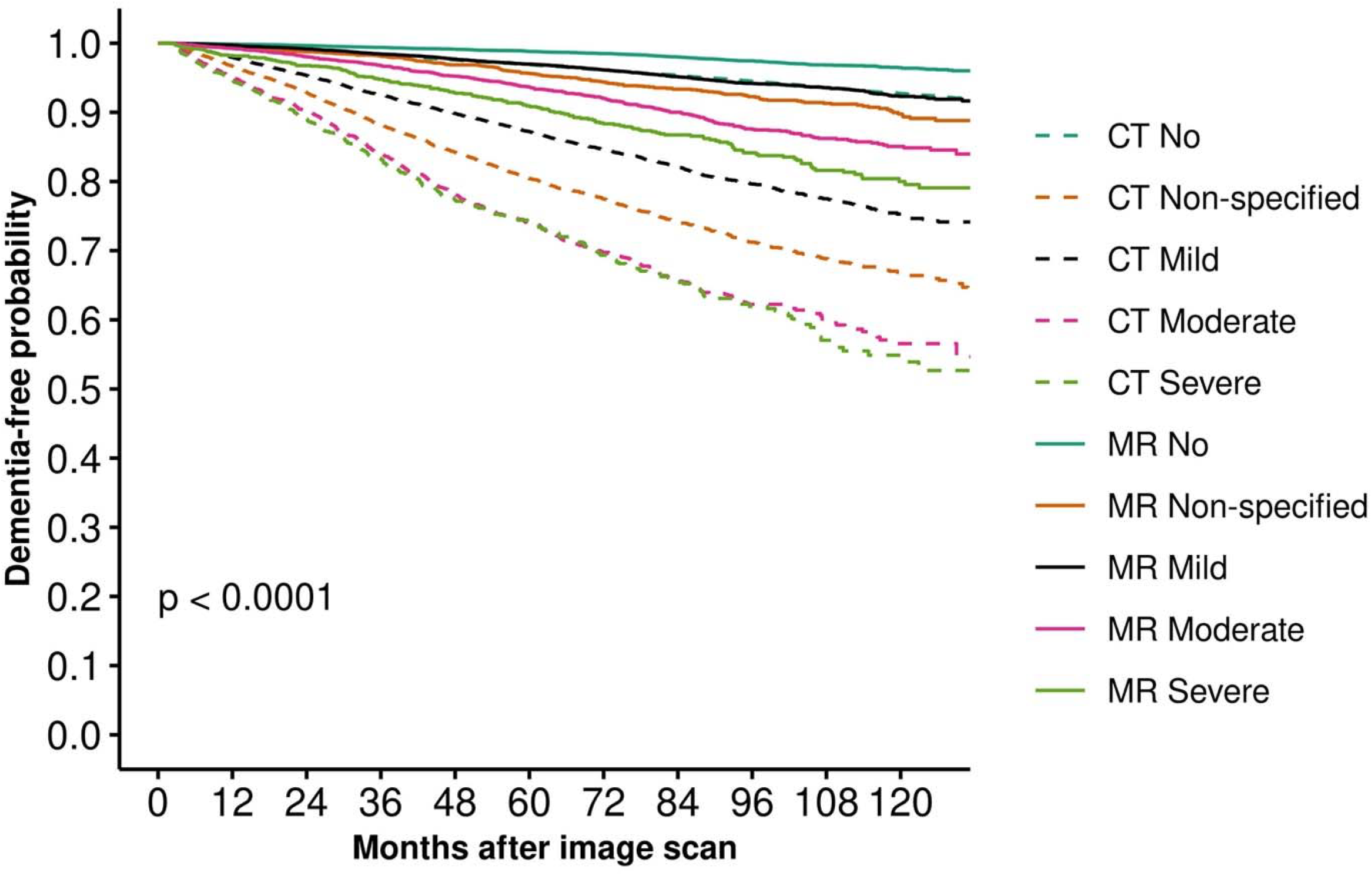
Kaplan-Meier plot of dementia-free survival by modality and white matter disease severity grade. CT: Computed tomography; MR: Magnetic resonance

## DISCUSSION

In this large observational cohort with approximately 250,000 subjects and over one million person-years of follow-up, we found that the presence of incidentally-discovered, NLP-identified CCD was strongly associated with an increased risk of future dementia. In patients between 50 and 70 years of age, CBI conferred a two-fold increased risk, after adjustment for other risk factors. On CT scan, patients with WMD had an adjusted risk almost three-fold that of those without WMD on CT scan; with MRI, the adjusted risk associated with WMD was approximately two-fold. These risk estimates are comparable to or greater than those for other major risk factors for dementia, including symptomatic ischemic stroke.^23-26^ While there remains some uncertainty regarding the potential heterogeneity of pathophysiology underlying CCD, these results support an interpretation of incidentally-discovered CBI or WMD as “stroke equivalents,” at least with regards to risk of dementia. The size of these effects was reduced in patients 70+ years of age, emphasizing the importance of early identification and the need for effective mid-life prevention strategies. These findings establish that routinely obtained neuroimaging reports carry substantial information that can be used to identify patients at high risk of progressing to dementia.

To our knowledge, this is the first cohort on which the prognostic significance of incidentally-discovered CCD with regard to future dementia has been studied. A prior study in this cohort found that CBI was a “stroke equivalent” in terms of the risk of future clinically evident stroke.^27^ While the findings here are similar to some studies in screened cohorts,^3,28^ there are additional insights not previously known. In particular, we found a dramatic increase in the incidence of dementia following CT-detected CCD compared to MRI-detected CCD. The incidence of dementia was two-fold higher when CBI was detected by CT scan versus MRI and four-fold higher when WMD was detected by CT scan versus MRI (**Table 2**). The difference in risk is presumably due to the lower sensitivity of CT scan, particularly for WMD. Thus, when changes are detected with CT scan, CCD is generally more advanced. This is consistent with our prior findings in this cohort of a much lower age-related incidence of WMD in patients imaged by CT scanning compared to MRI.^29^

Importantly, in 77% of patients with WMD, we were able to extract prognostically-informative disease severity information from routinely obtained neuroimaging reports, which underscored the difference between CT- and MRI-detected WMD. The cohort of patients without WMD by CT scan appears to have similar risk of dementia as compared to patients with mild WMD on MRI, presumably due in part to the presence of undetected WMD within the CT scan cohort. And patients with mild WMD on CT scan have a risk of dementia similar to or greater than those with severe WMD by MRI. Patients with severe WMD on CT have a dementia incidence approximately 20-fold higher than patients free of WMD on MRI, with an adjusted hazard ratio of 4, indicating that neuroimaging reports contain substantial information useful for risk stratification. Patients with incidentally-discovered CCD may be an attractive target for dementia prevention for several reasons. First, the overall incidence of dementia in these patients is high, well over 2% per year in the current study, which makes it a feasible outcome for a prevention trial. Second, it identifies patients with an elevated risk of dementias attributed to vascular risk factors for which there are already many known effective therapies. Third, the number of patients with CCD appears to be abundant and now identifiable: we found over 11,000 patients aged 50+ years of age with CBI and approximately 70,000 with WMD without prior symptomatic stroke or cognitive decline, including dementia, in a single health system, suggesting that neuroimaging reports might be a repository of a large number of high-risk patients suitable for trial enrollment. While prevention strategies for dementia are lacking, therapies used for secondary stroke prevention are attractive candidate interventions in this population, given the vascular contributions to the development of many dementias. And, in turn, this population may be an attractive high risk population for a trial. Antiplatelet medications have not previously been shown to be beneficial for prevention of dementia in a general population, but, in theory, may be helpful for patients with vascular brain injury (e.g. CCD).^30^ In this study, few patients were on prescribed antiplatelets (∼5%), an unknown percentage were on over-the-counter aspirin, and about one-third had other indications for antithrombotic therapy. Given the size of this cohort, it is likely that a large population of patients may be candidates for randomization to antiplatelets in a clinical trial. Similarly, only half of the patients in this cohort were on statins. Finally, besides age, hypertension remains the most consistently shared risk factor between CCD, symptomatic stroke, and dementia, making it a compelling target for intervention among patients with incidentally-discovered CCD.^31-35^ Novel agents currently being studied in stroke prevention may have unique benefits or a different risk-benefit balance with incidentally-discovered CCD.^36,37^

There are limitations to this study and the proposed approach to patient identification. Patients were selected into this cohort because they had clinically-indicated neuroimaging scans. Thus, the patients are not representative of the full spectrum of patients with WMD and CBI. Additionally, as these reports were obtained from routine care across multiple, heterogeneous clinical settings, there may be substantial limitations to the consistency of and the degree of details recoverable from these reports.

On the other hand, the use of routinely obtained neuroimaging reports as our source of data may also be viewed as the chief strength of our study. First, in the absence of population screening for CCD, incidentally-discovered CCD represents the most clinically relevant population. Further, by using reports obtained as part of routine care, we were able to estimate the prognostic value of imaging findings across diverse settings using heterogeneous imaging modalities without precise and standardized definitions of CBI and WMD, demonstrating the usefulness of routinely-obtained neuroimage reports. Interestingly, while MRI would be considered the gold standard to screen for CCD, CT scans comprised the majority of neuroimages included in our study; lesions discovered by CT scan appear to be substantially more prognostic than those discovered by MRI. Finally, unlike with artificial intelligence-driven direct analysis of neuroimages which may be subject to technical limitations of imaging data repositories (e.g., file size, transfer speed, cumbersome de-identification), we have demonstrated that neuroimaging reports are easily accessible and analyzable via NLP in the EHR of a large integrated health system.

## CONCLUSIONS

Incidentally-discovered CCD including CBI and WMD are common and associated with a high risk for future dementia. Identification of patients with these conditions with an NLP algorithm on routinely-obtained neuroimaging reports may facilitate dementia prevention studies and risk stratification.

## Data Availability

The datasets generated and/or analyzed during the current study are not publicly available due to ethical standards. The authors do not have permission to share data.

## ABBREVIATIONS

CBI: covert brain infarction
CCD: covert cerebrovascular diseases
CI: confidence intervals
CT: computer tomography
HR: hazard ratio
KPSC: Kaiser Permanente Southern California
NLP: natural language processing
WMD: white matter disease

## Author Contributorship

DMK contributed to conceptualization, funding acquisition, investigation, methodology, supervision, and wrote the original manuscript draft. YZ contributed to data curation, formal analysis, and investigation. LYL, PHL, DFK, and SF contributed to investigation and methodology. JN contributed to investigation. CZ contributed to data curation, investigation, and methodology. HL contributed to funding acquisition, investigation, methodology, and supervision. WC contributed to data curation, formal analysis, investigation, methodology, and supervision. All authors contributed equally to reviewing and editing the final manuscript.

## Data Availability Statement

The datasets generated and/or analyzed during the current study are not publicly available due to ethical standards. The authors do not have permission to share data. Authors Wansu Chen and Yichen Zhou had full access to all the data in the study and take responsibility for the integrity of the data and the accuracy of the data analysis.

## Ethical Approval and Patient Consent

This study was approved by the Tufts Health Sciences and KPSC Institutional Review Board. Patient consent was not required as all data was anonymized.

