## Supplement for "Association of Covert Cerebrovascular Disease Identified Using Natural Language Processing and Future Dementia"

**Supplementary Materials**

### **Supplement A: Supplementary methods**

#### **Supplement A1. ICD diagnosis codes and ICD/CPT procedure codes used to exclude patients from the cohort.**

| **ICD-9 Code** | **ICD-9 Description** | **ICD-10 Code** | **ICD-10 Description** |
| --- | --- | --- | --- |
| 006.5 | Amebic brain abscess | A06.6 | Amebic brain abscess |
| 013.20 | Tuberculoma of brain, unspecified | A17.81 | Tuberculoma of brain and spinal cord |
| 013.21 | Tuberculoma of brain, bacteriological or histological examination not done | A17.81 | Tuberculoma of brain and spinal cord |
| 013.22 | Tuberculoma of brain, bacteriological or histological examination unknown (at present) | A17.81 | Tuberculoma of brain and spinal cord |
| 013.23 | Tuberculoma of brain, tubercle bacilli found (in sputum) by microscopy | A17.81 | Tuberculoma of brain and spinal cord |
| 013.24 | Tuberculoma of brain, tubercle bacilli not found (in sputum) by microscopy, but found by bacterial culture | A17.81 | Tuberculoma of brain and spinal cord |
| 013.25 | Tuberculoma of brain, tubercle bacilli not found by bacteriological examination, but tuberculosis confirmed histologically | A17.81 | Tuberculoma of brain and spinal cord |
| 013.26 | Tuberculoma of brain, tubercle bacilli not found by bacteriological or histological examination, but tuberculosis confirmed by other methods [inoculation of animals] | A17.81 | Tuberculoma of brain and spinal cord |
| 013.30 | Tuberculous abscess of brain, unspecified | A17.81 | Tuberculoma of brain and spinal cord |
| 013.31 | Tuberculous abscess of brain, bacteriological or histological examination not done | A17.81 | Tuberculoma of brain and spinal cord |
| 013.32 | Tuberculous abscess of brain, bacteriological or histological examination unknown (at present) | A17.81 | Tuberculoma of brain and spinal cord |
| 013.33 | Tuberculous abscess of brain, tubercle bacilli found (in sputum) by microscopy | A17.81 | Tuberculoma of brain and spinal cord |
| 013.34 | Tuberculous abscess of brain, tubercle bacilli not found (in sputum) by microscopy, but found by bacterial culture | A17.81 | Tuberculoma of brain and spinal cord |
| 013.35 | Tuberculous abscess of brain, tubercle bacilli not found by bacteriological examination, but tuberculosis confirmed histologically | A17.81 | Tuberculoma of brain and spinal cord |
| 013.36 | Tuberculous abscess of brain, tubercle bacilli not found by bacteriological or histological examination, but tuberculosis confirmed by other methods [inoculation of animals] | A17.81 | Tuberculoma of brain and spinal cord |
| 191.0 | Malignant neoplasm of cerebrum, except lobes and ventricles | C71.0 | Malignant neoplasm of cerebrum, except lobes and ventricles |
| 191.1 | Malignant neoplasm of frontal lobe | C71.1 | Malignant neoplasm of frontal lobe |
| 191.2 | Malignant neoplasm of temporal lobe | C71.2 | Malignant neoplasm of temporal lobe |
| 191.3 | Malignant neoplasm of parietal lobe | C71.3 | Malignant neoplasm of parietal lobe |
| 191.4 | Malignant neoplasm of occipital lobe | C71.4 | Malignant neoplasm of occipital lobe |
| 191.5 | Malignant neoplasm of ventricles | C71.5 | Malignant neoplasm of cerebral ventricle |
| 191.6 | Malignant neoplasm of cerebellum nos | C71.6 | Malignant neoplasm of cerebellum |
| 191.7 | Malignant neoplasm of brain stem | C71.7 | Malignant neoplasm of brain stem |
| 191.8 | Malignant neoplasm of other parts of brain | C71.8 | Malignant neoplasm of overlapping sites of brain |
| 191.9 | Malignant neoplasm of brain, unspecified | C71.9 | Malignant neoplasm of brain, unspecified |
| 239.6 | Neoplasm of unspecified nature of brain | D49.6 | Neoplasm of unspecified behavior of brain |
| 290.0 | Senile dementia, uncomplicated | F03.90 | Unspecified dementia without behavioral disturbance |
| 290.10 | Presenile dementia, uncomplicated | F03.90 | Unspecified dementia without behavioral disturbance |
| 290.11 | Presenile dementia with delirium | F03.90 | Unspecified dementia without behavioral disturbance |
| 290.12 | Presenile dementia with delusional features | F03.90 | Unspecified dementia without behavioral disturbance |
| 290.12 | Presenile dementia with delusional features |  |  |
| 290.13 | Presenile dementia with depressive features | F03.90 | Unspecified dementia without behavioral disturbance |
| 290.20 | Senile dementia with delusional features | F03.90 | Unspecified dementia without behavioral disturbance |
| 290.20 | Senile dementia with delusional features |  |  |
| 290.21 | Senile dementia with depressive features | F03.90 | Unspecified dementia without behavioral disturbance |
| 290.3 | Senile dementia with delirium | F03.90 | Unspecified dementia without behavioral disturbance |
| 290.3 | Senile dementia with delirium |  |  |
| 290.40 | Vascular dementia, uncomplicated | F01.50 | Vascular dementia without behavioral disturbance |
| 290.41 | Vascular dementia, with delirium | F01.51 | Vascular dementia with behavioral disturbance |
| 290.42 | Vascular dementia, with delusions | F01.51 | Vascular dementia with behavioral disturbance |
| 290.43 | Vascular dementia, with depressed mood | F01.51 | Vascular dementia with behavioral disturbance |
| 290.8 | Other specified senile psychotic conditions | F03.90 | Unspecified dementia without behavioral disturbance |
| 290.9 | Unspecified senile psychotic condition | F03.90 | Unspecified dementia without behavioral disturbance |
| 291.2 | Alcohol-induced persisting dementia | F10.27 | Alcohol dependence with alcohol-induced persisting dementia |
| 292.82 | Drug-induced persisting dementia | F13.27 | Sedative, hypnotic or anxiolytic dependence with sedative, hypnotic or anxiolytic-induced persisting dementia |
| 292.82 | Drug-induced persisting dementia | F13.97 | Sedative, hypnotic or anxiolytic use, unspecified with sedative, hypnotic or anxiolytic-induced persisting dementia |
| 292.82 | Drug-induced persisting dementia | F18.17 | Inhalant abuse with inhalant-induced dementia |
| 292.82 | Drug-induced persisting dementia | F18.27 | Inhalant dependence with inhalant-induced dementia |
| 292.82 | Drug-induced persisting dementia | F18.97 | Inhalant use, unspecified with inhalant-induced persisting dementia |
| 292.82 | Drug-induced persisting dementia | F19.17 | Other psychoactive substance abuse with psychoactive substance-induced persisting dementia |
| 292.82 | Drug-induced persisting dementia | F19.27 | Other psychoactive substance dependence with psychoactive substance-induced persisting dementia |
| 292.82 | Drug-induced persisting dementia | F19.97 | Other psychoactive substance use, unspecified with psychoactive substance-induced persisting dementia |
| 294.10 | Dementia in conditions classified elsewhere without behavioral disturbance | F02.80 | Dementia in other diseases classified elsewhere without behavioral disturbance |
| 294.11 | Dementia in conditions classified elsewhere with behavioral disturbance | F02.81 | Dementia in other diseases classified elsewhere with behavioral disturbance |
| 294.20 | Dementia, unspecified, without behavioral disturbance | F03.90 | Unspecified dementia without behavioral disturbance |
| 294.21 | Dementia, unspecified, with behavioral disturbance | F03.91 | Unspecified dementia with behavioral disturbance |
| 315.31 | Expressive language disorder | F80.1 | Expressive language disorder |
| 324.0 | Intracranial abscess | G06.0 | Intracranial abscess and granuloma |
| 326. | Late effects of intracranial abscess or pyogenic infection | G09. | Sequelae of inflammatory diseases of central nervous system |
| 331.0 | Alzheimer's disease | G30.0 | Alzheimer's disease with early onset |
| 331.0 | Alzheimer's disease | G30.1 | Alzheimer's disease with late onset |
| 331.0 | Alzheimer's disease | G30.8 | Other Alzheimer's disease |
| 331.0 | Alzheimer's disease | G30.9 | Alzheimer's disease, unspecified |
| 331.11 | Pick's disease | G31.01 | Pick's disease |
| 331.19 | Other frontotemporal dementia | G31.09 | Other frontotemporal dementia |
| 331.2 | Senile degeneration of brain | G31.1 | Senile degeneration of brain, not elsewhere classified |
| 331.3 | Communicating hydrocephalus | G91.0 | Communicating hydrocephalus |
| 331.4 | Obstructive hydrocephalus | G91.1 | Obstructive hydrocephalus |
| 331.4 | Obstructive hydrocephalus | G91.3 | Post-traumatic hydrocephalus, unspecified |
| 331.4 | Obstructive hydrocephalus | G91.8 | Other hydrocephalus |
| 331.4 | Obstructive hydrocephalus | G91.9 | Hydrocephalus, unspecified |
| 331.5 | Idiopathic normal pressure hydrocephalus (INPH) | G91.2 | (Idiopathic) normal pressure hydrocephalus |
| 331.6 | Corticobasal degeneration | G31.85 | Corticobasal degeneration |
| 331.82 | Dementia with lewy bodies | G31.83 | Dementia with Lewy bodies |
| 340. | Multiple sclerosis | G35. | Multiple sclerosis |
| 342.00 | Flaccid hemiplegia and hemiparesis affecting unspecified side | G81.00 | Flaccid hemiplegia affecting unspecified side |
| 342.01 | Flaccid hemiplegia and hemiparesis affecting dominant side | G81.01 | Flaccid hemiplegia affecting right dominant side |
| 342.01 | Flaccid hemiplegia and hemiparesis affecting dominant side | G81.02 | Flaccid hemiplegia affecting left dominant side |
| 342.02 | Flaccid hemiplegia and hemiparesis affecting nondominant side | G81.03 | Flaccid hemiplegia affecting right nondominant side |
| 342.02 | Flaccid hemiplegia and hemiparesis affecting nondominant side | G81.04 | Flaccid hemiplegia affecting left nondominant side |
| 342.10 | Spastic hemiplegia and hemiparesis affecting unspecified side | G81.10 | Spastic hemiplegia affecting unspecified side |
| 342.11 | Spastic hemiplegia and hemiparesis affecting dominant side | G81.11 | Spastic hemiplegia affecting right dominant side |
| 342.11 | Spastic hemiplegia and hemiparesis affecting dominant side | G81.12 | Spastic hemiplegia affecting left dominant side |
| 342.12 | Spastic hemiplegia and hemiparesis affecting nondominant side | G81.13 | Spastic hemiplegia affecting right nondominant side |
| 342.12 | Spastic hemiplegia and hemiparesis affecting nondominant side | G81.14 | Spastic hemiplegia affecting left nondominant side |
| 342.80 | Other specified hemiplegia and hemiparesis affecting unspecified side | G81.90 | Hemiplegia, unspecified affecting unspecified side |
| 342.81 | Other specified hemiplegia and hemiparesis affecting dominant side | G81.91 | Hemiplegia, unspecified affecting right dominant side |
| 342.81 | Other specified hemiplegia and hemiparesis affecting dominant side | G81.92 | Hemiplegia, unspecified affecting left dominant side |
| 342.82 | Other specified hemiplegia and hemiparesis affecting nondominant side | G81.93 | Hemiplegia, unspecified affecting right nondominant side |
| 342.82 | Other specified hemiplegia and hemiparesis affecting nondominant side | G81.94 | Hemiplegia, unspecified affecting left nondominant side |
| 342.90 | Hemiplegia, unspecified, affecting unspecified side | G81.90 | Hemiplegia, unspecified affecting unspecified side |
| 342.91 | Hemiplegia, unspecified, affecting dominant side | G81.91 | Hemiplegia, unspecified affecting right dominant side |
| 342.91 | Hemiplegia, unspecified, affecting dominant side | G81.92 | Hemiplegia, unspecified affecting left dominant side |
| 342.92 | Hemiplegia, unspecified, affecting nondominant side | G81.93 | Hemiplegia, unspecified affecting right nondominant side |
| 342.92 | Hemiplegia, unspecified, affecting nondominant side | G81.94 | Hemiplegia, unspecified affecting left nondominant side |
| 344.30 | Monoplegia of lower limb affecting unspecified side | G83.10 | Monoplegia of lower limb affecting unspecified side |
| 344.31 | Monoplegia of lower limb affecting dominant side | G83.11 | Monoplegia of lower limb affecting right dominant side |
| 344.31 | Monoplegia of lower limb affecting dominant side | G83.12 | Monoplegia of lower limb affecting left dominant side |
| 344.32 | Monoplegia of lower limb affecting nondominant side | G83.13 | Monoplegia of lower limb affecting right nondominant side |
| 344.32 | Monoplegia of lower limb affecting nondominant side | G83.14 | Monoplegia of lower limb affecting left nondominant side |
| 344.40 | Monoplegia of upper limb affecting unspecified side | G83.20 | Monoplegia of upper limb affecting unspecified side |
| 344.41 | Monoplegia of upper limb affecting dominant side | G83.21 | Monoplegia of upper limb affecting right dominant side |
| 344.41 | Monoplegia of upper limb affecting dominant side | G83.22 | Monoplegia of upper limb affecting left dominant side |
| 344.42 | Monoplegia of upper limb affecting nondominant sde | G83.23 | Monoplegia of upper limb affecting right nondominant side |
| 344.42 | Monoplegia of upper limb affecting nondominant sde | G83.24 | Monoplegia of upper limb affecting left nondominant side |
| 344.5 | Unspecified monoplegia | G83.30 | Monoplegia, unspecified affecting unspecified side |
| 344.5 | Unspecified monoplegia | G83.31 | Monoplegia, unspecified affecting right dominant side |
| 344.5 | Unspecified monoplegia | G83.32 | Monoplegia, unspecified affecting left dominant side |
| 344.5 | Unspecified monoplegia | G83.33 | Monoplegia, unspecified affecting right nondominant side |
| 344.5 | Unspecified monoplegia | G83.34 | Monoplegia, unspecified affecting left nondominant side |
| 344.9 | Paralysis, unspecified | G83.9 | Paralytic syndrome, unspecified |
| 348.1 | Anoxic brain damage | G93.1 | Anoxic brain damage, not elsewhere classified |
| 362.30 | Retinal vascular occlusion, unspecified | H34.9 | Unspecified retinal vascular occlusion |
| 362.31 | Central retinal artery occlusion | H34.10 | Central retinal artery occlusion, unspecified eye |
| 362.31 | Central retinal artery occlusion | H34.11 | Central retinal artery occlusion, right eye |
| 362.31 | Central retinal artery occlusion | H34.12 | Central retinal artery occlusion, left eye |
| 362.31 | Central retinal artery occlusion | H34.13 | Central retinal artery occlusion, bilateral |
| 362.32 | Retinal arterial branch occlusion | H34.231 | Retinal artery branch occlusion, right eye |
| 362.32 | Retinal arterial branch occlusion | H34.232 | Retinal artery branch occlusion, left eye |
| 362.32 | Retinal arterial branch occlusion | H34.233 | Retinal artery branch occlusion, bilateral |
| 362.32 | Retinal arterial branch occlusion | H34.239 | Retinal artery branch occlusion, unspecified eye |
| 362.33 | Partial retinal arterial occlusion | H34.211 | Partial retinal artery occlusion, right eye |
| 362.33 | Partial retinal arterial occlusion | H34.212 | Partial retinal artery occlusion, left eye |
| 362.33 | Partial retinal arterial occlusion | H34.213 | Partial retinal artery occlusion, bilateral |
| 362.33 | Partial retinal arterial occlusion | H34.219 | Partial retinal artery occlusion, unspecified eye |
| 362.34 | Transient retinal arterial occlusion | G45.3 | Amaurosis fugax |
| 362.34 | Transient retinal arterial occlusion | H34.00 | Transient retinal artery occlusion, unspecified eye |
| 362.34 | Transient retinal arterial occlusion | H34.01 | Transient retinal artery occlusion, right eye |
| 362.34 | Transient retinal arterial occlusion | H34.02 | Transient retinal artery occlusion, left eye |
| 362.34 | Transient retinal arterial occlusion | H34.03 | Transient retinal artery occlusion, bilateral |
| 368.00 | Amblyopia, unspecified |  |  |
| 368.01 | Strabismic amblyopia |  |  |
| 368.02 | Deprivation amblyopia |  |  |
| 368.03 | Refractive amblyopia |  |  |
| 368.11 | Sudden visual loss | H53.131 | Sudden visual loss, right eye |
| 368.11 | Sudden visual loss | H53.132 | Sudden visual loss, left eye |
| 368.11 | Sudden visual loss | H53.133 | Sudden visual loss, bilateral |
| 368.11 | Sudden visual loss | H53.139 | Sudden visual loss, unspecified eye |
| 368.12 | Transient visual loss | H53.121 | Transient visual loss, right eye |
| 368.12 | Transient visual loss | H53.122 | Transient visual loss, left eye |
| 368.12 | Transient visual loss | H53.123 | Transient visual loss, bilateral |
| 368.12 | Transient visual loss | H53.129 | Transient visual loss, unspecified eye |
| 368.13 | Visual discomfort |  |  |
| 368.14 | Visual distortions of shape and size | H53.15 | Visual distortions of shape and size |
| 368.15 | Other visual distortions and entoptic phenomena |  |  |
| 368.16 | Psychophysical visual disturbances | R48.3 | Visual agnosia |
| 368.16 | Psychophysical visual disturbances |  |  |
| 368.2 | Diplopia | H53.2 | Diplopia |
| 368.30 | Binocular vision disorder, unspecified | H53.30 | Unspecified disorder of binocular vision |
| 368.31 | Suppression of binocular vision |  |  |
| 368.32 | Simultaneous visual perception without fusion |  |  |
| 368.33 | Fusion with defective stereopsis |  |  |
| 368.34 | Abnormal retinal correspondence |  |  |
| 368.40 | Visual field defect, unspecified | H53.40 | Unspecified visual field defects |
| 368.41 | Scotoma involving central area | H53.411 | Scotoma involving central area, right eye |
| 368.41 | Scotoma involving central area | H53.412 | Scotoma involving central area, left eye |
| 368.41 | Scotoma involving central area | H53.413 | Scotoma involving central area, bilateral |
| 368.41 | Scotoma involving central area | H53.419 | Scotoma involving central area, unspecified eye |
| 368.42 | Scotoma of blind spot area | H53.421 | Scotoma of blind spot area, right eye |
| 368.42 | Scotoma of blind spot area | H53.422 | Scotoma of blind spot area, left eye |
| 368.42 | Scotoma of blind spot area | H53.423 | Scotoma of blind spot area, bilateral |
| 368.42 | Scotoma of blind spot area | H53.429 | Scotoma of blind spot area, unspecified eye |
| 368.43 | Sector or arcuate visual field defects | H53.431 | Sector or arcuate defects, right eye |
| 368.43 | Sector or arcuate visual field defects | H53.432 | Sector or arcuate defects, left eye |
| 368.43 | Sector or arcuate visual field defects | H53.433 | Sector or arcuate defects, bilateral |
| 368.43 | Sector or arcuate visual field defects | H53.439 | Sector or arcuate defects, unspecified eye |
| 368.44 | Other localized visual field defect | H53.451 | Other localized visual field defect, right eye |
| 368.44 | Other localized visual field defect | H53.452 | Other localized visual field defect, left eye |
| 368.44 | Other localized visual field defect | H53.453 | Other localized visual field defect, bilateral |
| 368.44 | Other localized visual field defect | H53.459 | Other localized visual field defect, unspecified eye |
| 368.45 | Generalized visual field contraction or constriction | H53.481 | Generalized contraction of visual field, right eye |
| 368.45 | Generalized visual field contraction or constriction | H53.482 | Generalized contraction of visual field, left eye |
| 368.45 | Generalized visual field contraction or constriction | H53.483 | Generalized contraction of visual field, bilateral |
| 368.45 | Generalized visual field contraction or constriction | H53.489 | Generalized contraction of visual field, unspecified eye |
| 368.46 | Homonymous bilateral field defects | H53.461 | Homonymous bilateral field defects, right side |
| 368.46 | Homonymous bilateral field defects | H53.462 | Homonymous bilateral field defects, left side |
| 368.46 | Homonymous bilateral field defects | H53.469 | Homonymous bilateral field defects, unspecified side |
| 368.47 | Heteronymous bilateral field defects | H53.47 | Heteronymous bilateral field defects |
| 368.51 | Protan defect |  |  |
| 368.52 | Deutan defect |  |  |
| 368.53 | Tritan defect |  |  |
| 368.54 | Achromatopsia |  |  |
| 368.55 | Acquired color vision deficiencies |  |  |
| 368.59 | Other color vision deficiencies |  |  |
| 368.60 | Night blindness, unspecified |  |  |
| 368.61 | Congenital night blindness |  |  |
| 368.62 | Acquired night blindness |  |  |
| 368.63 | Abnormal dark adaptation curve |  |  |
| 368.69 | Other night blindness |  |  |
| 369.8 | Unqualified visual loss, one eye | H54.60 | Unqualified visual loss, one eye, unspecified |
| 369.8 | Unqualified visual loss, one eye | H54.61 | Unqualified visual loss, right eye, normal vision left eye |
| 369.8 | Unqualified visual loss, one eye | H54.62 | Unqualified visual loss, left eye, normal vision right eye |
| 369.9 | Unspecified visual loss | H54.7 | Unspecified visual loss |
| 377.62 | Disorders of other visual pathways associated with vascular disorders | H47.531 | Disorders of visual pathways in (due to) vascular disorders, right side |
| 377.62 | Disorders of other visual pathways associated with vascular disorders | H47.532 | Disorders of visual pathways in (due to) vascular disorders, left side |
| 377.62 | Disorders of other visual pathways associated with vascular disorders | H47.539 | Disorders of visual pathways in (due to) vascular disorders, unspecified side |
| 377.72 | Disorders of visual cortex associated with vascular disorders | H47.641 | Disorders of visual cortex in (due to) vascular disorders, right side of brain |
| 377.72 | Disorders of visual cortex associated with vascular disorders | H47.642 | Disorders of visual cortex in (due to) vascular disorders, left side of brain |
| 377.72 | Disorders of visual cortex associated with vascular disorders | H47.649 | Disorders of visual cortex in (due to) vascular disorders, unspecified side of brain |
| 430. | Subarachnoid hemorrhage | I60.00 | Nontraumatic subarachnoid hemorrhage from unspecified carotid siphon and bifurcation |
| 430. | Subarachnoid hemorrhage | I60.01 | Nontraumatic subarachnoid hemorrhage from right carotid siphon and bifurcation |
| 430. | Subarachnoid hemorrhage | I60.02 | Nontraumatic subarachnoid hemorrhage from left carotid siphon and bifurcation |
| 430. | Subarachnoid hemorrhage | I60.10 | Nontraumatic subarachnoid hemorrhage from unspecified middle cerebral artery |
| 430. | Subarachnoid hemorrhage | I60.11 | Nontraumatic subarachnoid hemorrhage from right middle cerebral artery |
| 430. | Subarachnoid hemorrhage | I60.12 | Nontraumatic subarachnoid hemorrhage from left middle cerebral artery |
| 430. | Subarachnoid hemorrhage | I60.2 | Nontraumatic subarachnoid hemorrhage from anterior communicating artery |
| 430. | Subarachnoid hemorrhage | I60.30 | Nontraumatic subarachnoid hemorrhage from unspecified posterior communicating artery |
| 430. | Subarachnoid hemorrhage | I60.31 | Nontraumatic subarachnoid hemorrhage from right posterior communicating artery |
| 430. | Subarachnoid hemorrhage | I60.32 | Nontraumatic subarachnoid hemorrhage from left posterior communicating artery |
| 430. | Subarachnoid hemorrhage | I60.4 | Nontraumatic subarachnoid hemorrhage from basilar artery |
| 430. | Subarachnoid hemorrhage | I60.50 | Nontraumatic subarachnoid hemorrhage from unspecified vertebral artery |
| 430. | Subarachnoid hemorrhage | I60.51 | Nontraumatic subarachnoid hemorrhage from right vertebral artery |
| 430. | Subarachnoid hemorrhage | I60.52 | Nontraumatic subarachnoid hemorrhage from left vertebral artery |
| 430. | Subarachnoid hemorrhage | I60.6 | Nontraumatic subarachnoid hemorrhage from other intracranial arteries |
| 430. | Subarachnoid hemorrhage | I60.7 | Nontraumatic subarachnoid hemorrhage from unspecified intracranial artery |
| 430. | Subarachnoid hemorrhage | I60.8 | Other nontraumatic subarachnoid hemorrhage |
| 430. | Subarachnoid hemorrhage | I60.9 | Nontraumatic subarachnoid hemorrhage, unspecified |
| 431. | Intracerebral hemorrhage | I61.0 | Nontraumatic intracerebral hemorrhage in hemisphere, subcortical |
| 431. | Intracerebral hemorrhage | I61.1 | Nontraumatic intracerebral hemorrhage in hemisphere, cortical |
| 431. | Intracerebral hemorrhage | I61.2 | Nontraumatic intracerebral hemorrhage in hemisphere, unspecified |
| 431. | Intracerebral hemorrhage | I61.3 | Nontraumatic intracerebral hemorrhage in brain stem |
| 431. | Intracerebral hemorrhage | I61.4 | Nontraumatic intracerebral hemorrhage in cerebellum |
| 431. | Intracerebral hemorrhage | I61.5 | Nontraumatic intracerebral hemorrhage, intraventricular |
| 431. | Intracerebral hemorrhage | I61.6 | Nontraumatic intracerebral hemorrhage, multiple localized |
| 431. | Intracerebral hemorrhage | I61.8 | Other nontraumatic intracerebral hemorrhage |
| 431. | Intracerebral hemorrhage | I61.9 | Nontraumatic intracerebral hemorrhage, unspecified |
| 432.0 | Nontraumatic extradural hemorrhage | I62.1 | Nontraumatic extradural hemorrhage |
| 432.1 | Subdural hemorrhage | I62.00 | Nontraumatic subdural hemorrhage, unspecified |
| 432.1 | Subdural hemorrhage | I62.01 | Nontraumatic acute subdural hemorrhage |
| 432.1 | Subdural hemorrhage | I62.02 | Nontraumatic subacute subdural hemorrhage |
| 432.1 | Subdural hemorrhage | I62.03 | Nontraumatic chronic subdural hemorrhage |
| 432.9 | Unspecified intracranial hemorrhage | I62.9 | Nontraumatic intracranial hemorrhage, unspecified |
| 433.01 | Occlusion and stenosis of basilar artery with cerebral infarction | I63.02 | Cerebral infarction due to thrombosis of basilar artery |
| 433.01 | Occlusion and stenosis of basilar artery with cerebral infarction | I63.12 | Cerebral infarction due to embolism of basilar artery |
| 433.01 | Occlusion and stenosis of basilar artery with cerebral infarction | I63.22 | Cerebral infarction due to unspecified occlusion or stenosis of basilar artery |
| 433.11 | Occlusion and stenosis of carotid artery with cerebral infarction | I63.031 | Cerebral infarction due to thrombosis of right carotid artery |
| 433.11 | Occlusion and stenosis of carotid artery with cerebral infarction | I63.032 | Cerebral infarction due to thrombosis of left carotid artery |
| 433.11 | Occlusion and stenosis of carotid artery with cerebral infarction | I63.033 | Cerebral infarction due to thrombosis of bilateral carotid arteries |
| 433.11 | Occlusion and stenosis of carotid artery with cerebral infarction | I63.039 | Cerebral infarction due to thrombosis of unspecified carotid artery |
| 433.11 | Occlusion and stenosis of carotid artery with cerebral infarction | I63.131 | Cerebral infarction due to embolism of right carotid artery |
| 433.11 | Occlusion and stenosis of carotid artery with cerebral infarction | I63.132 | Cerebral infarction due to embolism of left carotid artery |
| 433.11 | Occlusion and stenosis of carotid artery with cerebral infarction | I63.133 | Cerebral infarction due to embolism of bilateral carotid arteries |
| 433.11 | Occlusion and stenosis of carotid artery with cerebral infarction | I63.139 | Cerebral infarction due to embolism of unspecified carotid artery |
| 433.11 | Occlusion and stenosis of carotid artery with cerebral infarction | I63.231 | Cerebral infarction due to unspecified occlusion or stenosis of right carotid arteries |
| 433.11 | Occlusion and stenosis of carotid artery with cerebral infarction | I63.232 | Cerebral infarction due to unspecified occlusion or stenosis of left carotid arteries |
| 433.11 | Occlusion and stenosis of carotid artery with cerebral infarction | I63.233 | Cerebral infarction due to unspecified occlusion or stenosis of bilateral carotid arteries |
| 433.11 | Occlusion and stenosis of carotid artery with cerebral infarction | I63.239 | Cerebral infarction due to unspecified occlusion or stenosis of unspecified carotid arteries |
| 433.21 | Occlusion and stenosis of vertebral artery with cerebral infarction | I63.011 | Cerebral infarction due to thrombosis of right vertebral artery |
| 433.21 | Occlusion and stenosis of vertebral artery with cerebral infarction | I63.012 | Cerebral infarction due to thrombosis of left vertebral artery |
| 433.21 | Occlusion and stenosis of vertebral artery with cerebral infarction | I63.013 | Cerebral infarction due to thrombosis of bilateral vertebral arteries |
| 433.21 | Occlusion and stenosis of vertebral artery with cerebral infarction | I63.019 | Cerebral infarction due to thrombosis of unspecified vertebral artery |
| 433.21 | Occlusion and stenosis of vertebral artery with cerebral infarction | I63.111 | Cerebral infarction due to embolism of right vertebral artery |
| 433.21 | Occlusion and stenosis of vertebral artery with cerebral infarction | I63.112 | Cerebral infarction due to embolism of left vertebral artery |
| 433.21 | Occlusion and stenosis of vertebral artery with cerebral infarction | I63.113 | Cerebral infarction due to embolism of bilateral vertebral arteries |
| 433.21 | Occlusion and stenosis of vertebral artery with cerebral infarction | I63.119 | Cerebral infarction due to embolism of unspecified vertebral artery |
| 433.21 | Occlusion and stenosis of vertebral artery with cerebral infarction | I63.211 | Cerebral infarction due to unspecified occlusion or stenosis of right vertebral artery |
| 433.21 | Occlusion and stenosis of vertebral artery with cerebral infarction | I63.212 | Cerebral infarction due to unspecified occlusion or stenosis of left vertebral artery |
| 433.21 | Occlusion and stenosis of vertebral artery with cerebral infarction | I63.213 | Cerebral infarction due to unspecified occlusion or stenosis of bilateral vertebral arteries |
| 433.21 | Occlusion and stenosis of vertebral artery with cerebral infarction | I63.219 | Cerebral infarction due to unspecified occlusion or stenosis of unspecified vertebral arteries |
| 433.31 | Occlusion and stenosis of multiple and bilateral precerebral arteries with cerebral infarction | I63.59 | Cerebral infarction due to unspecified occlusion or stenosis of other cerebral artery |
| 433.81 | Occlusion and stenosis of other specified precerebral artery with cerebral infarction | I63.09 | Cerebral infarction due to thrombosis of other precerebral artery |
| 433.81 | Occlusion and stenosis of other specified precerebral artery with cerebral infarction | I63.19 | Cerebral infarction due to embolism of other precerebral artery |
| 433.81 | Occlusion and stenosis of other specified precerebral artery with cerebral infarction | I63.59 | Cerebral infarction due to unspecified occlusion or stenosis of other cerebral artery |
| 433.91 | Occlusion and stenosis of unspecified precerebral artery with cerebral infarction | I63.00 | Cerebral infarction due to thrombosis of unspecified precerebral artery |
| 433.91 | Occlusion and stenosis of unspecified precerebral artery with cerebral infarction | I63.10 | Cerebral infarction due to embolism of unspecified precerebral artery |
| 433.91 | Occlusion and stenosis of unspecified precerebral artery with cerebral infarction | I63.20 | Cerebral infarction due to unspecified occlusion or stenosis of unspecified precerebral arteries |
| 433.91 | Occlusion and stenosis of unspecified precerebral artery with cerebral infarction | I63.29 | Cerebral infarction due to unspecified occlusion or stenosis of other precerebral arteries |
| 434.00 | Cerebral thrombosis without mention of cerebral infarction |  |  |
| 434.01 | Cerebral thrombosis with cerebral infarction | I63.30 | Cerebral infarction due to thrombosis of unspecified cerebral artery |
| 434.01 | Cerebral thrombosis with cerebral infarction | I63.311 | Cerebral infarction due to thrombosis of right middle cerebral artery |
| 434.01 | Cerebral thrombosis with cerebral infarction | I63.312 | Cerebral infarction due to thrombosis of left middle cerebral artery |
| 434.01 | Cerebral thrombosis with cerebral infarction | I63.313 | Cerebral infarction due to thrombosis of bilateral middle cerebral arteries |
| 434.01 | Cerebral thrombosis with cerebral infarction | I63.319 | Cerebral infarction due to thrombosis of unspecified middle cerebral artery |
| 434.01 | Cerebral thrombosis with cerebral infarction | I63.321 | Cerebral infarction due to thrombosis of right anterior cerebral artery |
| 434.01 | Cerebral thrombosis with cerebral infarction | I63.322 | Cerebral infarction due to thrombosis of left anterior cerebral artery |
| 434.01 | Cerebral thrombosis with cerebral infarction | I63.323 | Cerebral infarction due to thrombosis of bilateral anterior cerebral arteries |
| 434.01 | Cerebral thrombosis with cerebral infarction | I63.329 | Cerebral infarction due to thrombosis of unspecified anterior cerebral artery |
| 434.01 | Cerebral thrombosis with cerebral infarction | I63.331 | Cerebral infarction due to thrombosis of right posterior cerebral artery |
| 434.01 | Cerebral thrombosis with cerebral infarction | I63.332 | Cerebral infarction due to thrombosis of left posterior cerebral artery |
| 434.01 | Cerebral thrombosis with cerebral infarction | I63.333 | Cerebral infarction to thrombosis of bilateral posterior cerebral arteries |
| 434.01 | Cerebral thrombosis with cerebral infarction | I63.339 | Cerebral infarction due to thrombosis of unspecified posterior cerebral artery |
| 434.01 | Cerebral thrombosis with cerebral infarction | I63.341 | Cerebral infarction due to thrombosis of right cerebellar artery |
| 434.01 | Cerebral thrombosis with cerebral infarction | I63.342 | Cerebral infarction due to thrombosis of left cerebellar artery |
| 434.01 | Cerebral thrombosis with cerebral infarction | I63.343 | Cerebral infarction to thrombosis of bilateral cerebellar arteries |
| 434.01 | Cerebral thrombosis with cerebral infarction | I63.349 | Cerebral infarction due to thrombosis of unspecified cerebellar artery |
| 434.01 | Cerebral thrombosis with cerebral infarction | I63.39 | Cerebral infarction due to thrombosis of other cerebral artery |
| 434.01 | Cerebral thrombosis with cerebral infarction | I63.6 | Cerebral infarction due to cerebral venous thrombosis, nonpyogenic |
| 434.10 | Cerebral embolism without mention of cerebral infarction |  |  |
| 434.11 | Cerebral embolism with cerebral infarction | I63.40 | Cerebral infarction due to embolism of unspecified cerebral artery |
| 434.11 | Cerebral embolism with cerebral infarction | I63.411 | Cerebral infarction due to embolism of right middle cerebral artery |
| 434.11 | Cerebral embolism with cerebral infarction | I63.412 | Cerebral infarction due to embolism of left middle cerebral artery |
| 434.11 | Cerebral embolism with cerebral infarction | I63.413 | Cerebral infarction due to embolism of bilateral middle cerebral arteries |
| 434.11 | Cerebral embolism with cerebral infarction | I63.419 | Cerebral infarction due to embolism of unspecified middle cerebral artery |
| 434.11 | Cerebral embolism with cerebral infarction | I63.421 | Cerebral infarction due to embolism of right anterior cerebral artery |
| 434.11 | Cerebral embolism with cerebral infarction | I63.422 | Cerebral infarction due to embolism of left anterior cerebral artery |
| 434.11 | Cerebral embolism with cerebral infarction | I63.423 | Cerebral infarction due to embolism of bilateral anterior cerebral arteries |
| 434.11 | Cerebral embolism with cerebral infarction | I63.429 | Cerebral infarction due to embolism of unspecified anterior cerebral artery |
| 434.11 | Cerebral embolism with cerebral infarction | I63.431 | Cerebral infarction due to embolism of right posterior cerebral artery |
| 434.11 | Cerebral embolism with cerebral infarction | I63.432 | Cerebral infarction due to embolism of left posterior cerebral artery |
| 434.11 | Cerebral embolism with cerebral infarction | I63.433 | Cerebral infarction due to embolism of bilateral posterior cerebral arteries |
| 434.11 | Cerebral embolism with cerebral infarction | I63.439 | Cerebral infarction due to embolism of unspecified posterior cerebral artery |
| 434.11 | Cerebral embolism with cerebral infarction | I63.441 | Cerebral infarction due to embolism of right cerebellar artery |
| 434.11 | Cerebral embolism with cerebral infarction | I63.442 | Cerebral infarction due to embolism of left cerebellar artery |
| 434.11 | Cerebral embolism with cerebral infarction | I63.443 | Cerebral infarction due to embolism of bilateral cerebellar arteries |
| 434.11 | Cerebral embolism with cerebral infarction | I63.449 | Cerebral infarction due to embolism of unspecified cerebellar artery |
| 434.11 | Cerebral embolism with cerebral infarction | I63.49 | Cerebral infarction due to embolism of other cerebral artery |
| 434.91 | Cerebral artery occlusion, unspecified with cerebral infarction | I63.50 | Cerebral infarction due to unspecified occlusion or stenosis of unspecified cerebral artery |
| 434.91 | Cerebral artery occlusion, unspecified with cerebral infarction | I63.511 | Cerebral infarction due to unspecified occlusion or stenosis of right middle cerebral artery |
| 434.91 | Cerebral artery occlusion, unspecified with cerebral infarction | I63.512 | Cerebral infarction due to unspecified occlusion or stenosis of left middle cerebral artery |
| 434.91 | Cerebral artery occlusion, unspecified with cerebral infarction | I63.513 | Cerebral infarction due to unspecified occlusion or stenosis of bilateral middle cerebral arteries |
| 434.91 | Cerebral artery occlusion, unspecified with cerebral infarction | I63.519 | Cerebral infarction due to unspecified occlusion or stenosis of unspecified middle cerebral artery |
| 434.91 | Cerebral artery occlusion, unspecified with cerebral infarction | I63.521 | Cerebral infarction due to unspecified occlusion or stenosis of right anterior cerebral artery |
| 434.91 | Cerebral artery occlusion, unspecified with cerebral infarction | I63.522 | Cerebral infarction due to unspecified occlusion or stenosis of left anterior cerebral artery |
| 434.91 | Cerebral artery occlusion, unspecified with cerebral infarction | I63.523 | Cerebral infarction due to unspecified occlusion or stenosis of bilateral anterior cerebral arteries |
| 434.91 | Cerebral artery occlusion, unspecified with cerebral infarction | I63.529 | Cerebral infarction due to unspecified occlusion or stenosis of unspecified anterior cerebral artery |
| 434.91 | Cerebral artery occlusion, unspecified with cerebral infarction | I63.531 | Cerebral infarction due to unspecified occlusion or stenosis of right posterior cerebral artery |
| 434.91 | Cerebral artery occlusion, unspecified with cerebral infarction | I63.532 | Cerebral infarction due to unspecified occlusion or stenosis of left posterior cerebral artery |
| 434.91 | Cerebral artery occlusion, unspecified with cerebral infarction | I63.533 | Cerebral infarction due to unspecified occlusion or stenosis of bilateral posterior cerebral arteries |
| 434.91 | Cerebral artery occlusion, unspecified with cerebral infarction | I63.539 | Cerebral infarction due to unspecified occlusion or stenosis of unspecified posterior cerebral artery |
| 434.91 | Cerebral artery occlusion, unspecified with cerebral infarction | I63.541 | Cerebral infarction due to unspecified occlusion or stenosis of right cerebellar artery |
| 434.91 | Cerebral artery occlusion, unspecified with cerebral infarction | I63.542 | Cerebral infarction due to unspecified occlusion or stenosis of left cerebellar artery |
| 434.91 | Cerebral artery occlusion, unspecified with cerebral infarction | I63.543 | Cerebral infarction due to unspecified occlusion or stenosis of bilateral cerebellar arteries |
| 434.91 | Cerebral artery occlusion, unspecified with cerebral infarction | I63.549 | Cerebral infarction due to unspecified occlusion or stenosis of unspecified cerebellar artery |
| 434.91 | Cerebral artery occlusion, unspecified with cerebral infarction | I63.59 | Cerebral infarction due to unspecified occlusion or stenosis of other cerebral artery |
| 434.91 | Cerebral artery occlusion, unspecified with cerebral infarction | I63.8 | Other cerebral infarction |
| 434.91 | Cerebral artery occlusion, unspecified with cerebral infarction | I63.9 | Cerebral infarction, unspecified |
| 435.8 | Other specified transient cerebral ischemias | G45.1 | Carotid artery syndrome (hemispheric) |
| 435.8 | Other specified transient cerebral ischemias | G45.2 | Multiple and bilateral precerebral artery syndromes |
| 435.8 | Other specified transient cerebral ischemias | G45.8 | Other transient cerebral ischemic attacks and related syndromes |
| 435.8 | Other specified transient cerebral ischemias | G46.0 | Middle cerebral artery syndrome |
| 435.8 | Other specified transient cerebral ischemias | G46.1 | Anterior cerebral artery syndrome |
| 435.8 | Other specified transient cerebral ischemias | G46.2 | Posterior cerebral artery syndrome |
| 435.9 | Unspecified transient cerebral ischemia | G45.9 | Transient cerebral ischemic attack, unspecified |
| 435.9 | Unspecified transient cerebral ischemia | I67.841 | Reversible cerebrovascular vasoconstriction syndrome |
| 435.9 | Unspecified transient cerebral ischemia |  |  |
| 436. | Acute, but ill-defined, cerebrovascular disease |  |  |
| 437.1 | Other generalized ischemic cerebrovascular disease | I67.82 | Cerebral ischemia |
| 437.1 | Other generalized ischemic cerebrovascular disease | I67.89 | Other cerebrovascular disease |
| 437.8 | Other ill-defined cerebrovascular disease | G46.3 | Brain stem stroke syndrome |
| 437.8 | Other ill-defined cerebrovascular disease | G46.4 | Cerebellar stroke syndrome |
| 437.8 | Other ill-defined cerebrovascular disease | G46.5 | Pure motor lacunar syndrome |
| 437.8 | Other ill-defined cerebrovascular disease | G46.6 | Pure sensory lacunar syndrome |
| 437.8 | Other ill-defined cerebrovascular disease | G46.7 | Other lacunar syndromes |
| 437.8 | Other ill-defined cerebrovascular disease | G46.8 | Other vascular syndromes of brain in cerebrovascular diseases |
| 437.8 | Other ill-defined cerebrovascular disease | I67.89 | Other cerebrovascular disease |
| 438.0 | Late effects of cerebrovascular disease, cognitive deficits | I69.010 | Attention and concentration deficit following nontraumatic subarachnoid hemorrhage |
| 438.0 | Late effects of cerebrovascular disease, cognitive deficits | I69.011 | Memory deficit following nontraumatic subarachnoid hemorrhage |
| 438.0 | Late effects of cerebrovascular disease, cognitive deficits | I69.012 | Visuospatial deficit and spatial neglect following nontraumatic subarachnoid hemorrhage |
| 438.0 | Late effects of cerebrovascular disease, cognitive deficits | I69.013 | Psychomotor deficit following nontraumatic subarachnoid hemorrhage |
| 438.0 | Late effects of cerebrovascular disease, cognitive deficits | I69.014 | Frontal lobe and executive function deficit following nontraumatic subarachnoid hemorrhage |
| 438.0 | Late effects of cerebrovascular disease, cognitive deficits | I69.015 | Cognitive social or emotional deficit following nontraumatic subarachnoid hemorrhage |
| 438.0 | Late effects of cerebrovascular disease, cognitive deficits | I69.018 | Other symptoms and signs involving cognitive functions following nontraumatic subarachnoid hemorrhage |
| 438.0 | Late effects of cerebrovascular disease, cognitive deficits | I69.019 | Unspecified symptoms and signs involving cognitive functions following nontraumatic subarachnoid hemorrhage |
| 438.0 | Late effects of cerebrovascular disease, cognitive deficits | I69.110 | Attention and concentration deficit following nontraumatic intracerebral hemorrhage |
| 438.0 | Late effects of cerebrovascular disease, cognitive deficits | I69.111 | Memory deficit following nontraumatic intracerebral hemorrhage |
| 438.0 | Late effects of cerebrovascular disease, cognitive deficits | I69.112 | Visuospatial deficit and spatial neglect following nontraumatic intracerebral hemorrhage |
| 438.0 | Late effects of cerebrovascular disease, cognitive deficits | I69.113 | Psychomotor deficit following nontraumatic intracerebral hemorrhage |
| 438.0 | Late effects of cerebrovascular disease, cognitive deficits | I69.114 | Frontal lobe and executive function deficit following nontraumatic intracerebral hemorrhage |
| 438.0 | Late effects of cerebrovascular disease, cognitive deficits | I69.115 | Cognitive social or emotional deficit following nontraumatic intracerebral hemorrhage |
| 438.0 | Late effects of cerebrovascular disease, cognitive deficits | I69.118 | Other symptoms and signs involving cognitive functions following nontraumatic intracerebral hemorrhage |
| 438.0 | Late effects of cerebrovascular disease, cognitive deficits | I69.119 | Unspecified symptoms and signs involving cognitive functions following nontraumatic intracerebral hemorrhage |
| 438.0 | Late effects of cerebrovascular disease, cognitive deficits | I69.210 | Attention and concentration deficit following other nontraumatic intracranial hemorrhage |
| 438.0 | Late effects of cerebrovascular disease, cognitive deficits | I69.211 | Memory deficit following other nontraumatic intracranial hemorrhage |
| 438.0 | Late effects of cerebrovascular disease, cognitive deficits | I69.212 | Visuospatial deficit and spatial neglect following other nontraumatic intracranial hemorrhage |
| 438.0 | Late effects of cerebrovascular disease, cognitive deficits | I69.213 | Psychomotor deficit following other nontraumatic intracranial hemorrhage |
| 438.0 | Late effects of cerebrovascular disease, cognitive deficits | I69.214 | Frontal lobe and executive function deficit following other nontraumatic intracranial hemorrhage |
| 438.0 | Late effects of cerebrovascular disease, cognitive deficits | I69.215 | Cognitive social or emotional deficit following other nontraumatic intracranial hemorrhage |
| 438.0 | Late effects of cerebrovascular disease, cognitive deficits | I69.218 | Other symptoms and signs involving cognitive functions following other nontraumatic intracranial hemorrhage |
| 438.0 | Late effects of cerebrovascular disease, cognitive deficits | I69.219 | Unspecified symptoms and signs involving cognitive functions following other nontraumatic intracranial hemorrhage |
| 438.0 | Late effects of cerebrovascular disease, cognitive deficits | I69.310 | Attention and concentration deficit following cerebral infarction |
| 438.0 | Late effects of cerebrovascular disease, cognitive deficits | I69.311 | Memory deficit following cerebral infarction |
| 438.0 | Late effects of cerebrovascular disease, cognitive deficits | I69.312 | Visuospatial deficit and spatial neglect following cerebral infarction |
| 438.0 | Late effects of cerebrovascular disease, cognitive deficits | I69.313 | Psychomotor deficit following cerebral infarction |
| 438.0 | Late effects of cerebrovascular disease, cognitive deficits | I69.314 | Frontal lobe and executive function deficit following cerebral infarction |
| 438.0 | Late effects of cerebrovascular disease, cognitive deficits | I69.315 | Cognitive social or emotional deficit following cerebral infarction |
| 438.0 | Late effects of cerebrovascular disease, cognitive deficits | I69.318 | Other symptoms and signs involving cognitive functions following cerebral infarction |
| 438.0 | Late effects of cerebrovascular disease, cognitive deficits | I69.319 | Unspecified symptoms and signs involving cognitive functions following cerebral infarction |
| 438.0 | Late effects of cerebrovascular disease, cognitive deficits | I69.810 | Attention and concentration deficit following other cerebrovascular disease |
| 438.0 | Late effects of cerebrovascular disease, cognitive deficits | I69.811 | Memory deficit following other cerebrovascular disease |
| 438.0 | Late effects of cerebrovascular disease, cognitive deficits | I69.812 | Visuospatial deficit and spatial neglect following other cerebrovascular disease |
| 438.0 | Late effects of cerebrovascular disease, cognitive deficits | I69.813 | Psychomotor deficit following other cerebrovascular disease |
| 438.0 | Late effects of cerebrovascular disease, cognitive deficits | I69.814 | Frontal lobe and executive function deficit following other cerebrovascular disease |
| 438.0 | Late effects of cerebrovascular disease, cognitive deficits | I69.815 | Cognitive social or emotional deficit following other cerebrovascular disease |
| 438.0 | Late effects of cerebrovascular disease, cognitive deficits | I69.818 | Other symptoms and signs involving cognitive functions following other cerebrovascular disease |
| 438.0 | Late effects of cerebrovascular disease, cognitive deficits | I69.819 | Unspecified symptoms and signs involving cognitive functions following other cerebrovascular disease |
| 438.0 | Late effects of cerebrovascular disease, cognitive deficits | I69.910 | Attention and concentration deficit following unspecified cerebrovascular disease |
| 438.0 | Late effects of cerebrovascular disease, cognitive deficits | I69.911 | Memory deficit following unspecified cerebrovascular disease |
| 438.0 | Late effects of cerebrovascular disease, cognitive deficits | I69.912 | Visuospatial deficit and spatial neglect following unspecified cerebrovascular disease |
| 438.0 | Late effects of cerebrovascular disease, cognitive deficits | I69.913 | Psychomotor deficit following unspecified cerebrovascular disease |
| 438.0 | Late effects of cerebrovascular disease, cognitive deficits | I69.914 | Frontal lobe and executive function deficit following unspecified cerebrovascular disease |
| 438.0 | Late effects of cerebrovascular disease, cognitive deficits | I69.915 | Cognitive social or emotional deficit following unspecified cerebrovascular disease |
| 438.0 | Late effects of cerebrovascular disease, cognitive deficits | I69.918 | Other symptoms and signs involving cognitive functions following unspecified cerebrovascular disease |
| 438.0 | Late effects of cerebrovascular disease, cognitive deficits | I69.919 | Unspecified symptoms and signs involving cognitive functions following unspecified cerebrovascular disease |
| 438.10 | Late effects of cerebrovascular disease, speech and language deficit, unspecified | I69.928 | Other speech and language deficits following unspecified cerebrovascular disease |
| 438.11 | Late effects of cerebrovascular disease, aphasia | I69.020 | Aphasia following nontraumatic subarachnoid hemorrhage |
| 438.11 | Late effects of cerebrovascular disease, aphasia | I69.120 | Aphasia following nontraumatic intracerebral hemorrhage |
| 438.11 | Late effects of cerebrovascular disease, aphasia | I69.220 | Aphasia following other nontraumatic intracranial hemorrhage |
| 438.11 | Late effects of cerebrovascular disease, aphasia | I69.320 | Aphasia following cerebral infarction |
| 438.11 | Late effects of cerebrovascular disease, aphasia | I69.820 | Aphasia following other cerebrovascular disease |
| 438.11 | Late effects of cerebrovascular disease, aphasia | I69.920 | Aphasia following unspecified cerebrovascular disease |
| 438.12 | Late effects of cerebrovascular disease, dysphasia | I69.021 | Dysphasia following nontraumatic subarachnoid hemorrhage |
| 438.12 | Late effects of cerebrovascular disease, dysphasia | I69.121 | Dysphasia following nontraumatic intracerebral hemorrhage |
| 438.12 | Late effects of cerebrovascular disease, dysphasia | I69.221 | Dysphasia following other nontraumatic intracranial hemorrhage |
| 438.12 | Late effects of cerebrovascular disease, dysphasia | I69.321 | Dysphasia following cerebral infarction |
| 438.12 | Late effects of cerebrovascular disease, dysphasia | I69.821 | Dysphasia following other cerebrovascular disease |
| 438.12 | Late effects of cerebrovascular disease, dysphasia | I69.921 | Dysphasia following unspecified cerebrovascular disease |
| 438.13 | Late effects of cerebrovascular disease, dysarthria | I69.022 | Dysarthria following nontraumatic subarachnoid hemorrhage |
| 438.13 | Late effects of cerebrovascular disease, dysarthria | I69.122 | Dysarthria following nontraumatic intracerebral hemorrhage |
| 438.13 | Late effects of cerebrovascular disease, dysarthria | I69.222 | Dysarthria following other nontraumatic intracranial hemorrhage |
| 438.13 | Late effects of cerebrovascular disease, dysarthria | I69.322 | Dysarthria following cerebral infarction |
| 438.13 | Late effects of cerebrovascular disease, dysarthria | I69.822 | Dysarthria following other cerebrovascular disease |
| 438.13 | Late effects of cerebrovascular disease, dysarthria | I69.922 | Dysarthria following unspecified cerebrovascular disease |
| 438.14 | Late effects of cerebrovascular disease, fluency disorder | I69.023 | Fluency disorder following nontraumatic subarachnoid hemorrhage |
| 438.14 | Late effects of cerebrovascular disease, fluency disorder | I69.123 | Fluency disorder following nontraumatic intracerebral hemorrhage |
| 438.14 | Late effects of cerebrovascular disease, fluency disorder | I69.223 | Fluency disorder following other nontraumatic intracranial hemorrhage |
| 438.14 | Late effects of cerebrovascular disease, fluency disorder | I69.323 | Fluency disorder following cerebral infarction |
| 438.14 | Late effects of cerebrovascular disease, fluency disorder | I69.823 | Fluency disorder following other cerebrovascular disease |
| 438.14 | Late effects of cerebrovascular disease, fluency disorder | I69.923 | Fluency disorder following unspecified cerebrovascular disease |
| 438.19 | Late effects of cerebrovascular disease, other speech and language deficits | I69.028 | Other speech and language deficits following nontraumatic subarachnoid hemorrhage |
| 438.19 | Late effects of cerebrovascular disease, other speech and language deficits | I69.128 | Other speech and language deficits following nontraumatic intracerebral hemorrhage |
| 438.19 | Late effects of cerebrovascular disease, other speech and language deficits | I69.228 | Other speech and language deficits following other nontraumatic intracranial hemorrhage |
| 438.19 | Late effects of cerebrovascular disease, other speech and language deficits | I69.328 | Other speech and language deficits following cerebral infarction |
| 438.19 | Late effects of cerebrovascular disease, other speech and language deficits | I69.828 | Other speech and language deficits following other cerebrovascular disease |
| 438.19 | Late effects of cerebrovascular disease, other speech and language deficits | I69.928 | Other speech and language deficits following unspecified cerebrovascular disease |
| 438.20 | Late effects of cerebrovascular disease, hemiplegia affecting unspecified side | I69.059 | Hemiplegia and hemiparesis following nontraumatic subarachnoid hemorrhage affecting unspecified side |
| 438.20 | Late effects of cerebrovascular disease, hemiplegia affecting unspecified side | I69.159 | Hemiplegia and hemiparesis following nontraumatic intracerebral hemorrhage affecting unspecified side |
| 438.20 | Late effects of cerebrovascular disease, hemiplegia affecting unspecified side | I69.259 | Hemiplegia and hemiparesis following other nontraumatic intracranial hemorrhage affecting unspecified side |
| 438.20 | Late effects of cerebrovascular disease, hemiplegia affecting unspecified side | I69.359 | Hemiplegia and hemiparesis following cerebral infarction affecting unspecified side |
| 438.20 | Late effects of cerebrovascular disease, hemiplegia affecting unspecified side | I69.859 | Hemiplegia and hemiparesis following other cerebrovascular disease affecting unspecified side |
| 438.20 | Late effects of cerebrovascular disease, hemiplegia affecting unspecified side | I69.959 | Hemiplegia and hemiparesis following unspecified cerebrovascular disease affecting unspecified side |
| 438.21 | Late effects of cerebrovascular disease, hemiplegia affecting dominant side | I69.051 | Hemiplegia and hemiparesis following nontraumatic subarachnoid hemorrhage affecting right dominant side |
| 438.21 | Late effects of cerebrovascular disease, hemiplegia affecting dominant side | I69.052 | Hemiplegia and hemiparesis following nontraumatic subarachnoid hemorrhage affecting left dominant side |
| 438.21 | Late effects of cerebrovascular disease, hemiplegia affecting dominant side | I69.151 | Hemiplegia and hemiparesis following nontraumatic intracerebral hemorrhage affecting right dominant side |
| 438.21 | Late effects of cerebrovascular disease, hemiplegia affecting dominant side | I69.152 | Hemiplegia and hemiparesis following nontraumatic intracerebral hemorrhage affecting left dominant side |
| 438.21 | Late effects of cerebrovascular disease, hemiplegia affecting dominant side | I69.251 | Hemiplegia and hemiparesis following other nontraumatic intracranial hemorrhage affecting right dominant side |
| 438.21 | Late effects of cerebrovascular disease, hemiplegia affecting dominant side | I69.252 | Hemiplegia and hemiparesis following other nontraumatic intracranial hemorrhage affecting left dominant side |
| 438.21 | Late effects of cerebrovascular disease, hemiplegia affecting dominant side | I69.351 | Hemiplegia and hemiparesis following cerebral infarction affecting right dominant side |
| 438.21 | Late effects of cerebrovascular disease, hemiplegia affecting dominant side | I69.352 | Hemiplegia and hemiparesis following cerebral infarction affecting left dominant side |
| 438.21 | Late effects of cerebrovascular disease, hemiplegia affecting dominant side | I69.851 | Hemiplegia and hemiparesis following other cerebrovascular disease affecting right dominant side |
| 438.21 | Late effects of cerebrovascular disease, hemiplegia affecting dominant side | I69.852 | Hemiplegia and hemiparesis following other cerebrovascular disease affecting left dominant side |
| 438.21 | Late effects of cerebrovascular disease, hemiplegia affecting dominant side | I69.951 | Hemiplegia and hemiparesis following unspecified cerebrovascular disease affecting right dominant side |
| 438.21 | Late effects of cerebrovascular disease, hemiplegia affecting dominant side | I69.952 | Hemiplegia and hemiparesis following unspecified cerebrovascular disease affecting left dominant side |
| 438.22 | Late effects of cerebrovascular disease, hemiplegia affecting nondominant side | I69.053 | Hemiplegia and hemiparesis following nontraumatic subarachnoid hemorrhage affecting right non-dominant side |
| 438.22 | Late effects of cerebrovascular disease, hemiplegia affecting nondominant side | I69.054 | Hemiplegia and hemiparesis following nontraumatic subarachnoid hemorrhage affecting left non-dominant side |
| 438.22 | Late effects of cerebrovascular disease, hemiplegia affecting nondominant side | I69.153 | Hemiplegia and hemiparesis following nontraumatic intracerebral hemorrhage affecting right non-dominant side |
| 438.22 | Late effects of cerebrovascular disease, hemiplegia affecting nondominant side | I69.154 | Hemiplegia and hemiparesis following nontraumatic intracerebral hemorrhage affecting left non-dominant side |
| 438.22 | Late effects of cerebrovascular disease, hemiplegia affecting nondominant side | I69.253 | Hemiplegia and hemiparesis following other nontraumatic intracranial hemorrhage affecting right non-dominant side |
| 438.22 | Late effects of cerebrovascular disease, hemiplegia affecting nondominant side | I69.254 | Hemiplegia and hemiparesis following other nontraumatic intracranial hemorrhage affecting left non-dominant side |
| 438.22 | Late effects of cerebrovascular disease, hemiplegia affecting nondominant side | I69.353 | Hemiplegia and hemiparesis following cerebral infarction affecting right non-dominant side |
| 438.22 | Late effects of cerebrovascular disease, hemiplegia affecting nondominant side | I69.354 | Hemiplegia and hemiparesis following cerebral infarction affecting left non-dominant side |
| 438.22 | Late effects of cerebrovascular disease, hemiplegia affecting nondominant side | I69.853 | Hemiplegia and hemiparesis following other cerebrovascular disease affecting right non-dominant side |
| 438.22 | Late effects of cerebrovascular disease, hemiplegia affecting nondominant side | I69.854 | Hemiplegia and hemiparesis following other cerebrovascular disease affecting left non-dominant side |
| 438.22 | Late effects of cerebrovascular disease, hemiplegia affecting nondominant side | I69.953 | Hemiplegia and hemiparesis following unspecified cerebrovascular disease affecting right non-dominant side |
| 438.22 | Late effects of cerebrovascular disease, hemiplegia affecting nondominant side | I69.954 | Hemiplegia and hemiparesis following unspecified cerebrovascular disease affecting left non-dominant side |
| 438.30 | Late effects of cerebrovascular disease, monoplegia of upper limb affecting unspecified side | I69.039 | Monoplegia of upper limb following nontraumatic subarachnoid hemorrhage affecting unspecified side |
| 438.30 | Late effects of cerebrovascular disease, monoplegia of upper limb affecting unspecified side | I69.139 | Monoplegia of upper limb following nontraumatic intracerebral hemorrhage affecting unspecified side |
| 438.30 | Late effects of cerebrovascular disease, monoplegia of upper limb affecting unspecified side | I69.239 | Monoplegia of upper limb following other nontraumatic intracranial hemorrhage affecting unspecified side |
| 438.30 | Late effects of cerebrovascular disease, monoplegia of upper limb affecting unspecified side | I69.339 | Monoplegia of upper limb following cerebral infarction affecting unspecified side |
| 438.30 | Late effects of cerebrovascular disease, monoplegia of upper limb affecting unspecified side | I69.839 | Monoplegia of upper limb following other cerebrovascular disease affecting unspecified side |
| 438.30 | Late effects of cerebrovascular disease, monoplegia of upper limb affecting unspecified side | I69.939 | Monoplegia of upper limb following unspecified cerebrovascular disease affecting unspecified side |
| 438.31 | Late effects of cerebrovascular disease, monoplegia of upper limb affecting dominant side | I69.031 | Monoplegia of upper limb following nontraumatic subarachnoid hemorrhage affecting right dominant side |
| 438.31 | Late effects of cerebrovascular disease, monoplegia of upper limb affecting dominant side | I69.032 | Monoplegia of upper limb following nontraumatic subarachnoid hemorrhage affecting left dominant side |
| 438.31 | Late effects of cerebrovascular disease, monoplegia of upper limb affecting dominant side | I69.131 | Monoplegia of upper limb following nontraumatic intracerebral hemorrhage affecting right dominant side |
| 438.31 | Late effects of cerebrovascular disease, monoplegia of upper limb affecting dominant side | I69.132 | Monoplegia of upper limb following nontraumatic intracerebral hemorrhage affecting left dominant side |
| 438.31 | Late effects of cerebrovascular disease, monoplegia of upper limb affecting dominant side | I69.231 | Monoplegia of upper limb following other nontraumatic intracranial hemorrhage affecting right dominant side |
| 438.31 | Late effects of cerebrovascular disease, monoplegia of upper limb affecting dominant side | I69.232 | Monoplegia of upper limb following other nontraumatic intracranial hemorrhage affecting left dominant side |
| 438.31 | Late effects of cerebrovascular disease, monoplegia of upper limb affecting dominant side | I69.331 | Monoplegia of upper limb following cerebral infarction affecting right dominant side |
| 438.31 | Late effects of cerebrovascular disease, monoplegia of upper limb affecting dominant side | I69.332 | Monoplegia of upper limb following cerebral infarction affecting left dominant side |
| 438.31 | Late effects of cerebrovascular disease, monoplegia of upper limb affecting dominant side | I69.831 | Monoplegia of upper limb following other cerebrovascular disease affecting right dominant side |
| 438.31 | Late effects of cerebrovascular disease, monoplegia of upper limb affecting dominant side | I69.832 | Monoplegia of upper limb following other cerebrovascular disease affecting left dominant side |
| 438.31 | Late effects of cerebrovascular disease, monoplegia of upper limb affecting dominant side | I69.931 | Monoplegia of upper limb following unspecified cerebrovascular disease affecting right dominant side |
| 438.31 | Late effects of cerebrovascular disease, monoplegia of upper limb affecting dominant side | I69.932 | Monoplegia of upper limb following unspecified cerebrovascular disease affecting left dominant side |
| 438.32 | Late effects of cerebrovascular disease, monoplegia of upper limb affecting nondominant side | I69.033 | Monoplegia of upper limb following nontraumatic subarachnoid hemorrhage affecting right non-dominant side |
| 438.32 | Late effects of cerebrovascular disease, monoplegia of upper limb affecting nondominant side | I69.034 | Monoplegia of upper limb following nontraumatic subarachnoid hemorrhage affecting left non-dominant side |
| 438.32 | Late effects of cerebrovascular disease, monoplegia of upper limb affecting nondominant side | I69.133 | Monoplegia of upper limb following nontraumatic intracerebral hemorrhage affecting right non-dominant side |
| 438.32 | Late effects of cerebrovascular disease, monoplegia of upper limb affecting nondominant side | I69.134 | Monoplegia of upper limb following nontraumatic intracerebral hemorrhage affecting left non-dominant side |
| 438.32 | Late effects of cerebrovascular disease, monoplegia of upper limb affecting nondominant side | I69.233 | Monoplegia of upper limb following other nontraumatic intracranial hemorrhage affecting right non-dominant side |
| 438.32 | Late effects of cerebrovascular disease, monoplegia of upper limb affecting nondominant side | I69.234 | Monoplegia of upper limb following other nontraumatic intracranial hemorrhage affecting left non-dominant side |
| 438.32 | Late effects of cerebrovascular disease, monoplegia of upper limb affecting nondominant side | I69.333 | Monoplegia of upper limb following cerebral infarction affecting right non-dominant side |
| 438.32 | Late effects of cerebrovascular disease, monoplegia of upper limb affecting nondominant side | I69.334 | Monoplegia of upper limb following cerebral infarction affecting left non-dominant side |
| 438.32 | Late effects of cerebrovascular disease, monoplegia of upper limb affecting nondominant side | I69.833 | Monoplegia of upper limb following other cerebrovascular disease affecting right non-dominant side |
| 438.32 | Late effects of cerebrovascular disease, monoplegia of upper limb affecting nondominant side | I69.834 | Monoplegia of upper limb following other cerebrovascular disease affecting left non-dominant side |
| 438.32 | Late effects of cerebrovascular disease, monoplegia of upper limb affecting nondominant side | I69.933 | Monoplegia of upper limb following unspecified cerebrovascular disease affecting right non-dominant side |
| 438.32 | Late effects of cerebrovascular disease, monoplegia of upper limb affecting nondominant side | I69.934 | Monoplegia of upper limb following unspecified cerebrovascular disease affecting left non-dominant side |
| 438.40 | Late effects of cerebrovascular disease, monoplegia of lower limb affecting unspecified side | I69.049 | Monoplegia of lower limb following nontraumatic subarachnoid hemorrhage affecting unspecified side |
| 438.40 | Late effects of cerebrovascular disease, monoplegia of lower limb affecting unspecified side | I69.149 | Monoplegia of lower limb following nontraumatic intracerebral hemorrhage affecting unspecified side |
| 438.40 | Late effects of cerebrovascular disease, monoplegia of lower limb affecting unspecified side | I69.249 | Monoplegia of lower limb following other nontraumatic intracranial hemorrhage affecting unspecified side |
| 438.40 | Late effects of cerebrovascular disease, monoplegia of lower limb affecting unspecified side | I69.349 | Monoplegia of lower limb following cerebral infarction affecting unspecified side |
| 438.40 | Late effects of cerebrovascular disease, monoplegia of lower limb affecting unspecified side | I69.849 | Monoplegia of lower limb following other cerebrovascular disease affecting unspecified side |
| 438.40 | Late effects of cerebrovascular disease, monoplegia of lower limb affecting unspecified side | I69.949 | Monoplegia of lower limb following unspecified cerebrovascular disease affecting unspecified side |
| 438.41 | Late effects of cerebrovascular disease, monoplegia of lower limb affecting dominant side | I69.041 | Monoplegia of lower limb following nontraumatic subarachnoid hemorrhage affecting right dominant side |
| 438.41 | Late effects of cerebrovascular disease, monoplegia of lower limb affecting dominant side | I69.042 | Monoplegia of lower limb following nontraumatic subarachnoid hemorrhage affecting left dominant side |
| 438.41 | Late effects of cerebrovascular disease, monoplegia of lower limb affecting dominant side | I69.141 | Monoplegia of lower limb following nontraumatic intracerebral hemorrhage affecting right dominant side |
| 438.41 | Late effects of cerebrovascular disease, monoplegia of lower limb affecting dominant side | I69.142 | Monoplegia of lower limb following nontraumatic intracerebral hemorrhage affecting left dominant side |
| 438.41 | Late effects of cerebrovascular disease, monoplegia of lower limb affecting dominant side | I69.241 | Monoplegia of lower limb following other nontraumatic intracranial hemorrhage affecting right dominant side |
| 438.41 | Late effects of cerebrovascular disease, monoplegia of lower limb affecting dominant side | I69.242 | Monoplegia of lower limb following other nontraumatic intracranial hemorrhage affecting left dominant side |
| 438.41 | Late effects of cerebrovascular disease, monoplegia of lower limb affecting dominant side | I69.341 | Monoplegia of lower limb following cerebral infarction affecting right dominant side |
| 438.41 | Late effects of cerebrovascular disease, monoplegia of lower limb affecting dominant side | I69.342 | Monoplegia of lower limb following cerebral infarction affecting left dominant side |
| 438.41 | Late effects of cerebrovascular disease, monoplegia of lower limb affecting dominant side | I69.841 | Monoplegia of lower limb following other cerebrovascular disease affecting right dominant side |
| 438.41 | Late effects of cerebrovascular disease, monoplegia of lower limb affecting dominant side | I69.842 | Monoplegia of lower limb following other cerebrovascular disease affecting left dominant side |
| 438.41 | Late effects of cerebrovascular disease, monoplegia of lower limb affecting dominant side | I69.941 | Monoplegia of lower limb following unspecified cerebrovascular disease affecting right dominant side |
| 438.41 | Late effects of cerebrovascular disease, monoplegia of lower limb affecting dominant side | I69.942 | Monoplegia of lower limb following unspecified cerebrovascular disease affecting left dominant side |
| 438.42 | Late effects of cerebrovascular disease, monoplegia of lower limb affecting nondominant side | I69.043 | Monoplegia of lower limb following nontraumatic subarachnoid hemorrhage affecting right non-dominant side |
| 438.42 | Late effects of cerebrovascular disease, monoplegia of lower limb affecting nondominant side | I69.044 | Monoplegia of lower limb following nontraumatic subarachnoid hemorrhage affecting left non-dominant side |
| 438.42 | Late effects of cerebrovascular disease, monoplegia of lower limb affecting nondominant side | I69.143 | Monoplegia of lower limb following nontraumatic intracerebral hemorrhage affecting right non-dominant side |
| 438.42 | Late effects of cerebrovascular disease, monoplegia of lower limb affecting nondominant side | I69.144 | Monoplegia of lower limb following nontraumatic intracerebral hemorrhage affecting left non-dominant side |
| 438.42 | Late effects of cerebrovascular disease, monoplegia of lower limb affecting nondominant side | I69.243 | Monoplegia of lower limb following other nontraumatic intracranial hemorrhage affecting right non-dominant side |
| 438.42 | Late effects of cerebrovascular disease, monoplegia of lower limb affecting nondominant side | I69.244 | Monoplegia of lower limb following other nontraumatic intracranial hemorrhage affecting left non-dominant side |
| 438.42 | Late effects of cerebrovascular disease, monoplegia of lower limb affecting nondominant side | I69.343 | Monoplegia of lower limb following cerebral infarction affecting right non-dominant side |
| 438.42 | Late effects of cerebrovascular disease, monoplegia of lower limb affecting nondominant side | I69.344 | Monoplegia of lower limb following cerebral infarction affecting left non-dominant side |
| 438.42 | Late effects of cerebrovascular disease, monoplegia of lower limb affecting nondominant side | I69.843 | Monoplegia of lower limb following other cerebrovascular disease affecting right non-dominant side |
| 438.42 | Late effects of cerebrovascular disease, monoplegia of lower limb affecting nondominant side | I69.844 | Monoplegia of lower limb following other cerebrovascular disease affecting left non-dominant side |
| 438.42 | Late effects of cerebrovascular disease, monoplegia of lower limb affecting nondominant side | I69.943 | Monoplegia of lower limb following unspecified cerebrovascular disease affecting right non-dominant side |
| 438.42 | Late effects of cerebrovascular disease, monoplegia of lower limb affecting nondominant side | I69.944 | Monoplegia of lower limb following unspecified cerebrovascular disease affecting left non-dominant side |
| 438.50 | Late effects of cerebrovascular disease, other paralytic syndrome affecting unspecified side | I69.069 | Other paralytic syndrome following nontraumatic subarachnoid hemorrhage affecting unspecified side |
| 438.50 | Late effects of cerebrovascular disease, other paralytic syndrome affecting unspecified side | I69.169 | Other paralytic syndrome following nontraumatic intracerebral hemorrhage affecting unspecified side |
| 438.50 | Late effects of cerebrovascular disease, other paralytic syndrome affecting unspecified side | I69.269 | Other paralytic syndrome following other nontraumatic intracranial hemorrhage affecting unspecified side |
| 438.50 | Late effects of cerebrovascular disease, other paralytic syndrome affecting unspecified side | I69.369 | Other paralytic syndrome following cerebral infarction affecting unspecified side |
| 438.50 | Late effects of cerebrovascular disease, other paralytic syndrome affecting unspecified side | I69.869 | Other paralytic syndrome following other cerebrovascular disease affecting unspecified side |
| 438.50 | Late effects of cerebrovascular disease, other paralytic syndrome affecting unspecified side | I69.969 | Other paralytic syndrome following unspecified cerebrovascular disease affecting unspecified side |
| 438.51 | Late effects of cerebrovascular disease, other paralytic syndrome affecting dominant side | I69.061 | Other paralytic syndrome following nontraumatic subarachnoid hemorrhage affecting right dominant side |
| 438.51 | Late effects of cerebrovascular disease, other paralytic syndrome affecting dominant side | I69.062 | Other paralytic syndrome following nontraumatic subarachnoid hemorrhage affecting left dominant side |
| 438.51 | Late effects of cerebrovascular disease, other paralytic syndrome affecting dominant side | I69.161 | Other paralytic syndrome following nontraumatic intracerebral hemorrhage affecting right dominant side |
| 438.51 | Late effects of cerebrovascular disease, other paralytic syndrome affecting dominant side | I69.162 | Other paralytic syndrome following nontraumatic intracerebral hemorrhage affecting left dominant side |
| 438.51 | Late effects of cerebrovascular disease, other paralytic syndrome affecting dominant side | I69.261 | Other paralytic syndrome following other nontraumatic intracranial hemorrhage affecting right dominant side |
| 438.51 | Late effects of cerebrovascular disease, other paralytic syndrome affecting dominant side | I69.262 | Other paralytic syndrome following other nontraumatic intracranial hemorrhage affecting left dominant side |
| 438.51 | Late effects of cerebrovascular disease, other paralytic syndrome affecting dominant side | I69.361 | Other paralytic syndrome following cerebral infarction affecting right dominant side |
| 438.51 | Late effects of cerebrovascular disease, other paralytic syndrome affecting dominant side | I69.362 | Other paralytic syndrome following cerebral infarction affecting left dominant side |
| 438.51 | Late effects of cerebrovascular disease, other paralytic syndrome affecting dominant side | I69.861 | Other paralytic syndrome following other cerebrovascular disease affecting right dominant side |
| 438.51 | Late effects of cerebrovascular disease, other paralytic syndrome affecting dominant side | I69.862 | Other paralytic syndrome following other cerebrovascular disease affecting left dominant side |
| 438.51 | Late effects of cerebrovascular disease, other paralytic syndrome affecting dominant side | I69.961 | Other paralytic syndrome following unspecified cerebrovascular disease affecting right dominant side |
| 438.51 | Late effects of cerebrovascular disease, other paralytic syndrome affecting dominant side | I69.962 | Other paralytic syndrome following unspecified cerebrovascular disease affecting left dominant side |
| 438.52 | Late effects of cerebrovascular disease, other paralytic syndrome affecting nondominant side | I69.063 | Other paralytic syndrome following nontraumatic subarachnoid hemorrhage affecting right non-dominant side |
| 438.52 | Late effects of cerebrovascular disease, other paralytic syndrome affecting nondominant side | I69.064 | Other paralytic syndrome following nontraumatic subarachnoid hemorrhage affecting left non-dominant side |
| 438.52 | Late effects of cerebrovascular disease, other paralytic syndrome affecting nondominant side | I69.163 | Other paralytic syndrome following nontraumatic intracerebral hemorrhage affecting right non-dominant side |
| 438.52 | Late effects of cerebrovascular disease, other paralytic syndrome affecting nondominant side | I69.164 | Other paralytic syndrome following nontraumatic intracerebral hemorrhage affecting left non-dominant side |
| 438.52 | Late effects of cerebrovascular disease, other paralytic syndrome affecting nondominant side | I69.263 | Other paralytic syndrome following other nontraumatic intracranial hemorrhage affecting right non-dominant side |
| 438.52 | Late effects of cerebrovascular disease, other paralytic syndrome affecting nondominant side | I69.264 | Other paralytic syndrome following other nontraumatic intracranial hemorrhage affecting left non-dominant side |
| 438.52 | Late effects of cerebrovascular disease, other paralytic syndrome affecting nondominant side | I69.363 | Other paralytic syndrome following cerebral infarction affecting right non-dominant side |
| 438.52 | Late effects of cerebrovascular disease, other paralytic syndrome affecting nondominant side | I69.364 | Other paralytic syndrome following cerebral infarction affecting left non-dominant side |
| 438.52 | Late effects of cerebrovascular disease, other paralytic syndrome affecting nondominant side | I69.863 | Other paralytic syndrome following other cerebrovascular disease affecting right non-dominant side |
| 438.52 | Late effects of cerebrovascular disease, other paralytic syndrome affecting nondominant side | I69.864 | Other paralytic syndrome following other cerebrovascular disease affecting left non-dominant side |
| 438.52 | Late effects of cerebrovascular disease, other paralytic syndrome affecting nondominant side | I69.963 | Other paralytic syndrome following unspecified cerebrovascular disease affecting right non-dominant side |
| 438.52 | Late effects of cerebrovascular disease, other paralytic syndrome affecting nondominant side | I69.964 | Other paralytic syndrome following unspecified cerebrovascular disease affecting left non-dominant side |
| 438.53 | Late effects of cerebrovascular disease, other paralytic syndrome, bilateral | I69.065 | Other paralytic syndrome following nontraumatic subarachnoid hemorrhage, bilateral |
| 438.53 | Late effects of cerebrovascular disease, other paralytic syndrome, bilateral | I69.165 | Other paralytic syndrome following nontraumatic intracerebral hemorrhage, bilateral |
| 438.53 | Late effects of cerebrovascular disease, other paralytic syndrome, bilateral | I69.265 | Other paralytic syndrome following other nontraumatic intracranial hemorrhage, bilateral |
| 438.53 | Late effects of cerebrovascular disease, other paralytic syndrome, bilateral | I69.365 | Other paralytic syndrome following cerebral infarction, bilateral |
| 438.53 | Late effects of cerebrovascular disease, other paralytic syndrome, bilateral | I69.865 | Other paralytic syndrome following other cerebrovascular disease, bilateral |
| 438.53 | Late effects of cerebrovascular disease, other paralytic syndrome, bilateral | I69.965 | Other paralytic syndrome following unspecified cerebrovascular disease, bilateral |
| 438.6 | Late effects of cerebrovascular disease, alterations of sensations | I69.998 | Other sequelae following unspecified cerebrovascular disease |
| 438.7 | Late effects of cerebrovascular disease, disturbances of vision | I69.998 | Other sequelae following unspecified cerebrovascular disease |
| 438.81 | Other late effects of cerebrovascular disease, apraxia | I69.090 | Apraxia following nontraumatic subarachnoid hemorrhage |
| 438.81 | Other late effects of cerebrovascular disease, apraxia | I69.190 | Apraxia following nontraumatic intracerebral hemorrhage |
| 438.81 | Other late effects of cerebrovascular disease, apraxia | I69.290 | Apraxia following other nontraumatic intracranial hemorrhage |
| 438.81 | Other late effects of cerebrovascular disease, apraxia | I69.390 | Apraxia following cerebral infarction |
| 438.81 | Other late effects of cerebrovascular disease, apraxia | I69.890 | Apraxia following other cerebrovascular disease |
| 438.81 | Other late effects of cerebrovascular disease, apraxia | I69.990 | Apraxia following unspecified cerebrovascular disease |
| 438.82 | Other late effects of cerebrovascular disease, dysphagia | I69.091 | Dysphagia following nontraumatic subarachnoid hemorrhage |
| 438.82 | Other late effects of cerebrovascular disease, dysphagia | I69.191 | Dysphagia following nontraumatic intracerebral hemorrhage |
| 438.82 | Other late effects of cerebrovascular disease, dysphagia | I69.291 | Dysphagia following other nontraumatic intracranial hemorrhage |
| 438.82 | Other late effects of cerebrovascular disease, dysphagia | I69.391 | Dysphagia following cerebral infarction |
| 438.82 | Other late effects of cerebrovascular disease, dysphagia | I69.891 | Dysphagia following other cerebrovascular disease |
| 438.82 | Other late effects of cerebrovascular disease, dysphagia | I69.991 | Dysphagia following unspecified cerebrovascular disease |
| 438.83 | Other late effects of cerebrovascular disease, facial weakness | I69.092 | Facial weakness following nontraumatic subarachnoid hemorrhage |
| 438.83 | Other late effects of cerebrovascular disease, facial weakness | I69.192 | Facial weakness following nontraumatic intracerebral hemorrhage |
| 438.83 | Other late effects of cerebrovascular disease, facial weakness | I69.292 | Facial weakness following other nontraumatic intracranial hemorrhage |
| 438.83 | Other late effects of cerebrovascular disease, facial weakness | I69.392 | Facial weakness following cerebral infarction |
| 438.83 | Other late effects of cerebrovascular disease, facial weakness | I69.892 | Facial weakness following other cerebrovascular disease |
| 438.83 | Other late effects of cerebrovascular disease, facial weakness | I69.992 | Facial weakness following unspecified cerebrovascular disease |
| 438.84 | Other late effects of cerebrovascular disease, ataxia | I69.093 | Ataxia following nontraumatic subarachnoid hemorrhage |
| 438.84 | Other late effects of cerebrovascular disease, ataxia | I69.193 | Ataxia following nontraumatic intracerebral hemorrhage |
| 438.84 | Other late effects of cerebrovascular disease, ataxia | I69.293 | Ataxia following other nontraumatic intracranial hemorrhage |
| 438.84 | Other late effects of cerebrovascular disease, ataxia | I69.393 | Ataxia following cerebral infarction |
| 438.84 | Other late effects of cerebrovascular disease, ataxia | I69.893 | Ataxia following other cerebrovascular disease |
| 438.84 | Other late effects of cerebrovascular disease, ataxia | I69.993 | Ataxia following unspecified cerebrovascular disease |
| 438.85 | Other late effects of cerebrovascular disease, vertigo | I69.998 | Other sequelae following unspecified cerebrovascular disease |
| 438.89 | Other late effects of cerebrovascular disease | I69.098 | Other sequelae following nontraumatic subarachnoid hemorrhage |
| 438.89 | Other late effects of cerebrovascular disease | I69.198 | Other sequelae of nontraumatic intracerebral hemorrhage |
| 438.89 | Other late effects of cerebrovascular disease | I69.298 | Other sequelae of other nontraumatic intracranial hemorrhage |
| 438.89 | Other late effects of cerebrovascular disease | I69.398 | Other sequelae of cerebral infarction |
| 438.89 | Other late effects of cerebrovascular disease | I69.898 | Other sequelae of other cerebrovascular disease |
| 438.89 | Other late effects of cerebrovascular disease | I69.998 | Other sequelae following unspecified cerebrovascular disease |
| 438.9 | Unspecified late effects of cerebrovascular disease | I69.00 | Unspecified sequelae of nontraumatic subarachnoid hemorrhage |
| 438.9 | Unspecified late effects of cerebrovascular disease | I69.10 | Unspecified sequelae of nontraumatic intracerebral hemorrhage |
| 438.9 | Unspecified late effects of cerebrovascular disease | I69.20 | Unspecified sequelae of other nontraumatic intracranial hemorrhage |
| 438.9 | Unspecified late effects of cerebrovascular disease | I69.30 | Unspecified sequelae of cerebral infarction |
| 438.9 | Unspecified late effects of cerebrovascular disease | I69.80 | Unspecified sequelae of other cerebrovascular disease |
| 438.9 | Unspecified late effects of cerebrovascular disease | I69.90 | Unspecified sequelae of unspecified cerebrovascular disease |
| 781.4 | Transient paralysis of limb | R29.5 | Transient paralysis |
| 781.8 | Neurologic neglect syndrome | R41.4 | Neurologic neglect syndrome |
| 781.94 | Facial weakness | R29.810 | Facial weakness |
| 784.3 | Aphasia | R47.01 | Aphasia |
| 784.51 | Dysarthria | R47.1 | Dysarthria and anarthria |
| 787.20 | Dysphagia, unspecified | R13.0 | Aphagia |
| 787.20 | Dysphagia, unspecified | R13.10 | Dysphagia, unspecified |
| 787.21 | Dysphagia, oral phase | R13.11 | Dysphagia, oral phase |
| 787.22 | Dysphagia, oropharyngeal phase | R13.12 | Dysphagia, oropharyngeal phase |
| 787.23 | Dysphagia, pharyngeal phase | R13.13 | Dysphagia, pharyngeal phase |
| 787.24 | Dysphagia, pharyngoesophageal phase | R13.14 | Dysphagia, pharyngoesophageal phase |
| 787.29 | Other dysphagia | R13.19 | Other dysphagia |
| 799.53 | Visuospatial deficit | R41.842 | Visuospatial deficit |
| 800.10 | Closed fracture of vault of skull with cerebral laceration and contusion, unspecified state of consciousness | S02.0XXA | Fracture of vault of skull, initial encounter for closed fracture |
| 800.10 | Closed fracture of vault of skull with cerebral laceration and contusion, unspecified state of consciousness | S06.330A | Contusion and laceration of cerebrum, unspecified, without loss of consciousness, initial encounter |
| 800.11 | Closed fracture of vault of skull with cerebral laceration and contusion, with no loss of consciousness | S02.0XXA | Fracture of vault of skull, initial encounter for closed fracture |
| 800.11 | Closed fracture of vault of skull with cerebral laceration and contusion, with no loss of consciousness | S06.330A | Contusion and laceration of cerebrum, unspecified, without loss of consciousness, initial encounter |
| 800.12 | Closed fracture of vault of skull with cerebral laceration and contusion, with brief [less than one hour] loss of consciousness | S02.0XXA | Fracture of vault of skull, initial encounter for closed fracture |
| 800.12 | Closed fracture of vault of skull with cerebral laceration and contusion, with brief [less than one hour] loss of consciousness | S06.331A | Contusion and laceration of cerebrum, unspecified, with loss of consciousness of 30 minutes or less, initial encounter |
| 800.12 | Closed fracture of vault of skull with cerebral laceration and contusion, with brief [less than one hour] loss of consciousness | S06.332A | Contusion and laceration of cerebrum, unspecified, with loss of consciousness of 31 minutes to 59 minutes, initial encounter |
| 800.13 | Closed fracture of vault of skull with cerebral laceration and contusion, with moderate [1-24 hours] loss of consciousness | S02.0XXA | Fracture of vault of skull, initial encounter for closed fracture |
| 800.13 | Closed fracture of vault of skull with cerebral laceration and contusion, with moderate [1-24 hours] loss of consciousness | S06.333A | Contusion and laceration of cerebrum, unspecified, with loss of consciousness of 1 hour to 5 hours 59 minutes, initial encounter |
| 800.13 | Closed fracture of vault of skull with cerebral laceration and contusion, with moderate [1-24 hours] loss of consciousness | S06.334A | Contusion and laceration of cerebrum, unspecified, with loss of consciousness of 6 hours to 24 hours, initial encounter |
| 800.14 | Closed fracture of vault of skull with cerebral laceration and contusion, with prolonged [more than 24 hours] loss of consciousness and return to pre-existing conscious level | S02.0XXA | Fracture of vault of skull, initial encounter for closed fracture |
| 800.14 | Closed fracture of vault of skull with cerebral laceration and contusion, with prolonged [more than 24 hours] loss of consciousness and return to pre-existing conscious level | S06.335A | Contusion and laceration of cerebrum, unspecified, with loss of consciousness greater than 24 hours with return to pre-existing conscious level, initial encounter |
| 800.15 | Closed fracture of vault of skull with cerebral laceration and contusion, with prolonged [more than 24 hours] loss of consciousness, without return to pre-existing conscious level | S02.0XXA | Fracture of vault of skull, initial encounter for closed fracture |
| 800.15 | Closed fracture of vault of skull with cerebral laceration and contusion, with prolonged [more than 24 hours] loss of consciousness, without return to pre-existing conscious level | S06.336A | Contusion and laceration of cerebrum, unspecified, with loss of consciousness greater than 24 hours without return to pre-existing conscious level with patient surviving, initial encounter |
| 800.15 | Closed fracture of vault of skull with cerebral laceration and contusion, with prolonged [more than 24 hours] loss of consciousness, without return to pre-existing conscious level | S06.337A | Contusion and laceration of cerebrum, unspecified, with loss of consciousness of any duration with death due to brain injury prior to regaining consciousness, initial encounter |
| 800.15 | Closed fracture of vault of skull with cerebral laceration and contusion, with prolonged [more than 24 hours] loss of consciousness, without return to pre-existing conscious level | S06.338A | Contusion and laceration of cerebrum, unspecified, with loss of consciousness of any duration with death due to other cause prior to regaining consciousness, initial encounter |
| 800.16 | Closed fracture of vault of skull with cerebral laceration and contusion, with loss of consciousness of unspecified duration | S02.0XXA | Fracture of vault of skull, initial encounter for closed fracture |
| 800.16 | Closed fracture of vault of skull with cerebral laceration and contusion, with loss of consciousness of unspecified duration | S06.339A | Contusion and laceration of cerebrum, unspecified, with loss of consciousness of unspecified duration, initial encounter |
| 800.19 | Closed fracture of vault of skull with cerebral laceration and contusion, with concussion, unspecified | S02.0XXA | Fracture of vault of skull, initial encounter for closed fracture |
| 800.19 | Closed fracture of vault of skull with cerebral laceration and contusion, with concussion, unspecified | S06.330A | Contusion and laceration of cerebrum, unspecified, without loss of consciousness, initial encounter |
| 800.20 | Closed fracture of vault of skull with subarachnoid, subdural, and extradural hemorrhage, unspecified state of consciousness | S06.4X0A | Epidural hemorrhage without loss of consciousness, initial encounter |
| 800.20 | Closed fracture of vault of skull with subarachnoid, subdural, and extradural hemorrhage, unspecified state of consciousness | S06.5X0A | Traumatic subdural hemorrhage without loss of consciousness, initial encounter |
| 800.20 | Closed fracture of vault of skull with subarachnoid, subdural, and extradural hemorrhage, unspecified state of consciousness | S06.6X0A | Traumatic subarachnoid hemorrhage without loss of consciousness, initial encounter |
| 800.20 | Closed fracture of vault of skull with subarachnoid, subdural, and extradural hemorrhage, unspecified state of consciousness |  |  |
| 800.21 | Closed fracture of vault of skull with subarachnoid, subdural, and extradural hemorrhage, with no loss of consciousness | S06.4X0A | Epidural hemorrhage without loss of consciousness, initial encounter |
| 800.21 | Closed fracture of vault of skull with subarachnoid, subdural, and extradural hemorrhage, with no loss of consciousness | S06.5X0A | Traumatic subdural hemorrhage without loss of consciousness, initial encounter |
| 800.21 | Closed fracture of vault of skull with subarachnoid, subdural, and extradural hemorrhage, with no loss of consciousness | S06.6X0A | Traumatic subarachnoid hemorrhage without loss of consciousness, initial encounter |
| 800.21 | Closed fracture of vault of skull with subarachnoid, subdural, and extradural hemorrhage, with no loss of consciousness |  |  |
| 800.22 | Closed fracture of vault of skull with subarachnoid, subdural, and extradural hemorrhage, with brief [less than one hour] loss of consciousness | S06.4X1A | Epidural hemorrhage with loss of consciousness of 30 minutes or less, initial encounter |
| 800.22 | Closed fracture of vault of skull with subarachnoid, subdural, and extradural hemorrhage, with brief [less than one hour] loss of consciousness | S06.4X2A | Epidural hemorrhage with loss of consciousness of 31 minutes to 59 minutes, initial encounter |
| 800.22 | Closed fracture of vault of skull with subarachnoid, subdural, and extradural hemorrhage, with brief [less than one hour] loss of consciousness | S06.5X1A | Traumatic subdural hemorrhage with loss of consciousness of 30 minutes or less, initial encounter |
| 800.22 | Closed fracture of vault of skull with subarachnoid, subdural, and extradural hemorrhage, with brief [less than one hour] loss of consciousness | S06.5X2A | Traumatic subdural hemorrhage with loss of consciousness of 31 minutes to 59 minutes, initial encounter |
| 800.22 | Closed fracture of vault of skull with subarachnoid, subdural, and extradural hemorrhage, with brief [less than one hour] loss of consciousness | S06.6X1A | Traumatic subarachnoid hemorrhage with loss of consciousness of 30 minutes or less, initial encounter |
| 800.22 | Closed fracture of vault of skull with subarachnoid, subdural, and extradural hemorrhage, with brief [less than one hour] loss of consciousness | S06.6X2A | Traumatic subarachnoid hemorrhage with loss of consciousness of 31 minutes to 59 minutes, initial encounter |
| 800.22 | Closed fracture of vault of skull with subarachnoid, subdural, and extradural hemorrhage, with brief [less than one hour] loss of consciousness |  |  |
| 800.23 | Closed fracture of vault of skull with subarachnoid, subdural, and extradural hemorrhage, with moderate [1-24 hours] loss of consciousness | S06.4X3A | Epidural hemorrhage with loss of consciousness of 1 hour to 5 hours 59 minutes, initial encounter |
| 800.23 | Closed fracture of vault of skull with subarachnoid, subdural, and extradural hemorrhage, with moderate [1-24 hours] loss of consciousness | S06.4X4A | Epidural hemorrhage with loss of consciousness of 6 hours to 24 hours, initial encounter |
| 800.23 | Closed fracture of vault of skull with subarachnoid, subdural, and extradural hemorrhage, with moderate [1-24 hours] loss of consciousness | S06.5X3A | Traumatic subdural hemorrhage with loss of consciousness of 1 hour to 5 hours 59 minutes, initial encounter |
| 800.23 | Closed fracture of vault of skull with subarachnoid, subdural, and extradural hemorrhage, with moderate [1-24 hours] loss of consciousness | S06.5X4A | Traumatic subdural hemorrhage with loss of consciousness of 6 hours to 24 hours, initial encounter |
| 800.23 | Closed fracture of vault of skull with subarachnoid, subdural, and extradural hemorrhage, with moderate [1-24 hours] loss of consciousness | S06.6X3A | Traumatic subarachnoid hemorrhage with loss of consciousness of 1 hour to 5 hours 59 minutes, initial encounter |
| 800.23 | Closed fracture of vault of skull with subarachnoid, subdural, and extradural hemorrhage, with moderate [1-24 hours] loss of consciousness | S06.6X4A | Traumatic subarachnoid hemorrhage with loss of consciousness of 6 hours to 24 hours, initial encounter |
| 800.23 | Closed fracture of vault of skull with subarachnoid, subdural, and extradural hemorrhage, with moderate [1-24 hours] loss of consciousness |  |  |
| 800.24 | Closed fracture of vault of skull with subarachnoid, subdural, and extradural hemorrhage, with prolonged [more than 24 hours] loss of consciousness and return to pre-existing conscious level | S02.0XXA | Fracture of vault of skull, initial encounter for closed fracture |
| 800.24 | Closed fracture of vault of skull with subarachnoid, subdural, and extradural hemorrhage, with prolonged [more than 24 hours] loss of consciousness and return to pre-existing conscious level | S06.4X5A | Epidural hemorrhage with loss of consciousness greater than 24 hours with return to pre-existing conscious level, initial encounter |
| 800.24 | Closed fracture of vault of skull with subarachnoid, subdural, and extradural hemorrhage, with prolonged [more than 24 hours] loss of consciousness and return to pre-existing conscious level | S06.5X5A | Traumatic subdural hemorrhage with loss of consciousness greater than 24 hours with return to pre-existing conscious level, initial encounter |
| 800.24 | Closed fracture of vault of skull with subarachnoid, subdural, and extradural hemorrhage, with prolonged [more than 24 hours] loss of consciousness and return to pre-existing conscious level | S06.6X5A | Traumatic subarachnoid hemorrhage with loss of consciousness greater than 24 hours with return to pre-existing conscious level, initial encounter |
| 800.25 | Closed fracture of vault of skull with subarachnoid, subdural, and extradural hemorrhage, with prolonged [more than 24 hours] loss of consciousness, without return to pre-existing conscious level | S06.4X6A | Epidural hemorrhage with loss of consciousness greater than 24 hours without return to pre-existing conscious level with patient surviving, initial encounter |
| 800.25 | Closed fracture of vault of skull with subarachnoid, subdural, and extradural hemorrhage, with prolonged [more than 24 hours] loss of consciousness, without return to pre-existing conscious level | S06.4X7A | Epidural hemorrhage with loss of consciousness of any duration with death due to brain injury prior to regaining consciousness, initial encounter |
| 800.25 | Closed fracture of vault of skull with subarachnoid, subdural, and extradural hemorrhage, with prolonged [more than 24 hours] loss of consciousness, without return to pre-existing conscious level | S06.4X8A | Epidural hemorrhage with loss of consciousness of any duration with death due to other causes prior to regaining consciousness, initial encounter |
| 800.25 | Closed fracture of vault of skull with subarachnoid, subdural, and extradural hemorrhage, with prolonged [more than 24 hours] loss of consciousness, without return to pre-existing conscious level | S06.5X6A | Traumatic subdural hemorrhage with loss of consciousness greater than 24 hours without return to pre-existing conscious level with patient surviving, initial encounter |
| 800.25 | Closed fracture of vault of skull with subarachnoid, subdural, and extradural hemorrhage, with prolonged [more than 24 hours] loss of consciousness, without return to pre-existing conscious level | S06.5X7A | Traumatic subdural hemorrhage with loss of consciousness of any duration with death due to brain injury before regaining consciousness, initial encounter |
| 800.25 | Closed fracture of vault of skull with subarachnoid, subdural, and extradural hemorrhage, with prolonged [more than 24 hours] loss of consciousness, without return to pre-existing conscious level | S06.5X8A | Traumatic subdural hemorrhage with loss of consciousness of any duration with death due to other cause before regaining consciousness, initial encounter |
| 800.25 | Closed fracture of vault of skull with subarachnoid, subdural, and extradural hemorrhage, with prolonged [more than 24 hours] loss of consciousness, without return to pre-existing conscious level | S06.6X6A | Traumatic subarachnoid hemorrhage with loss of consciousness greater than 24 hours without return to pre-existing conscious level with patient surviving, initial encounter |
| 800.25 | Closed fracture of vault of skull with subarachnoid, subdural, and extradural hemorrhage, with prolonged [more than 24 hours] loss of consciousness, without return to pre-existing conscious level | S06.6X7A | Traumatic subarachnoid hemorrhage with loss of consciousness of any duration with death due to brain injury prior to regaining consciousness, initial encounter |
| 800.25 | Closed fracture of vault of skull with subarachnoid, subdural, and extradural hemorrhage, with prolonged [more than 24 hours] loss of consciousness, without return to pre-existing conscious level | S06.6X8A | Traumatic subarachnoid hemorrhage with loss of consciousness of any duration with death due to other cause prior to regaining consciousness, initial encounter |
| 800.25 | Closed fracture of vault of skull with subarachnoid, subdural, and extradural hemorrhage, with prolonged [more than 24 hours] loss of consciousness, without return to pre-existing conscious level |  |  |
| 800.26 | Closed fracture of vault of skull with subarachnoid, subdural, and extradural hemorrhage, with loss of consciousness of unspecified duration | S06.4X9A | Epidural hemorrhage with loss of consciousness of unspecified duration, initial encounter |
| 800.26 | Closed fracture of vault of skull with subarachnoid, subdural, and extradural hemorrhage, with loss of consciousness of unspecified duration | S06.5X9A | Traumatic subdural hemorrhage with loss of consciousness of unspecified duration, initial encounter |
| 800.26 | Closed fracture of vault of skull with subarachnoid, subdural, and extradural hemorrhage, with loss of consciousness of unspecified duration | S06.6X9A | Traumatic subarachnoid hemorrhage with loss of consciousness of unspecified duration, initial encounter |
| 800.26 | Closed fracture of vault of skull with subarachnoid, subdural, and extradural hemorrhage, with loss of consciousness of unspecified duration |  |  |
| 800.29 | Closed fracture of vault of skull with subarachnoid, subdural, and extradural hemorrhage, with concussion, unspecified | S06.4X0A | Epidural hemorrhage without loss of consciousness, initial encounter |
| 800.29 | Closed fracture of vault of skull with subarachnoid, subdural, and extradural hemorrhage, with concussion, unspecified | S06.5X0A | Traumatic subdural hemorrhage without loss of consciousness, initial encounter |
| 800.29 | Closed fracture of vault of skull with subarachnoid, subdural, and extradural hemorrhage, with concussion, unspecified | S06.6X0A | Traumatic subarachnoid hemorrhage without loss of consciousness, initial encounter |
| 800.29 | Closed fracture of vault of skull with subarachnoid, subdural, and extradural hemorrhage, with concussion, unspecified |  |  |
| 800.30 | Closed fracture of vault of skull with other and unspecified intracranial hemorrhage, unspecified state of consciousness | S02.0XXA | Fracture of vault of skull, initial encounter for closed fracture |
| 800.30 | Closed fracture of vault of skull with other and unspecified intracranial hemorrhage, unspecified state of consciousness | S06.360A | Traumatic hemorrhage of cerebrum, unspecified, without loss of consciousness, initial encounter |
| 800.31 | Closed fracture of vault of skull with other and unspecified intracranial hemorrhage, with no loss of consciousness | S02.0XXA | Fracture of vault of skull, initial encounter for closed fracture |
| 800.31 | Closed fracture of vault of skull with other and unspecified intracranial hemorrhage, with no loss of consciousness | S06.360A | Traumatic hemorrhage of cerebrum, unspecified, without loss of consciousness, initial encounter |
| 800.32 | Closed fracture of vault of skull with other and unspecified intracranial hemorrhage, with brief [less than one hour] loss of consciousness | S02.0XXA | Fracture of vault of skull, initial encounter for closed fracture |
| 800.32 | Closed fracture of vault of skull with other and unspecified intracranial hemorrhage, with brief [less than one hour] loss of consciousness | S06.361A | Traumatic hemorrhage of cerebrum, unspecified, with loss of consciousness of 30 minutes or less, initial encounter |
| 800.32 | Closed fracture of vault of skull with other and unspecified intracranial hemorrhage, with brief [less than one hour] loss of consciousness | S06.362A | Traumatic hemorrhage of cerebrum, unspecified, with loss of consciousness of 31 minutes to 59 minutes, initial encounter |
| 800.33 | Closed fracture of vault of skull with other and unspecified intracranial hemorrhage, with moderate [1-24 hours] loss of consciousness | S02.0XXA | Fracture of vault of skull, initial encounter for closed fracture |
| 800.33 | Closed fracture of vault of skull with other and unspecified intracranial hemorrhage, with moderate [1-24 hours] loss of consciousness | S06.363A | Traumatic hemorrhage of cerebrum, unspecified, with loss of consciousness of 1 hours to 5 hours 59 minutes, initial encounter |
| 800.33 | Closed fracture of vault of skull with other and unspecified intracranial hemorrhage, with moderate [1-24 hours] loss of consciousness | S06.364A | Traumatic hemorrhage of cerebrum, unspecified, with loss of consciousness of 6 hours to 24 hours, initial encounter |
| 800.34 | Closed fracture of vault of skull with other and unspecified intracranial hemorrhage, with prolonged [more than 24 hours] loss of consciousness and return to pre-existing conscious level | S02.0XXA | Fracture of vault of skull, initial encounter for closed fracture |
| 800.34 | Closed fracture of vault of skull with other and unspecified intracranial hemorrhage, with prolonged [more than 24 hours] loss of consciousness and return to pre-existing conscious level | S06.365A | Traumatic hemorrhage of cerebrum, unspecified, with loss of consciousness greater than 24 hours with return to pre-existing conscious level, initial encounter |
| 800.35 | Closed fracture of vault of skull with other and unspecified intracranial hemorrhage, with prolonged [more than 24 hours] loss of consciousness, without return to pre-existing conscious level | S02.0XXA | Fracture of vault of skull, initial encounter for closed fracture |
| 800.35 | Closed fracture of vault of skull with other and unspecified intracranial hemorrhage, with prolonged [more than 24 hours] loss of consciousness, without return to pre-existing conscious level | S06.366A | Traumatic hemorrhage of cerebrum, unspecified, with loss of consciousness greater than 24 hours without return to pre-existing conscious level with patient surviving, initial encounter |
| 800.35 | Closed fracture of vault of skull with other and unspecified intracranial hemorrhage, with prolonged [more than 24 hours] loss of consciousness, without return to pre-existing conscious level | S06.367A | Traumatic hemorrhage of cerebrum, unspecified, with loss of consciousness of any duration with death due to brain injury prior to regaining consciousness, initial encounter |
| 800.35 | Closed fracture of vault of skull with other and unspecified intracranial hemorrhage, with prolonged [more than 24 hours] loss of consciousness, without return to pre-existing conscious level | S06.368A | Traumatic hemorrhage of cerebrum, unspecified, with loss of consciousness of any duration with death due to other cause prior to regaining consciousness, initial encounter |
| 800.36 | Closed fracture of vault of skull with other and unspecified intracranial hemorrhage, with loss of consciousness of unspecified duration | S02.0XXA | Fracture of vault of skull, initial encounter for closed fracture |
| 800.36 | Closed fracture of vault of skull with other and unspecified intracranial hemorrhage, with loss of consciousness of unspecified duration | S06.369A | Traumatic hemorrhage of cerebrum, unspecified, with loss of consciousness of unspecified duration, initial encounter |
| 800.39 | Closed fracture of vault of skull with other and unspecified intracranial hemorrhage, with concussion, unspecified | S02.0XXA | Fracture of vault of skull, initial encounter for closed fracture |
| 800.39 | Closed fracture of vault of skull with other and unspecified intracranial hemorrhage, with concussion, unspecified | S06.360A | Traumatic hemorrhage of cerebrum, unspecified, without loss of consciousness, initial encounter |
| 800.40 | Closed fracture of vault of skull with intracranial injury of other and unspecified nature, unspecified state of consciousness | S02.0XXA | Fracture of vault of skull, initial encounter for closed fracture |
| 800.40 | Closed fracture of vault of skull with intracranial injury of other and unspecified nature, unspecified state of consciousness | S06.890A | Other specified intracranial injury without loss of consciousness, initial encounter |
| 800.40 | Closed fracture of vault of skull with intracranial injury of other and unspecified nature, unspecified state of consciousness | S06.9X0A | Unspecified intracranial injury without loss of consciousness, initial encounter |
| 800.41 | Closed fracture of vault of skull with intracranial injury of other and unspecified nature, with no loss of consciousness | S02.0XXA | Fracture of vault of skull, initial encounter for closed fracture |
| 800.41 | Closed fracture of vault of skull with intracranial injury of other and unspecified nature, with no loss of consciousness | S06.890A | Other specified intracranial injury without loss of consciousness, initial encounter |
| 800.41 | Closed fracture of vault of skull with intracranial injury of other and unspecified nature, with no loss of consciousness | S06.9X0A | Unspecified intracranial injury without loss of consciousness, initial encounter |
| 800.42 | Closed fracture of vault of skull with intracranial injury of other and unspecified nature, with brief [less than one hour] loss of consciousness | S02.0XXA | Fracture of vault of skull, initial encounter for closed fracture |
| 800.42 | Closed fracture of vault of skull with intracranial injury of other and unspecified nature, with brief [less than one hour] loss of consciousness | S06.891A | Other specified intracranial injury with loss of consciousness of 30 minutes or less, initial encounter |
| 800.42 | Closed fracture of vault of skull with intracranial injury of other and unspecified nature, with brief [less than one hour] loss of consciousness | S06.892A | Other specified intracranial injury with loss of consciousness of 31 minutes to 59 minutes, initial encounter |
| 800.42 | Closed fracture of vault of skull with intracranial injury of other and unspecified nature, with brief [less than one hour] loss of consciousness | S06.9X1A | Unspecified intracranial injury with loss of consciousness of 30 minutes or less, initial encounter |
| 800.42 | Closed fracture of vault of skull with intracranial injury of other and unspecified nature, with brief [less than one hour] loss of consciousness | S06.9X2A | Unspecified intracranial injury with loss of consciousness of 31 minutes to 59 minutes, initial encounter |
| 800.43 | Closed fracture of vault of skull with intracranial injury of other and unspecified nature, with moderate [1-24 hours] loss of consciousness | S02.0XXA | Fracture of vault of skull, initial encounter for closed fracture |
| 800.43 | Closed fracture of vault of skull with intracranial injury of other and unspecified nature, with moderate [1-24 hours] loss of consciousness | S06.893A | Other specified intracranial injury with loss of consciousness of 1 hour to 5 hours 59 minutes, initial encounter |
| 800.43 | Closed fracture of vault of skull with intracranial injury of other and unspecified nature, with moderate [1-24 hours] loss of consciousness | S06.894A | Other specified intracranial injury with loss of consciousness of 6 hours to 24 hours, initial encounter |
| 800.43 | Closed fracture of vault of skull with intracranial injury of other and unspecified nature, with moderate [1-24 hours] loss of consciousness | S06.9X3A | Unspecified intracranial injury with loss of consciousness of 1 hour to 5 hours 59 minutes, initial encounter |
| 800.43 | Closed fracture of vault of skull with intracranial injury of other and unspecified nature, with moderate [1-24 hours] loss of consciousness | S06.9X4A | Unspecified intracranial injury with loss of consciousness of 6 hours to 24 hours, initial encounter |
| 800.44 | Closed fracture of vault of skull with intracranial injury of other and unspecified nature, with prolonged [more than 24 hours] loss of consciousness and return to pre-existing conscious level | S02.0XXA | Fracture of vault of skull, initial encounter for closed fracture |
| 800.44 | Closed fracture of vault of skull with intracranial injury of other and unspecified nature, with prolonged [more than 24 hours] loss of consciousness and return to pre-existing conscious level | S06.895A | Other specified intracranial injury with loss of consciousness greater than 24 hours with return to pre-existing conscious level, initial encounter |
| 800.44 | Closed fracture of vault of skull with intracranial injury of other and unspecified nature, with prolonged [more than 24 hours] loss of consciousness and return to pre-existing conscious level | S06.9X5A | Unspecified intracranial injury with loss of consciousness greater than 24 hours with return to pre-existing conscious level, initial encounter |
| 800.45 | Closed fracture of vault of skull with intracranial injury of other and unspecified nature, with prolonged [more than 24 hours] loss of consciousness, without return to pre-existing conscious level | S02.0XXA | Fracture of vault of skull, initial encounter for closed fracture |
| 800.45 | Closed fracture of vault of skull with intracranial injury of other and unspecified nature, with prolonged [more than 24 hours] loss of consciousness, without return to pre-existing conscious level | S06.896A | Other specified intracranial injury with loss of consciousness greater than 24 hours without return to pre-existing conscious level with patient surviving, initial encounter |
| 800.45 | Closed fracture of vault of skull with intracranial injury of other and unspecified nature, with prolonged [more than 24 hours] loss of consciousness, without return to pre-existing conscious level | S06.897A | Other specified intracranial injury with loss of consciousness of any duration with death due to brain injury prior to regaining consciousness, initial encounter |
| 800.45 | Closed fracture of vault of skull with intracranial injury of other and unspecified nature, with prolonged [more than 24 hours] loss of consciousness, without return to pre-existing conscious level | S06.898A | Other specified intracranial injury with loss of consciousness of any duration with death due to other cause prior to regaining consciousness, initial encounter |
| 800.45 | Closed fracture of vault of skull with intracranial injury of other and unspecified nature, with prolonged [more than 24 hours] loss of consciousness, without return to pre-existing conscious level | S06.9X6A | Unspecified intracranial injury with loss of consciousness greater than 24 hours without return to pre-existing conscious level with patient surviving, initial encounter |
| 800.45 | Closed fracture of vault of skull with intracranial injury of other and unspecified nature, with prolonged [more than 24 hours] loss of consciousness, without return to pre-existing conscious level | S06.9X7A | Unspecified intracranial injury with loss of consciousness of any duration with death due to brain injury prior to regaining consciousness, initial encounter |
| 800.45 | Closed fracture of vault of skull with intracranial injury of other and unspecified nature, with prolonged [more than 24 hours] loss of consciousness, without return to pre-existing conscious level | S06.9X8A | Unspecified intracranial injury with loss of consciousness of any duration with death due to other cause prior to regaining consciousness, initial encounter |
| 800.46 | Closed fracture of vault of skull with intracranial injury of other and unspecified nature, with loss of consciousness of unspecified duration | S02.0XXA | Fracture of vault of skull, initial encounter for closed fracture |
| 800.46 | Closed fracture of vault of skull with intracranial injury of other and unspecified nature, with loss of consciousness of unspecified duration | S06.899A | Other specified intracranial injury with loss of consciousness of unspecified duration, initial encounter |
| 800.46 | Closed fracture of vault of skull with intracranial injury of other and unspecified nature, with loss of consciousness of unspecified duration | S06.9X9A | Unspecified intracranial injury with loss of consciousness of unspecified duration, initial encounter |
| 800.49 | Closed fracture of vault of skull with intracranial injury of other and unspecified nature, with concussion, unspecified | S02.0XXA | Fracture of vault of skull, initial encounter for closed fracture |
| 800.49 | Closed fracture of vault of skull with intracranial injury of other and unspecified nature, with concussion, unspecified | S06.890A | Other specified intracranial injury without loss of consciousness, initial encounter |
| 800.49 | Closed fracture of vault of skull with intracranial injury of other and unspecified nature, with concussion, unspecified | S06.9X0A | Unspecified intracranial injury without loss of consciousness, initial encounter |
| 800.60 | Open fracture of vault of skull with cerebral laceration and contusion, unspecified state of consciousness | S02.0XXB | Fracture of vault of skull, initial encounter for open fracture |
| 800.60 | Open fracture of vault of skull with cerebral laceration and contusion, unspecified state of consciousness | S06.330A | Contusion and laceration of cerebrum, unspecified, without loss of consciousness, initial encounter |
| 800.61 | Open fracture of vault of skull with cerebral laceration and contusion, with no loss of consciousness | S02.0XXB | Fracture of vault of skull, initial encounter for open fracture |
| 800.61 | Open fracture of vault of skull with cerebral laceration and contusion, with no loss of consciousness | S06.330A | Contusion and laceration of cerebrum, unspecified, without loss of consciousness, initial encounter |
| 800.62 | Open fracture of vault of skull with cerebral laceration and contusion, with brief [less than one hour] loss of consciousness | S02.0XXB | Fracture of vault of skull, initial encounter for open fracture |
| 800.62 | Open fracture of vault of skull with cerebral laceration and contusion, with brief [less than one hour] loss of consciousness | S06.331A | Contusion and laceration of cerebrum, unspecified, with loss of consciousness of 30 minutes or less, initial encounter |
| 800.62 | Open fracture of vault of skull with cerebral laceration and contusion, with brief [less than one hour] loss of consciousness | S06.332A | Contusion and laceration of cerebrum, unspecified, with loss of consciousness of 31 minutes to 59 minutes, initial encounter |
| 800.63 | Open fracture of vault of skull with cerebral laceration and contusion, with moderate [1-24 hours] loss of consciousness | S02.0XXB | Fracture of vault of skull, initial encounter for open fracture |
| 800.63 | Open fracture of vault of skull with cerebral laceration and contusion, with moderate [1-24 hours] loss of consciousness | S06.333A | Contusion and laceration of cerebrum, unspecified, with loss of consciousness of 1 hour to 5 hours 59 minutes, initial encounter |
| 800.63 | Open fracture of vault of skull with cerebral laceration and contusion, with moderate [1-24 hours] loss of consciousness | S06.334A | Contusion and laceration of cerebrum, unspecified, with loss of consciousness of 6 hours to 24 hours, initial encounter |
| 800.64 | Open fracture of vault of skull with cerebral laceration and contusion, with prolonged [more than 24 hours] loss of consciousness and return to pre-existing conscious level | S02.0XXB | Fracture of vault of skull, initial encounter for open fracture |
| 800.64 | Open fracture of vault of skull with cerebral laceration and contusion, with prolonged [more than 24 hours] loss of consciousness and return to pre-existing conscious level | S06.335A | Contusion and laceration of cerebrum, unspecified, with loss of consciousness greater than 24 hours with return to pre-existing conscious level, initial encounter |
| 800.65 | Open fracture of vault of skull with cerebral laceration and contusion, with prolonged [more than 24 hours] loss of consciousness, without return to pre-existing conscious level | S02.0XXB | Fracture of vault of skull, initial encounter for open fracture |
| 800.65 | Open fracture of vault of skull with cerebral laceration and contusion, with prolonged [more than 24 hours] loss of consciousness, without return to pre-existing conscious level | S06.336A | Contusion and laceration of cerebrum, unspecified, with loss of consciousness greater than 24 hours without return to pre-existing conscious level with patient surviving, initial encounter |
| 800.65 | Open fracture of vault of skull with cerebral laceration and contusion, with prolonged [more than 24 hours] loss of consciousness, without return to pre-existing conscious level | S06.337A | Contusion and laceration of cerebrum, unspecified, with loss of consciousness of any duration with death due to brain injury prior to regaining consciousness, initial encounter |
| 800.65 | Open fracture of vault of skull with cerebral laceration and contusion, with prolonged [more than 24 hours] loss of consciousness, without return to pre-existing conscious level | S06.338A | Contusion and laceration of cerebrum, unspecified, with loss of consciousness of any duration with death due to other cause prior to regaining consciousness, initial encounter |
| 800.66 | Open fracture of vault of skull with cerebral laceration and contusion, with loss of consciousness of unspecified duration | S02.0XXB | Fracture of vault of skull, initial encounter for open fracture |
| 800.66 | Open fracture of vault of skull with cerebral laceration and contusion, with loss of consciousness of unspecified duration | S06.339A | Contusion and laceration of cerebrum, unspecified, with loss of consciousness of unspecified duration, initial encounter |
| 800.69 | Open fracture of vault of skull with cerebral laceration and contusion, with concussion, unspecified | S02.0XXB | Fracture of vault of skull, initial encounter for open fracture |
| 800.69 | Open fracture of vault of skull with cerebral laceration and contusion, with concussion, unspecified | S06.330A | Contusion and laceration of cerebrum, unspecified, without loss of consciousness, initial encounter |
| 800.70 | Open fracture of vault of skull with subarachnoid, subdural, and extradural hemorrhage, unspecified state of consciousness | S06.4X0A | Epidural hemorrhage without loss of consciousness, initial encounter |
| 800.70 | Open fracture of vault of skull with subarachnoid, subdural, and extradural hemorrhage, unspecified state of consciousness | S06.5X0A | Traumatic subdural hemorrhage without loss of consciousness, initial encounter |
| 800.70 | Open fracture of vault of skull with subarachnoid, subdural, and extradural hemorrhage, unspecified state of consciousness | S06.6X0A | Traumatic subarachnoid hemorrhage without loss of consciousness, initial encounter |
| 800.70 | Open fracture of vault of skull with subarachnoid, subdural, and extradural hemorrhage, unspecified state of consciousness |  |  |
| 800.71 | Open fracture of vault of skull with subarachnoid, subdural, and extradural hemorrhage, with no loss of consciousness | S06.4X0A | Epidural hemorrhage without loss of consciousness, initial encounter |
| 800.71 | Open fracture of vault of skull with subarachnoid, subdural, and extradural hemorrhage, with no loss of consciousness | S06.5X0A | Traumatic subdural hemorrhage without loss of consciousness, initial encounter |
| 800.71 | Open fracture of vault of skull with subarachnoid, subdural, and extradural hemorrhage, with no loss of consciousness | S06.6X0A | Traumatic subarachnoid hemorrhage without loss of consciousness, initial encounter |
| 800.71 | Open fracture of vault of skull with subarachnoid, subdural, and extradural hemorrhage, with no loss of consciousness |  |  |
| 800.72 | Open fracture of vault of skull with subarachnoid, subdural, and extradural hemorrhage, with brief [less than one hour] loss of consciousness | S06.4X1A | Epidural hemorrhage with loss of consciousness of 30 minutes or less, initial encounter |
| 800.72 | Open fracture of vault of skull with subarachnoid, subdural, and extradural hemorrhage, with brief [less than one hour] loss of consciousness | S06.4X2A | Epidural hemorrhage with loss of consciousness of 31 minutes to 59 minutes, initial encounter |
| 800.72 | Open fracture of vault of skull with subarachnoid, subdural, and extradural hemorrhage, with brief [less than one hour] loss of consciousness | S06.5X1A | Traumatic subdural hemorrhage with loss of consciousness of 30 minutes or less, initial encounter |
| 800.72 | Open fracture of vault of skull with subarachnoid, subdural, and extradural hemorrhage, with brief [less than one hour] loss of consciousness | S06.5X2A | Traumatic subdural hemorrhage with loss of consciousness of 31 minutes to 59 minutes, initial encounter |
| 800.72 | Open fracture of vault of skull with subarachnoid, subdural, and extradural hemorrhage, with brief [less than one hour] loss of consciousness | S06.6X1A | Traumatic subarachnoid hemorrhage with loss of consciousness of 30 minutes or less, initial encounter |
| 800.72 | Open fracture of vault of skull with subarachnoid, subdural, and extradural hemorrhage, with brief [less than one hour] loss of consciousness | S06.6X2A | Traumatic subarachnoid hemorrhage with loss of consciousness of 31 minutes to 59 minutes, initial encounter |
| 800.72 | Open fracture of vault of skull with subarachnoid, subdural, and extradural hemorrhage, with brief [less than one hour] loss of consciousness |  |  |
| 800.73 | Open fracture of vault of skull with subarachnoid, subdural, and extradural hemorrhage, with moderate [1-24 hours] loss of consciousness | S06.4X3A | Epidural hemorrhage with loss of consciousness of 1 hour to 5 hours 59 minutes, initial encounter |
| 800.73 | Open fracture of vault of skull with subarachnoid, subdural, and extradural hemorrhage, with moderate [1-24 hours] loss of consciousness | S06.4X4A | Epidural hemorrhage with loss of consciousness of 6 hours to 24 hours, initial encounter |
| 800.73 | Open fracture of vault of skull with subarachnoid, subdural, and extradural hemorrhage, with moderate [1-24 hours] loss of consciousness | S06.5X3A | Traumatic subdural hemorrhage with loss of consciousness of 1 hour to 5 hours 59 minutes, initial encounter |
| 800.73 | Open fracture of vault of skull with subarachnoid, subdural, and extradural hemorrhage, with moderate [1-24 hours] loss of consciousness | S06.5X4A | Traumatic subdural hemorrhage with loss of consciousness of 6 hours to 24 hours, initial encounter |
| 800.73 | Open fracture of vault of skull with subarachnoid, subdural, and extradural hemorrhage, with moderate [1-24 hours] loss of consciousness | S06.6X3A | Traumatic subarachnoid hemorrhage with loss of consciousness of 1 hour to 5 hours 59 minutes, initial encounter |
| 800.73 | Open fracture of vault of skull with subarachnoid, subdural, and extradural hemorrhage, with moderate [1-24 hours] loss of consciousness | S06.6X4A | Traumatic subarachnoid hemorrhage with loss of consciousness of 6 hours to 24 hours, initial encounter |
| 800.73 | Open fracture of vault of skull with subarachnoid, subdural, and extradural hemorrhage, with moderate [1-24 hours] loss of consciousness |  |  |
| 800.74 | Open fracture of vault of skull with subarachnoid, subdural, and extradural hemorrhage, with prolonged [more than 24 hours] loss of consciousness and return to pre-existing conscious level | S06.4X5A | Epidural hemorrhage with loss of consciousness greater than 24 hours with return to pre-existing conscious level, initial encounter |
| 800.74 | Open fracture of vault of skull with subarachnoid, subdural, and extradural hemorrhage, with prolonged [more than 24 hours] loss of consciousness and return to pre-existing conscious level | S06.5X5A | Traumatic subdural hemorrhage with loss of consciousness greater than 24 hours with return to pre-existing conscious level, initial encounter |
| 800.74 | Open fracture of vault of skull with subarachnoid, subdural, and extradural hemorrhage, with prolonged [more than 24 hours] loss of consciousness and return to pre-existing conscious level | S06.6X5A | Traumatic subarachnoid hemorrhage with loss of consciousness greater than 24 hours with return to pre-existing conscious level, initial encounter |
| 800.74 | Open fracture of vault of skull with subarachnoid, subdural, and extradural hemorrhage, with prolonged [more than 24 hours] loss of consciousness and return to pre-existing conscious level |  |  |
| 800.75 | Open fracture of vault of skull with subarachnoid, subdural, and extradural hemorrhage, with prolonged [more than 24 hours] loss of consciousness, without return to pre-existing conscious level | S06.4X6A | Epidural hemorrhage with loss of consciousness greater than 24 hours without return to pre-existing conscious level with patient surviving, initial encounter |
| 800.75 | Open fracture of vault of skull with subarachnoid, subdural, and extradural hemorrhage, with prolonged [more than 24 hours] loss of consciousness, without return to pre-existing conscious level | S06.4X7A | Epidural hemorrhage with loss of consciousness of any duration with death due to brain injury prior to regaining consciousness, initial encounter |
| 800.75 | Open fracture of vault of skull with subarachnoid, subdural, and extradural hemorrhage, with prolonged [more than 24 hours] loss of consciousness, without return to pre-existing conscious level | S06.4X8A | Epidural hemorrhage with loss of consciousness of any duration with death due to other causes prior to regaining consciousness, initial encounter |
| 800.75 | Open fracture of vault of skull with subarachnoid, subdural, and extradural hemorrhage, with prolonged [more than 24 hours] loss of consciousness, without return to pre-existing conscious level | S06.5X6A | Traumatic subdural hemorrhage with loss of consciousness greater than 24 hours without return to pre-existing conscious level with patient surviving, initial encounter |
| 800.75 | Open fracture of vault of skull with subarachnoid, subdural, and extradural hemorrhage, with prolonged [more than 24 hours] loss of consciousness, without return to pre-existing conscious level | S06.5X7A | Traumatic subdural hemorrhage with loss of consciousness of any duration with death due to brain injury before regaining consciousness, initial encounter |
| 800.75 | Open fracture of vault of skull with subarachnoid, subdural, and extradural hemorrhage, with prolonged [more than 24 hours] loss of consciousness, without return to pre-existing conscious level | S06.5X8A | Traumatic subdural hemorrhage with loss of consciousness of any duration with death due to other cause before regaining consciousness, initial encounter |
| 800.75 | Open fracture of vault of skull with subarachnoid, subdural, and extradural hemorrhage, with prolonged [more than 24 hours] loss of consciousness, without return to pre-existing conscious level | S06.6X6A | Traumatic subarachnoid hemorrhage with loss of consciousness greater than 24 hours without return to pre-existing conscious level with patient surviving, initial encounter |
| 800.75 | Open fracture of vault of skull with subarachnoid, subdural, and extradural hemorrhage, with prolonged [more than 24 hours] loss of consciousness, without return to pre-existing conscious level | S06.6X7A | Traumatic subarachnoid hemorrhage with loss of consciousness of any duration with death due to brain injury prior to regaining consciousness, initial encounter |
| 800.75 | Open fracture of vault of skull with subarachnoid, subdural, and extradural hemorrhage, with prolonged [more than 24 hours] loss of consciousness, without return to pre-existing conscious level | S06.6X8A | Traumatic subarachnoid hemorrhage with loss of consciousness of any duration with death due to other cause prior to regaining consciousness, initial encounter |
| 800.75 | Open fracture of vault of skull with subarachnoid, subdural, and extradural hemorrhage, with prolonged [more than 24 hours] loss of consciousness, without return to pre-existing conscious level |  |  |
| 800.76 | Open fracture of vault of skull with subarachnoid, subdural, and extradural hemorrhage, with loss of consciousness of unspecified duration | S06.4X9A | Epidural hemorrhage with loss of consciousness of unspecified duration, initial encounter |
| 800.76 | Open fracture of vault of skull with subarachnoid, subdural, and extradural hemorrhage, with loss of consciousness of unspecified duration | S06.5X9A | Traumatic subdural hemorrhage with loss of consciousness of unspecified duration, initial encounter |
| 800.76 | Open fracture of vault of skull with subarachnoid, subdural, and extradural hemorrhage, with loss of consciousness of unspecified duration | S06.6X9A | Traumatic subarachnoid hemorrhage with loss of consciousness of unspecified duration, initial encounter |
| 800.76 | Open fracture of vault of skull with subarachnoid, subdural, and extradural hemorrhage, with loss of consciousness of unspecified duration |  |  |
| 800.79 | Open fracture of vault of skull with subarachnoid, subdural, and extradural hemorrhage, with concussion, unspecified | S06.4X0A | Epidural hemorrhage without loss of consciousness, initial encounter |
| 800.79 | Open fracture of vault of skull with subarachnoid, subdural, and extradural hemorrhage, with concussion, unspecified | S06.5X0A | Traumatic subdural hemorrhage without loss of consciousness, initial encounter |
| 800.79 | Open fracture of vault of skull with subarachnoid, subdural, and extradural hemorrhage, with concussion, unspecified | S06.6X0A | Traumatic subarachnoid hemorrhage without loss of consciousness, initial encounter |
| 800.79 | Open fracture of vault of skull with subarachnoid, subdural, and extradural hemorrhage, with concussion, unspecified |  |  |
| 800.80 | Open fracture of vault of skull with other and unspecified intracranial hemorrhage, unspecified state of consciousness | S02.0XXB | Fracture of vault of skull, initial encounter for open fracture |
| 800.80 | Open fracture of vault of skull with other and unspecified intracranial hemorrhage, unspecified state of consciousness | S06.360A | Traumatic hemorrhage of cerebrum, unspecified, without loss of consciousness, initial encounter |
| 800.81 | Open fracture of vault of skull with other and unspecified intracranial hemorrhage, with no loss of consciousness | S02.0XXB | Fracture of vault of skull, initial encounter for open fracture |
| 800.81 | Open fracture of vault of skull with other and unspecified intracranial hemorrhage, with no loss of consciousness | S06.360A | Traumatic hemorrhage of cerebrum, unspecified, without loss of consciousness, initial encounter |
| 800.82 | Open fracture of vault of skull with other and unspecified intracranial hemorrhage, with brief [less than one hour] loss of consciousness | S02.0XXB | Fracture of vault of skull, initial encounter for open fracture |
| 800.82 | Open fracture of vault of skull with other and unspecified intracranial hemorrhage, with brief [less than one hour] loss of consciousness | S06.361A | Traumatic hemorrhage of cerebrum, unspecified, with loss of consciousness of 30 minutes or less, initial encounter |
| 800.82 | Open fracture of vault of skull with other and unspecified intracranial hemorrhage, with brief [less than one hour] loss of consciousness | S06.362A | Traumatic hemorrhage of cerebrum, unspecified, with loss of consciousness of 31 minutes to 59 minutes, initial encounter |
| 800.83 | Open fracture of vault of skull with other and unspecified intracranial hemorrhage, with moderate [1-24 hours] loss of consciousness | S02.0XXB | Fracture of vault of skull, initial encounter for open fracture |
| 800.83 | Open fracture of vault of skull with other and unspecified intracranial hemorrhage, with moderate [1-24 hours] loss of consciousness | S06.363A | Traumatic hemorrhage of cerebrum, unspecified, with loss of consciousness of 1 hours to 5 hours 59 minutes, initial encounter |
| 800.83 | Open fracture of vault of skull with other and unspecified intracranial hemorrhage, with moderate [1-24 hours] loss of consciousness | S06.364A | Traumatic hemorrhage of cerebrum, unspecified, with loss of consciousness of 6 hours to 24 hours, initial encounter |
| 800.84 | Open fracture of vault of skull with other and unspecified intracranial hemorrhage, with prolonged [more than 24 hours] loss of consciousness and return to pre-existing conscious level | S02.0XXB | Fracture of vault of skull, initial encounter for open fracture |
| 800.84 | Open fracture of vault of skull with other and unspecified intracranial hemorrhage, with prolonged [more than 24 hours] loss of consciousness and return to pre-existing conscious level | S06.365A | Traumatic hemorrhage of cerebrum, unspecified, with loss of consciousness greater than 24 hours with return to pre-existing conscious level, initial encounter |
| 800.85 | Open fracture of vault of skull with other and unspecified intracranial hemorrhage, with prolonged [more than 24 hours] loss of consciousness, without return to pre-existing conscious level | S02.0XXB | Fracture of vault of skull, initial encounter for open fracture |
| 800.85 | Open fracture of vault of skull with other and unspecified intracranial hemorrhage, with prolonged [more than 24 hours] loss of consciousness, without return to pre-existing conscious level | S06.366A | Traumatic hemorrhage of cerebrum, unspecified, with loss of consciousness greater than 24 hours without return to pre-existing conscious level with patient surviving, initial encounter |
| 800.85 | Open fracture of vault of skull with other and unspecified intracranial hemorrhage, with prolonged [more than 24 hours] loss of consciousness, without return to pre-existing conscious level | S06.367A | Traumatic hemorrhage of cerebrum, unspecified, with loss of consciousness of any duration with death due to brain injury prior to regaining consciousness, initial encounter |
| 800.85 | Open fracture of vault of skull with other and unspecified intracranial hemorrhage, with prolonged [more than 24 hours] loss of consciousness, without return to pre-existing conscious level | S06.368A | Traumatic hemorrhage of cerebrum, unspecified, with loss of consciousness of any duration with death due to other cause prior to regaining consciousness, initial encounter |
| 800.86 | Open fracture of vault of skull with other and unspecified intracranial hemorrhage, with loss of consciousness of unspecified duration | S02.0XXB | Fracture of vault of skull, initial encounter for open fracture |
| 800.86 | Open fracture of vault of skull with other and unspecified intracranial hemorrhage, with loss of consciousness of unspecified duration | S06.369A | Traumatic hemorrhage of cerebrum, unspecified, with loss of consciousness of unspecified duration, initial encounter |
| 800.89 | Open fracture of vault of skull with other and unspecified intracranial hemorrhage, with concussion, unspecified | S02.0XXB | Fracture of vault of skull, initial encounter for open fracture |
| 800.89 | Open fracture of vault of skull with other and unspecified intracranial hemorrhage, with concussion, unspecified | S06.360A | Traumatic hemorrhage of cerebrum, unspecified, without loss of consciousness, initial encounter |
| 800.90 | Open fracture of vault of skull with intracranial injury of other and unspecified nature, unspecified state of consciousness | S02.0XXB | Fracture of vault of skull, initial encounter for open fracture |
| 800.90 | Open fracture of vault of skull with intracranial injury of other and unspecified nature, unspecified state of consciousness | S06.890A | Other specified intracranial injury without loss of consciousness, initial encounter |
| 800.90 | Open fracture of vault of skull with intracranial injury of other and unspecified nature, unspecified state of consciousness | S06.9X0A | Unspecified intracranial injury without loss of consciousness, initial encounter |
| 800.91 | Open fracture of vault of skull with intracranial injury of other and unspecified nature, with no loss of consciousness | S02.0XXB | Fracture of vault of skull, initial encounter for open fracture |
| 800.91 | Open fracture of vault of skull with intracranial injury of other and unspecified nature, with no loss of consciousness | S06.890A | Other specified intracranial injury without loss of consciousness, initial encounter |
| 800.91 | Open fracture of vault of skull with intracranial injury of other and unspecified nature, with no loss of consciousness | S06.9X0A | Unspecified intracranial injury without loss of consciousness, initial encounter |
| 800.92 | Open fracture of vault of skull with intracranial injury of other and unspecified nature, with brief [less than one hour] loss of consciousness | S02.0XXB | Fracture of vault of skull, initial encounter for open fracture |
| 800.92 | Open fracture of vault of skull with intracranial injury of other and unspecified nature, with brief [less than one hour] loss of consciousness | S06.891A | Other specified intracranial injury with loss of consciousness of 30 minutes or less, initial encounter |
| 800.92 | Open fracture of vault of skull with intracranial injury of other and unspecified nature, with brief [less than one hour] loss of consciousness | S06.892A | Other specified intracranial injury with loss of consciousness of 31 minutes to 59 minutes, initial encounter |
| 800.92 | Open fracture of vault of skull with intracranial injury of other and unspecified nature, with brief [less than one hour] loss of consciousness | S06.9X1A | Unspecified intracranial injury with loss of consciousness of 30 minutes or less, initial encounter |
| 800.92 | Open fracture of vault of skull with intracranial injury of other and unspecified nature, with brief [less than one hour] loss of consciousness | S06.9X2A | Unspecified intracranial injury with loss of consciousness of 31 minutes to 59 minutes, initial encounter |
| 800.93 | Open fracture of vault of skull with intracranial injury of other and unspecified nature, with moderate [1-24 hours] loss of consciousness | S02.0XXB | Fracture of vault of skull, initial encounter for open fracture |
| 800.93 | Open fracture of vault of skull with intracranial injury of other and unspecified nature, with moderate [1-24 hours] loss of consciousness | S06.893A | Other specified intracranial injury with loss of consciousness of 1 hour to 5 hours 59 minutes, initial encounter |
| 800.93 | Open fracture of vault of skull with intracranial injury of other and unspecified nature, with moderate [1-24 hours] loss of consciousness | S06.894A | Other specified intracranial injury with loss of consciousness of 6 hours to 24 hours, initial encounter |
| 800.93 | Open fracture of vault of skull with intracranial injury of other and unspecified nature, with moderate [1-24 hours] loss of consciousness | S06.9X3A | Unspecified intracranial injury with loss of consciousness of 1 hour to 5 hours 59 minutes, initial encounter |
| 800.93 | Open fracture of vault of skull with intracranial injury of other and unspecified nature, with moderate [1-24 hours] loss of consciousness | S06.9X4A | Unspecified intracranial injury with loss of consciousness of 6 hours to 24 hours, initial encounter |
| 800.94 | Open fracture of vault of skull with intracranial injury of other and unspecified nature, with prolonged [more than 24 hours] loss of consciousness and return to pre-existing conscious level | S02.0XXB | Fracture of vault of skull, initial encounter for open fracture |
| 800.94 | Open fracture of vault of skull with intracranial injury of other and unspecified nature, with prolonged [more than 24 hours] loss of consciousness and return to pre-existing conscious level | S06.895A | Other specified intracranial injury with loss of consciousness greater than 24 hours with return to pre-existing conscious level, initial encounter |
| 800.94 | Open fracture of vault of skull with intracranial injury of other and unspecified nature, with prolonged [more than 24 hours] loss of consciousness and return to pre-existing conscious level | S06.9X5A | Unspecified intracranial injury with loss of consciousness greater than 24 hours with return to pre-existing conscious level, initial encounter |
| 800.95 | Open fracture of vault of skull with intracranial injury of other and unspecified nature, with prolonged [more than 24 hours] loss of consciousness, without return to pre-existing conscious level | S02.0XXB | Fracture of vault of skull, initial encounter for open fracture |
| 800.95 | Open fracture of vault of skull with intracranial injury of other and unspecified nature, with prolonged [more than 24 hours] loss of consciousness, without return to pre-existing conscious level | S06.896A | Other specified intracranial injury with loss of consciousness greater than 24 hours without return to pre-existing conscious level with patient surviving, initial encounter |
| 800.95 | Open fracture of vault of skull with intracranial injury of other and unspecified nature, with prolonged [more than 24 hours] loss of consciousness, without return to pre-existing conscious level | S06.897A | Other specified intracranial injury with loss of consciousness of any duration with death due to brain injury prior to regaining consciousness, initial encounter |
| 800.95 | Open fracture of vault of skull with intracranial injury of other and unspecified nature, with prolonged [more than 24 hours] loss of consciousness, without return to pre-existing conscious level | S06.898A | Other specified intracranial injury with loss of consciousness of any duration with death due to other cause prior to regaining consciousness, initial encounter |
| 800.95 | Open fracture of vault of skull with intracranial injury of other and unspecified nature, with prolonged [more than 24 hours] loss of consciousness, without return to pre-existing conscious level | S06.9X6A | Unspecified intracranial injury with loss of consciousness greater than 24 hours without return to pre-existing conscious level with patient surviving, initial encounter |
| 800.95 | Open fracture of vault of skull with intracranial injury of other and unspecified nature, with prolonged [more than 24 hours] loss of consciousness, without return to pre-existing conscious level | S06.9X7A | Unspecified intracranial injury with loss of consciousness of any duration with death due to brain injury prior to regaining consciousness, initial encounter |
| 800.95 | Open fracture of vault of skull with intracranial injury of other and unspecified nature, with prolonged [more than 24 hours] loss of consciousness, without return to pre-existing conscious level | S06.9X8A | Unspecified intracranial injury with loss of consciousness of any duration with death due to other cause prior to regaining consciousness, initial encounter |
| 800.96 | Open fracture of vault of skull with intracranial injury of other and unspecified nature, with loss of consciousness of unspecified duration | S02.0XXB | Fracture of vault of skull, initial encounter for open fracture |
| 800.96 | Open fracture of vault of skull with intracranial injury of other and unspecified nature, with loss of consciousness of unspecified duration | S06.899A | Other specified intracranial injury with loss of consciousness of unspecified duration, initial encounter |
| 800.96 | Open fracture of vault of skull with intracranial injury of other and unspecified nature, with loss of consciousness of unspecified duration | S06.9X9A | Unspecified intracranial injury with loss of consciousness of unspecified duration, initial encounter |
| 800.99 | Open fracture of vault of skull with intracranial injury of other and unspecified nature, with concussion, unspecified | S02.0XXB | Fracture of vault of skull, initial encounter for open fracture |
| 800.99 | Open fracture of vault of skull with intracranial injury of other and unspecified nature, with concussion, unspecified | S06.890A | Other specified intracranial injury without loss of consciousness, initial encounter |
| 800.99 | Open fracture of vault of skull with intracranial injury of other and unspecified nature, with concussion, unspecified | S06.9X0A | Unspecified intracranial injury without loss of consciousness, initial encounter |
| 801.10 | Closed fracture of base of skull with cerebral laceration and contusion, unspecified state of consciousness | S02.109A | Fracture of base of skull, unspecified side, initial encounter for closed fracture |
| 801.10 | Closed fracture of base of skull with cerebral laceration and contusion, unspecified state of consciousness | S06.330A | Contusion and laceration of cerebrum, unspecified, without loss of consciousness, initial encounter |
| 801.11 | Closed fracture of base of skull with cerebral laceration and contusion, with no loss of consciousness | S02.109A | Fracture of base of skull, unspecified side, initial encounter for closed fracture |
| 801.11 | Closed fracture of base of skull with cerebral laceration and contusion, with no loss of consciousness | S06.330A | Contusion and laceration of cerebrum, unspecified, without loss of consciousness, initial encounter |
| 801.12 | Closed fracture of base of skull with cerebral laceration and contusion, with brief [less than one hour] loss of consciousness | S02.109A | Fracture of base of skull, unspecified side, initial encounter for closed fracture |
| 801.12 | Closed fracture of base of skull with cerebral laceration and contusion, with brief [less than one hour] loss of consciousness | S06.331A | Contusion and laceration of cerebrum, unspecified, with loss of consciousness of 30 minutes or less, initial encounter |
| 801.12 | Closed fracture of base of skull with cerebral laceration and contusion, with brief [less than one hour] loss of consciousness | S06.332A | Contusion and laceration of cerebrum, unspecified, with loss of consciousness of 31 minutes to 59 minutes, initial encounter |
| 801.13 | Closed fracture of base of skull with cerebral laceration and contusion, with moderate [1-24 hours] loss of consciousness | S02.109A | Fracture of base of skull, unspecified side, initial encounter for closed fracture |
| 801.13 | Closed fracture of base of skull with cerebral laceration and contusion, with moderate [1-24 hours] loss of consciousness | S06.333A | Contusion and laceration of cerebrum, unspecified, with loss of consciousness of 1 hour to 5 hours 59 minutes, initial encounter |
| 801.13 | Closed fracture of base of skull with cerebral laceration and contusion, with moderate [1-24 hours] loss of consciousness | S06.334A | Contusion and laceration of cerebrum, unspecified, with loss of consciousness of 6 hours to 24 hours, initial encounter |
| 801.14 | Closed fracture of base of skull with cerebral laceration and contusion, with prolonged [more than 24 hours] loss of consciousness and return to pre-existing conscious level | S02.109A | Fracture of base of skull, unspecified side, initial encounter for closed fracture |
| 801.14 | Closed fracture of base of skull with cerebral laceration and contusion, with prolonged [more than 24 hours] loss of consciousness and return to pre-existing conscious level | S06.335A | Contusion and laceration of cerebrum, unspecified, with loss of consciousness greater than 24 hours with return to pre-existing conscious level, initial encounter |
| 801.15 | Closed fracture of base of skull with cerebral laceration and contusion, with prolonged [more than 24 hours] loss of consciousness, without return to pre-existing conscious level | S02.109A | Fracture of base of skull, unspecified side, initial encounter for closed fracture |
| 801.15 | Closed fracture of base of skull with cerebral laceration and contusion, with prolonged [more than 24 hours] loss of consciousness, without return to pre-existing conscious level | S06.336A | Contusion and laceration of cerebrum, unspecified, with loss of consciousness greater than 24 hours without return to pre-existing conscious level with patient surviving, initial encounter |
| 801.15 | Closed fracture of base of skull with cerebral laceration and contusion, with prolonged [more than 24 hours] loss of consciousness, without return to pre-existing conscious level | S06.337A | Contusion and laceration of cerebrum, unspecified, with loss of consciousness of any duration with death due to brain injury prior to regaining consciousness, initial encounter |
| 801.15 | Closed fracture of base of skull with cerebral laceration and contusion, with prolonged [more than 24 hours] loss of consciousness, without return to pre-existing conscious level | S06.338A | Contusion and laceration of cerebrum, unspecified, with loss of consciousness of any duration with death due to other cause prior to regaining consciousness, initial encounter |
| 801.16 | Closed fracture of base of skull with cerebral laceration and contusion, with loss of consciousness of unspecified duration | S02.109A | Fracture of base of skull, unspecified side, initial encounter for closed fracture |
| 801.16 | Closed fracture of base of skull with cerebral laceration and contusion, with loss of consciousness of unspecified duration | S06.339A | Contusion and laceration of cerebrum, unspecified, with loss of consciousness of unspecified duration, initial encounter |
| 801.19 | Closed fracture of base of skull with cerebral laceration and contusion, with concussion, unspecified | S02.109A | Fracture of base of skull, unspecified side, initial encounter for closed fracture |
| 801.19 | Closed fracture of base of skull with cerebral laceration and contusion, with concussion, unspecified | S06.330A | Contusion and laceration of cerebrum, unspecified, without loss of consciousness, initial encounter |
| 801.20 | Closed fracture of base of skull with subarachnoid, subdural, and extradural hemorrhage, unspecified state of consciousness | S06.4X0A | Epidural hemorrhage without loss of consciousness, initial encounter |
| 801.20 | Closed fracture of base of skull with subarachnoid, subdural, and extradural hemorrhage, unspecified state of consciousness | S06.5X0A | Traumatic subdural hemorrhage without loss of consciousness, initial encounter |
| 801.20 | Closed fracture of base of skull with subarachnoid, subdural, and extradural hemorrhage, unspecified state of consciousness | S06.6X0A | Traumatic subarachnoid hemorrhage without loss of consciousness, initial encounter |
| 801.20 | Closed fracture of base of skull with subarachnoid, subdural, and extradural hemorrhage, unspecified state of consciousness |  |  |
| 801.21 | Closed fracture of base of skull with subarachnoid, subdural, and extradural hemorrhage, with no loss of consciousness | S06.4X0A | Epidural hemorrhage without loss of consciousness, initial encounter |
| 801.21 | Closed fracture of base of skull with subarachnoid, subdural, and extradural hemorrhage, with no loss of consciousness | S06.5X0A | Traumatic subdural hemorrhage without loss of consciousness, initial encounter |
| 801.21 | Closed fracture of base of skull with subarachnoid, subdural, and extradural hemorrhage, with no loss of consciousness | S06.6X0A | Traumatic subarachnoid hemorrhage without loss of consciousness, initial encounter |
| 801.21 | Closed fracture of base of skull with subarachnoid, subdural, and extradural hemorrhage, with no loss of consciousness |  |  |
| 801.22 | Closed fracture of base of skull with subarachnoid, subdural, and extradural hemorrhage, with brief [less than one hour] loss of consciousness | S06.4X1A | Epidural hemorrhage with loss of consciousness of 30 minutes or less, initial encounter |
| 801.22 | Closed fracture of base of skull with subarachnoid, subdural, and extradural hemorrhage, with brief [less than one hour] loss of consciousness | S06.4X2A | Epidural hemorrhage with loss of consciousness of 31 minutes to 59 minutes, initial encounter |
| 801.22 | Closed fracture of base of skull with subarachnoid, subdural, and extradural hemorrhage, with brief [less than one hour] loss of consciousness | S06.5X1A | Traumatic subdural hemorrhage with loss of consciousness of 30 minutes or less, initial encounter |
| 801.22 | Closed fracture of base of skull with subarachnoid, subdural, and extradural hemorrhage, with brief [less than one hour] loss of consciousness | S06.5X2A | Traumatic subdural hemorrhage with loss of consciousness of 31 minutes to 59 minutes, initial encounter |
| 801.22 | Closed fracture of base of skull with subarachnoid, subdural, and extradural hemorrhage, with brief [less than one hour] loss of consciousness | S06.6X1A | Traumatic subarachnoid hemorrhage with loss of consciousness of 30 minutes or less, initial encounter |
| 801.22 | Closed fracture of base of skull with subarachnoid, subdural, and extradural hemorrhage, with brief [less than one hour] loss of consciousness | S06.6X2A | Traumatic subarachnoid hemorrhage with loss of consciousness of 31 minutes to 59 minutes, initial encounter |
| 801.22 | Closed fracture of base of skull with subarachnoid, subdural, and extradural hemorrhage, with brief [less than one hour] loss of consciousness |  |  |
| 801.23 | Closed fracture of base of skull with subarachnoid, subdural, and extradural hemorrhage, with moderate [1-24 hours] loss of consciousness | S06.4X3A | Epidural hemorrhage with loss of consciousness of 1 hour to 5 hours 59 minutes, initial encounter |
| 801.23 | Closed fracture of base of skull with subarachnoid, subdural, and extradural hemorrhage, with moderate [1-24 hours] loss of consciousness | S06.4X4A | Epidural hemorrhage with loss of consciousness of 6 hours to 24 hours, initial encounter |
| 801.23 | Closed fracture of base of skull with subarachnoid, subdural, and extradural hemorrhage, with moderate [1-24 hours] loss of consciousness | S06.5X3A | Traumatic subdural hemorrhage with loss of consciousness of 1 hour to 5 hours 59 minutes, initial encounter |
| 801.23 | Closed fracture of base of skull with subarachnoid, subdural, and extradural hemorrhage, with moderate [1-24 hours] loss of consciousness | S06.5X4A | Traumatic subdural hemorrhage with loss of consciousness of 6 hours to 24 hours, initial encounter |
| 801.23 | Closed fracture of base of skull with subarachnoid, subdural, and extradural hemorrhage, with moderate [1-24 hours] loss of consciousness | S06.6X3A | Traumatic subarachnoid hemorrhage with loss of consciousness of 1 hour to 5 hours 59 minutes, initial encounter |
| 801.23 | Closed fracture of base of skull with subarachnoid, subdural, and extradural hemorrhage, with moderate [1-24 hours] loss of consciousness | S06.6X4A | Traumatic subarachnoid hemorrhage with loss of consciousness of 6 hours to 24 hours, initial encounter |
| 801.23 | Closed fracture of base of skull with subarachnoid, subdural, and extradural hemorrhage, with moderate [1-24 hours] loss of consciousness |  |  |
| 801.24 | Closed fracture of base of skull with subarachnoid, subdural, and extradural hemorrhage, with prolonged [more than 24 hours] loss of consciousness and return to pre-existing conscious level | S06.4X5A | Epidural hemorrhage with loss of consciousness greater than 24 hours with return to pre-existing conscious level, initial encounter |
| 801.24 | Closed fracture of base of skull with subarachnoid, subdural, and extradural hemorrhage, with prolonged [more than 24 hours] loss of consciousness and return to pre-existing conscious level | S06.5X5A | Traumatic subdural hemorrhage with loss of consciousness greater than 24 hours with return to pre-existing conscious level, initial encounter |
| 801.24 | Closed fracture of base of skull with subarachnoid, subdural, and extradural hemorrhage, with prolonged [more than 24 hours] loss of consciousness and return to pre-existing conscious level | S06.6X5A | Traumatic subarachnoid hemorrhage with loss of consciousness greater than 24 hours with return to pre-existing conscious level, initial encounter |
| 801.24 | Closed fracture of base of skull with subarachnoid, subdural, and extradural hemorrhage, with prolonged [more than 24 hours] loss of consciousness and return to pre-existing conscious level |  |  |
| 801.25 | Closed fracture of base of skull with subarachnoid, subdural, and extradural hemorrhage, with prolonged [more than 24 hours] loss of consciousness, without return to pre-existing conscious level | S06.4X6A | Epidural hemorrhage with loss of consciousness greater than 24 hours without return to pre-existing conscious level with patient surviving, initial encounter |
| 801.25 | Closed fracture of base of skull with subarachnoid, subdural, and extradural hemorrhage, with prolonged [more than 24 hours] loss of consciousness, without return to pre-existing conscious level | S06.4X7A | Epidural hemorrhage with loss of consciousness of any duration with death due to brain injury prior to regaining consciousness, initial encounter |
| 801.25 | Closed fracture of base of skull with subarachnoid, subdural, and extradural hemorrhage, with prolonged [more than 24 hours] loss of consciousness, without return to pre-existing conscious level | S06.4X8A | Epidural hemorrhage with loss of consciousness of any duration with death due to other causes prior to regaining consciousness, initial encounter |
| 801.25 | Closed fracture of base of skull with subarachnoid, subdural, and extradural hemorrhage, with prolonged [more than 24 hours] loss of consciousness, without return to pre-existing conscious level | S06.5X6A | Traumatic subdural hemorrhage with loss of consciousness greater than 24 hours without return to pre-existing conscious level with patient surviving, initial encounter |
| 801.25 | Closed fracture of base of skull with subarachnoid, subdural, and extradural hemorrhage, with prolonged [more than 24 hours] loss of consciousness, without return to pre-existing conscious level | S06.5X7A | Traumatic subdural hemorrhage with loss of consciousness of any duration with death due to brain injury before regaining consciousness, initial encounter |
| 801.25 | Closed fracture of base of skull with subarachnoid, subdural, and extradural hemorrhage, with prolonged [more than 24 hours] loss of consciousness, without return to pre-existing conscious level | S06.5X8A | Traumatic subdural hemorrhage with loss of consciousness of any duration with death due to other cause before regaining consciousness, initial encounter |
| 801.25 | Closed fracture of base of skull with subarachnoid, subdural, and extradural hemorrhage, with prolonged [more than 24 hours] loss of consciousness, without return to pre-existing conscious level | S06.6X6A | Traumatic subarachnoid hemorrhage with loss of consciousness greater than 24 hours without return to pre-existing conscious level with patient surviving, initial encounter |
| 801.25 | Closed fracture of base of skull with subarachnoid, subdural, and extradural hemorrhage, with prolonged [more than 24 hours] loss of consciousness, without return to pre-existing conscious level | S06.6X7A | Traumatic subarachnoid hemorrhage with loss of consciousness of any duration with death due to brain injury prior to regaining consciousness, initial encounter |
| 801.25 | Closed fracture of base of skull with subarachnoid, subdural, and extradural hemorrhage, with prolonged [more than 24 hours] loss of consciousness, without return to pre-existing conscious level | S06.6X8A | Traumatic subarachnoid hemorrhage with loss of consciousness of any duration with death due to other cause prior to regaining consciousness, initial encounter |
| 801.25 | Closed fracture of base of skull with subarachnoid, subdural, and extradural hemorrhage, with prolonged [more than 24 hours] loss of consciousness, without return to pre-existing conscious level |  |  |
| 801.26 | Closed fracture of base of skull with subarachnoid, subdural, and extradural hemorrhage, with loss of consciousness of unspecified duration | S06.4X9A | Epidural hemorrhage with loss of consciousness of unspecified duration, initial encounter |
| 801.26 | Closed fracture of base of skull with subarachnoid, subdural, and extradural hemorrhage, with loss of consciousness of unspecified duration | S06.5X9A | Traumatic subdural hemorrhage with loss of consciousness of unspecified duration, initial encounter |
| 801.26 | Closed fracture of base of skull with subarachnoid, subdural, and extradural hemorrhage, with loss of consciousness of unspecified duration | S06.6X9A | Traumatic subarachnoid hemorrhage with loss of consciousness of unspecified duration, initial encounter |
| 801.26 | Closed fracture of base of skull with subarachnoid, subdural, and extradural hemorrhage, with loss of consciousness of unspecified duration |  |  |
| 801.29 | Closed fracture of base of skull with subarachnoid, subdural, and extradural hemorrhage, with concussion, unspecified | S06.4X0A | Epidural hemorrhage without loss of consciousness, initial encounter |
| 801.29 | Closed fracture of base of skull with subarachnoid, subdural, and extradural hemorrhage, with concussion, unspecified | S06.5X0A | Traumatic subdural hemorrhage without loss of consciousness, initial encounter |
| 801.29 | Closed fracture of base of skull with subarachnoid, subdural, and extradural hemorrhage, with concussion, unspecified | S06.6X0A | Traumatic subarachnoid hemorrhage without loss of consciousness, initial encounter |
| 801.29 | Closed fracture of base of skull with subarachnoid, subdural, and extradural hemorrhage, with concussion, unspecified |  |  |
| 801.30 | Closed fracture of base of skull with other and unspecified intracranial hemorrhage, unspecified state of consciousness | S02.109A | Fracture of base of skull, unspecified side, initial encounter for closed fracture |
| 801.30 | Closed fracture of base of skull with other and unspecified intracranial hemorrhage, unspecified state of consciousness | S06.360A | Traumatic hemorrhage of cerebrum, unspecified, without loss of consciousness, initial encounter |
| 801.31 | Closed fracture of base of skull with other and unspecified intracranial hemorrhage, with no loss of consciousness | S02.109A | Fracture of base of skull, unspecified side, initial encounter for closed fracture |
| 801.31 | Closed fracture of base of skull with other and unspecified intracranial hemorrhage, with no loss of consciousness | S06.360A | Traumatic hemorrhage of cerebrum, unspecified, without loss of consciousness, initial encounter |
| 801.32 | Closed fracture of base of skull with other and unspecified intracranial hemorrhage, with brief [less than one hour] loss of consciousness | S02.109A | Fracture of base of skull, unspecified side, initial encounter for closed fracture |
| 801.32 | Closed fracture of base of skull with other and unspecified intracranial hemorrhage, with brief [less than one hour] loss of consciousness | S06.361A | Traumatic hemorrhage of cerebrum, unspecified, with loss of consciousness of 30 minutes or less, initial encounter |
| 801.32 | Closed fracture of base of skull with other and unspecified intracranial hemorrhage, with brief [less than one hour] loss of consciousness | S06.362A | Traumatic hemorrhage of cerebrum, unspecified, with loss of consciousness of 31 minutes to 59 minutes, initial encounter |
| 801.33 | Closed fracture of base of skull with other and unspecified intracranial hemorrhage, with moderate [1-24 hours] loss of consciousness | S02.109A | Fracture of base of skull, unspecified side, initial encounter for closed fracture |
| 801.33 | Closed fracture of base of skull with other and unspecified intracranial hemorrhage, with moderate [1-24 hours] loss of consciousness | S06.363A | Traumatic hemorrhage of cerebrum, unspecified, with loss of consciousness of 1 hours to 5 hours 59 minutes, initial encounter |
| 801.33 | Closed fracture of base of skull with other and unspecified intracranial hemorrhage, with moderate [1-24 hours] loss of consciousness | S06.364A | Traumatic hemorrhage of cerebrum, unspecified, with loss of consciousness of 6 hours to 24 hours, initial encounter |
| 801.34 | Closed fracture of base of skull with other and unspecified intracranial hemorrhage, with prolonged [more than 24 hours] loss of consciousness and return to pre-existing conscious level | S02.109A | Fracture of base of skull, unspecified side, initial encounter for closed fracture |
| 801.34 | Closed fracture of base of skull with other and unspecified intracranial hemorrhage, with prolonged [more than 24 hours] loss of consciousness and return to pre-existing conscious level | S06.365A | Traumatic hemorrhage of cerebrum, unspecified, with loss of consciousness greater than 24 hours with return to pre-existing conscious level, initial encounter |
| 801.35 | Closed fracture of base of skull with other and unspecified intracranial hemorrhage, with prolonged [more than 24 hours] loss of consciousness, without return to pre-existing conscious level | S02.109A | Fracture of base of skull, unspecified side, initial encounter for closed fracture |
| 801.35 | Closed fracture of base of skull with other and unspecified intracranial hemorrhage, with prolonged [more than 24 hours] loss of consciousness, without return to pre-existing conscious level | S06.366A | Traumatic hemorrhage of cerebrum, unspecified, with loss of consciousness greater than 24 hours without return to pre-existing conscious level with patient surviving, initial encounter |
| 801.35 | Closed fracture of base of skull with other and unspecified intracranial hemorrhage, with prolonged [more than 24 hours] loss of consciousness, without return to pre-existing conscious level | S06.367A | Traumatic hemorrhage of cerebrum, unspecified, with loss of consciousness of any duration with death due to brain injury prior to regaining consciousness, initial encounter |
| 801.35 | Closed fracture of base of skull with other and unspecified intracranial hemorrhage, with prolonged [more than 24 hours] loss of consciousness, without return to pre-existing conscious level | S06.368A | Traumatic hemorrhage of cerebrum, unspecified, with loss of consciousness of any duration with death due to other cause prior to regaining consciousness, initial encounter |
| 801.36 | Closed fracture of base of skull with other and unspecified intracranial hemorrhage, with loss of consciousness of unspecified duration | S02.109A | Fracture of base of skull, unspecified side, initial encounter for closed fracture |
| 801.36 | Closed fracture of base of skull with other and unspecified intracranial hemorrhage, with loss of consciousness of unspecified duration | S06.369A | Traumatic hemorrhage of cerebrum, unspecified, with loss of consciousness of unspecified duration, initial encounter |
| 801.39 | Closed fracture of base of skull with other and unspecified intracranial hemorrhage, with concussion, unspecified | S02.109A | Fracture of base of skull, unspecified side, initial encounter for closed fracture |
| 801.39 | Closed fracture of base of skull with other and unspecified intracranial hemorrhage, with concussion, unspecified | S06.360A | Traumatic hemorrhage of cerebrum, unspecified, without loss of consciousness, initial encounter |
| 801.40 | Closed fracture of base of skull with intracranial injury of other and unspecified nature, unspecified state of consciousness | S02.109A | Fracture of base of skull, unspecified side, initial encounter for closed fracture |
| 801.40 | Closed fracture of base of skull with intracranial injury of other and unspecified nature, unspecified state of consciousness | S06.890A | Other specified intracranial injury without loss of consciousness, initial encounter |
| 801.40 | Closed fracture of base of skull with intracranial injury of other and unspecified nature, unspecified state of consciousness | S06.9X0A | Unspecified intracranial injury without loss of consciousness, initial encounter |
| 801.41 | Closed fracture of base of skull with intracranial injury of other and unspecified nature, with no loss of consciousness | S02.109A | Fracture of base of skull, unspecified side, initial encounter for closed fracture |
| 801.41 | Closed fracture of base of skull with intracranial injury of other and unspecified nature, with no loss of consciousness | S06.890A | Other specified intracranial injury without loss of consciousness, initial encounter |
| 801.41 | Closed fracture of base of skull with intracranial injury of other and unspecified nature, with no loss of consciousness | S06.9X0A | Unspecified intracranial injury without loss of consciousness, initial encounter |
| 801.42 | Closed fracture of base of skull with intracranial injury of other and unspecified nature, with brief [less than one hour] loss of consciousness | S02.109A | Fracture of base of skull, unspecified side, initial encounter for closed fracture |
| 801.42 | Closed fracture of base of skull with intracranial injury of other and unspecified nature, with brief [less than one hour] loss of consciousness | S06.891A | Other specified intracranial injury with loss of consciousness of 30 minutes or less, initial encounter |
| 801.42 | Closed fracture of base of skull with intracranial injury of other and unspecified nature, with brief [less than one hour] loss of consciousness | S06.892A | Other specified intracranial injury with loss of consciousness of 31 minutes to 59 minutes, initial encounter |
| 801.42 | Closed fracture of base of skull with intracranial injury of other and unspecified nature, with brief [less than one hour] loss of consciousness | S06.9X1A | Unspecified intracranial injury with loss of consciousness of 30 minutes or less, initial encounter |
| 801.42 | Closed fracture of base of skull with intracranial injury of other and unspecified nature, with brief [less than one hour] loss of consciousness | S06.9X2A | Unspecified intracranial injury with loss of consciousness of 31 minutes to 59 minutes, initial encounter |
| 801.43 | Closed fracture of base of skull with intracranial injury of other and unspecified nature, with moderate [1-24 hours] loss of consciousness | S02.109A | Fracture of base of skull, unspecified side, initial encounter for closed fracture |
| 801.43 | Closed fracture of base of skull with intracranial injury of other and unspecified nature, with moderate [1-24 hours] loss of consciousness | S06.893A | Other specified intracranial injury with loss of consciousness of 1 hour to 5 hours 59 minutes, initial encounter |
| 801.43 | Closed fracture of base of skull with intracranial injury of other and unspecified nature, with moderate [1-24 hours] loss of consciousness | S06.894A | Other specified intracranial injury with loss of consciousness of 6 hours to 24 hours, initial encounter |
| 801.43 | Closed fracture of base of skull with intracranial injury of other and unspecified nature, with moderate [1-24 hours] loss of consciousness | S06.9X3A | Unspecified intracranial injury with loss of consciousness of 1 hour to 5 hours 59 minutes, initial encounter |
| 801.43 | Closed fracture of base of skull with intracranial injury of other and unspecified nature, with moderate [1-24 hours] loss of consciousness | S06.9X4A | Unspecified intracranial injury with loss of consciousness of 6 hours to 24 hours, initial encounter |
| 801.44 | Closed fracture of base of skull with intracranial injury of other and unspecified nature, with prolonged [more than 24 hours) loss of consciousness and return to pre-existing conscious level | S02.109A | Fracture of base of skull, unspecified side, initial encounter for closed fracture |
| 801.44 | Closed fracture of base of skull with intracranial injury of other and unspecified nature, with prolonged [more than 24 hours) loss of consciousness and return to pre-existing conscious level | S06.895A | Other specified intracranial injury with loss of consciousness greater than 24 hours with return to pre-existing conscious level, initial encounter |
| 801.44 | Closed fracture of base of skull with intracranial injury of other and unspecified nature, with prolonged [more than 24 hours) loss of consciousness and return to pre-existing conscious level | S06.9X5A | Unspecified intracranial injury with loss of consciousness greater than 24 hours with return to pre-existing conscious level, initial encounter |
| 801.45 | Closed fracture of base of skull with intracranial injury of other and unspecified nature, with prolonged [more than 24 hours] loss of consciousness, without return to pre-existing conscious level | S02.109A | Fracture of base of skull, unspecified side, initial encounter for closed fracture |
| 801.45 | Closed fracture of base of skull with intracranial injury of other and unspecified nature, with prolonged [more than 24 hours] loss of consciousness, without return to pre-existing conscious level | S06.896A | Other specified intracranial injury with loss of consciousness greater than 24 hours without return to pre-existing conscious level with patient surviving, initial encounter |
| 801.45 | Closed fracture of base of skull with intracranial injury of other and unspecified nature, with prolonged [more than 24 hours] loss of consciousness, without return to pre-existing conscious level | S06.897A | Other specified intracranial injury with loss of consciousness of any duration with death due to brain injury prior to regaining consciousness, initial encounter |
| 801.45 | Closed fracture of base of skull with intracranial injury of other and unspecified nature, with prolonged [more than 24 hours] loss of consciousness, without return to pre-existing conscious level | S06.898A | Other specified intracranial injury with loss of consciousness of any duration with death due to other cause prior to regaining consciousness, initial encounter |
| 801.45 | Closed fracture of base of skull with intracranial injury of other and unspecified nature, with prolonged [more than 24 hours] loss of consciousness, without return to pre-existing conscious level | S06.9X6A | Unspecified intracranial injury with loss of consciousness greater than 24 hours without return to pre-existing conscious level with patient surviving, initial encounter |
| 801.45 | Closed fracture of base of skull with intracranial injury of other and unspecified nature, with prolonged [more than 24 hours] loss of consciousness, without return to pre-existing conscious level | S06.9X7A | Unspecified intracranial injury with loss of consciousness of any duration with death due to brain injury prior to regaining consciousness, initial encounter |
| 801.45 | Closed fracture of base of skull with intracranial injury of other and unspecified nature, with prolonged [more than 24 hours] loss of consciousness, without return to pre-existing conscious level | S06.9X8A | Unspecified intracranial injury with loss of consciousness of any duration with death due to other cause prior to regaining consciousness, initial encounter |
| 801.46 | Closed fracture of base of skull with intracranial injury of other and unspecified nature, with loss of consciousness of unspecified duration | S02.109A | Fracture of base of skull, unspecified side, initial encounter for closed fracture |
| 801.46 | Closed fracture of base of skull with intracranial injury of other and unspecified nature, with loss of consciousness of unspecified duration | S06.899A | Other specified intracranial injury with loss of consciousness of unspecified duration, initial encounter |
| 801.46 | Closed fracture of base of skull with intracranial injury of other and unspecified nature, with loss of consciousness of unspecified duration | S06.9X9A | Unspecified intracranial injury with loss of consciousness of unspecified duration, initial encounter |
| 801.49 | Closed fracture of base of skull with intracranial injury of other and unspecified nature, with concussion, unspecified | S02.109A | Fracture of base of skull, unspecified side, initial encounter for closed fracture |
| 801.49 | Closed fracture of base of skull with intracranial injury of other and unspecified nature, with concussion, unspecified | S06.890A | Other specified intracranial injury without loss of consciousness, initial encounter |
| 801.49 | Closed fracture of base of skull with intracranial injury of other and unspecified nature, with concussion, unspecified | S06.9X0A | Unspecified intracranial injury without loss of consciousness, initial encounter |
| 801.60 | Open fracture of base of skull with cerebral laceration and contusion, unspecified state of consciousness | S02.109B | Fracture of base of skull, unspecified side, initial encounter for open fracture |
| 801.60 | Open fracture of base of skull with cerebral laceration and contusion, unspecified state of consciousness | S06.330A | Contusion and laceration of cerebrum, unspecified, without loss of consciousness, initial encounter |
| 801.61 | Open fracture of base of skull with cerebral laceration and contusion, with no loss of consciousness | S02.109B | Fracture of base of skull, unspecified side, initial encounter for open fracture |
| 801.61 | Open fracture of base of skull with cerebral laceration and contusion, with no loss of consciousness | S06.330A | Contusion and laceration of cerebrum, unspecified, without loss of consciousness, initial encounter |
| 801.62 | Open fracture of base of skull with cerebral laceration and contusion, with brief [less than one hour] loss of consciousness | S02.109B | Fracture of base of skull, unspecified side, initial encounter for open fracture |
| 801.62 | Open fracture of base of skull with cerebral laceration and contusion, with brief [less than one hour] loss of consciousness | S06.331A | Contusion and laceration of cerebrum, unspecified, with loss of consciousness of 30 minutes or less, initial encounter |
| 801.62 | Open fracture of base of skull with cerebral laceration and contusion, with brief [less than one hour] loss of consciousness | S06.332A | Contusion and laceration of cerebrum, unspecified, with loss of consciousness of 31 minutes to 59 minutes, initial encounter |
| 801.63 | Open fracture of base of skull with cerebral laceration and contusion, with moderate [1-24 hours] loss of consciousness | S02.109B | Fracture of base of skull, unspecified side, initial encounter for open fracture |
| 801.63 | Open fracture of base of skull with cerebral laceration and contusion, with moderate [1-24 hours] loss of consciousness | S06.333A | Contusion and laceration of cerebrum, unspecified, with loss of consciousness of 1 hour to 5 hours 59 minutes, initial encounter |
| 801.63 | Open fracture of base of skull with cerebral laceration and contusion, with moderate [1-24 hours] loss of consciousness | S06.334A | Contusion and laceration of cerebrum, unspecified, with loss of consciousness of 6 hours to 24 hours, initial encounter |
| 801.64 | Open fracture of base of skull with cerebral laceration and contusion, with prolonged [more than 24 hours] loss of consciousness and return to pre-existing conscious level | S02.109B | Fracture of base of skull, unspecified side, initial encounter for open fracture |
| 801.64 | Open fracture of base of skull with cerebral laceration and contusion, with prolonged [more than 24 hours] loss of consciousness and return to pre-existing conscious level | S06.335A | Contusion and laceration of cerebrum, unspecified, with loss of consciousness greater than 24 hours with return to pre-existing conscious level, initial encounter |
| 801.65 | Open fracture of base of skull with cerebral laceration and contusion, with prolonged [more than 24 hours] loss of consciousness, without return to pre-existing conscious level | S02.109B | Fracture of base of skull, unspecified side, initial encounter for open fracture |
| 801.65 | Open fracture of base of skull with cerebral laceration and contusion, with prolonged [more than 24 hours] loss of consciousness, without return to pre-existing conscious level | S06.336A | Contusion and laceration of cerebrum, unspecified, with loss of consciousness greater than 24 hours without return to pre-existing conscious level with patient surviving, initial encounter |
| 801.65 | Open fracture of base of skull with cerebral laceration and contusion, with prolonged [more than 24 hours] loss of consciousness, without return to pre-existing conscious level | S06.337A | Contusion and laceration of cerebrum, unspecified, with loss of consciousness of any duration with death due to brain injury prior to regaining consciousness, initial encounter |
| 801.65 | Open fracture of base of skull with cerebral laceration and contusion, with prolonged [more than 24 hours] loss of consciousness, without return to pre-existing conscious level | S06.338A | Contusion and laceration of cerebrum, unspecified, with loss of consciousness of any duration with death due to other cause prior to regaining consciousness, initial encounter |
| 801.66 | Open fracture of base of skull with cerebral laceration and contusion, with loss of consciousness of unspecified duration | S02.109B | Fracture of base of skull, unspecified side, initial encounter for open fracture |
| 801.66 | Open fracture of base of skull with cerebral laceration and contusion, with loss of consciousness of unspecified duration | S06.339A | Contusion and laceration of cerebrum, unspecified, with loss of consciousness of unspecified duration, initial encounter |
| 801.69 | Open fracture of base of skull with cerebral laceration and contusion, with concussion, unspecified | S02.109B | Fracture of base of skull, unspecified side, initial encounter for open fracture |
| 801.69 | Open fracture of base of skull with cerebral laceration and contusion, with concussion, unspecified | S06.330A | Contusion and laceration of cerebrum, unspecified, without loss of consciousness, initial encounter |
| 801.70 | Open fracture of base of skull with subarachnoid, subdural, and extradural hemorrhage, unspecified state of consciousness | S06.4X0A | Epidural hemorrhage without loss of consciousness, initial encounter |
| 801.70 | Open fracture of base of skull with subarachnoid, subdural, and extradural hemorrhage, unspecified state of consciousness | S06.5X0A | Traumatic subdural hemorrhage without loss of consciousness, initial encounter |
| 801.70 | Open fracture of base of skull with subarachnoid, subdural, and extradural hemorrhage, unspecified state of consciousness | S06.6X0A | Traumatic subarachnoid hemorrhage without loss of consciousness, initial encounter |
| 801.70 | Open fracture of base of skull with subarachnoid, subdural, and extradural hemorrhage, unspecified state of consciousness |  |  |
| 801.71 | Open fracture of base of skull with subarachnoid, subdural, and extradural hemorrhage, with no loss of consciousness | S06.4X0A | Epidural hemorrhage without loss of consciousness, initial encounter |
| 801.71 | Open fracture of base of skull with subarachnoid, subdural, and extradural hemorrhage, with no loss of consciousness | S06.5X0A | Traumatic subdural hemorrhage without loss of consciousness, initial encounter |
| 801.71 | Open fracture of base of skull with subarachnoid, subdural, and extradural hemorrhage, with no loss of consciousness | S06.6X0A | Traumatic subarachnoid hemorrhage without loss of consciousness, initial encounter |
| 801.71 | Open fracture of base of skull with subarachnoid, subdural, and extradural hemorrhage, with no loss of consciousness |  |  |
| 801.72 | Open fracture of base of skull with subarachnoid, subdural, and extradural hemorrhage, with brief [less than one hour] loss of consciousness | S06.4X1A | Epidural hemorrhage with loss of consciousness of 30 minutes or less, initial encounter |
| 801.72 | Open fracture of base of skull with subarachnoid, subdural, and extradural hemorrhage, with brief [less than one hour] loss of consciousness | S06.4X2A | Epidural hemorrhage with loss of consciousness of 31 minutes to 59 minutes, initial encounter |
| 801.72 | Open fracture of base of skull with subarachnoid, subdural, and extradural hemorrhage, with brief [less than one hour] loss of consciousness | S06.5X1A | Traumatic subdural hemorrhage with loss of consciousness of 30 minutes or less, initial encounter |
| 801.72 | Open fracture of base of skull with subarachnoid, subdural, and extradural hemorrhage, with brief [less than one hour] loss of consciousness | S06.5X2A | Traumatic subdural hemorrhage with loss of consciousness of 31 minutes to 59 minutes, initial encounter |
| 801.72 | Open fracture of base of skull with subarachnoid, subdural, and extradural hemorrhage, with brief [less than one hour] loss of consciousness | S06.6X1A | Traumatic subarachnoid hemorrhage with loss of consciousness of 30 minutes or less, initial encounter |
| 801.72 | Open fracture of base of skull with subarachnoid, subdural, and extradural hemorrhage, with brief [less than one hour] loss of consciousness | S06.6X2A | Traumatic subarachnoid hemorrhage with loss of consciousness of 31 minutes to 59 minutes, initial encounter |
| 801.72 | Open fracture of base of skull with subarachnoid, subdural, and extradural hemorrhage, with brief [less than one hour] loss of consciousness |  |  |
| 801.73 | Open fracture of base of skull with subarachnoid, subdural, and extradural hemorrhage, with moderate [1-24 hours] loss of consciousness | S06.4X3A | Epidural hemorrhage with loss of consciousness of 1 hour to 5 hours 59 minutes, initial encounter |
| 801.73 | Open fracture of base of skull with subarachnoid, subdural, and extradural hemorrhage, with moderate [1-24 hours] loss of consciousness | S06.4X4A | Epidural hemorrhage with loss of consciousness of 6 hours to 24 hours, initial encounter |
| 801.73 | Open fracture of base of skull with subarachnoid, subdural, and extradural hemorrhage, with moderate [1-24 hours] loss of consciousness | S06.5X3A | Traumatic subdural hemorrhage with loss of consciousness of 1 hour to 5 hours 59 minutes, initial encounter |
| 801.73 | Open fracture of base of skull with subarachnoid, subdural, and extradural hemorrhage, with moderate [1-24 hours] loss of consciousness | S06.5X4A | Traumatic subdural hemorrhage with loss of consciousness of 6 hours to 24 hours, initial encounter |
| 801.73 | Open fracture of base of skull with subarachnoid, subdural, and extradural hemorrhage, with moderate [1-24 hours] loss of consciousness | S06.6X3A | Traumatic subarachnoid hemorrhage with loss of consciousness of 1 hour to 5 hours 59 minutes, initial encounter |
| 801.73 | Open fracture of base of skull with subarachnoid, subdural, and extradural hemorrhage, with moderate [1-24 hours] loss of consciousness | S06.6X4A | Traumatic subarachnoid hemorrhage with loss of consciousness of 6 hours to 24 hours, initial encounter |
| 801.73 | Open fracture of base of skull with subarachnoid, subdural, and extradural hemorrhage, with moderate [1-24 hours] loss of consciousness |  |  |
| 801.74 | Open fracture of base of skull with subarachnoid, subdural, and extradural hemorrhage, with prolonged [more than 24 hours] loss of consciousness and return to pre-existing conscious level | S06.4X5A | Epidural hemorrhage with loss of consciousness greater than 24 hours with return to pre-existing conscious level, initial encounter |
| 801.74 | Open fracture of base of skull with subarachnoid, subdural, and extradural hemorrhage, with prolonged [more than 24 hours] loss of consciousness and return to pre-existing conscious level | S06.5X5A | Traumatic subdural hemorrhage with loss of consciousness greater than 24 hours with return to pre-existing conscious level, initial encounter |
| 801.74 | Open fracture of base of skull with subarachnoid, subdural, and extradural hemorrhage, with prolonged [more than 24 hours] loss of consciousness and return to pre-existing conscious level | S06.6X5A | Traumatic subarachnoid hemorrhage with loss of consciousness greater than 24 hours with return to pre-existing conscious level, initial encounter |
| 801.74 | Open fracture of base of skull with subarachnoid, subdural, and extradural hemorrhage, with prolonged [more than 24 hours] loss of consciousness and return to pre-existing conscious level |  |  |
| 801.75 | Open fracture of base of skull with subarachnoid, subdural, and extradural hemorrhage, with prolonged [more than 24 hours] loss of consciousness, without return to pre-existing conscious level | S06.4X6A | Epidural hemorrhage with loss of consciousness greater than 24 hours without return to pre-existing conscious level with patient surviving, initial encounter |
| 801.75 | Open fracture of base of skull with subarachnoid, subdural, and extradural hemorrhage, with prolonged [more than 24 hours] loss of consciousness, without return to pre-existing conscious level | S06.4X7A | Epidural hemorrhage with loss of consciousness of any duration with death due to brain injury prior to regaining consciousness, initial encounter |
| 801.75 | Open fracture of base of skull with subarachnoid, subdural, and extradural hemorrhage, with prolonged [more than 24 hours] loss of consciousness, without return to pre-existing conscious level | S06.4X8A | Epidural hemorrhage with loss of consciousness of any duration with death due to other causes prior to regaining consciousness, initial encounter |
| 801.75 | Open fracture of base of skull with subarachnoid, subdural, and extradural hemorrhage, with prolonged [more than 24 hours] loss of consciousness, without return to pre-existing conscious level | S06.5X6A | Traumatic subdural hemorrhage with loss of consciousness greater than 24 hours without return to pre-existing conscious level with patient surviving, initial encounter |
| 801.75 | Open fracture of base of skull with subarachnoid, subdural, and extradural hemorrhage, with prolonged [more than 24 hours] loss of consciousness, without return to pre-existing conscious level | S06.5X7A | Traumatic subdural hemorrhage with loss of consciousness of any duration with death due to brain injury before regaining consciousness, initial encounter |
| 801.75 | Open fracture of base of skull with subarachnoid, subdural, and extradural hemorrhage, with prolonged [more than 24 hours] loss of consciousness, without return to pre-existing conscious level | S06.5X8A | Traumatic subdural hemorrhage with loss of consciousness of any duration with death due to other cause before regaining consciousness, initial encounter |
| 801.75 | Open fracture of base of skull with subarachnoid, subdural, and extradural hemorrhage, with prolonged [more than 24 hours] loss of consciousness, without return to pre-existing conscious level | S06.6X6A | Traumatic subarachnoid hemorrhage with loss of consciousness greater than 24 hours without return to pre-existing conscious level with patient surviving, initial encounter |
| 801.75 | Open fracture of base of skull with subarachnoid, subdural, and extradural hemorrhage, with prolonged [more than 24 hours] loss of consciousness, without return to pre-existing conscious level | S06.6X7A | Traumatic subarachnoid hemorrhage with loss of consciousness of any duration with death due to brain injury prior to regaining consciousness, initial encounter |
| 801.75 | Open fracture of base of skull with subarachnoid, subdural, and extradural hemorrhage, with prolonged [more than 24 hours] loss of consciousness, without return to pre-existing conscious level | S06.6X8A | Traumatic subarachnoid hemorrhage with loss of consciousness of any duration with death due to other cause prior to regaining consciousness, initial encounter |
| 801.75 | Open fracture of base of skull with subarachnoid, subdural, and extradural hemorrhage, with prolonged [more than 24 hours] loss of consciousness, without return to pre-existing conscious level |  |  |
| 801.76 | Open fracture of base of skull with subarachnoid, subdural, and extradural hemorrhage, with loss of consciousness of unspecified duration | S06.4X9A | Epidural hemorrhage with loss of consciousness of unspecified duration, initial encounter |
| 801.76 | Open fracture of base of skull with subarachnoid, subdural, and extradural hemorrhage, with loss of consciousness of unspecified duration | S06.5X9A | Traumatic subdural hemorrhage with loss of consciousness of unspecified duration, initial encounter |
| 801.76 | Open fracture of base of skull with subarachnoid, subdural, and extradural hemorrhage, with loss of consciousness of unspecified duration | S06.6X9A | Traumatic subarachnoid hemorrhage with loss of consciousness of unspecified duration, initial encounter |
| 801.76 | Open fracture of base of skull with subarachnoid, subdural, and extradural hemorrhage, with loss of consciousness of unspecified duration |  |  |
| 801.79 | Open fracture of base of skull with subarachnoid, subdural, and extradural hemorrhage, with concussion, unspecified | S06.4X0A | Epidural hemorrhage without loss of consciousness, initial encounter |
| 801.79 | Open fracture of base of skull with subarachnoid, subdural, and extradural hemorrhage, with concussion, unspecified | S06.5X0A | Traumatic subdural hemorrhage without loss of consciousness, initial encounter |
| 801.79 | Open fracture of base of skull with subarachnoid, subdural, and extradural hemorrhage, with concussion, unspecified | S06.6X0A | Traumatic subarachnoid hemorrhage without loss of consciousness, initial encounter |
| 801.79 | Open fracture of base of skull with subarachnoid, subdural, and extradural hemorrhage, with concussion, unspecified |  |  |
| 801.80 | Open fracture of base of skull with other and unspecified intracranial hemorrhage, unspecified state of consciousness | S02.109B | Fracture of base of skull, unspecified side, initial encounter for open fracture |
| 801.80 | Open fracture of base of skull with other and unspecified intracranial hemorrhage, unspecified state of consciousness | S06.360A | Traumatic hemorrhage of cerebrum, unspecified, without loss of consciousness, initial encounter |
| 801.81 | Open fracture of base of skull with other and unspecified intracranial hemorrhage, with no loss of consciousness | S02.109B | Fracture of base of skull, unspecified side, initial encounter for open fracture |
| 801.81 | Open fracture of base of skull with other and unspecified intracranial hemorrhage, with no loss of consciousness | S06.360A | Traumatic hemorrhage of cerebrum, unspecified, without loss of consciousness, initial encounter |
| 801.82 | Open fracture of base of skull with other and unspecified intracranial hemorrhage, with brief [less than one hour] loss of consciousness | S02.109B | Fracture of base of skull, unspecified side, initial encounter for open fracture |
| 801.82 | Open fracture of base of skull with other and unspecified intracranial hemorrhage, with brief [less than one hour] loss of consciousness | S06.361A | Traumatic hemorrhage of cerebrum, unspecified, with loss of consciousness of 30 minutes or less, initial encounter |
| 801.82 | Open fracture of base of skull with other and unspecified intracranial hemorrhage, with brief [less than one hour] loss of consciousness | S06.362A | Traumatic hemorrhage of cerebrum, unspecified, with loss of consciousness of 31 minutes to 59 minutes, initial encounter |
| 801.83 | Open fracture of base of skull with other and unspecified intracranial hemorrhage, with moderate [1-24 hours] loss of consciousness | S02.109B | Fracture of base of skull, unspecified side, initial encounter for open fracture |
| 801.83 | Open fracture of base of skull with other and unspecified intracranial hemorrhage, with moderate [1-24 hours] loss of consciousness | S06.363A | Traumatic hemorrhage of cerebrum, unspecified, with loss of consciousness of 1 hours to 5 hours 59 minutes, initial encounter |
| 801.83 | Open fracture of base of skull with other and unspecified intracranial hemorrhage, with moderate [1-24 hours] loss of consciousness | S06.364A | Traumatic hemorrhage of cerebrum, unspecified, with loss of consciousness of 6 hours to 24 hours, initial encounter |
| 801.84 | Open fracture of base of skull with other and unspecified intracranial hemorrhage, with prolonged [more than 24 hours] loss of consciousness and return to pre-existing conscious level | S02.109B | Fracture of base of skull, unspecified side, initial encounter for open fracture |
| 801.84 | Open fracture of base of skull with other and unspecified intracranial hemorrhage, with prolonged [more than 24 hours] loss of consciousness and return to pre-existing conscious level | S06.365A | Traumatic hemorrhage of cerebrum, unspecified, with loss of consciousness greater than 24 hours with return to pre-existing conscious level, initial encounter |
| 801.85 | Open fracture of base of skull with other and unspecified intracranial hemorrhage, with prolonged [more than 24 hours] loss of consciousness, without return to pre-existing conscious level | S02.109B | Fracture of base of skull, unspecified side, initial encounter for open fracture |
| 801.85 | Open fracture of base of skull with other and unspecified intracranial hemorrhage, with prolonged [more than 24 hours] loss of consciousness, without return to pre-existing conscious level | S06.366A | Traumatic hemorrhage of cerebrum, unspecified, with loss of consciousness greater than 24 hours without return to pre-existing conscious level with patient surviving, initial encounter |
| 801.85 | Open fracture of base of skull with other and unspecified intracranial hemorrhage, with prolonged [more than 24 hours] loss of consciousness, without return to pre-existing conscious level | S06.367A | Traumatic hemorrhage of cerebrum, unspecified, with loss of consciousness of any duration with death due to brain injury prior to regaining consciousness, initial encounter |
| 801.85 | Open fracture of base of skull with other and unspecified intracranial hemorrhage, with prolonged [more than 24 hours] loss of consciousness, without return to pre-existing conscious level | S06.368A | Traumatic hemorrhage of cerebrum, unspecified, with loss of consciousness of any duration with death due to other cause prior to regaining consciousness, initial encounter |
| 801.86 | Open fracture of base of skull with other and unspecified intracranial hemorrhage, with loss of consciousness of unspecified duration | S02.109B | Fracture of base of skull, unspecified side, initial encounter for open fracture |
| 801.86 | Open fracture of base of skull with other and unspecified intracranial hemorrhage, with loss of consciousness of unspecified duration | S06.369A | Traumatic hemorrhage of cerebrum, unspecified, with loss of consciousness of unspecified duration, initial encounter |
| 801.89 | Open fracture of base of skull with other and unspecified intracranial hemorrhage, with concussion, unspecified | S02.109B | Fracture of base of skull, unspecified side, initial encounter for open fracture |
| 801.89 | Open fracture of base of skull with other and unspecified intracranial hemorrhage, with concussion, unspecified | S06.360A | Traumatic hemorrhage of cerebrum, unspecified, without loss of consciousness, initial encounter |
| 801.90 | Open fracture of base of skull with intracranial injury of other and unspecified nature, unspecified state of consciousness | S02.109B | Fracture of base of skull, unspecified side, initial encounter for open fracture |
| 801.90 | Open fracture of base of skull with intracranial injury of other and unspecified nature, unspecified state of consciousness | S06.890A | Other specified intracranial injury without loss of consciousness, initial encounter |
| 801.90 | Open fracture of base of skull with intracranial injury of other and unspecified nature, unspecified state of consciousness | S06.9X0A | Unspecified intracranial injury without loss of consciousness, initial encounter |
| 801.91 | Open fracture of base of skull with intracranial injury of other and unspecified nature, with no loss of consciousness | S02.109B | Fracture of base of skull, unspecified side, initial encounter for open fracture |
| 801.91 | Open fracture of base of skull with intracranial injury of other and unspecified nature, with no loss of consciousness | S06.890A | Other specified intracranial injury without loss of consciousness, initial encounter |
| 801.91 | Open fracture of base of skull with intracranial injury of other and unspecified nature, with no loss of consciousness | S06.9X0A | Unspecified intracranial injury without loss of consciousness, initial encounter |
| 801.92 | Open fracture of base of skull with intracranial injury of other and unspecified nature, with brief [less than one hour] loss of consciousness | S02.109B | Fracture of base of skull, unspecified side, initial encounter for open fracture |
| 801.92 | Open fracture of base of skull with intracranial injury of other and unspecified nature, with brief [less than one hour] loss of consciousness | S06.891A | Other specified intracranial injury with loss of consciousness of 30 minutes or less, initial encounter |
| 801.92 | Open fracture of base of skull with intracranial injury of other and unspecified nature, with brief [less than one hour] loss of consciousness | S06.892A | Other specified intracranial injury with loss of consciousness of 31 minutes to 59 minutes, initial encounter |
| 801.92 | Open fracture of base of skull with intracranial injury of other and unspecified nature, with brief [less than one hour] loss of consciousness | S06.9X1A | Unspecified intracranial injury with loss of consciousness of 30 minutes or less, initial encounter |
| 801.92 | Open fracture of base of skull with intracranial injury of other and unspecified nature, with brief [less than one hour] loss of consciousness | S06.9X2A | Unspecified intracranial injury with loss of consciousness of 31 minutes to 59 minutes, initial encounter |
| 801.93 | Open fracture of base of skull with intracranial injury of other and unspecified nature, with moderate [1-24 hours] loss of consciousness | S02.109B | Fracture of base of skull, unspecified side, initial encounter for open fracture |
| 801.93 | Open fracture of base of skull with intracranial injury of other and unspecified nature, with moderate [1-24 hours] loss of consciousness | S06.893A | Other specified intracranial injury with loss of consciousness of 1 hour to 5 hours 59 minutes, initial encounter |
| 801.93 | Open fracture of base of skull with intracranial injury of other and unspecified nature, with moderate [1-24 hours] loss of consciousness | S06.894A | Other specified intracranial injury with loss of consciousness of 6 hours to 24 hours, initial encounter |
| 801.93 | Open fracture of base of skull with intracranial injury of other and unspecified nature, with moderate [1-24 hours] loss of consciousness | S06.9X3A | Unspecified intracranial injury with loss of consciousness of 1 hour to 5 hours 59 minutes, initial encounter |
| 801.93 | Open fracture of base of skull with intracranial injury of other and unspecified nature, with moderate [1-24 hours] loss of consciousness | S06.9X4A | Unspecified intracranial injury with loss of consciousness of 6 hours to 24 hours, initial encounter |
| 801.94 | Open fracture of base of skull with intracranial injury of other and unspecified nature, with prolonged [more than 24 hours] loss of consciousness and return to pre-existing conscious level | S02.109B | Fracture of base of skull, unspecified side, initial encounter for open fracture |
| 801.94 | Open fracture of base of skull with intracranial injury of other and unspecified nature, with prolonged [more than 24 hours] loss of consciousness and return to pre-existing conscious level | S06.895A | Other specified intracranial injury with loss of consciousness greater than 24 hours with return to pre-existing conscious level, initial encounter |
| 801.94 | Open fracture of base of skull with intracranial injury of other and unspecified nature, with prolonged [more than 24 hours] loss of consciousness and return to pre-existing conscious level | S06.9X5A | Unspecified intracranial injury with loss of consciousness greater than 24 hours with return to pre-existing conscious level, initial encounter |
| 801.95 | Open fracture of base of skull with intracranial injury of other and unspecified nature, with prolonged [more than 24 hours] loss of consciousness, without return to pre-existing conscious level | S02.109B | Fracture of base of skull, unspecified side, initial encounter for open fracture |
| 801.95 | Open fracture of base of skull with intracranial injury of other and unspecified nature, with prolonged [more than 24 hours] loss of consciousness, without return to pre-existing conscious level | S06.896A | Other specified intracranial injury with loss of consciousness greater than 24 hours without return to pre-existing conscious level with patient surviving, initial encounter |
| 801.95 | Open fracture of base of skull with intracranial injury of other and unspecified nature, with prolonged [more than 24 hours] loss of consciousness, without return to pre-existing conscious level | S06.897A | Other specified intracranial injury with loss of consciousness of any duration with death due to brain injury prior to regaining consciousness, initial encounter |
| 801.95 | Open fracture of base of skull with intracranial injury of other and unspecified nature, with prolonged [more than 24 hours] loss of consciousness, without return to pre-existing conscious level | S06.898A | Other specified intracranial injury with loss of consciousness of any duration with death due to other cause prior to regaining consciousness, initial encounter |
| 801.95 | Open fracture of base of skull with intracranial injury of other and unspecified nature, with prolonged [more than 24 hours] loss of consciousness, without return to pre-existing conscious level | S06.9X6A | Unspecified intracranial injury with loss of consciousness greater than 24 hours without return to pre-existing conscious level with patient surviving, initial encounter |
| 801.95 | Open fracture of base of skull with intracranial injury of other and unspecified nature, with prolonged [more than 24 hours] loss of consciousness, without return to pre-existing conscious level | S06.9X7A | Unspecified intracranial injury with loss of consciousness of any duration with death due to brain injury prior to regaining consciousness, initial encounter |
| 801.95 | Open fracture of base of skull with intracranial injury of other and unspecified nature, with prolonged [more than 24 hours] loss of consciousness, without return to pre-existing conscious level | S06.9X8A | Unspecified intracranial injury with loss of consciousness of any duration with death due to other cause prior to regaining consciousness, initial encounter |
| 801.96 | Open fracture of base of skull with intracranial injury of other and unspecified nature, with loss of consciousness of unspecified duration | S02.109B | Fracture of base of skull, unspecified side, initial encounter for open fracture |
| 801.96 | Open fracture of base of skull with intracranial injury of other and unspecified nature, with loss of consciousness of unspecified duration | S06.899A | Other specified intracranial injury with loss of consciousness of unspecified duration, initial encounter |
| 801.96 | Open fracture of base of skull with intracranial injury of other and unspecified nature, with loss of consciousness of unspecified duration | S06.9X9A | Unspecified intracranial injury with loss of consciousness of unspecified duration, initial encounter |
| 801.99 | Open fracture of base of skull with intracranial injury of other and unspecified nature, with concussion, unspecified | S02.109B | Fracture of base of skull, unspecified side, initial encounter for open fracture |
| 801.99 | Open fracture of base of skull with intracranial injury of other and unspecified nature, with concussion, unspecified | S06.890A | Other specified intracranial injury without loss of consciousness, initial encounter |
| 801.99 | Open fracture of base of skull with intracranial injury of other and unspecified nature, with concussion, unspecified | S06.9X0A | Unspecified intracranial injury without loss of consciousness, initial encounter |
| 803.10 | Other closed skull fracture with cerebral laceration and contusion, unspecified state of consciousness | S02.91XA | Unspecified fracture of skull, initial encounter for closed fracture |
| 803.10 | Other closed skull fracture with cerebral laceration and contusion, unspecified state of consciousness | S06.330A | Contusion and laceration of cerebrum, unspecified, without loss of consciousness, initial encounter |
| 803.11 | Other closed skull fracture with cerebral laceration and contusion, with no loss of consciousness | S02.91XA | Unspecified fracture of skull, initial encounter for closed fracture |
| 803.11 | Other closed skull fracture with cerebral laceration and contusion, with no loss of consciousness | S06.330A | Contusion and laceration of cerebrum, unspecified, without loss of consciousness, initial encounter |
| 803.12 | Other closed skull fracture with cerebral laceration and contusion, with brief [less than one hour] loss of consciousness | S02.91XA | Unspecified fracture of skull, initial encounter for closed fracture |
| 803.12 | Other closed skull fracture with cerebral laceration and contusion, with brief [less than one hour] loss of consciousness | S06.331A | Contusion and laceration of cerebrum, unspecified, with loss of consciousness of 30 minutes or less, initial encounter |
| 803.12 | Other closed skull fracture with cerebral laceration and contusion, with brief [less than one hour] loss of consciousness | S06.332A | Contusion and laceration of cerebrum, unspecified, with loss of consciousness of 31 minutes to 59 minutes, initial encounter |
| 803.13 | Other closed skull fracture with cerebral laceration and contusion, with moderate [1-24 hours] loss of consciousness | S02.91XA | Unspecified fracture of skull, initial encounter for closed fracture |
| 803.13 | Other closed skull fracture with cerebral laceration and contusion, with moderate [1-24 hours] loss of consciousness | S06.333A | Contusion and laceration of cerebrum, unspecified, with loss of consciousness of 1 hour to 5 hours 59 minutes, initial encounter |
| 803.13 | Other closed skull fracture with cerebral laceration and contusion, with moderate [1-24 hours] loss of consciousness | S06.334A | Contusion and laceration of cerebrum, unspecified, with loss of consciousness of 6 hours to 24 hours, initial encounter |
| 803.14 | Other closed skull fracture with cerebral laceration and contusion, with prolonged [more than 24 hours] loss of consciousness and return to pre-existing conscious level | S02.91XA | Unspecified fracture of skull, initial encounter for closed fracture |
| 803.14 | Other closed skull fracture with cerebral laceration and contusion, with prolonged [more than 24 hours] loss of consciousness and return to pre-existing conscious level | S06.335A | Contusion and laceration of cerebrum, unspecified, with loss of consciousness greater than 24 hours with return to pre-existing conscious level, initial encounter |
| 803.15 | Other closed skull fracture with cerebral laceration and contusion, with prolonged [more than 24 hours] loss of consciousness, without return to pre-existing conscious level | S02.91XA | Unspecified fracture of skull, initial encounter for closed fracture |
| 803.15 | Other closed skull fracture with cerebral laceration and contusion, with prolonged [more than 24 hours] loss of consciousness, without return to pre-existing conscious level | S06.336A | Contusion and laceration of cerebrum, unspecified, with loss of consciousness greater than 24 hours without return to pre-existing conscious level with patient surviving, initial encounter |
| 803.15 | Other closed skull fracture with cerebral laceration and contusion, with prolonged [more than 24 hours] loss of consciousness, without return to pre-existing conscious level | S06.337A | Contusion and laceration of cerebrum, unspecified, with loss of consciousness of any duration with death due to brain injury prior to regaining consciousness, initial encounter |
| 803.15 | Other closed skull fracture with cerebral laceration and contusion, with prolonged [more than 24 hours] loss of consciousness, without return to pre-existing conscious level | S06.338A | Contusion and laceration of cerebrum, unspecified, with loss of consciousness of any duration with death due to other cause prior to regaining consciousness, initial encounter |
| 803.16 | Other closed skull fracture with cerebral laceration and contusion, with loss of consciousness of unspecified duration | S02.91XA | Unspecified fracture of skull, initial encounter for closed fracture |
| 803.16 | Other closed skull fracture with cerebral laceration and contusion, with loss of consciousness of unspecified duration | S06.339A | Contusion and laceration of cerebrum, unspecified, with loss of consciousness of unspecified duration, initial encounter |
| 803.19 | Other closed skull fracture with cerebral laceration and contusion, with concussion, unspecified | S02.91XA | Unspecified fracture of skull, initial encounter for closed fracture |
| 803.19 | Other closed skull fracture with cerebral laceration and contusion, with concussion, unspecified | S06.330A | Contusion and laceration of cerebrum, unspecified, without loss of consciousness, initial encounter |
| 803.20 | Other closed skull fracture with subarachnoid, subdural, and extradural hemorrhage, unspecified state of consciousness | S06.4X0A | Epidural hemorrhage without loss of consciousness, initial encounter |
| 803.20 | Other closed skull fracture with subarachnoid, subdural, and extradural hemorrhage, unspecified state of consciousness | S06.5X0A | Traumatic subdural hemorrhage without loss of consciousness, initial encounter |
| 803.20 | Other closed skull fracture with subarachnoid, subdural, and extradural hemorrhage, unspecified state of consciousness | S06.6X0A | Traumatic subarachnoid hemorrhage without loss of consciousness, initial encounter |
| 803.20 | Other closed skull fracture with subarachnoid, subdural, and extradural hemorrhage, unspecified state of consciousness |  |  |
| 803.21 | Other closed skull fracture with subarachnoid, subdural, and extradural hemorrhage, with no loss of consciousness | S06.4X0A | Epidural hemorrhage without loss of consciousness, initial encounter |
| 803.21 | Other closed skull fracture with subarachnoid, subdural, and extradural hemorrhage, with no loss of consciousness | S06.5X0A | Traumatic subdural hemorrhage without loss of consciousness, initial encounter |
| 803.21 | Other closed skull fracture with subarachnoid, subdural, and extradural hemorrhage, with no loss of consciousness | S06.6X0A | Traumatic subarachnoid hemorrhage without loss of consciousness, initial encounter |
| 803.21 | Other closed skull fracture with subarachnoid, subdural, and extradural hemorrhage, with no loss of consciousness |  |  |
| 803.22 | Other closed skull fracture with subarachnoid, subdural, and extradural hemorrhage, with brief [less than one hour] loss of consciousness | S06.4X1A | Epidural hemorrhage with loss of consciousness of 30 minutes or less, initial encounter |
| 803.22 | Other closed skull fracture with subarachnoid, subdural, and extradural hemorrhage, with brief [less than one hour] loss of consciousness | S06.4X2A | Epidural hemorrhage with loss of consciousness of 31 minutes to 59 minutes, initial encounter |
| 803.22 | Other closed skull fracture with subarachnoid, subdural, and extradural hemorrhage, with brief [less than one hour] loss of consciousness | S06.5X1A | Traumatic subdural hemorrhage with loss of consciousness of 30 minutes or less, initial encounter |
| 803.22 | Other closed skull fracture with subarachnoid, subdural, and extradural hemorrhage, with brief [less than one hour] loss of consciousness | S06.5X2A | Traumatic subdural hemorrhage with loss of consciousness of 31 minutes to 59 minutes, initial encounter |
| 803.22 | Other closed skull fracture with subarachnoid, subdural, and extradural hemorrhage, with brief [less than one hour] loss of consciousness | S06.6X1A | Traumatic subarachnoid hemorrhage with loss of consciousness of 30 minutes or less, initial encounter |
| 803.22 | Other closed skull fracture with subarachnoid, subdural, and extradural hemorrhage, with brief [less than one hour] loss of consciousness | S06.6X2A | Traumatic subarachnoid hemorrhage with loss of consciousness of 31 minutes to 59 minutes, initial encounter |
| 803.22 | Other closed skull fracture with subarachnoid, subdural, and extradural hemorrhage, with brief [less than one hour] loss of consciousness |  |  |
| 803.23 | Other closed skull fracture with subarachnoid, subdural, and extradural hemorrhage, with moderate [1-24 hours] loss of consciousness | S06.4X3A | Epidural hemorrhage with loss of consciousness of 1 hour to 5 hours 59 minutes, initial encounter |
| 803.23 | Other closed skull fracture with subarachnoid, subdural, and extradural hemorrhage, with moderate [1-24 hours] loss of consciousness | S06.4X4A | Epidural hemorrhage with loss of consciousness of 6 hours to 24 hours, initial encounter |
| 803.23 | Other closed skull fracture with subarachnoid, subdural, and extradural hemorrhage, with moderate [1-24 hours] loss of consciousness | S06.5X3A | Traumatic subdural hemorrhage with loss of consciousness of 1 hour to 5 hours 59 minutes, initial encounter |
| 803.23 | Other closed skull fracture with subarachnoid, subdural, and extradural hemorrhage, with moderate [1-24 hours] loss of consciousness | S06.5X4A | Traumatic subdural hemorrhage with loss of consciousness of 6 hours to 24 hours, initial encounter |
| 803.23 | Other closed skull fracture with subarachnoid, subdural, and extradural hemorrhage, with moderate [1-24 hours] loss of consciousness | S06.6X3A | Traumatic subarachnoid hemorrhage with loss of consciousness of 1 hour to 5 hours 59 minutes, initial encounter |
| 803.23 | Other closed skull fracture with subarachnoid, subdural, and extradural hemorrhage, with moderate [1-24 hours] loss of consciousness | S06.6X4A | Traumatic subarachnoid hemorrhage with loss of consciousness of 6 hours to 24 hours, initial encounter |
| 803.23 | Other closed skull fracture with subarachnoid, subdural, and extradural hemorrhage, with moderate [1-24 hours] loss of consciousness |  |  |
| 803.24 | Other closed skull fracture with subarachnoid, subdural, and extradural hemorrhage, with prolonged [more than 24 hours] loss of consciousness and return to pre-existing conscious level | S06.4X5A | Epidural hemorrhage with loss of consciousness greater than 24 hours with return to pre-existing conscious level, initial encounter |
| 803.24 | Other closed skull fracture with subarachnoid, subdural, and extradural hemorrhage, with prolonged [more than 24 hours] loss of consciousness and return to pre-existing conscious level | S06.5X5A | Traumatic subdural hemorrhage with loss of consciousness greater than 24 hours with return to pre-existing conscious level, initial encounter |
| 803.24 | Other closed skull fracture with subarachnoid, subdural, and extradural hemorrhage, with prolonged [more than 24 hours] loss of consciousness and return to pre-existing conscious level | S06.6X5A | Traumatic subarachnoid hemorrhage with loss of consciousness greater than 24 hours with return to pre-existing conscious level, initial encounter |
| 803.24 | Other closed skull fracture with subarachnoid, subdural, and extradural hemorrhage, with prolonged [more than 24 hours] loss of consciousness and return to pre-existing conscious level |  |  |
| 803.25 | Other closed skull fracture with subarachnoid, subdural, and extradural hemorrhage, with prolonged [more than 24 hours] loss of consciousness, without return to pre-existing conscious level | S06.4X6A | Epidural hemorrhage with loss of consciousness greater than 24 hours without return to pre-existing conscious level with patient surviving, initial encounter |
| 803.25 | Other closed skull fracture with subarachnoid, subdural, and extradural hemorrhage, with prolonged [more than 24 hours] loss of consciousness, without return to pre-existing conscious level | S06.4X7A | Epidural hemorrhage with loss of consciousness of any duration with death due to brain injury prior to regaining consciousness, initial encounter |
| 803.25 | Other closed skull fracture with subarachnoid, subdural, and extradural hemorrhage, with prolonged [more than 24 hours] loss of consciousness, without return to pre-existing conscious level | S06.4X8A | Epidural hemorrhage with loss of consciousness of any duration with death due to other causes prior to regaining consciousness, initial encounter |
| 803.25 | Other closed skull fracture with subarachnoid, subdural, and extradural hemorrhage, with prolonged [more than 24 hours] loss of consciousness, without return to pre-existing conscious level | S06.5X6A | Traumatic subdural hemorrhage with loss of consciousness greater than 24 hours without return to pre-existing conscious level with patient surviving, initial encounter |
| 803.25 | Other closed skull fracture with subarachnoid, subdural, and extradural hemorrhage, with prolonged [more than 24 hours] loss of consciousness, without return to pre-existing conscious level | S06.5X7A | Traumatic subdural hemorrhage with loss of consciousness of any duration with death due to brain injury before regaining consciousness, initial encounter |
| 803.25 | Other closed skull fracture with subarachnoid, subdural, and extradural hemorrhage, with prolonged [more than 24 hours] loss of consciousness, without return to pre-existing conscious level | S06.5X8A | Traumatic subdural hemorrhage with loss of consciousness of any duration with death due to other cause before regaining consciousness, initial encounter |
| 803.25 | Other closed skull fracture with subarachnoid, subdural, and extradural hemorrhage, with prolonged [more than 24 hours] loss of consciousness, without return to pre-existing conscious level | S06.6X6A | Traumatic subarachnoid hemorrhage with loss of consciousness greater than 24 hours without return to pre-existing conscious level with patient surviving, initial encounter |
| 803.25 | Other closed skull fracture with subarachnoid, subdural, and extradural hemorrhage, with prolonged [more than 24 hours] loss of consciousness, without return to pre-existing conscious level | S06.6X7A | Traumatic subarachnoid hemorrhage with loss of consciousness of any duration with death due to brain injury prior to regaining consciousness, initial encounter |
| 803.25 | Other closed skull fracture with subarachnoid, subdural, and extradural hemorrhage, with prolonged [more than 24 hours] loss of consciousness, without return to pre-existing conscious level | S06.6X8A | Traumatic subarachnoid hemorrhage with loss of consciousness of any duration with death due to other cause prior to regaining consciousness, initial encounter |
| 803.25 | Other closed skull fracture with subarachnoid, subdural, and extradural hemorrhage, with prolonged [more than 24 hours] loss of consciousness, without return to pre-existing conscious level |  |  |
| 803.26 | Other closed skull fracture with subarachnoid, subdural, and extradural hemorrhage, with loss of consciousness of unspecified duration | S06.4X9A | Epidural hemorrhage with loss of consciousness of unspecified duration, initial encounter |
| 803.26 | Other closed skull fracture with subarachnoid, subdural, and extradural hemorrhage, with loss of consciousness of unspecified duration | S06.5X9A | Traumatic subdural hemorrhage with loss of consciousness of unspecified duration, initial encounter |
| 803.26 | Other closed skull fracture with subarachnoid, subdural, and extradural hemorrhage, with loss of consciousness of unspecified duration | S06.6X9A | Traumatic subarachnoid hemorrhage with loss of consciousness of unspecified duration, initial encounter |
| 803.26 | Other closed skull fracture with subarachnoid, subdural, and extradural hemorrhage, with loss of consciousness of unspecified duration |  |  |
| 803.29 | Other closed skull fracture with subarachnoid, subdural, and extradural hemorrhage, with concussion, unspecified | S06.4X0A | Epidural hemorrhage without loss of consciousness, initial encounter |
| 803.29 | Other closed skull fracture with subarachnoid, subdural, and extradural hemorrhage, with concussion, unspecified | S06.5X0A | Traumatic subdural hemorrhage without loss of consciousness, initial encounter |
| 803.29 | Other closed skull fracture with subarachnoid, subdural, and extradural hemorrhage, with concussion, unspecified | S06.6X0A | Traumatic subarachnoid hemorrhage without loss of consciousness, initial encounter |
| 803.29 | Other closed skull fracture with subarachnoid, subdural, and extradural hemorrhage, with concussion, unspecified |  |  |
| 803.30 | Other closed skull fracture with other and unspecified intracranial hemorrhage, unspecified state of unconsciousness | S02.91XA | Unspecified fracture of skull, initial encounter for closed fracture |
| 803.30 | Other closed skull fracture with other and unspecified intracranial hemorrhage, unspecified state of unconsciousness | S06.369A | Traumatic hemorrhage of cerebrum, unspecified, with loss of consciousness of unspecified duration, initial encounter |
| 803.31 | Other closed skull fracture with other and unspecified intracranial hemorrhage, with no loss of consciousness | S02.91XA | Unspecified fracture of skull, initial encounter for closed fracture |
| 803.31 | Other closed skull fracture with other and unspecified intracranial hemorrhage, with no loss of consciousness | S06.360A | Traumatic hemorrhage of cerebrum, unspecified, without loss of consciousness, initial encounter |
| 803.32 | Other closed skull fracture with other and unspecified intracranial hemorrhage, with brief [less than one hour] loss of consciousness | S02.91XA | Unspecified fracture of skull, initial encounter for closed fracture |
| 803.32 | Other closed skull fracture with other and unspecified intracranial hemorrhage, with brief [less than one hour] loss of consciousness | S06.361A | Traumatic hemorrhage of cerebrum, unspecified, with loss of consciousness of 30 minutes or less, initial encounter |
| 803.32 | Other closed skull fracture with other and unspecified intracranial hemorrhage, with brief [less than one hour] loss of consciousness | S06.362A | Traumatic hemorrhage of cerebrum, unspecified, with loss of consciousness of 31 minutes to 59 minutes, initial encounter |
| 803.33 | Other closed skull fracture with other and unspecified intracranial hemorrhage, with moderate [1-24 hours] loss of consciousness | S02.91XA | Unspecified fracture of skull, initial encounter for closed fracture |
| 803.33 | Other closed skull fracture with other and unspecified intracranial hemorrhage, with moderate [1-24 hours] loss of consciousness | S06.363A | Traumatic hemorrhage of cerebrum, unspecified, with loss of consciousness of 1 hours to 5 hours 59 minutes, initial encounter |
| 803.33 | Other closed skull fracture with other and unspecified intracranial hemorrhage, with moderate [1-24 hours] loss of consciousness | S06.364A | Traumatic hemorrhage of cerebrum, unspecified, with loss of consciousness of 6 hours to 24 hours, initial encounter |
| 803.34 | Other closed skull fracture with other and unspecified intracranial hemorrhage, with prolonged [more than 24 hours] loss of consciousness and return to pre-existing conscious level | S02.91XA | Unspecified fracture of skull, initial encounter for closed fracture |
| 803.34 | Other closed skull fracture with other and unspecified intracranial hemorrhage, with prolonged [more than 24 hours] loss of consciousness and return to pre-existing conscious level | S06.365A | Traumatic hemorrhage of cerebrum, unspecified, with loss of consciousness greater than 24 hours with return to pre-existing conscious level, initial encounter |
| 803.35 | Other closed skull fracture with other and unspecified intracranial hemorrhage, with prolonged [more than 24 hours] loss of consciousness, without return to pre-existing conscious level | S02.91XA | Unspecified fracture of skull, initial encounter for closed fracture |
| 803.35 | Other closed skull fracture with other and unspecified intracranial hemorrhage, with prolonged [more than 24 hours] loss of consciousness, without return to pre-existing conscious level | S06.366A | Traumatic hemorrhage of cerebrum, unspecified, with loss of consciousness greater than 24 hours without return to pre-existing conscious level with patient surviving, initial encounter |
| 803.35 | Other closed skull fracture with other and unspecified intracranial hemorrhage, with prolonged [more than 24 hours] loss of consciousness, without return to pre-existing conscious level | S06.367A | Traumatic hemorrhage of cerebrum, unspecified, with loss of consciousness of any duration with death due to brain injury prior to regaining consciousness, initial encounter |
| 803.35 | Other closed skull fracture with other and unspecified intracranial hemorrhage, with prolonged [more than 24 hours] loss of consciousness, without return to pre-existing conscious level | S06.368A | Traumatic hemorrhage of cerebrum, unspecified, with loss of consciousness of any duration with death due to other cause prior to regaining consciousness, initial encounter |
| 803.36 | Other closed skull fracture with other and unspecified intracranial hemorrhage, with loss of consciousness of unspecified duration | S02.91XA | Unspecified fracture of skull, initial encounter for closed fracture |
| 803.36 | Other closed skull fracture with other and unspecified intracranial hemorrhage, with loss of consciousness of unspecified duration | S06.369A | Traumatic hemorrhage of cerebrum, unspecified, with loss of consciousness of unspecified duration, initial encounter |
| 803.39 | Other closed skull fracture with other and unspecified intracranial hemorrhage, with concussion, unspecified | S02.91XA | Unspecified fracture of skull, initial encounter for closed fracture |
| 803.39 | Other closed skull fracture with other and unspecified intracranial hemorrhage, with concussion, unspecified | S06.360A | Traumatic hemorrhage of cerebrum, unspecified, without loss of consciousness, initial encounter |
| 803.40 | Other closed skull fracture with intracranial injury of other and unspecified nature, unspecified state of consciousness | S02.91XA | Unspecified fracture of skull, initial encounter for closed fracture |
| 803.40 | Other closed skull fracture with intracranial injury of other and unspecified nature, unspecified state of consciousness | S06.890A | Other specified intracranial injury without loss of consciousness, initial encounter |
| 803.40 | Other closed skull fracture with intracranial injury of other and unspecified nature, unspecified state of consciousness | S06.9X0A | Unspecified intracranial injury without loss of consciousness, initial encounter |
| 803.41 | Other closed skull fracture with intracranial injury of other and unspecified nature, with no loss of consciousness | S02.91XA | Unspecified fracture of skull, initial encounter for closed fracture |
| 803.41 | Other closed skull fracture with intracranial injury of other and unspecified nature, with no loss of consciousness | S06.890A | Other specified intracranial injury without loss of consciousness, initial encounter |
| 803.41 | Other closed skull fracture with intracranial injury of other and unspecified nature, with no loss of consciousness | S06.9X0A | Unspecified intracranial injury without loss of consciousness, initial encounter |
| 803.42 | Other closed skull fracture with intracranial injury of other and unspecified nature, with brief [less than one hour] loss of consciousness | S02.91XA | Unspecified fracture of skull, initial encounter for closed fracture |
| 803.42 | Other closed skull fracture with intracranial injury of other and unspecified nature, with brief [less than one hour] loss of consciousness | S06.891A | Other specified intracranial injury with loss of consciousness of 30 minutes or less, initial encounter |
| 803.42 | Other closed skull fracture with intracranial injury of other and unspecified nature, with brief [less than one hour] loss of consciousness | S06.892A | Other specified intracranial injury with loss of consciousness of 31 minutes to 59 minutes, initial encounter |
| 803.42 | Other closed skull fracture with intracranial injury of other and unspecified nature, with brief [less than one hour] loss of consciousness | S06.9X1A | Unspecified intracranial injury with loss of consciousness of 30 minutes or less, initial encounter |
| 803.42 | Other closed skull fracture with intracranial injury of other and unspecified nature, with brief [less than one hour] loss of consciousness | S06.9X2A | Unspecified intracranial injury with loss of consciousness of 31 minutes to 59 minutes, initial encounter |
| 803.43 | Other closed skull fracture with intracranial injury of other and unspecified nature, with moderate [1-24 hours] loss of consciousness | S02.91XA | Unspecified fracture of skull, initial encounter for closed fracture |
| 803.43 | Other closed skull fracture with intracranial injury of other and unspecified nature, with moderate [1-24 hours] loss of consciousness | S06.893A | Other specified intracranial injury with loss of consciousness of 1 hour to 5 hours 59 minutes, initial encounter |
| 803.43 | Other closed skull fracture with intracranial injury of other and unspecified nature, with moderate [1-24 hours] loss of consciousness | S06.894A | Other specified intracranial injury with loss of consciousness of 6 hours to 24 hours, initial encounter |
| 803.43 | Other closed skull fracture with intracranial injury of other and unspecified nature, with moderate [1-24 hours] loss of consciousness | S06.9X3A | Unspecified intracranial injury with loss of consciousness of 1 hour to 5 hours 59 minutes, initial encounter |
| 803.43 | Other closed skull fracture with intracranial injury of other and unspecified nature, with moderate [1-24 hours] loss of consciousness | S06.9X4A | Unspecified intracranial injury with loss of consciousness of 6 hours to 24 hours, initial encounter |
| 803.44 | Other closed skull fracture with intracranial injury of other and unspecified nature, with prolonged [more than 24 hours] loss of consciousness and return to pre-existing conscious level | S02.91XA | Unspecified fracture of skull, initial encounter for closed fracture |
| 803.44 | Other closed skull fracture with intracranial injury of other and unspecified nature, with prolonged [more than 24 hours] loss of consciousness and return to pre-existing conscious level | S06.895A | Other specified intracranial injury with loss of consciousness greater than 24 hours with return to pre-existing conscious level, initial encounter |
| 803.44 | Other closed skull fracture with intracranial injury of other and unspecified nature, with prolonged [more than 24 hours] loss of consciousness and return to pre-existing conscious level | S06.9X5A | Unspecified intracranial injury with loss of consciousness greater than 24 hours with return to pre-existing conscious level, initial encounter |
| 803.45 | Other closed skull fracture with intracranial injury of other and unspecified nature, with prolonged [more than 24 hours] loss of consciousness, without return to pre-existing conscious level | S02.91XA | Unspecified fracture of skull, initial encounter for closed fracture |
| 803.45 | Other closed skull fracture with intracranial injury of other and unspecified nature, with prolonged [more than 24 hours] loss of consciousness, without return to pre-existing conscious level | S06.896A | Other specified intracranial injury with loss of consciousness greater than 24 hours without return to pre-existing conscious level with patient surviving, initial encounter |
| 803.45 | Other closed skull fracture with intracranial injury of other and unspecified nature, with prolonged [more than 24 hours] loss of consciousness, without return to pre-existing conscious level | S06.897A | Other specified intracranial injury with loss of consciousness of any duration with death due to brain injury prior to regaining consciousness, initial encounter |
| 803.45 | Other closed skull fracture with intracranial injury of other and unspecified nature, with prolonged [more than 24 hours] loss of consciousness, without return to pre-existing conscious level | S06.898A | Other specified intracranial injury with loss of consciousness of any duration with death due to other cause prior to regaining consciousness, initial encounter |
| 803.45 | Other closed skull fracture with intracranial injury of other and unspecified nature, with prolonged [more than 24 hours] loss of consciousness, without return to pre-existing conscious level | S06.9X6A | Unspecified intracranial injury with loss of consciousness greater than 24 hours without return to pre-existing conscious level with patient surviving, initial encounter |
| 803.45 | Other closed skull fracture with intracranial injury of other and unspecified nature, with prolonged [more than 24 hours] loss of consciousness, without return to pre-existing conscious level | S06.9X7A | Unspecified intracranial injury with loss of consciousness of any duration with death due to brain injury prior to regaining consciousness, initial encounter |
| 803.45 | Other closed skull fracture with intracranial injury of other and unspecified nature, with prolonged [more than 24 hours] loss of consciousness, without return to pre-existing conscious level | S06.9X8A | Unspecified intracranial injury with loss of consciousness of any duration with death due to other cause prior to regaining consciousness, initial encounter |
| 803.46 | Other closed skull fracture with intracranial injury of other and unspecified nature, with loss of consciousness of unspecified duration | S02.91XA | Unspecified fracture of skull, initial encounter for closed fracture |
| 803.46 | Other closed skull fracture with intracranial injury of other and unspecified nature, with loss of consciousness of unspecified duration | S06.899A | Other specified intracranial injury with loss of consciousness of unspecified duration, initial encounter |
| 803.46 | Other closed skull fracture with intracranial injury of other and unspecified nature, with loss of consciousness of unspecified duration | S06.9X9A | Unspecified intracranial injury with loss of consciousness of unspecified duration, initial encounter |
| 803.49 | Other closed skull fracture with intracranial injury of other and unspecified nature, with concussion, unspecified | S02.91XA | Unspecified fracture of skull, initial encounter for closed fracture |
| 803.49 | Other closed skull fracture with intracranial injury of other and unspecified nature, with concussion, unspecified | S06.890A | Other specified intracranial injury without loss of consciousness, initial encounter |
| 803.49 | Other closed skull fracture with intracranial injury of other and unspecified nature, with concussion, unspecified | S06.9X0A | Unspecified intracranial injury without loss of consciousness, initial encounter |
| 803.60 | Other open skull fracture with cerebral laceration and contusion, unspecified state of consciousness | S02.91XB | Unspecified fracture of skull, initial encounter for open fracture |
| 803.60 | Other open skull fracture with cerebral laceration and contusion, unspecified state of consciousness | S06.330A | Contusion and laceration of cerebrum, unspecified, without loss of consciousness, initial encounter |
| 803.61 | Other open skull fracture with cerebral laceration and contusion, with no loss of consciousness | S02.91XB | Unspecified fracture of skull, initial encounter for open fracture |
| 803.61 | Other open skull fracture with cerebral laceration and contusion, with no loss of consciousness | S06.330A | Contusion and laceration of cerebrum, unspecified, without loss of consciousness, initial encounter |
| 803.62 | Other open skull fracture with cerebral laceration and contusion, with brief [less than one hour] loss of consciousness | S02.91XB | Unspecified fracture of skull, initial encounter for open fracture |
| 803.62 | Other open skull fracture with cerebral laceration and contusion, with brief [less than one hour] loss of consciousness | S06.331A | Contusion and laceration of cerebrum, unspecified, with loss of consciousness of 30 minutes or less, initial encounter |
| 803.62 | Other open skull fracture with cerebral laceration and contusion, with brief [less than one hour] loss of consciousness | S06.332A | Contusion and laceration of cerebrum, unspecified, with loss of consciousness of 31 minutes to 59 minutes, initial encounter |
| 803.63 | Other open skull fracture with cerebral laceration and contusion, with moderate [1-24 hours] loss of consciousness | S02.91XB | Unspecified fracture of skull, initial encounter for open fracture |
| 803.63 | Other open skull fracture with cerebral laceration and contusion, with moderate [1-24 hours] loss of consciousness | S06.333A | Contusion and laceration of cerebrum, unspecified, with loss of consciousness of 1 hour to 5 hours 59 minutes, initial encounter |
| 803.63 | Other open skull fracture with cerebral laceration and contusion, with moderate [1-24 hours] loss of consciousness | S06.334A | Contusion and laceration of cerebrum, unspecified, with loss of consciousness of 6 hours to 24 hours, initial encounter |
| 803.64 | Other open skull fracture with cerebral laceration and contusion, with prolonged [more than 24 hours] loss of consciousness and return to pre-existing conscious level | S02.91XB | Unspecified fracture of skull, initial encounter for open fracture |
| 803.64 | Other open skull fracture with cerebral laceration and contusion, with prolonged [more than 24 hours] loss of consciousness and return to pre-existing conscious level | S06.335A | Contusion and laceration of cerebrum, unspecified, with loss of consciousness greater than 24 hours with return to pre-existing conscious level, initial encounter |
| 803.65 | Other open skull fracture with cerebral laceration and contusion, with prolonged [more than 24 hours] loss of consciousness, without return to pre-existing conscious level | S02.91XB | Unspecified fracture of skull, initial encounter for open fracture |
| 803.65 | Other open skull fracture with cerebral laceration and contusion, with prolonged [more than 24 hours] loss of consciousness, without return to pre-existing conscious level | S06.336A | Contusion and laceration of cerebrum, unspecified, with loss of consciousness greater than 24 hours without return to pre-existing conscious level with patient surviving, initial encounter |
| 803.65 | Other open skull fracture with cerebral laceration and contusion, with prolonged [more than 24 hours] loss of consciousness, without return to pre-existing conscious level | S06.337A | Contusion and laceration of cerebrum, unspecified, with loss of consciousness of any duration with death due to brain injury prior to regaining consciousness, initial encounter |
| 803.65 | Other open skull fracture with cerebral laceration and contusion, with prolonged [more than 24 hours] loss of consciousness, without return to pre-existing conscious level | S06.338A | Contusion and laceration of cerebrum, unspecified, with loss of consciousness of any duration with death due to other cause prior to regaining consciousness, initial encounter |
| 803.66 | Other open skull fracture with cerebral laceration and contusion, with loss of consciousness of unspecified duration | S02.91XB | Unspecified fracture of skull, initial encounter for open fracture |
| 803.66 | Other open skull fracture with cerebral laceration and contusion, with loss of consciousness of unspecified duration | S06.339A | Contusion and laceration of cerebrum, unspecified, with loss of consciousness of unspecified duration, initial encounter |
| 803.69 | Other open skull fracture with cerebral laceration and contusion, with concussion, unspecified | S02.91XB | Unspecified fracture of skull, initial encounter for open fracture |
| 803.69 | Other open skull fracture with cerebral laceration and contusion, with concussion, unspecified | S06.330A | Contusion and laceration of cerebrum, unspecified, without loss of consciousness, initial encounter |
| 803.70 | Other open skull fracture with subarachnoid, subdural, and extradural hemorrhage, unspecified state of consciousness | S06.4X0A | Epidural hemorrhage without loss of consciousness, initial encounter |
| 803.70 | Other open skull fracture with subarachnoid, subdural, and extradural hemorrhage, unspecified state of consciousness | S06.5X0A | Traumatic subdural hemorrhage without loss of consciousness, initial encounter |
| 803.70 | Other open skull fracture with subarachnoid, subdural, and extradural hemorrhage, unspecified state of consciousness | S06.6X0A | Traumatic subarachnoid hemorrhage without loss of consciousness, initial encounter |
| 803.70 | Other open skull fracture with subarachnoid, subdural, and extradural hemorrhage, unspecified state of consciousness |  |  |
| 803.71 | Other open skull fracture with subarachnoid, subdural, and extradural hemorrhage, with no loss of consciousness | S06.4X0A | Epidural hemorrhage without loss of consciousness, initial encounter |
| 803.71 | Other open skull fracture with subarachnoid, subdural, and extradural hemorrhage, with no loss of consciousness | S06.5X0A | Traumatic subdural hemorrhage without loss of consciousness, initial encounter |
| 803.71 | Other open skull fracture with subarachnoid, subdural, and extradural hemorrhage, with no loss of consciousness | S06.6X0A | Traumatic subarachnoid hemorrhage without loss of consciousness, initial encounter |
| 803.71 | Other open skull fracture with subarachnoid, subdural, and extradural hemorrhage, with no loss of consciousness |  |  |
| 803.72 | Other open skull fracture with subarachnoid, subdural, and extradural hemorrhage, with brief [less than one hour] loss of consciousness | S06.4X1A | Epidural hemorrhage with loss of consciousness of 30 minutes or less, initial encounter |
| 803.72 | Other open skull fracture with subarachnoid, subdural, and extradural hemorrhage, with brief [less than one hour] loss of consciousness | S06.4X2A | Epidural hemorrhage with loss of consciousness of 31 minutes to 59 minutes, initial encounter |
| 803.72 | Other open skull fracture with subarachnoid, subdural, and extradural hemorrhage, with brief [less than one hour] loss of consciousness | S06.5X1A | Traumatic subdural hemorrhage with loss of consciousness of 30 minutes or less, initial encounter |
| 803.72 | Other open skull fracture with subarachnoid, subdural, and extradural hemorrhage, with brief [less than one hour] loss of consciousness | S06.5X2A | Traumatic subdural hemorrhage with loss of consciousness of 31 minutes to 59 minutes, initial encounter |
| 803.72 | Other open skull fracture with subarachnoid, subdural, and extradural hemorrhage, with brief [less than one hour] loss of consciousness | S06.6X1A | Traumatic subarachnoid hemorrhage with loss of consciousness of 30 minutes or less, initial encounter |
| 803.72 | Other open skull fracture with subarachnoid, subdural, and extradural hemorrhage, with brief [less than one hour] loss of consciousness | S06.6X2A | Traumatic subarachnoid hemorrhage with loss of consciousness of 31 minutes to 59 minutes, initial encounter |
| 803.72 | Other open skull fracture with subarachnoid, subdural, and extradural hemorrhage, with brief [less than one hour] loss of consciousness |  |  |
| 803.73 | Other open skull fracture with subarachnoid, subdural, and extradural hemorrhage, with moderate [1-24 hours] loss of consciousness | S06.4X3A | Epidural hemorrhage with loss of consciousness of 1 hour to 5 hours 59 minutes, initial encounter |
| 803.73 | Other open skull fracture with subarachnoid, subdural, and extradural hemorrhage, with moderate [1-24 hours] loss of consciousness | S06.4X4A | Epidural hemorrhage with loss of consciousness of 6 hours to 24 hours, initial encounter |
| 803.73 | Other open skull fracture with subarachnoid, subdural, and extradural hemorrhage, with moderate [1-24 hours] loss of consciousness | S06.5X3A | Traumatic subdural hemorrhage with loss of consciousness of 1 hour to 5 hours 59 minutes, initial encounter |
| 803.73 | Other open skull fracture with subarachnoid, subdural, and extradural hemorrhage, with moderate [1-24 hours] loss of consciousness | S06.5X4A | Traumatic subdural hemorrhage with loss of consciousness of 6 hours to 24 hours, initial encounter |
| 803.73 | Other open skull fracture with subarachnoid, subdural, and extradural hemorrhage, with moderate [1-24 hours] loss of consciousness | S06.6X3A | Traumatic subarachnoid hemorrhage with loss of consciousness of 1 hour to 5 hours 59 minutes, initial encounter |
| 803.73 | Other open skull fracture with subarachnoid, subdural, and extradural hemorrhage, with moderate [1-24 hours] loss of consciousness | S06.6X4A | Traumatic subarachnoid hemorrhage with loss of consciousness of 6 hours to 24 hours, initial encounter |
| 803.73 | Other open skull fracture with subarachnoid, subdural, and extradural hemorrhage, with moderate [1-24 hours] loss of consciousness |  |  |
| 803.74 | Other open skull fracture with subarachnoid, subdural, and extradural hemorrhage, with prolonged [more than 24 hours] loss of consciousness and return to pre-existing conscious level | S06.4X5A | Epidural hemorrhage with loss of consciousness greater than 24 hours with return to pre-existing conscious level, initial encounter |
| 803.74 | Other open skull fracture with subarachnoid, subdural, and extradural hemorrhage, with prolonged [more than 24 hours] loss of consciousness and return to pre-existing conscious level | S06.5X5A | Traumatic subdural hemorrhage with loss of consciousness greater than 24 hours with return to pre-existing conscious level, initial encounter |
| 803.74 | Other open skull fracture with subarachnoid, subdural, and extradural hemorrhage, with prolonged [more than 24 hours] loss of consciousness and return to pre-existing conscious level | S06.6X5A | Traumatic subarachnoid hemorrhage with loss of consciousness greater than 24 hours with return to pre-existing conscious level, initial encounter |
| 803.74 | Other open skull fracture with subarachnoid, subdural, and extradural hemorrhage, with prolonged [more than 24 hours] loss of consciousness and return to pre-existing conscious level |  |  |
| 803.75 | Other open skull fracture with subarachnoid, subdural, and extradural hemorrhage, with prolonged [more than 24 hours] loss of consciousness, without return to pre-existing conscious level | S06.4X6A | Epidural hemorrhage with loss of consciousness greater than 24 hours without return to pre-existing conscious level with patient surviving, initial encounter |
| 803.75 | Other open skull fracture with subarachnoid, subdural, and extradural hemorrhage, with prolonged [more than 24 hours] loss of consciousness, without return to pre-existing conscious level | S06.4X7A | Epidural hemorrhage with loss of consciousness of any duration with death due to brain injury prior to regaining consciousness, initial encounter |
| 803.75 | Other open skull fracture with subarachnoid, subdural, and extradural hemorrhage, with prolonged [more than 24 hours] loss of consciousness, without return to pre-existing conscious level | S06.4X8A | Epidural hemorrhage with loss of consciousness of any duration with death due to other causes prior to regaining consciousness, initial encounter |
| 803.75 | Other open skull fracture with subarachnoid, subdural, and extradural hemorrhage, with prolonged [more than 24 hours] loss of consciousness, without return to pre-existing conscious level | S06.5X6A | Traumatic subdural hemorrhage with loss of consciousness greater than 24 hours without return to pre-existing conscious level with patient surviving, initial encounter |
| 803.75 | Other open skull fracture with subarachnoid, subdural, and extradural hemorrhage, with prolonged [more than 24 hours] loss of consciousness, without return to pre-existing conscious level | S06.5X7A | Traumatic subdural hemorrhage with loss of consciousness of any duration with death due to brain injury before regaining consciousness, initial encounter |
| 803.75 | Other open skull fracture with subarachnoid, subdural, and extradural hemorrhage, with prolonged [more than 24 hours] loss of consciousness, without return to pre-existing conscious level | S06.5X8A | Traumatic subdural hemorrhage with loss of consciousness of any duration with death due to other cause before regaining consciousness, initial encounter |
| 803.75 | Other open skull fracture with subarachnoid, subdural, and extradural hemorrhage, with prolonged [more than 24 hours] loss of consciousness, without return to pre-existing conscious level | S06.6X6A | Traumatic subarachnoid hemorrhage with loss of consciousness greater than 24 hours without return to pre-existing conscious level with patient surviving, initial encounter |
| 803.75 | Other open skull fracture with subarachnoid, subdural, and extradural hemorrhage, with prolonged [more than 24 hours] loss of consciousness, without return to pre-existing conscious level | S06.6X7A | Traumatic subarachnoid hemorrhage with loss of consciousness of any duration with death due to brain injury prior to regaining consciousness, initial encounter |
| 803.75 | Other open skull fracture with subarachnoid, subdural, and extradural hemorrhage, with prolonged [more than 24 hours] loss of consciousness, without return to pre-existing conscious level | S06.6X8A | Traumatic subarachnoid hemorrhage with loss of consciousness of any duration with death due to other cause prior to regaining consciousness, initial encounter |
| 803.75 | Other open skull fracture with subarachnoid, subdural, and extradural hemorrhage, with prolonged [more than 24 hours] loss of consciousness, without return to pre-existing conscious level |  |  |
| 803.76 | Other open skull fracture with subarachnoid, subdural, and extradural hemorrhage, with loss of consciousness of unspecified duration | S06.4X9A | Epidural hemorrhage with loss of consciousness of unspecified duration, initial encounter |
| 803.76 | Other open skull fracture with subarachnoid, subdural, and extradural hemorrhage, with loss of consciousness of unspecified duration | S06.5X9A | Traumatic subdural hemorrhage with loss of consciousness of unspecified duration, initial encounter |
| 803.76 | Other open skull fracture with subarachnoid, subdural, and extradural hemorrhage, with loss of consciousness of unspecified duration | S06.6X9A | Traumatic subarachnoid hemorrhage with loss of consciousness of unspecified duration, initial encounter |
| 803.76 | Other open skull fracture with subarachnoid, subdural, and extradural hemorrhage, with loss of consciousness of unspecified duration |  |  |
| 803.79 | Other open skull fracture with subarachnoid, subdural, and extradural hemorrhage, with concussion, unspecified | S06.4X0A | Epidural hemorrhage without loss of consciousness, initial encounter |
| 803.79 | Other open skull fracture with subarachnoid, subdural, and extradural hemorrhage, with concussion, unspecified | S06.5X0A | Traumatic subdural hemorrhage without loss of consciousness, initial encounter |
| 803.79 | Other open skull fracture with subarachnoid, subdural, and extradural hemorrhage, with concussion, unspecified | S06.6X0A | Traumatic subarachnoid hemorrhage without loss of consciousness, initial encounter |
| 803.79 | Other open skull fracture with subarachnoid, subdural, and extradural hemorrhage, with concussion, unspecified |  |  |
| 803.80 | Other open skull fracture with other and unspecified intracranial hemorrhage, unspecified state of consciousness | S02.91XB | Unspecified fracture of skull, initial encounter for open fracture |
| 803.80 | Other open skull fracture with other and unspecified intracranial hemorrhage, unspecified state of consciousness | S06.360A | Traumatic hemorrhage of cerebrum, unspecified, without loss of consciousness, initial encounter |
| 803.81 | Other open skull fracture with other and unspecified intracranial hemorrhage, with no loss of consciousness | S02.91XB | Unspecified fracture of skull, initial encounter for open fracture |
| 803.81 | Other open skull fracture with other and unspecified intracranial hemorrhage, with no loss of consciousness | S06.360A | Traumatic hemorrhage of cerebrum, unspecified, without loss of consciousness, initial encounter |
| 803.82 | Other open skull fracture with other and unspecified intracranial hemorrhage, with brief [less than one hour] loss of consciousness | S02.91XB | Unspecified fracture of skull, initial encounter for open fracture |
| 803.82 | Other open skull fracture with other and unspecified intracranial hemorrhage, with brief [less than one hour] loss of consciousness | S06.361A | Traumatic hemorrhage of cerebrum, unspecified, with loss of consciousness of 30 minutes or less, initial encounter |
| 803.82 | Other open skull fracture with other and unspecified intracranial hemorrhage, with brief [less than one hour] loss of consciousness | S06.362A | Traumatic hemorrhage of cerebrum, unspecified, with loss of consciousness of 31 minutes to 59 minutes, initial encounter |
| 803.83 | Other open skull fracture with other and unspecified intracranial hemorrhage, with moderate [1-24 hours] loss of consciousness | S02.91XB | Unspecified fracture of skull, initial encounter for open fracture |
| 803.83 | Other open skull fracture with other and unspecified intracranial hemorrhage, with moderate [1-24 hours] loss of consciousness | S06.363A | Traumatic hemorrhage of cerebrum, unspecified, with loss of consciousness of 1 hours to 5 hours 59 minutes, initial encounter |
| 803.83 | Other open skull fracture with other and unspecified intracranial hemorrhage, with moderate [1-24 hours] loss of consciousness | S06.364A | Traumatic hemorrhage of cerebrum, unspecified, with loss of consciousness of 6 hours to 24 hours, initial encounter |
| 803.84 | Other open skull fracture with other and unspecified intracranial hemorrhage, with prolonged [more than 24 hours] loss of consciousness and return to pre-existing conscious level | S02.91XB | Unspecified fracture of skull, initial encounter for open fracture |
| 803.84 | Other open skull fracture with other and unspecified intracranial hemorrhage, with prolonged [more than 24 hours] loss of consciousness and return to pre-existing conscious level | S06.365A | Traumatic hemorrhage of cerebrum, unspecified, with loss of consciousness greater than 24 hours with return to pre-existing conscious level, initial encounter |
| 803.85 | Other open skull fracture with other and unspecified intracranial hemorrhage, with prolonged [more than 24 hours] loss of consciousness, without return to pre-existing conscious level | S02.91XB | Unspecified fracture of skull, initial encounter for open fracture |
| 803.85 | Other open skull fracture with other and unspecified intracranial hemorrhage, with prolonged [more than 24 hours] loss of consciousness, without return to pre-existing conscious level | S06.366A | Traumatic hemorrhage of cerebrum, unspecified, with loss of consciousness greater than 24 hours without return to pre-existing conscious level with patient surviving, initial encounter |
| 803.85 | Other open skull fracture with other and unspecified intracranial hemorrhage, with prolonged [more than 24 hours] loss of consciousness, without return to pre-existing conscious level | S06.367A | Traumatic hemorrhage of cerebrum, unspecified, with loss of consciousness of any duration with death due to brain injury prior to regaining consciousness, initial encounter |
| 803.85 | Other open skull fracture with other and unspecified intracranial hemorrhage, with prolonged [more than 24 hours] loss of consciousness, without return to pre-existing conscious level | S06.368A | Traumatic hemorrhage of cerebrum, unspecified, with loss of consciousness of any duration with death due to other cause prior to regaining consciousness, initial encounter |
| 803.86 | Other open skull fracture with other and unspecified intracranial hemorrhage, with loss of consciousness of unspecified duration | S02.91XB | Unspecified fracture of skull, initial encounter for open fracture |
| 803.86 | Other open skull fracture with other and unspecified intracranial hemorrhage, with loss of consciousness of unspecified duration | S06.369A | Traumatic hemorrhage of cerebrum, unspecified, with loss of consciousness of unspecified duration, initial encounter |
| 803.89 | Other open skull fracture with other and unspecified intracranial hemorrhage, with concussion, unspecified | S02.91XB | Unspecified fracture of skull, initial encounter for open fracture |
| 803.89 | Other open skull fracture with other and unspecified intracranial hemorrhage, with concussion, unspecified | S06.360A | Traumatic hemorrhage of cerebrum, unspecified, without loss of consciousness, initial encounter |
| 803.90 | Other open skull fracture with intracranial injury of other and unspecified nature, unspecified state of consciousness | S02.91XB | Unspecified fracture of skull, initial encounter for open fracture |
| 803.90 | Other open skull fracture with intracranial injury of other and unspecified nature, unspecified state of consciousness | S06.890A | Other specified intracranial injury without loss of consciousness, initial encounter |
| 803.90 | Other open skull fracture with intracranial injury of other and unspecified nature, unspecified state of consciousness | S06.9X0A | Unspecified intracranial injury without loss of consciousness, initial encounter |
| 803.91 | Other open skull fracture with intracranial injury of other and unspecified nature, with no loss of consciousness | S02.91XB | Unspecified fracture of skull, initial encounter for open fracture |
| 803.91 | Other open skull fracture with intracranial injury of other and unspecified nature, with no loss of consciousness | S06.890A | Other specified intracranial injury without loss of consciousness, initial encounter |
| 803.91 | Other open skull fracture with intracranial injury of other and unspecified nature, with no loss of consciousness | S06.9X0A | Unspecified intracranial injury without loss of consciousness, initial encounter |
| 803.92 | Other open skull fracture with intracranial injury of other and unspecified nature, with brief [less than one hour] loss of consciousness | S02.91XB | Unspecified fracture of skull, initial encounter for open fracture |
| 803.92 | Other open skull fracture with intracranial injury of other and unspecified nature, with brief [less than one hour] loss of consciousness | S06.891A | Other specified intracranial injury with loss of consciousness of 30 minutes or less, initial encounter |
| 803.92 | Other open skull fracture with intracranial injury of other and unspecified nature, with brief [less than one hour] loss of consciousness | S06.892A | Other specified intracranial injury with loss of consciousness of 31 minutes to 59 minutes, initial encounter |
| 803.92 | Other open skull fracture with intracranial injury of other and unspecified nature, with brief [less than one hour] loss of consciousness | S06.9X1A | Unspecified intracranial injury with loss of consciousness of 30 minutes or less, initial encounter |
| 803.92 | Other open skull fracture with intracranial injury of other and unspecified nature, with brief [less than one hour] loss of consciousness | S06.9X2A | Unspecified intracranial injury with loss of consciousness of 31 minutes to 59 minutes, initial encounter |
| 803.93 | Other open skull fracture with intracranial injury of other and unspecified nature, with moderate [1-24 hours] loss of consciousness | S02.91XB | Unspecified fracture of skull, initial encounter for open fracture |
| 803.93 | Other open skull fracture with intracranial injury of other and unspecified nature, with moderate [1-24 hours] loss of consciousness | S06.893A | Other specified intracranial injury with loss of consciousness of 1 hour to 5 hours 59 minutes, initial encounter |
| 803.93 | Other open skull fracture with intracranial injury of other and unspecified nature, with moderate [1-24 hours] loss of consciousness | S06.894A | Other specified intracranial injury with loss of consciousness of 6 hours to 24 hours, initial encounter |
| 803.93 | Other open skull fracture with intracranial injury of other and unspecified nature, with moderate [1-24 hours] loss of consciousness | S06.9X3A | Unspecified intracranial injury with loss of consciousness of 1 hour to 5 hours 59 minutes, initial encounter |
| 803.93 | Other open skull fracture with intracranial injury of other and unspecified nature, with moderate [1-24 hours] loss of consciousness | S06.9X4A | Unspecified intracranial injury with loss of consciousness of 6 hours to 24 hours, initial encounter |
| 803.94 | Other open skull fracture with intracranial injury of other and unspecified nature, with prolonged [more than 24 hours] loss of consciousness and return to pre-existing conscious level | S02.91XB | Unspecified fracture of skull, initial encounter for open fracture |
| 803.94 | Other open skull fracture with intracranial injury of other and unspecified nature, with prolonged [more than 24 hours] loss of consciousness and return to pre-existing conscious level | S06.895A | Other specified intracranial injury with loss of consciousness greater than 24 hours with return to pre-existing conscious level, initial encounter |
| 803.94 | Other open skull fracture with intracranial injury of other and unspecified nature, with prolonged [more than 24 hours] loss of consciousness and return to pre-existing conscious level | S06.9X5A | Unspecified intracranial injury with loss of consciousness greater than 24 hours with return to pre-existing conscious level, initial encounter |
| 803.95 | Other open skull fracture with intracranial injury of other and unspecified nature, with prolonged [more than 24 hours] loss of consciousness, without return to pre-existing conscious level | S02.91XB | Unspecified fracture of skull, initial encounter for open fracture |
| 803.95 | Other open skull fracture with intracranial injury of other and unspecified nature, with prolonged [more than 24 hours] loss of consciousness, without return to pre-existing conscious level | S06.896A | Other specified intracranial injury with loss of consciousness greater than 24 hours without return to pre-existing conscious level with patient surviving, initial encounter |
| 803.95 | Other open skull fracture with intracranial injury of other and unspecified nature, with prolonged [more than 24 hours] loss of consciousness, without return to pre-existing conscious level | S06.897A | Other specified intracranial injury with loss of consciousness of any duration with death due to brain injury prior to regaining consciousness, initial encounter |
| 803.95 | Other open skull fracture with intracranial injury of other and unspecified nature, with prolonged [more than 24 hours] loss of consciousness, without return to pre-existing conscious level | S06.898A | Other specified intracranial injury with loss of consciousness of any duration with death due to other cause prior to regaining consciousness, initial encounter |
| 803.95 | Other open skull fracture with intracranial injury of other and unspecified nature, with prolonged [more than 24 hours] loss of consciousness, without return to pre-existing conscious level | S06.9X6A | Unspecified intracranial injury with loss of consciousness greater than 24 hours without return to pre-existing conscious level with patient surviving, initial encounter |
| 803.95 | Other open skull fracture with intracranial injury of other and unspecified nature, with prolonged [more than 24 hours] loss of consciousness, without return to pre-existing conscious level | S06.9X7A | Unspecified intracranial injury with loss of consciousness of any duration with death due to brain injury prior to regaining consciousness, initial encounter |
| 803.95 | Other open skull fracture with intracranial injury of other and unspecified nature, with prolonged [more than 24 hours] loss of consciousness, without return to pre-existing conscious level | S06.9X8A | Unspecified intracranial injury with loss of consciousness of any duration with death due to other cause prior to regaining consciousness, initial encounter |
| 803.96 | Other open skull fracture with intracranial injury of other and unspecified nature, with loss of consciousness of unspecified duration | S02.91XB | Unspecified fracture of skull, initial encounter for open fracture |
| 803.96 | Other open skull fracture with intracranial injury of other and unspecified nature, with loss of consciousness of unspecified duration | S06.899A | Other specified intracranial injury with loss of consciousness of unspecified duration, initial encounter |
| 803.96 | Other open skull fracture with intracranial injury of other and unspecified nature, with loss of consciousness of unspecified duration | S06.9X9A | Unspecified intracranial injury with loss of consciousness of unspecified duration, initial encounter |
| 803.99 | Other open skull fracture with intracranial injury of other and unspecified nature, with concussion, unspecified | S02.91XB | Unspecified fracture of skull, initial encounter for open fracture |
| 803.99 | Other open skull fracture with intracranial injury of other and unspecified nature, with concussion, unspecified | S06.890A | Other specified intracranial injury without loss of consciousness, initial encounter |
| 803.99 | Other open skull fracture with intracranial injury of other and unspecified nature, with concussion, unspecified | S06.9X0A | Unspecified intracranial injury without loss of consciousness, initial encounter |
| 804.10 | Closed fractures involving skull or face with other bones, with cerebral laceration and contusion, unspecified state of consciousness | S02.91XA | Unspecified fracture of skull, initial encounter for closed fracture |
| 804.10 | Closed fractures involving skull or face with other bones, with cerebral laceration and contusion, unspecified state of consciousness | S06.330A | Contusion and laceration of cerebrum, unspecified, without loss of consciousness, initial encounter |
| 804.11 | Closed fractures involving skull or face with other bones, with cerebral laceration and contusion, with no loss of consciousness | S02.91XA | Unspecified fracture of skull, initial encounter for closed fracture |
| 804.11 | Closed fractures involving skull or face with other bones, with cerebral laceration and contusion, with no loss of consciousness | S06.330A | Contusion and laceration of cerebrum, unspecified, without loss of consciousness, initial encounter |
| 804.12 | Closed fractures involving skull or face with other bones, with cerebral laceration and contusion, with brief [less than one hour] loss of consciousness | S02.91XA | Unspecified fracture of skull, initial encounter for closed fracture |
| 804.12 | Closed fractures involving skull or face with other bones, with cerebral laceration and contusion, with brief [less than one hour] loss of consciousness | S06.331A | Contusion and laceration of cerebrum, unspecified, with loss of consciousness of 30 minutes or less, initial encounter |
| 804.12 | Closed fractures involving skull or face with other bones, with cerebral laceration and contusion, with brief [less than one hour] loss of consciousness | S06.332A | Contusion and laceration of cerebrum, unspecified, with loss of consciousness of 31 minutes to 59 minutes, initial encounter |
| 804.13 | Closed fractures involving skull or face with other bones, with cerebral laceration and contusion, with moderate [1-24 hours] loss of consciousness | S02.91XA | Unspecified fracture of skull, initial encounter for closed fracture |
| 804.13 | Closed fractures involving skull or face with other bones, with cerebral laceration and contusion, with moderate [1-24 hours] loss of consciousness | S06.333A | Contusion and laceration of cerebrum, unspecified, with loss of consciousness of 1 hour to 5 hours 59 minutes, initial encounter |
| 804.13 | Closed fractures involving skull or face with other bones, with cerebral laceration and contusion, with moderate [1-24 hours] loss of consciousness | S06.334A | Contusion and laceration of cerebrum, unspecified, with loss of consciousness of 6 hours to 24 hours, initial encounter |
| 804.14 | Closed fractures involving skull or face with other bones, with cerebral laceration and contusion, with prolonged [more than 24 hours] loss of consciousness and return to pre-existing conscious level | S02.91XA | Unspecified fracture of skull, initial encounter for closed fracture |
| 804.14 | Closed fractures involving skull or face with other bones, with cerebral laceration and contusion, with prolonged [more than 24 hours] loss of consciousness and return to pre-existing conscious level | S06.335A | Contusion and laceration of cerebrum, unspecified, with loss of consciousness greater than 24 hours with return to pre-existing conscious level, initial encounter |
| 804.15 | Closed fractures involving skull or face with other bones, with cerebral laceration and contusion, with prolonged [more than 24 hours] loss of consciousness, without return to pre-existing conscious level | S02.91XA | Unspecified fracture of skull, initial encounter for closed fracture |
| 804.15 | Closed fractures involving skull or face with other bones, with cerebral laceration and contusion, with prolonged [more than 24 hours] loss of consciousness, without return to pre-existing conscious level | S06.336A | Contusion and laceration of cerebrum, unspecified, with loss of consciousness greater than 24 hours without return to pre-existing conscious level with patient surviving, initial encounter |
| 804.15 | Closed fractures involving skull or face with other bones, with cerebral laceration and contusion, with prolonged [more than 24 hours] loss of consciousness, without return to pre-existing conscious level | S06.337A | Contusion and laceration of cerebrum, unspecified, with loss of consciousness of any duration with death due to brain injury prior to regaining consciousness, initial encounter |
| 804.15 | Closed fractures involving skull or face with other bones, with cerebral laceration and contusion, with prolonged [more than 24 hours] loss of consciousness, without return to pre-existing conscious level | S06.338A | Contusion and laceration of cerebrum, unspecified, with loss of consciousness of any duration with death due to other cause prior to regaining consciousness, initial encounter |
| 804.16 | Closed fractures involving skull or face with other bones, with cerebral laceration and contusion, with loss of consciousness of unspecified duration | S02.91XA | Unspecified fracture of skull, initial encounter for closed fracture |
| 804.16 | Closed fractures involving skull or face with other bones, with cerebral laceration and contusion, with loss of consciousness of unspecified duration | S06.339A | Contusion and laceration of cerebrum, unspecified, with loss of consciousness of unspecified duration, initial encounter |
| 804.19 | Closed fractures involving skull or face with other bones, with cerebral laceration and contusion, with concussion, unspecified | S02.91XA | Unspecified fracture of skull, initial encounter for closed fracture |
| 804.19 | Closed fractures involving skull or face with other bones, with cerebral laceration and contusion, with concussion, unspecified | S06.330A | Contusion and laceration of cerebrum, unspecified, without loss of consciousness, initial encounter |
| 804.20 | Closed fractures involving skull or face with other bones with subarachnoid, subdural, and extradural hemorrhage, unspecified state of consciousness | S06.4X0A | Epidural hemorrhage without loss of consciousness, initial encounter |
| 804.20 | Closed fractures involving skull or face with other bones with subarachnoid, subdural, and extradural hemorrhage, unspecified state of consciousness | S06.5X0A | Traumatic subdural hemorrhage without loss of consciousness, initial encounter |
| 804.20 | Closed fractures involving skull or face with other bones with subarachnoid, subdural, and extradural hemorrhage, unspecified state of consciousness | S06.6X0A | Traumatic subarachnoid hemorrhage without loss of consciousness, initial encounter |
| 804.20 | Closed fractures involving skull or face with other bones with subarachnoid, subdural, and extradural hemorrhage, unspecified state of consciousness |  |  |
| 804.21 | Closed fractures involving skull or face with other bones with subarachnoid, subdural, and extradural hemorrhage, with no loss of consciousness | S06.4X0A | Epidural hemorrhage without loss of consciousness, initial encounter |
| 804.21 | Closed fractures involving skull or face with other bones with subarachnoid, subdural, and extradural hemorrhage, with no loss of consciousness | S06.5X0A | Traumatic subdural hemorrhage without loss of consciousness, initial encounter |
| 804.21 | Closed fractures involving skull or face with other bones with subarachnoid, subdural, and extradural hemorrhage, with no loss of consciousness | S06.6X0A | Traumatic subarachnoid hemorrhage without loss of consciousness, initial encounter |
| 804.21 | Closed fractures involving skull or face with other bones with subarachnoid, subdural, and extradural hemorrhage, with no loss of consciousness |  |  |
| 804.22 | Closed fractures involving skull or face with other bones with subarachnoid, subdural, and extradural hemorrhage, with brief [less than one hour] loss of consciousness | S06.4X1A | Epidural hemorrhage with loss of consciousness of 30 minutes or less, initial encounter |
| 804.22 | Closed fractures involving skull or face with other bones with subarachnoid, subdural, and extradural hemorrhage, with brief [less than one hour] loss of consciousness | S06.4X2A | Epidural hemorrhage with loss of consciousness of 31 minutes to 59 minutes, initial encounter |
| 804.22 | Closed fractures involving skull or face with other bones with subarachnoid, subdural, and extradural hemorrhage, with brief [less than one hour] loss of consciousness | S06.5X1A | Traumatic subdural hemorrhage with loss of consciousness of 30 minutes or less, initial encounter |
| 804.22 | Closed fractures involving skull or face with other bones with subarachnoid, subdural, and extradural hemorrhage, with brief [less than one hour] loss of consciousness | S06.5X2A | Traumatic subdural hemorrhage with loss of consciousness of 31 minutes to 59 minutes, initial encounter |
| 804.22 | Closed fractures involving skull or face with other bones with subarachnoid, subdural, and extradural hemorrhage, with brief [less than one hour] loss of consciousness | S06.6X1A | Traumatic subarachnoid hemorrhage with loss of consciousness of 30 minutes or less, initial encounter |
| 804.22 | Closed fractures involving skull or face with other bones with subarachnoid, subdural, and extradural hemorrhage, with brief [less than one hour] loss of consciousness | S06.6X2A | Traumatic subarachnoid hemorrhage with loss of consciousness of 31 minutes to 59 minutes, initial encounter |
| 804.22 | Closed fractures involving skull or face with other bones with subarachnoid, subdural, and extradural hemorrhage, with brief [less than one hour] loss of consciousness |  |  |
| 804.23 | Closed fractures involving skull or face with other bones with subarachnoid, subdural, and extradural hemorrhage, with moderate [1-24 hours] loss of consciousness | S06.4X3A | Epidural hemorrhage with loss of consciousness of 1 hour to 5 hours 59 minutes, initial encounter |
| 804.23 | Closed fractures involving skull or face with other bones with subarachnoid, subdural, and extradural hemorrhage, with moderate [1-24 hours] loss of consciousness | S06.4X4A | Epidural hemorrhage with loss of consciousness of 6 hours to 24 hours, initial encounter |
| 804.23 | Closed fractures involving skull or face with other bones with subarachnoid, subdural, and extradural hemorrhage, with moderate [1-24 hours] loss of consciousness | S06.5X3A | Traumatic subdural hemorrhage with loss of consciousness of 1 hour to 5 hours 59 minutes, initial encounter |
| 804.23 | Closed fractures involving skull or face with other bones with subarachnoid, subdural, and extradural hemorrhage, with moderate [1-24 hours] loss of consciousness | S06.5X4A | Traumatic subdural hemorrhage with loss of consciousness of 6 hours to 24 hours, initial encounter |
| 804.23 | Closed fractures involving skull or face with other bones with subarachnoid, subdural, and extradural hemorrhage, with moderate [1-24 hours] loss of consciousness | S06.6X3A | Traumatic subarachnoid hemorrhage with loss of consciousness of 1 hour to 5 hours 59 minutes, initial encounter |
| 804.23 | Closed fractures involving skull or face with other bones with subarachnoid, subdural, and extradural hemorrhage, with moderate [1-24 hours] loss of consciousness | S06.6X4A | Traumatic subarachnoid hemorrhage with loss of consciousness of 6 hours to 24 hours, initial encounter |
| 804.23 | Closed fractures involving skull or face with other bones with subarachnoid, subdural, and extradural hemorrhage, with moderate [1-24 hours] loss of consciousness |  |  |
| 804.24 | Closed fractures involving skull or face with other bones with subarachnoid, subdural, and extradural hemorrhage, with prolonged [more than 24 hours] loss of consciousness and return to pre-existing conscious level | S06.4X5A | Epidural hemorrhage with loss of consciousness greater than 24 hours with return to pre-existing conscious level, initial encounter |
| 804.24 | Closed fractures involving skull or face with other bones with subarachnoid, subdural, and extradural hemorrhage, with prolonged [more than 24 hours] loss of consciousness and return to pre-existing conscious level | S06.5X5A | Traumatic subdural hemorrhage with loss of consciousness greater than 24 hours with return to pre-existing conscious level, initial encounter |
| 804.24 | Closed fractures involving skull or face with other bones with subarachnoid, subdural, and extradural hemorrhage, with prolonged [more than 24 hours] loss of consciousness and return to pre-existing conscious level | S06.6X5A | Traumatic subarachnoid hemorrhage with loss of consciousness greater than 24 hours with return to pre-existing conscious level, initial encounter |
| 804.24 | Closed fractures involving skull or face with other bones with subarachnoid, subdural, and extradural hemorrhage, with prolonged [more than 24 hours] loss of consciousness and return to pre-existing conscious level |  |  |
| 804.25 | Closed fractures involving skull or face with other bones with subarachnoid, subdural, and extradural hemorrhage, with prolonged [more than 24 hours] loss of consciousness, without return to pre-existing conscious level | S06.4X6A | Epidural hemorrhage with loss of consciousness greater than 24 hours without return to pre-existing conscious level with patient surviving, initial encounter |
| 804.25 | Closed fractures involving skull or face with other bones with subarachnoid, subdural, and extradural hemorrhage, with prolonged [more than 24 hours] loss of consciousness, without return to pre-existing conscious level | S06.4X7A | Epidural hemorrhage with loss of consciousness of any duration with death due to brain injury prior to regaining consciousness, initial encounter |
| 804.25 | Closed fractures involving skull or face with other bones with subarachnoid, subdural, and extradural hemorrhage, with prolonged [more than 24 hours] loss of consciousness, without return to pre-existing conscious level | S06.4X8A | Epidural hemorrhage with loss of consciousness of any duration with death due to other causes prior to regaining consciousness, initial encounter |
| 804.25 | Closed fractures involving skull or face with other bones with subarachnoid, subdural, and extradural hemorrhage, with prolonged [more than 24 hours] loss of consciousness, without return to pre-existing conscious level | S06.5X6A | Traumatic subdural hemorrhage with loss of consciousness greater than 24 hours without return to pre-existing conscious level with patient surviving, initial encounter |
| 804.25 | Closed fractures involving skull or face with other bones with subarachnoid, subdural, and extradural hemorrhage, with prolonged [more than 24 hours] loss of consciousness, without return to pre-existing conscious level | S06.5X7A | Traumatic subdural hemorrhage with loss of consciousness of any duration with death due to brain injury before regaining consciousness, initial encounter |
| 804.25 | Closed fractures involving skull or face with other bones with subarachnoid, subdural, and extradural hemorrhage, with prolonged [more than 24 hours] loss of consciousness, without return to pre-existing conscious level | S06.5X8A | Traumatic subdural hemorrhage with loss of consciousness of any duration with death due to other cause before regaining consciousness, initial encounter |
| 804.25 | Closed fractures involving skull or face with other bones with subarachnoid, subdural, and extradural hemorrhage, with prolonged [more than 24 hours] loss of consciousness, without return to pre-existing conscious level | S06.6X6A | Traumatic subarachnoid hemorrhage with loss of consciousness greater than 24 hours without return to pre-existing conscious level with patient surviving, initial encounter |
| 804.25 | Closed fractures involving skull or face with other bones with subarachnoid, subdural, and extradural hemorrhage, with prolonged [more than 24 hours] loss of consciousness, without return to pre-existing conscious level | S06.6X7A | Traumatic subarachnoid hemorrhage with loss of consciousness of any duration with death due to brain injury prior to regaining consciousness, initial encounter |
| 804.25 | Closed fractures involving skull or face with other bones with subarachnoid, subdural, and extradural hemorrhage, with prolonged [more than 24 hours] loss of consciousness, without return to pre-existing conscious level | S06.6X8A | Traumatic subarachnoid hemorrhage with loss of consciousness of any duration with death due to other cause prior to regaining consciousness, initial encounter |
| 804.25 | Closed fractures involving skull or face with other bones with subarachnoid, subdural, and extradural hemorrhage, with prolonged [more than 24 hours] loss of consciousness, without return to pre-existing conscious level |  |  |
| 804.26 | Closed fractures involving skull or face with other bones with subarachnoid, subdural, and extradural hemorrhage, with loss of consciousness of unspecified duration | S06.4X9A | Epidural hemorrhage with loss of consciousness of unspecified duration, initial encounter |
| 804.26 | Closed fractures involving skull or face with other bones with subarachnoid, subdural, and extradural hemorrhage, with loss of consciousness of unspecified duration | S06.5X9A | Traumatic subdural hemorrhage with loss of consciousness of unspecified duration, initial encounter |
| 804.26 | Closed fractures involving skull or face with other bones with subarachnoid, subdural, and extradural hemorrhage, with loss of consciousness of unspecified duration | S06.6X9A | Traumatic subarachnoid hemorrhage with loss of consciousness of unspecified duration, initial encounter |
| 804.26 | Closed fractures involving skull or face with other bones with subarachnoid, subdural, and extradural hemorrhage, with loss of consciousness of unspecified duration |  |  |
| 804.29 | Closed fractures involving skull or face with other bones with subarachnoid, subdural, and extradural hemorrhage, with concussion, unspecified | S06.4X0A | Epidural hemorrhage without loss of consciousness, initial encounter |
| 804.29 | Closed fractures involving skull or face with other bones with subarachnoid, subdural, and extradural hemorrhage, with concussion, unspecified | S06.5X0A | Traumatic subdural hemorrhage without loss of consciousness, initial encounter |
| 804.29 | Closed fractures involving skull or face with other bones with subarachnoid, subdural, and extradural hemorrhage, with concussion, unspecified | S06.6X0A | Traumatic subarachnoid hemorrhage without loss of consciousness, initial encounter |
| 804.29 | Closed fractures involving skull or face with other bones with subarachnoid, subdural, and extradural hemorrhage, with concussion, unspecified |  |  |
| 804.30 | Closed fractures involving skull or face with other bones, with other and unspecified intracranial hemorrhage, unspecified state of consciousness | S02.91XA | Unspecified fracture of skull, initial encounter for closed fracture |
| 804.30 | Closed fractures involving skull or face with other bones, with other and unspecified intracranial hemorrhage, unspecified state of consciousness | S06.360A | Traumatic hemorrhage of cerebrum, unspecified, without loss of consciousness, initial encounter |
| 804.31 | Closed fractures involving skull or face with other bones, with other and unspecified intracranial hemorrhage, with no loss of consciousness | S02.91XA | Unspecified fracture of skull, initial encounter for closed fracture |
| 804.31 | Closed fractures involving skull or face with other bones, with other and unspecified intracranial hemorrhage, with no loss of consciousness | S06.360A | Traumatic hemorrhage of cerebrum, unspecified, without loss of consciousness, initial encounter |
| 804.32 | Closed fractures involving skull or face with other bones, with other and unspecified intracranial hemorrhage, with brief [less than one hour] loss of consciousness | S02.91XA | Unspecified fracture of skull, initial encounter for closed fracture |
| 804.32 | Closed fractures involving skull or face with other bones, with other and unspecified intracranial hemorrhage, with brief [less than one hour] loss of consciousness | S06.361A | Traumatic hemorrhage of cerebrum, unspecified, with loss of consciousness of 30 minutes or less, initial encounter |
| 804.32 | Closed fractures involving skull or face with other bones, with other and unspecified intracranial hemorrhage, with brief [less than one hour] loss of consciousness | S06.362A | Traumatic hemorrhage of cerebrum, unspecified, with loss of consciousness of 31 minutes to 59 minutes, initial encounter |
| 804.33 | Closed fractures involving skull or face with other bones, with other and unspecified intracranial hemorrhage, with moderate [1-24 hours] loss of consciousness | S02.91XA | Unspecified fracture of skull, initial encounter for closed fracture |
| 804.33 | Closed fractures involving skull or face with other bones, with other and unspecified intracranial hemorrhage, with moderate [1-24 hours] loss of consciousness | S06.363A | Traumatic hemorrhage of cerebrum, unspecified, with loss of consciousness of 1 hours to 5 hours 59 minutes, initial encounter |
| 804.33 | Closed fractures involving skull or face with other bones, with other and unspecified intracranial hemorrhage, with moderate [1-24 hours] loss of consciousness | S06.364A | Traumatic hemorrhage of cerebrum, unspecified, with loss of consciousness of 6 hours to 24 hours, initial encounter |
| 804.34 | Closed fractures involving skull or face with other bones, with other and unspecified intracranial hemorrhage, with prolonged [more than 24 hours] loss of consciousness and return to pre- existing conscious level | S02.91XA | Unspecified fracture of skull, initial encounter for closed fracture |
| 804.34 | Closed fractures involving skull or face with other bones, with other and unspecified intracranial hemorrhage, with prolonged [more than 24 hours] loss of consciousness and return to pre- existing conscious level | S06.365A | Traumatic hemorrhage of cerebrum, unspecified, with loss of consciousness greater than 24 hours with return to pre-existing conscious level, initial encounter |
| 804.35 | Closed fractures involving skull or face with other bones, with other and unspecified intracranial hemorrhage, with prolonged [more than 24 hours] loss of consciousness, without return to pre-existing conscious level | S02.91XA | Unspecified fracture of skull, initial encounter for closed fracture |
| 804.35 | Closed fractures involving skull or face with other bones, with other and unspecified intracranial hemorrhage, with prolonged [more than 24 hours] loss of consciousness, without return to pre-existing conscious level | S06.366A | Traumatic hemorrhage of cerebrum, unspecified, with loss of consciousness greater than 24 hours without return to pre-existing conscious level with patient surviving, initial encounter |
| 804.35 | Closed fractures involving skull or face with other bones, with other and unspecified intracranial hemorrhage, with prolonged [more than 24 hours] loss of consciousness, without return to pre-existing conscious level | S06.367A | Traumatic hemorrhage of cerebrum, unspecified, with loss of consciousness of any duration with death due to brain injury prior to regaining consciousness, initial encounter |
| 804.35 | Closed fractures involving skull or face with other bones, with other and unspecified intracranial hemorrhage, with prolonged [more than 24 hours] loss of consciousness, without return to pre-existing conscious level | S06.368A | Traumatic hemorrhage of cerebrum, unspecified, with loss of consciousness of any duration with death due to other cause prior to regaining consciousness, initial encounter |
| 804.36 | Closed fractures involving skull or face with other bones, with other and unspecified intracranial hemorrhage, with loss of consciousness of unspecified duration | S02.91XA | Unspecified fracture of skull, initial encounter for closed fracture |
| 804.36 | Closed fractures involving skull or face with other bones, with other and unspecified intracranial hemorrhage, with loss of consciousness of unspecified duration | S06.369A | Traumatic hemorrhage of cerebrum, unspecified, with loss of consciousness of unspecified duration, initial encounter |
| 804.39 | Closed fractures involving skull or face with other bones, with other and unspecified intracranial hemorrhage, with concussion, unspecified | S02.91XA | Unspecified fracture of skull, initial encounter for closed fracture |
| 804.39 | Closed fractures involving skull or face with other bones, with other and unspecified intracranial hemorrhage, with concussion, unspecified | S06.360A | Traumatic hemorrhage of cerebrum, unspecified, without loss of consciousness, initial encounter |
| 804.40 | Closed fractures involving skull or face with other bones, with intracranial injury of other and unspecified nature, unspecified state of consciousness | S02.91XA | Unspecified fracture of skull, initial encounter for closed fracture |
| 804.40 | Closed fractures involving skull or face with other bones, with intracranial injury of other and unspecified nature, unspecified state of consciousness | S06.890A | Other specified intracranial injury without loss of consciousness, initial encounter |
| 804.40 | Closed fractures involving skull or face with other bones, with intracranial injury of other and unspecified nature, unspecified state of consciousness | S06.9X0A | Unspecified intracranial injury without loss of consciousness, initial encounter |
| 804.41 | Closed fractures involving skull or face with other bones, with intracranial injury of other and unspecified nature, with no loss of consciousness | S02.91XA | Unspecified fracture of skull, initial encounter for closed fracture |
| 804.41 | Closed fractures involving skull or face with other bones, with intracranial injury of other and unspecified nature, with no loss of consciousness | S06.890A | Other specified intracranial injury without loss of consciousness, initial encounter |
| 804.41 | Closed fractures involving skull or face with other bones, with intracranial injury of other and unspecified nature, with no loss of consciousness | S06.9X0A | Unspecified intracranial injury without loss of consciousness, initial encounter |
| 804.42 | Closed fractures involving skull or face with other bones, with intracranial injury of other and unspecified nature, with brief [less than one hour] loss of consciousness | S02.91XA | Unspecified fracture of skull, initial encounter for closed fracture |
| 804.42 | Closed fractures involving skull or face with other bones, with intracranial injury of other and unspecified nature, with brief [less than one hour] loss of consciousness | S06.891A | Other specified intracranial injury with loss of consciousness of 30 minutes or less, initial encounter |
| 804.42 | Closed fractures involving skull or face with other bones, with intracranial injury of other and unspecified nature, with brief [less than one hour] loss of consciousness | S06.892A | Other specified intracranial injury with loss of consciousness of 31 minutes to 59 minutes, initial encounter |
| 804.42 | Closed fractures involving skull or face with other bones, with intracranial injury of other and unspecified nature, with brief [less than one hour] loss of consciousness | S06.9X1A | Unspecified intracranial injury with loss of consciousness of 30 minutes or less, initial encounter |
| 804.42 | Closed fractures involving skull or face with other bones, with intracranial injury of other and unspecified nature, with brief [less than one hour] loss of consciousness | S06.9X2A | Unspecified intracranial injury with loss of consciousness of 31 minutes to 59 minutes, initial encounter |
| 804.43 | Closed fractures involving skull or face with other bones, with intracranial injury of other and unspecified nature, with moderate [1-24 hours] loss of consciousness | S02.91XA | Unspecified fracture of skull, initial encounter for closed fracture |
| 804.43 | Closed fractures involving skull or face with other bones, with intracranial injury of other and unspecified nature, with moderate [1-24 hours] loss of consciousness | S06.893A | Other specified intracranial injury with loss of consciousness of 1 hour to 5 hours 59 minutes, initial encounter |
| 804.43 | Closed fractures involving skull or face with other bones, with intracranial injury of other and unspecified nature, with moderate [1-24 hours] loss of consciousness | S06.894A | Other specified intracranial injury with loss of consciousness of 6 hours to 24 hours, initial encounter |
| 804.43 | Closed fractures involving skull or face with other bones, with intracranial injury of other and unspecified nature, with moderate [1-24 hours] loss of consciousness | S06.9X4A | Unspecified intracranial injury with loss of consciousness of 6 hours to 24 hours, initial encounter |
| 804.44 | Closed fractures involving skull or face with other bones, with intracranial injury of other and unspecified nature, with prolonged [more than 24 hours] loss of consciousness and return to pre-existing conscious level | S02.91XA | Unspecified fracture of skull, initial encounter for closed fracture |
| 804.44 | Closed fractures involving skull or face with other bones, with intracranial injury of other and unspecified nature, with prolonged [more than 24 hours] loss of consciousness and return to pre-existing conscious level | S06.895A | Other specified intracranial injury with loss of consciousness greater than 24 hours with return to pre-existing conscious level, initial encounter |
| 804.44 | Closed fractures involving skull or face with other bones, with intracranial injury of other and unspecified nature, with prolonged [more than 24 hours] loss of consciousness and return to pre-existing conscious level | S06.9X5A | Unspecified intracranial injury with loss of consciousness greater than 24 hours with return to pre-existing conscious level, initial encounter |
| 804.45 | Closed fractures involving skull or face with other bones, with intracranial injury of other and unspecified nature, with prolonged [more than 24 hours] loss of consciousness, without return to pre-existing conscious level | S02.91XA | Unspecified fracture of skull, initial encounter for closed fracture |
| 804.45 | Closed fractures involving skull or face with other bones, with intracranial injury of other and unspecified nature, with prolonged [more than 24 hours] loss of consciousness, without return to pre-existing conscious level | S06.896A | Other specified intracranial injury with loss of consciousness greater than 24 hours without return to pre-existing conscious level with patient surviving, initial encounter |
| 804.45 | Closed fractures involving skull or face with other bones, with intracranial injury of other and unspecified nature, with prolonged [more than 24 hours] loss of consciousness, without return to pre-existing conscious level | S06.897A | Other specified intracranial injury with loss of consciousness of any duration with death due to brain injury prior to regaining consciousness, initial encounter |
| 804.45 | Closed fractures involving skull or face with other bones, with intracranial injury of other and unspecified nature, with prolonged [more than 24 hours] loss of consciousness, without return to pre-existing conscious level | S06.898A | Other specified intracranial injury with loss of consciousness of any duration with death due to other cause prior to regaining consciousness, initial encounter |
| 804.45 | Closed fractures involving skull or face with other bones, with intracranial injury of other and unspecified nature, with prolonged [more than 24 hours] loss of consciousness, without return to pre-existing conscious level | S06.9X6A | Unspecified intracranial injury with loss of consciousness greater than 24 hours without return to pre-existing conscious level with patient surviving, initial encounter |
| 804.45 | Closed fractures involving skull or face with other bones, with intracranial injury of other and unspecified nature, with prolonged [more than 24 hours] loss of consciousness, without return to pre-existing conscious level | S06.9X7A | Unspecified intracranial injury with loss of consciousness of any duration with death due to brain injury prior to regaining consciousness, initial encounter |
| 804.45 | Closed fractures involving skull or face with other bones, with intracranial injury of other and unspecified nature, with prolonged [more than 24 hours] loss of consciousness, without return to pre-existing conscious level | S06.9X8A | Unspecified intracranial injury with loss of consciousness of any duration with death due to other cause prior to regaining consciousness, initial encounter |
| 804.46 | Closed fractures involving skull or face with other bones, with intracranial injury of other and unspecified nature, with loss of consciousness of unspecified duration | S02.91XA | Unspecified fracture of skull, initial encounter for closed fracture |
| 804.46 | Closed fractures involving skull or face with other bones, with intracranial injury of other and unspecified nature, with loss of consciousness of unspecified duration | S06.899A | Other specified intracranial injury with loss of consciousness of unspecified duration, initial encounter |
| 804.46 | Closed fractures involving skull or face with other bones, with intracranial injury of other and unspecified nature, with loss of consciousness of unspecified duration | S06.9X9A | Unspecified intracranial injury with loss of consciousness of unspecified duration, initial encounter |
| 804.49 | Closed fractures involving skull or face with other bones, with intracranial injury of other and unspecified nature, with concussion, unspecified | S02.91XA | Unspecified fracture of skull, initial encounter for closed fracture |
| 804.49 | Closed fractures involving skull or face with other bones, with intracranial injury of other and unspecified nature, with concussion, unspecified | S06.890A | Other specified intracranial injury without loss of consciousness, initial encounter |
| 804.49 | Closed fractures involving skull or face with other bones, with intracranial injury of other and unspecified nature, with concussion, unspecified | S06.9X9A | Unspecified intracranial injury with loss of consciousness of unspecified duration, initial encounter |
| 804.60 | Open fractures involving skull or face with other bones, with cerebral laceration and contusion, unspecified state of consciousness | S02.91XB | Unspecified fracture of skull, initial encounter for open fracture |
| 804.60 | Open fractures involving skull or face with other bones, with cerebral laceration and contusion, unspecified state of consciousness | S06.330A | Contusion and laceration of cerebrum, unspecified, without loss of consciousness, initial encounter |
| 804.61 | Open fractures involving skull or face with other bones, with cerebral laceration and contusion, with no loss of consciousness | S02.91XB | Unspecified fracture of skull, initial encounter for open fracture |
| 804.61 | Open fractures involving skull or face with other bones, with cerebral laceration and contusion, with no loss of consciousness | S06.330A | Contusion and laceration of cerebrum, unspecified, without loss of consciousness, initial encounter |
| 804.62 | Open fractures involving skull or face with other bones, with cerebral laceration and contusion, with brief [less than one hour] loss of consciousness | S02.91XB | Unspecified fracture of skull, initial encounter for open fracture |
| 804.62 | Open fractures involving skull or face with other bones, with cerebral laceration and contusion, with brief [less than one hour] loss of consciousness | S06.331A | Contusion and laceration of cerebrum, unspecified, with loss of consciousness of 30 minutes or less, initial encounter |
| 804.62 | Open fractures involving skull or face with other bones, with cerebral laceration and contusion, with brief [less than one hour] loss of consciousness | S06.332A | Contusion and laceration of cerebrum, unspecified, with loss of consciousness of 31 minutes to 59 minutes, initial encounter |
| 804.63 | Open fractures involving skull or face with other bones, with cerebral laceration and contusion, with moderate [1-24 hours] loss of consciousness | S02.91XB | Unspecified fracture of skull, initial encounter for open fracture |
| 804.63 | Open fractures involving skull or face with other bones, with cerebral laceration and contusion, with moderate [1-24 hours] loss of consciousness | S06.333A | Contusion and laceration of cerebrum, unspecified, with loss of consciousness of 1 hour to 5 hours 59 minutes, initial encounter |
| 804.63 | Open fractures involving skull or face with other bones, with cerebral laceration and contusion, with moderate [1-24 hours] loss of consciousness | S06.334A | Contusion and laceration of cerebrum, unspecified, with loss of consciousness of 6 hours to 24 hours, initial encounter |
| 804.64 | Open fractures involving skull or face with other bones, with cerebral laceration and contusion, with prolonged [more than 24 hours] loss of consciousness and return to pre-existing conscious level | S02.91XB | Unspecified fracture of skull, initial encounter for open fracture |
| 804.64 | Open fractures involving skull or face with other bones, with cerebral laceration and contusion, with prolonged [more than 24 hours] loss of consciousness and return to pre-existing conscious level | S06.335A | Contusion and laceration of cerebrum, unspecified, with loss of consciousness greater than 24 hours with return to pre-existing conscious level, initial encounter |
| 804.65 | Open fractures involving skull or face with other bones, with cerebral laceration and contusion, with prolonged [more than 24 hours] loss of consciousness, without return to pre-existing conscious level | S02.91XB | Unspecified fracture of skull, initial encounter for open fracture |
| 804.65 | Open fractures involving skull or face with other bones, with cerebral laceration and contusion, with prolonged [more than 24 hours] loss of consciousness, without return to pre-existing conscious level | S06.336A | Contusion and laceration of cerebrum, unspecified, with loss of consciousness greater than 24 hours without return to pre-existing conscious level with patient surviving, initial encounter |
| 804.65 | Open fractures involving skull or face with other bones, with cerebral laceration and contusion, with prolonged [more than 24 hours] loss of consciousness, without return to pre-existing conscious level | S06.337A | Contusion and laceration of cerebrum, unspecified, with loss of consciousness of any duration with death due to brain injury prior to regaining consciousness, initial encounter |
| 804.65 | Open fractures involving skull or face with other bones, with cerebral laceration and contusion, with prolonged [more than 24 hours] loss of consciousness, without return to pre-existing conscious level | S06.338A | Contusion and laceration of cerebrum, unspecified, with loss of consciousness of any duration with death due to other cause prior to regaining consciousness, initial encounter |
| 804.66 | Open fractures involving skull or face with other bones, with cerebral laceration and contusion, with loss of consciousness of unspecified duration | S02.91XB | Unspecified fracture of skull, initial encounter for open fracture |
| 804.66 | Open fractures involving skull or face with other bones, with cerebral laceration and contusion, with loss of consciousness of unspecified duration | S06.339A | Contusion and laceration of cerebrum, unspecified, with loss of consciousness of unspecified duration, initial encounter |
| 804.69 | Open fractures involving skull or face with other bones, with cerebral laceration and contusion, with concussion, unspecified | S02.91XB | Unspecified fracture of skull, initial encounter for open fracture |
| 804.69 | Open fractures involving skull or face with other bones, with cerebral laceration and contusion, with concussion, unspecified | S06.330A | Contusion and laceration of cerebrum, unspecified, without loss of consciousness, initial encounter |
| 804.70 | Open fractures involving skull or face with other bones with subarachnoid, subdural, and extradural hemorrhage, unspecified state of consciousness | S06.4X0A | Epidural hemorrhage without loss of consciousness, initial encounter |
| 804.70 | Open fractures involving skull or face with other bones with subarachnoid, subdural, and extradural hemorrhage, unspecified state of consciousness | S06.5X0A | Traumatic subdural hemorrhage without loss of consciousness, initial encounter |
| 804.70 | Open fractures involving skull or face with other bones with subarachnoid, subdural, and extradural hemorrhage, unspecified state of consciousness | S06.6X0A | Traumatic subarachnoid hemorrhage without loss of consciousness, initial encounter |
| 804.70 | Open fractures involving skull or face with other bones with subarachnoid, subdural, and extradural hemorrhage, unspecified state of consciousness |  |  |
| 804.71 | Open fractures involving skull or face with other bones with subarachnoid, subdural, and extradural hemorrhage, with no loss of consciousness | S06.4X0A | Epidural hemorrhage without loss of consciousness, initial encounter |
| 804.71 | Open fractures involving skull or face with other bones with subarachnoid, subdural, and extradural hemorrhage, with no loss of consciousness | S06.5X0A | Traumatic subdural hemorrhage without loss of consciousness, initial encounter |
| 804.71 | Open fractures involving skull or face with other bones with subarachnoid, subdural, and extradural hemorrhage, with no loss of consciousness | S06.6X0A | Traumatic subarachnoid hemorrhage without loss of consciousness, initial encounter |
| 804.71 | Open fractures involving skull or face with other bones with subarachnoid, subdural, and extradural hemorrhage, with no loss of consciousness |  |  |
| 804.72 | Open fractures involving skull or face with other bones with subarachnoid, subdural, and extradural hemorrhage, with brief [less than one hour] loss of consciousness | S06.4X1A | Epidural hemorrhage with loss of consciousness of 30 minutes or less, initial encounter |
| 804.72 | Open fractures involving skull or face with other bones with subarachnoid, subdural, and extradural hemorrhage, with brief [less than one hour] loss of consciousness | S06.4X2A | Epidural hemorrhage with loss of consciousness of 31 minutes to 59 minutes, initial encounter |
| 804.72 | Open fractures involving skull or face with other bones with subarachnoid, subdural, and extradural hemorrhage, with brief [less than one hour] loss of consciousness | S06.5X1A | Traumatic subdural hemorrhage with loss of consciousness of 30 minutes or less, initial encounter |
| 804.72 | Open fractures involving skull or face with other bones with subarachnoid, subdural, and extradural hemorrhage, with brief [less than one hour] loss of consciousness | S06.5X2A | Traumatic subdural hemorrhage with loss of consciousness of 31 minutes to 59 minutes, initial encounter |
| 804.72 | Open fractures involving skull or face with other bones with subarachnoid, subdural, and extradural hemorrhage, with brief [less than one hour] loss of consciousness | S06.6X1A | Traumatic subarachnoid hemorrhage with loss of consciousness of 30 minutes or less, initial encounter |
| 804.72 | Open fractures involving skull or face with other bones with subarachnoid, subdural, and extradural hemorrhage, with brief [less than one hour] loss of consciousness | S06.6X2A | Traumatic subarachnoid hemorrhage with loss of consciousness of 31 minutes to 59 minutes, initial encounter |
| 804.72 | Open fractures involving skull or face with other bones with subarachnoid, subdural, and extradural hemorrhage, with brief [less than one hour] loss of consciousness |  |  |
| 804.73 | Open fractures involving skull or face with other bones with subarachnoid, subdural, and extradural hemorrhage, with moderate [1-24 hours] loss of consciousness | S06.4X3A | Epidural hemorrhage with loss of consciousness of 1 hour to 5 hours 59 minutes, initial encounter |
| 804.73 | Open fractures involving skull or face with other bones with subarachnoid, subdural, and extradural hemorrhage, with moderate [1-24 hours] loss of consciousness | S06.4X4A | Epidural hemorrhage with loss of consciousness of 6 hours to 24 hours, initial encounter |
| 804.73 | Open fractures involving skull or face with other bones with subarachnoid, subdural, and extradural hemorrhage, with moderate [1-24 hours] loss of consciousness | S06.5X3A | Traumatic subdural hemorrhage with loss of consciousness of 1 hour to 5 hours 59 minutes, initial encounter |
| 804.73 | Open fractures involving skull or face with other bones with subarachnoid, subdural, and extradural hemorrhage, with moderate [1-24 hours] loss of consciousness | S06.5X4A | Traumatic subdural hemorrhage with loss of consciousness of 6 hours to 24 hours, initial encounter |
| 804.73 | Open fractures involving skull or face with other bones with subarachnoid, subdural, and extradural hemorrhage, with moderate [1-24 hours] loss of consciousness | S06.6X3A | Traumatic subarachnoid hemorrhage with loss of consciousness of 1 hour to 5 hours 59 minutes, initial encounter |
| 804.73 | Open fractures involving skull or face with other bones with subarachnoid, subdural, and extradural hemorrhage, with moderate [1-24 hours] loss of consciousness | S06.6X4A | Traumatic subarachnoid hemorrhage with loss of consciousness of 6 hours to 24 hours, initial encounter |
| 804.73 | Open fractures involving skull or face with other bones with subarachnoid, subdural, and extradural hemorrhage, with moderate [1-24 hours] loss of consciousness |  |  |
| 804.74 | Open fractures involving skull or face with other bones with subarachnoid, subdural, and extradural hemorrhage, with prolonged [more than 24 hours] loss of consciousness and return to pre-existing conscious level | S06.4X5A | Epidural hemorrhage with loss of consciousness greater than 24 hours with return to pre-existing conscious level, initial encounter |
| 804.74 | Open fractures involving skull or face with other bones with subarachnoid, subdural, and extradural hemorrhage, with prolonged [more than 24 hours] loss of consciousness and return to pre-existing conscious level | S06.5X5A | Traumatic subdural hemorrhage with loss of consciousness greater than 24 hours with return to pre-existing conscious level, initial encounter |
| 804.74 | Open fractures involving skull or face with other bones with subarachnoid, subdural, and extradural hemorrhage, with prolonged [more than 24 hours] loss of consciousness and return to pre-existing conscious level | S06.6X5A | Traumatic subarachnoid hemorrhage with loss of consciousness greater than 24 hours with return to pre-existing conscious level, initial encounter |
| 804.74 | Open fractures involving skull or face with other bones with subarachnoid, subdural, and extradural hemorrhage, with prolonged [more than 24 hours] loss of consciousness and return to pre-existing conscious level |  |  |
| 804.75 | Open fractures involving skull or face with other bones with subarachnoid, subdural, and extradural hemorrhage, with prolonged [more than 24 hours] loss of consciousness, without return to pre-existing conscious level | S06.4X6A | Epidural hemorrhage with loss of consciousness greater than 24 hours without return to pre-existing conscious level with patient surviving, initial encounter |
| 804.75 | Open fractures involving skull or face with other bones with subarachnoid, subdural, and extradural hemorrhage, with prolonged [more than 24 hours] loss of consciousness, without return to pre-existing conscious level | S06.4X7A | Epidural hemorrhage with loss of consciousness of any duration with death due to brain injury prior to regaining consciousness, initial encounter |
| 804.75 | Open fractures involving skull or face with other bones with subarachnoid, subdural, and extradural hemorrhage, with prolonged [more than 24 hours] loss of consciousness, without return to pre-existing conscious level | S06.4X8A | Epidural hemorrhage with loss of consciousness of any duration with death due to other causes prior to regaining consciousness, initial encounter |
| 804.75 | Open fractures involving skull or face with other bones with subarachnoid, subdural, and extradural hemorrhage, with prolonged [more than 24 hours] loss of consciousness, without return to pre-existing conscious level | S06.5X6A | Traumatic subdural hemorrhage with loss of consciousness greater than 24 hours without return to pre-existing conscious level with patient surviving, initial encounter |
| 804.75 | Open fractures involving skull or face with other bones with subarachnoid, subdural, and extradural hemorrhage, with prolonged [more than 24 hours] loss of consciousness, without return to pre-existing conscious level | S06.5X7A | Traumatic subdural hemorrhage with loss of consciousness of any duration with death due to brain injury before regaining consciousness, initial encounter |
| 804.75 | Open fractures involving skull or face with other bones with subarachnoid, subdural, and extradural hemorrhage, with prolonged [more than 24 hours] loss of consciousness, without return to pre-existing conscious level | S06.5X8A | Traumatic subdural hemorrhage with loss of consciousness of any duration with death due to other cause before regaining consciousness, initial encounter |
| 804.75 | Open fractures involving skull or face with other bones with subarachnoid, subdural, and extradural hemorrhage, with prolonged [more than 24 hours] loss of consciousness, without return to pre-existing conscious level | S06.6X6A | Traumatic subarachnoid hemorrhage with loss of consciousness greater than 24 hours without return to pre-existing conscious level with patient surviving, initial encounter |
| 804.75 | Open fractures involving skull or face with other bones with subarachnoid, subdural, and extradural hemorrhage, with prolonged [more than 24 hours] loss of consciousness, without return to pre-existing conscious level | S06.6X7A | Traumatic subarachnoid hemorrhage with loss of consciousness of any duration with death due to brain injury prior to regaining consciousness, initial encounter |
| 804.75 | Open fractures involving skull or face with other bones with subarachnoid, subdural, and extradural hemorrhage, with prolonged [more than 24 hours] loss of consciousness, without return to pre-existing conscious level | S06.6X8A | Traumatic subarachnoid hemorrhage with loss of consciousness of any duration with death due to other cause prior to regaining consciousness, initial encounter |
| 804.75 | Open fractures involving skull or face with other bones with subarachnoid, subdural, and extradural hemorrhage, with prolonged [more than 24 hours] loss of consciousness, without return to pre-existing conscious level |  |  |
| 804.76 | Open fractures involving skull or face with other bones with subarachnoid, subdural, and extradural hemorrhage, with loss of consciousness of unspecified duration | S06.4X9A | Epidural hemorrhage with loss of consciousness of unspecified duration, initial encounter |
| 804.76 | Open fractures involving skull or face with other bones with subarachnoid, subdural, and extradural hemorrhage, with loss of consciousness of unspecified duration | S06.5X9A | Traumatic subdural hemorrhage with loss of consciousness of unspecified duration, initial encounter |
| 804.76 | Open fractures involving skull or face with other bones with subarachnoid, subdural, and extradural hemorrhage, with loss of consciousness of unspecified duration | S06.6X9A | Traumatic subarachnoid hemorrhage with loss of consciousness of unspecified duration, initial encounter |
| 804.76 | Open fractures involving skull or face with other bones with subarachnoid, subdural, and extradural hemorrhage, with loss of consciousness of unspecified duration |  |  |
| 804.79 | Open fractures involving skull or face with other bones with subarachnoid, subdural, and extradural hemorrhage, with concussion, unspecified | S06.4X0A | Epidural hemorrhage without loss of consciousness, initial encounter |
| 804.79 | Open fractures involving skull or face with other bones with subarachnoid, subdural, and extradural hemorrhage, with concussion, unspecified | S06.5X0A | Traumatic subdural hemorrhage without loss of consciousness, initial encounter |
| 804.79 | Open fractures involving skull or face with other bones with subarachnoid, subdural, and extradural hemorrhage, with concussion, unspecified | S06.6X0A | Traumatic subarachnoid hemorrhage without loss of consciousness, initial encounter |
| 804.79 | Open fractures involving skull or face with other bones with subarachnoid, subdural, and extradural hemorrhage, with concussion, unspecified |  |  |
| 804.80 | Open fractures involving skull or face with other bones, with other and unspecified intracranial hemorrhage, unspecified state of consciousness | S02.91XB | Unspecified fracture of skull, initial encounter for open fracture |
| 804.80 | Open fractures involving skull or face with other bones, with other and unspecified intracranial hemorrhage, unspecified state of consciousness | S06.360A | Traumatic hemorrhage of cerebrum, unspecified, without loss of consciousness, initial encounter |
| 804.81 | Open fractures involving skull or face with other bones, with other and unspecified intracranial hemorrhage, with no loss of consciousness | S02.91XB | Unspecified fracture of skull, initial encounter for open fracture |
| 804.81 | Open fractures involving skull or face with other bones, with other and unspecified intracranial hemorrhage, with no loss of consciousness | S06.360A | Traumatic hemorrhage of cerebrum, unspecified, without loss of consciousness, initial encounter |
| 804.82 | Open fractures involving skull or face with other bones, with other and unspecified intracranial hemorrhage, with brief [less than one hour] loss of consciousness | S02.91XB | Unspecified fracture of skull, initial encounter for open fracture |
| 804.82 | Open fractures involving skull or face with other bones, with other and unspecified intracranial hemorrhage, with brief [less than one hour] loss of consciousness | S06.361A | Traumatic hemorrhage of cerebrum, unspecified, with loss of consciousness of 30 minutes or less, initial encounter |
| 804.82 | Open fractures involving skull or face with other bones, with other and unspecified intracranial hemorrhage, with brief [less than one hour] loss of consciousness | S06.362A | Traumatic hemorrhage of cerebrum, unspecified, with loss of consciousness of 31 minutes to 59 minutes, initial encounter |
| 804.83 | Open fractures involving skull or face with other bones, with other and unspecified intracranial hemorrhage, with moderate [1-24 hours] loss of consciousness | S02.91XB | Unspecified fracture of skull, initial encounter for open fracture |
| 804.83 | Open fractures involving skull or face with other bones, with other and unspecified intracranial hemorrhage, with moderate [1-24 hours] loss of consciousness | S06.363A | Traumatic hemorrhage of cerebrum, unspecified, with loss of consciousness of 1 hours to 5 hours 59 minutes, initial encounter |
| 804.83 | Open fractures involving skull or face with other bones, with other and unspecified intracranial hemorrhage, with moderate [1-24 hours] loss of consciousness | S06.364A | Traumatic hemorrhage of cerebrum, unspecified, with loss of consciousness of 6 hours to 24 hours, initial encounter |
| 804.84 | Open fractures involving skull or face with other bones, with other and unspecified intracranial hemorrhage, with prolonged [more than 24 hours] loss of consciousness and return to pre-existing conscious level | S02.91XB | Unspecified fracture of skull, initial encounter for open fracture |
| 804.84 | Open fractures involving skull or face with other bones, with other and unspecified intracranial hemorrhage, with prolonged [more than 24 hours] loss of consciousness and return to pre-existing conscious level | S06.365A | Traumatic hemorrhage of cerebrum, unspecified, with loss of consciousness greater than 24 hours with return to pre-existing conscious level, initial encounter |
| 804.85 | Open fractures involving skull or face with other bones, with other and unspecified intracranial hemorrhage, with prolonged [more than 24 hours] loss consciousness, without return to pre-existing conscious level | S02.91XB | Unspecified fracture of skull, initial encounter for open fracture |
| 804.85 | Open fractures involving skull or face with other bones, with other and unspecified intracranial hemorrhage, with prolonged [more than 24 hours] loss consciousness, without return to pre-existing conscious level | S06.366A | Traumatic hemorrhage of cerebrum, unspecified, with loss of consciousness greater than 24 hours without return to pre-existing conscious level with patient surviving, initial encounter |
| 804.85 | Open fractures involving skull or face with other bones, with other and unspecified intracranial hemorrhage, with prolonged [more than 24 hours] loss consciousness, without return to pre-existing conscious level | S06.367A | Traumatic hemorrhage of cerebrum, unspecified, with loss of consciousness of any duration with death due to brain injury prior to regaining consciousness, initial encounter |
| 804.85 | Open fractures involving skull or face with other bones, with other and unspecified intracranial hemorrhage, with prolonged [more than 24 hours] loss consciousness, without return to pre-existing conscious level | S06.368A | Traumatic hemorrhage of cerebrum, unspecified, with loss of consciousness of any duration with death due to other cause prior to regaining consciousness, initial encounter |
| 804.86 | Open fractures involving skull or face with other bones, with other and unspecified intracranial hemorrhage, with loss of consciousness of unspecified duration | S02.91XB | Unspecified fracture of skull, initial encounter for open fracture |
| 804.86 | Open fractures involving skull or face with other bones, with other and unspecified intracranial hemorrhage, with loss of consciousness of unspecified duration | S06.369A | Traumatic hemorrhage of cerebrum, unspecified, with loss of consciousness of unspecified duration, initial encounter |
| 804.89 | Open fractures involving skull or face with other bones, with other and unspecified intracranial hemorrhage, with concussion, unspecified | S02.91XB | Unspecified fracture of skull, initial encounter for open fracture |
| 804.89 | Open fractures involving skull or face with other bones, with other and unspecified intracranial hemorrhage, with concussion, unspecified | S06.360A | Traumatic hemorrhage of cerebrum, unspecified, without loss of consciousness, initial encounter |
| 804.90 | Open fractures involving skull or face with other bones, with intracranial injury of other and unspecified nature, unspecified state of consciousness | S02.91XB | Unspecified fracture of skull, initial encounter for open fracture |
| 804.90 | Open fractures involving skull or face with other bones, with intracranial injury of other and unspecified nature, unspecified state of consciousness | S06.890A | Other specified intracranial injury without loss of consciousness, initial encounter |
| 804.90 | Open fractures involving skull or face with other bones, with intracranial injury of other and unspecified nature, unspecified state of consciousness | S06.9X0A | Unspecified intracranial injury without loss of consciousness, initial encounter |
| 804.91 | Open fractures involving skull or face with other bones, with intracranial injury of other and unspecified nature, with no loss of consciousness | S02.91XB | Unspecified fracture of skull, initial encounter for open fracture |
| 804.91 | Open fractures involving skull or face with other bones, with intracranial injury of other and unspecified nature, with no loss of consciousness | S06.890A | Other specified intracranial injury without loss of consciousness, initial encounter |
| 804.91 | Open fractures involving skull or face with other bones, with intracranial injury of other and unspecified nature, with no loss of consciousness | S06.9X0A | Unspecified intracranial injury without loss of consciousness, initial encounter |
| 804.92 | Open fractures involving skull or face with other bones, with intracranial injury of other and unspecified nature, with brief [less than one hour] loss of consciousness | S02.91XB | Unspecified fracture of skull, initial encounter for open fracture |
| 804.92 | Open fractures involving skull or face with other bones, with intracranial injury of other and unspecified nature, with brief [less than one hour] loss of consciousness | S06.891A | Other specified intracranial injury with loss of consciousness of 30 minutes or less, initial encounter |
| 804.92 | Open fractures involving skull or face with other bones, with intracranial injury of other and unspecified nature, with brief [less than one hour] loss of consciousness | S06.892A | Other specified intracranial injury with loss of consciousness of 31 minutes to 59 minutes, initial encounter |
| 804.92 | Open fractures involving skull or face with other bones, with intracranial injury of other and unspecified nature, with brief [less than one hour] loss of consciousness | S06.9X1A | Unspecified intracranial injury with loss of consciousness of 30 minutes or less, initial encounter |
| 804.92 | Open fractures involving skull or face with other bones, with intracranial injury of other and unspecified nature, with brief [less than one hour] loss of consciousness | S06.9X2A | Unspecified intracranial injury with loss of consciousness of 31 minutes to 59 minutes, initial encounter |
| 804.93 | Open fractures involving skull or face with other bones, with intracranial injury of other and unspecified nature, with moderate [1-24 hours] loss of consciousness | S02.91XB | Unspecified fracture of skull, initial encounter for open fracture |
| 804.93 | Open fractures involving skull or face with other bones, with intracranial injury of other and unspecified nature, with moderate [1-24 hours] loss of consciousness | S06.893A | Other specified intracranial injury with loss of consciousness of 1 hour to 5 hours 59 minutes, initial encounter |
| 804.93 | Open fractures involving skull or face with other bones, with intracranial injury of other and unspecified nature, with moderate [1-24 hours] loss of consciousness | S06.894A | Other specified intracranial injury with loss of consciousness of 6 hours to 24 hours, initial encounter |
| 804.93 | Open fractures involving skull or face with other bones, with intracranial injury of other and unspecified nature, with moderate [1-24 hours] loss of consciousness | S06.9X4A | Unspecified intracranial injury with loss of consciousness of 6 hours to 24 hours, initial encounter |
| 804.94 | Open fractures involving skull or face with other bones, with intracranial injury of other and unspecified nature, with prolonged [more than 24 hours] loss of consciousness and return to pre-existing conscious level | S02.91XB | Unspecified fracture of skull, initial encounter for open fracture |
| 804.94 | Open fractures involving skull or face with other bones, with intracranial injury of other and unspecified nature, with prolonged [more than 24 hours] loss of consciousness and return to pre-existing conscious level | S06.895A | Other specified intracranial injury with loss of consciousness greater than 24 hours with return to pre-existing conscious level, initial encounter |
| 804.94 | Open fractures involving skull or face with other bones, with intracranial injury of other and unspecified nature, with prolonged [more than 24 hours] loss of consciousness and return to pre-existing conscious level | S06.9X5A | Unspecified intracranial injury with loss of consciousness greater than 24 hours with return to pre-existing conscious level, initial encounter |
| 804.95 | Open fractures involving skull or face with other bones, with intracranial injury of other and unspecified nature, with prolonged [more than 24 hours] loss of consciousness without return to pre-existing conscious level | S02.91XB | Unspecified fracture of skull, initial encounter for open fracture |
| 804.95 | Open fractures involving skull or face with other bones, with intracranial injury of other and unspecified nature, with prolonged [more than 24 hours] loss of consciousness without return to pre-existing conscious level | S06.896A | Other specified intracranial injury with loss of consciousness greater than 24 hours without return to pre-existing conscious level with patient surviving, initial encounter |
| 804.95 | Open fractures involving skull or face with other bones, with intracranial injury of other and unspecified nature, with prolonged [more than 24 hours] loss of consciousness without return to pre-existing conscious level | S06.897A | Other specified intracranial injury with loss of consciousness of any duration with death due to brain injury prior to regaining consciousness, initial encounter |
| 804.95 | Open fractures involving skull or face with other bones, with intracranial injury of other and unspecified nature, with prolonged [more than 24 hours] loss of consciousness without return to pre-existing conscious level | S06.898A | Other specified intracranial injury with loss of consciousness of any duration with death due to other cause prior to regaining consciousness, initial encounter |
| 804.95 | Open fractures involving skull or face with other bones, with intracranial injury of other and unspecified nature, with prolonged [more than 24 hours] loss of consciousness without return to pre-existing conscious level | S06.9X6A | Unspecified intracranial injury with loss of consciousness greater than 24 hours without return to pre-existing conscious level with patient surviving, initial encounter |
| 804.95 | Open fractures involving skull or face with other bones, with intracranial injury of other and unspecified nature, with prolonged [more than 24 hours] loss of consciousness without return to pre-existing conscious level | S06.9X7A | Unspecified intracranial injury with loss of consciousness of any duration with death due to brain injury prior to regaining consciousness, initial encounter |
| 804.95 | Open fractures involving skull or face with other bones, with intracranial injury of other and unspecified nature, with prolonged [more than 24 hours] loss of consciousness without return to pre-existing conscious level | S06.9X8A | Unspecified intracranial injury with loss of consciousness of any duration with death due to other cause prior to regaining consciousness, initial encounter |
| 804.96 | Open fractures involving skull or face with other bones, with intracranial injury of other and unspecified nature, with loss of consciousness of unspecified duration | S02.91XB | Unspecified fracture of skull, initial encounter for open fracture |
| 804.96 | Open fractures involving skull or face with other bones, with intracranial injury of other and unspecified nature, with loss of consciousness of unspecified duration | S06.899A | Other specified intracranial injury with loss of consciousness of unspecified duration, initial encounter |
| 804.96 | Open fractures involving skull or face with other bones, with intracranial injury of other and unspecified nature, with loss of consciousness of unspecified duration | S06.9X9A | Unspecified intracranial injury with loss of consciousness of unspecified duration, initial encounter |
| 804.99 | Open fractures involving skull or face with other bones, with intracranial injury of other and unspecified nature, with concussion, unspecified | S02.91XB | Unspecified fracture of skull, initial encounter for open fracture |
| 804.99 | Open fractures involving skull or face with other bones, with intracranial injury of other and unspecified nature, with concussion, unspecified | S06.890A | Other specified intracranial injury without loss of consciousness, initial encounter |
| 804.99 | Open fractures involving skull or face with other bones, with intracranial injury of other and unspecified nature, with concussion, unspecified | S06.9X0A | Unspecified intracranial injury without loss of consciousness, initial encounter |
| 851.00 | Cortex (cerebral) contusion without mention of open intracranial wound, unspecified state of consciousness | S06.330A | Contusion and laceration of cerebrum, unspecified, without loss of consciousness, initial encounter |
| 851.01 | Cortex (cerebral) contusion without mention of open intracranial wound, with no loss of consciousness | S06.330A | Contusion and laceration of cerebrum, unspecified, without loss of consciousness, initial encounter |
| 851.02 | Cortex (cerebral) contusion without mention of open intracranial wound, with brief [less than one hour] loss of consciousness | S06.331A | Contusion and laceration of cerebrum, unspecified, with loss of consciousness of 30 minutes or less, initial encounter |
| 851.03 | Cortex (cerebral) contusion without mention of open intracranial wound, with moderate [1-24 hours] loss of consciousness | S06.333A | Contusion and laceration of cerebrum, unspecified, with loss of consciousness of 1 hour to 5 hours 59 minutes, initial encounter |
| 851.04 | Cortex (cerebral) contusion without mention of open intracranial wound, with prolonged [more than 24 hours] loss of consciousness and return to pre-existing conscious level | S06.335A | Contusion and laceration of cerebrum, unspecified, with loss of consciousness greater than 24 hours with return to pre-existing conscious level, initial encounter |
| 851.05 | Cortex (cerebral) contusion without mention of open intracranial wound, with prolonged [more than 24 hours] loss of consciousness without return to pre-existing conscious level | S06.336A | Contusion and laceration of cerebrum, unspecified, with loss of consciousness greater than 24 hours without return to pre-existing conscious level with patient surviving, initial encounter |
| 851.06 | Cortex (cerebral) contusion without mention of open intracranial wound, with loss of consciousness of unspecified duration | S06.339A | Contusion and laceration of cerebrum, unspecified, with loss of consciousness of unspecified duration, initial encounter |
| 851.09 | Cortex (cerebral) contusion without mention of open intracranial wound, with concussion, unspecified | S06.330A | Contusion and laceration of cerebrum, unspecified, without loss of consciousness, initial encounter |
| 851.10 | Cortex (cerebral) contusion with open intracranial wound, unspecified state of consciousness | S06.330A | Contusion and laceration of cerebrum, unspecified, without loss of consciousness, initial encounter |
| 851.10 | Cortex (cerebral) contusion with open intracranial wound, unspecified state of consciousness |  |  |
| 851.11 | Cortex (cerebral) contusion with open intracranial wound, with no loss of consciousness | S06.330A | Contusion and laceration of cerebrum, unspecified, without loss of consciousness, initial encounter |
| 851.11 | Cortex (cerebral) contusion with open intracranial wound, with no loss of consciousness |  |  |
| 851.12 | Cortex (cerebral) contusion with open intracranial wound, with brief [less than one hour] loss of consciousness | S06.331A | Contusion and laceration of cerebrum, unspecified, with loss of consciousness of 30 minutes or less, initial encounter |
| 851.12 | Cortex (cerebral) contusion with open intracranial wound, with brief [less than one hour] loss of consciousness | S06.332A | Contusion and laceration of cerebrum, unspecified, with loss of consciousness of 31 minutes to 59 minutes, initial encounter |
| 851.12 | Cortex (cerebral) contusion with open intracranial wound, with brief [less than one hour] loss of consciousness |  |  |
| 851.13 | Cortex (cerebral) contusion with open intracranial wound, with moderate [1-24 hours] loss of consciousness | S06.333A | Contusion and laceration of cerebrum, unspecified, with loss of consciousness of 1 hour to 5 hours 59 minutes, initial encounter |
| 851.13 | Cortex (cerebral) contusion with open intracranial wound, with moderate [1-24 hours] loss of consciousness | S06.334A | Contusion and laceration of cerebrum, unspecified, with loss of consciousness of 6 hours to 24 hours, initial encounter |
| 851.13 | Cortex (cerebral) contusion with open intracranial wound, with moderate [1-24 hours] loss of consciousness |  |  |
| 851.14 | Cortex (cerebral) contusion with open intracranial wound, with prolonged [more than 24 hours] loss of consciousness and return to pre-existing conscious level | S06.335A | Contusion and laceration of cerebrum, unspecified, with loss of consciousness greater than 24 hours with return to pre-existing conscious level, initial encounter |
| 851.14 | Cortex (cerebral) contusion with open intracranial wound, with prolonged [more than 24 hours] loss of consciousness and return to pre-existing conscious level |  |  |
| 851.15 | Cortex (cerebral) contusion with open intracranial wound, with prolonged [more than 24 hours] loss of consciousness without return to pre-existing conscious level | S06.336A | Contusion and laceration of cerebrum, unspecified, with loss of consciousness greater than 24 hours without return to pre-existing conscious level with patient surviving, initial encounter |
| 851.15 | Cortex (cerebral) contusion with open intracranial wound, with prolonged [more than 24 hours] loss of consciousness without return to pre-existing conscious level | S06.337A | Contusion and laceration of cerebrum, unspecified, with loss of consciousness of any duration with death due to brain injury prior to regaining consciousness, initial encounter |
| 851.15 | Cortex (cerebral) contusion with open intracranial wound, with prolonged [more than 24 hours] loss of consciousness without return to pre-existing conscious level | S06.338A | Contusion and laceration of cerebrum, unspecified, with loss of consciousness of any duration with death due to other cause prior to regaining consciousness, initial encounter |
| 851.15 | Cortex (cerebral) contusion with open intracranial wound, with prolonged [more than 24 hours] loss of consciousness without return to pre-existing conscious level |  |  |
| 851.16 | Cortex (cerebral) contusion with open intracranial wound, with loss of consciousness of unspecified duration | S06.339A | Contusion and laceration of cerebrum, unspecified, with loss of consciousness of unspecified duration, initial encounter |
| 851.16 | Cortex (cerebral) contusion with open intracranial wound, with loss of consciousness of unspecified duration |  |  |
| 851.19 | Cortex (cerebral) contusion with open intracranial wound, with concussion, unspecified | S06.330A | Contusion and laceration of cerebrum, unspecified, without loss of consciousness, initial encounter |
| 851.19 | Cortex (cerebral) contusion with open intracranial wound, with concussion, unspecified | S06.339A | Contusion and laceration of cerebrum, unspecified, with loss of consciousness of unspecified duration, initial encounter |
| 851.19 | Cortex (cerebral) contusion with open intracranial wound, with concussion, unspecified |  |  |
| 851.20 | Cortex (cerebral) laceration without mention of open intracranial wound, unspecified state of consciousness | S06.330A | Contusion and laceration of cerebrum, unspecified, without loss of consciousness, initial encounter |
| 851.21 | Cortex (cerebral) laceration without mention of open intracranial wound, with no loss of consciousness | S06.330A | Contusion and laceration of cerebrum, unspecified, without loss of consciousness, initial encounter |
| 851.22 | Cortex (cerebral) laceration without mention of open intracranial wound, with brief [less than one hour] loss of consciousness | S06.331A | Contusion and laceration of cerebrum, unspecified, with loss of consciousness of 30 minutes or less, initial encounter |
| 851.23 | Cortex (cerebral) laceration without mention of open intracranial wound, with moderate [1-24 hours] loss of consciousness | S06.333A | Contusion and laceration of cerebrum, unspecified, with loss of consciousness of 1 hour to 5 hours 59 minutes, initial encounter |
| 851.24 | Cortex (cerebral) laceration without mention of open intracranial wound, with prolonged [more than 24 hours] loss of consciousness and return to pre-existing conscious level | S06.335A | Contusion and laceration of cerebrum, unspecified, with loss of consciousness greater than 24 hours with return to pre-existing conscious level, initial encounter |
| 851.25 | Cortex (cerebral) laceration without mention of open intracranial wound, with prolonged [more than 24 hours] loss of consciousness without return to pre-existing conscious level | S06.336A | Contusion and laceration of cerebrum, unspecified, with loss of consciousness greater than 24 hours without return to pre-existing conscious level with patient surviving, initial encounter |
| 851.26 | Cortex (cerebral) laceration without mention of open intracranial wound, with loss of consciousness of unspecified duration | S06.339A | Contusion and laceration of cerebrum, unspecified, with loss of consciousness of unspecified duration, initial encounter |
| 851.29 | Cortex (cerebral) laceration without mention of open intracranial wound, with concussion, unspecified | S06.330A | Contusion and laceration of cerebrum, unspecified, without loss of consciousness, initial encounter |
| 851.29 | Cortex (cerebral) laceration without mention of open intracranial wound, with concussion, unspecified | S06.339A | Contusion and laceration of cerebrum, unspecified, with loss of consciousness of unspecified duration, initial encounter |
| 851.30 | Cortex (cerebral) laceration with open intracranial wound, unspecified state of consciousness | S06.330A | Contusion and laceration of cerebrum, unspecified, without loss of consciousness, initial encounter |
| 851.30 | Cortex (cerebral) laceration with open intracranial wound, unspecified state of consciousness |  |  |
| 851.31 | Cortex (cerebral) laceration with open intracranial wound, with no loss of consciousness | S06.330A | Contusion and laceration of cerebrum, unspecified, without loss of consciousness, initial encounter |
| 851.31 | Cortex (cerebral) laceration with open intracranial wound, with no loss of consciousness |  |  |
| 851.32 | Cortex (cerebral) laceration with open intracranial wound, with brief [less than one hour] loss of consciousness | S06.331A | Contusion and laceration of cerebrum, unspecified, with loss of consciousness of 30 minutes or less, initial encounter |
| 851.32 | Cortex (cerebral) laceration with open intracranial wound, with brief [less than one hour] loss of consciousness | S06.332A | Contusion and laceration of cerebrum, unspecified, with loss of consciousness of 31 minutes to 59 minutes, initial encounter |
| 851.32 | Cortex (cerebral) laceration with open intracranial wound, with brief [less than one hour] loss of consciousness |  |  |
| 851.33 | Cortex (cerebral) laceration with open intracranial wound, with moderate [1-24 hours] loss of consciousness | S06.333A | Contusion and laceration of cerebrum, unspecified, with loss of consciousness of 1 hour to 5 hours 59 minutes, initial encounter |
| 851.33 | Cortex (cerebral) laceration with open intracranial wound, with moderate [1-24 hours] loss of consciousness | S06.334A | Contusion and laceration of cerebrum, unspecified, with loss of consciousness of 6 hours to 24 hours, initial encounter |
| 851.33 | Cortex (cerebral) laceration with open intracranial wound, with moderate [1-24 hours] loss of consciousness |  |  |
| 851.34 | Cortex (cerebral) laceration with open intracranial wound, with prolonged [more than 24 hours] loss of consciousness and return to pre-existing conscious level | S06.335A | Contusion and laceration of cerebrum, unspecified, with loss of consciousness greater than 24 hours with return to pre-existing conscious level, initial encounter |
| 851.34 | Cortex (cerebral) laceration with open intracranial wound, with prolonged [more than 24 hours] loss of consciousness and return to pre-existing conscious level |  |  |
| 851.35 | Cortex (cerebral) laceration with open intracranial wound, with prolonged [more than 24 hours] loss of consciousness without return to pre-existing conscious level | S06.336A | Contusion and laceration of cerebrum, unspecified, with loss of consciousness greater than 24 hours without return to pre-existing conscious level with patient surviving, initial encounter |
| 851.35 | Cortex (cerebral) laceration with open intracranial wound, with prolonged [more than 24 hours] loss of consciousness without return to pre-existing conscious level | S06.337A | Contusion and laceration of cerebrum, unspecified, with loss of consciousness of any duration with death due to brain injury prior to regaining consciousness, initial encounter |
| 851.35 | Cortex (cerebral) laceration with open intracranial wound, with prolonged [more than 24 hours] loss of consciousness without return to pre-existing conscious level | S06.338A | Contusion and laceration of cerebrum, unspecified, with loss of consciousness of any duration with death due to other cause prior to regaining consciousness, initial encounter |
| 851.35 | Cortex (cerebral) laceration with open intracranial wound, with prolonged [more than 24 hours] loss of consciousness without return to pre-existing conscious level |  |  |
| 851.36 | Cortex (cerebral) laceration with open intracranial wound, with loss of consciousness of unspecified duration | S06.339A | Contusion and laceration of cerebrum, unspecified, with loss of consciousness of unspecified duration, initial encounter |
| 851.36 | Cortex (cerebral) laceration with open intracranial wound, with loss of consciousness of unspecified duration |  |  |
| 851.39 | Cortex (cerebral) laceration with open intracranial wound, with concussion, unspecified | S06.330A | Contusion and laceration of cerebrum, unspecified, without loss of consciousness, initial encounter |
| 851.39 | Cortex (cerebral) laceration with open intracranial wound, with concussion, unspecified |  |  |
| 851.40 | Cerebellar or brain stem contusion without mention of open intracranial wound, unspecified state of consciousness | S06.370A | Contusion, laceration, and hemorrhage of cerebellum without loss of consciousness, initial encounter |
| 851.40 | Cerebellar or brain stem contusion without mention of open intracranial wound, unspecified state of consciousness | S06.380A | Contusion, laceration, and hemorrhage of brainstem without loss of consciousness, initial encounter |
| 851.41 | Cerebellar or brain stem contusion without mention of open intracranial wound, with no loss of consciousness | S06.370A | Contusion, laceration, and hemorrhage of cerebellum without loss of consciousness, initial encounter |
| 851.41 | Cerebellar or brain stem contusion without mention of open intracranial wound, with no loss of consciousness | S06.380A | Contusion, laceration, and hemorrhage of brainstem without loss of consciousness, initial encounter |
| 851.42 | Cerebellar or brain stem contusion without mention of open intracranial wound, with brief [less than one hour] loss of consciousness | S06.371A | Contusion, laceration, and hemorrhage of cerebellum with loss of consciousness of 30 minutes or less, initial encounter |
| 851.42 | Cerebellar or brain stem contusion without mention of open intracranial wound, with brief [less than one hour] loss of consciousness | S06.372A | Contusion, laceration, and hemorrhage of cerebellum with loss of consciousness of 31 minutes to 59 minutes, initial encounter |
| 851.42 | Cerebellar or brain stem contusion without mention of open intracranial wound, with brief [less than one hour] loss of consciousness | S06.381A | Contusion, laceration, and hemorrhage of brainstem with loss of consciousness of 30 minutes or less, initial encounter |
| 851.42 | Cerebellar or brain stem contusion without mention of open intracranial wound, with brief [less than one hour] loss of consciousness | S06.382A | Contusion, laceration, and hemorrhage of brainstem with loss of consciousness of 31 minutes to 59 minutes, initial encounter |
| 851.43 | Cerebellar or brain stem contusion without mention of open intracranial wound, with moderate [1-24 hours] loss of consciousness | S06.373A | Contusion, laceration, and hemorrhage of cerebellum with loss of consciousness of 1 hour to 5 hours 59 minutes, initial encounter |
| 851.43 | Cerebellar or brain stem contusion without mention of open intracranial wound, with moderate [1-24 hours] loss of consciousness | S06.374A | Contusion, laceration, and hemorrhage of cerebellum with loss of consciousness of 6 hours to 24 hours, initial encounter |
| 851.43 | Cerebellar or brain stem contusion without mention of open intracranial wound, with moderate [1-24 hours] loss of consciousness | S06.383A | Contusion, laceration, and hemorrhage of brainstem with loss of consciousness of 1 hour to 5 hours 59 minutes, initial encounter |
| 851.43 | Cerebellar or brain stem contusion without mention of open intracranial wound, with moderate [1-24 hours] loss of consciousness | S06.384A | Contusion, laceration, and hemorrhage of brainstem with loss of consciousness of 6 hours to 24 hours, initial encounter |
| 851.44 | Cerebellar or brain stem contusion without mention of open intracranial wound, with prolonged [more than 24 hours] loss consciousness and return to pre-existing conscious level | S06.375A | Contusion, laceration, and hemorrhage of cerebellum with loss of consciousness greater than 24 hours with return to pre-existing conscious level, initial encounter |
| 851.44 | Cerebellar or brain stem contusion without mention of open intracranial wound, with prolonged [more than 24 hours] loss consciousness and return to pre-existing conscious level | S06.385A | Contusion, laceration, and hemorrhage of brainstem with loss of consciousness greater than 24 hours with return to pre-existing conscious level, initial encounter |
| 851.45 | Cerebellar or brain stem contusion without mention of open intracranial wound, with prolonged [more than 24 hours] loss of consciousness without return to pre-existing conscious level | S06.376A | Contusion, laceration, and hemorrhage of cerebellum with loss of consciousness greater than 24 hours without return to pre-existing conscious level with patient surviving, initial encounter |
| 851.45 | Cerebellar or brain stem contusion without mention of open intracranial wound, with prolonged [more than 24 hours] loss of consciousness without return to pre-existing conscious level | S06.377A | Contusion, laceration, and hemorrhage of cerebellum with loss of consciousness of any duration with death due to brain injury prior to regaining consciousness, initial encounter |
| 851.45 | Cerebellar or brain stem contusion without mention of open intracranial wound, with prolonged [more than 24 hours] loss of consciousness without return to pre-existing conscious level | S06.378A | Contusion, laceration, and hemorrhage of cerebellum with loss of consciousness of any duration with death due to other cause prior to regaining consciousness, initial encounter |
| 851.45 | Cerebellar or brain stem contusion without mention of open intracranial wound, with prolonged [more than 24 hours] loss of consciousness without return to pre-existing conscious level | S06.386A | Contusion, laceration, and hemorrhage of brainstem with loss of consciousness greater than 24 hours without return to pre-existing conscious level with patient surviving, initial encounter |
| 851.45 | Cerebellar or brain stem contusion without mention of open intracranial wound, with prolonged [more than 24 hours] loss of consciousness without return to pre-existing conscious level | S06.387A | Contusion, laceration, and hemorrhage of brainstem with loss of consciousness of any duration with death due to brain injury prior to regaining consciousness, initial encounter |
| 851.45 | Cerebellar or brain stem contusion without mention of open intracranial wound, with prolonged [more than 24 hours] loss of consciousness without return to pre-existing conscious level | S06.388A | Contusion, laceration, and hemorrhage of brainstem with loss of consciousness of any duration with death due to other cause prior to regaining consciousness, initial encounter |
| 851.46 | Cerebellar or brain stem contusion without mention of open intracranial wound, with loss of consciousness of unspecified duration | S06.379A | Contusion, laceration, and hemorrhage of cerebellum with loss of consciousness of unspecified duration, initial encounter |
| 851.46 | Cerebellar or brain stem contusion without mention of open intracranial wound, with loss of consciousness of unspecified duration | S06.389A | Contusion, laceration, and hemorrhage of brainstem with loss of consciousness of unspecified duration, initial encounter |
| 851.49 | Cerebellar or brain stem contusion without mention of open intracranial wound, with concussion, unspecified | S06.370A | Contusion, laceration, and hemorrhage of cerebellum without loss of consciousness, initial encounter |
| 851.49 | Cerebellar or brain stem contusion without mention of open intracranial wound, with concussion, unspecified | S06.380A | Contusion, laceration, and hemorrhage of brainstem without loss of consciousness, initial encounter |
| 851.50 | Cerebellar or brain stem contusion with open intracranial wound, unspecified state of consciousness | S06.370A | Contusion, laceration, and hemorrhage of cerebellum without loss of consciousness, initial encounter |
| 851.50 | Cerebellar or brain stem contusion with open intracranial wound, unspecified state of consciousness | S06.380A | Contusion, laceration, and hemorrhage of brainstem without loss of consciousness, initial encounter |
| 851.50 | Cerebellar or brain stem contusion with open intracranial wound, unspecified state of consciousness |  |  |
| 851.51 | Cerebellar or brain stem contusion with open intracranial wound, with no loss of consciousness | S06.370A | Contusion, laceration, and hemorrhage of cerebellum without loss of consciousness, initial encounter |
| 851.51 | Cerebellar or brain stem contusion with open intracranial wound, with no loss of consciousness | S06.380A | Contusion, laceration, and hemorrhage of brainstem without loss of consciousness, initial encounter |
| 851.51 | Cerebellar or brain stem contusion with open intracranial wound, with no loss of consciousness |  |  |
| 851.52 | Cerebellar or brain stem contusion with open intracranial wound, with brief [less than one hour] loss of consciousness | S06.371A | Contusion, laceration, and hemorrhage of cerebellum with loss of consciousness of 30 minutes or less, initial encounter |
| 851.52 | Cerebellar or brain stem contusion with open intracranial wound, with brief [less than one hour] loss of consciousness | S06.372A | Contusion, laceration, and hemorrhage of cerebellum with loss of consciousness of 31 minutes to 59 minutes, initial encounter |
| 851.52 | Cerebellar or brain stem contusion with open intracranial wound, with brief [less than one hour] loss of consciousness | S06.381A | Contusion, laceration, and hemorrhage of brainstem with loss of consciousness of 30 minutes or less, initial encounter |
| 851.52 | Cerebellar or brain stem contusion with open intracranial wound, with brief [less than one hour] loss of consciousness | S06.382A | Contusion, laceration, and hemorrhage of brainstem with loss of consciousness of 31 minutes to 59 minutes, initial encounter |
| 851.52 | Cerebellar or brain stem contusion with open intracranial wound, with brief [less than one hour] loss of consciousness |  |  |
| 851.53 | Cerebellar or brain stem contusion with open intracranial wound, with moderate [1-24 hours] loss of consciousness | S06.373A | Contusion, laceration, and hemorrhage of cerebellum with loss of consciousness of 1 hour to 5 hours 59 minutes, initial encounter |
| 851.53 | Cerebellar or brain stem contusion with open intracranial wound, with moderate [1-24 hours] loss of consciousness | S06.374A | Contusion, laceration, and hemorrhage of cerebellum with loss of consciousness of 6 hours to 24 hours, initial encounter |
| 851.53 | Cerebellar or brain stem contusion with open intracranial wound, with moderate [1-24 hours] loss of consciousness | S06.383A | Contusion, laceration, and hemorrhage of brainstem with loss of consciousness of 1 hour to 5 hours 59 minutes, initial encounter |
| 851.53 | Cerebellar or brain stem contusion with open intracranial wound, with moderate [1-24 hours] loss of consciousness | S06.384A | Contusion, laceration, and hemorrhage of brainstem with loss of consciousness of 6 hours to 24 hours, initial encounter |
| 851.53 | Cerebellar or brain stem contusion with open intracranial wound, with moderate [1-24 hours] loss of consciousness |  |  |
| 851.54 | Cerebellar or brain stem contusion with open intracranial wound, with prolonged [more than 24 hours] loss of consciousness and return to pre-existing conscious level | S06.375A | Contusion, laceration, and hemorrhage of cerebellum with loss of consciousness greater than 24 hours with return to pre-existing conscious level, initial encounter |
| 851.54 | Cerebellar or brain stem contusion with open intracranial wound, with prolonged [more than 24 hours] loss of consciousness and return to pre-existing conscious level | S06.385A | Contusion, laceration, and hemorrhage of brainstem with loss of consciousness greater than 24 hours with return to pre-existing conscious level, initial encounter |
| 851.54 | Cerebellar or brain stem contusion with open intracranial wound, with prolonged [more than 24 hours] loss of consciousness and return to pre-existing conscious level |  |  |
| 851.55 | Cerebellar or brain stem contusion with open intracranial wound, with prolonged [more than 24 hours] loss of consciousness without return to pre-existing conscious level | S06.376A | Contusion, laceration, and hemorrhage of cerebellum with loss of consciousness greater than 24 hours without return to pre-existing conscious level with patient surviving, initial encounter |
| 851.55 | Cerebellar or brain stem contusion with open intracranial wound, with prolonged [more than 24 hours] loss of consciousness without return to pre-existing conscious level | S06.377A | Contusion, laceration, and hemorrhage of cerebellum with loss of consciousness of any duration with death due to brain injury prior to regaining consciousness, initial encounter |
| 851.55 | Cerebellar or brain stem contusion with open intracranial wound, with prolonged [more than 24 hours] loss of consciousness without return to pre-existing conscious level | S06.378A | Contusion, laceration, and hemorrhage of cerebellum with loss of consciousness of any duration with death due to other cause prior to regaining consciousness, initial encounter |
| 851.55 | Cerebellar or brain stem contusion with open intracranial wound, with prolonged [more than 24 hours] loss of consciousness without return to pre-existing conscious level | S06.386A | Contusion, laceration, and hemorrhage of brainstem with loss of consciousness greater than 24 hours without return to pre-existing conscious level with patient surviving, initial encounter |
| 851.55 | Cerebellar or brain stem contusion with open intracranial wound, with prolonged [more than 24 hours] loss of consciousness without return to pre-existing conscious level | S06.387A | Contusion, laceration, and hemorrhage of brainstem with loss of consciousness of any duration with death due to brain injury prior to regaining consciousness, initial encounter |
| 851.55 | Cerebellar or brain stem contusion with open intracranial wound, with prolonged [more than 24 hours] loss of consciousness without return to pre-existing conscious level | S06.388A | Contusion, laceration, and hemorrhage of brainstem with loss of consciousness of any duration with death due to other cause prior to regaining consciousness, initial encounter |
| 851.55 | Cerebellar or brain stem contusion with open intracranial wound, with prolonged [more than 24 hours] loss of consciousness without return to pre-existing conscious level |  |  |
| 851.56 | Cerebellar or brain stem contusion with open intracranial wound, with loss of consciousness of unspecified duration | S06.379A | Contusion, laceration, and hemorrhage of cerebellum with loss of consciousness of unspecified duration, initial encounter |
| 851.56 | Cerebellar or brain stem contusion with open intracranial wound, with loss of consciousness of unspecified duration | S06.389A | Contusion, laceration, and hemorrhage of brainstem with loss of consciousness of unspecified duration, initial encounter |
| 851.56 | Cerebellar or brain stem contusion with open intracranial wound, with loss of consciousness of unspecified duration |  |  |
| 851.59 | Cerebellar or brain stem contusion with open intracranial wound, with concussion, unspecified | S06.370A | Contusion, laceration, and hemorrhage of cerebellum without loss of consciousness, initial encounter |
| 851.59 | Cerebellar or brain stem contusion with open intracranial wound, with concussion, unspecified | S06.380A | Contusion, laceration, and hemorrhage of brainstem without loss of consciousness, initial encounter |
| 851.59 | Cerebellar or brain stem contusion with open intracranial wound, with concussion, unspecified |  |  |
| 851.60 | Cerebellar or brain stem laceration without mention of open intracranial wound, unspecified state of consciousness | S06.370A | Contusion, laceration, and hemorrhage of cerebellum without loss of consciousness, initial encounter |
| 851.60 | Cerebellar or brain stem laceration without mention of open intracranial wound, unspecified state of consciousness | S06.380A | Contusion, laceration, and hemorrhage of brainstem without loss of consciousness, initial encounter |
| 851.61 | Cerebellar or brain stem laceration without mention of open intracranial wound, with no loss of consciousness | S06.370A | Contusion, laceration, and hemorrhage of cerebellum without loss of consciousness, initial encounter |
| 851.61 | Cerebellar or brain stem laceration without mention of open intracranial wound, with no loss of consciousness | S06.380A | Contusion, laceration, and hemorrhage of brainstem without loss of consciousness, initial encounter |
| 851.62 | Cerebellar or brain stem laceration without mention of open intracranial wound, with brief [less than 1 hour] loss of consciousness | S06.371A | Contusion, laceration, and hemorrhage of cerebellum with loss of consciousness of 30 minutes or less, initial encounter |
| 851.62 | Cerebellar or brain stem laceration without mention of open intracranial wound, with brief [less than 1 hour] loss of consciousness | S06.372A | Contusion, laceration, and hemorrhage of cerebellum with loss of consciousness of 31 minutes to 59 minutes, initial encounter |
| 851.62 | Cerebellar or brain stem laceration without mention of open intracranial wound, with brief [less than 1 hour] loss of consciousness | S06.381A | Contusion, laceration, and hemorrhage of brainstem with loss of consciousness of 30 minutes or less, initial encounter |
| 851.62 | Cerebellar or brain stem laceration without mention of open intracranial wound, with brief [less than 1 hour] loss of consciousness | S06.382A | Contusion, laceration, and hemorrhage of brainstem with loss of consciousness of 31 minutes to 59 minutes, initial encounter |
| 851.63 | Cerebellar or brain stem laceration without mention of open intracranial wound, with moderate [1-24 hours] loss of consciousness | S06.373A | Contusion, laceration, and hemorrhage of cerebellum with loss of consciousness of 1 hour to 5 hours 59 minutes, initial encounter |
| 851.63 | Cerebellar or brain stem laceration without mention of open intracranial wound, with moderate [1-24 hours] loss of consciousness | S06.374A | Contusion, laceration, and hemorrhage of cerebellum with loss of consciousness of 6 hours to 24 hours, initial encounter |
| 851.63 | Cerebellar or brain stem laceration without mention of open intracranial wound, with moderate [1-24 hours] loss of consciousness | S06.383A | Contusion, laceration, and hemorrhage of brainstem with loss of consciousness of 1 hour to 5 hours 59 minutes, initial encounter |
| 851.63 | Cerebellar or brain stem laceration without mention of open intracranial wound, with moderate [1-24 hours] loss of consciousness | S06.384A | Contusion, laceration, and hemorrhage of brainstem with loss of consciousness of 6 hours to 24 hours, initial encounter |
| 851.64 | Cerebellar or brain stem laceration without mention of open intracranial wound, with prolonged [more than 24 hours] loss of consciousness and return to pre-existing conscious level | S06.375A | Contusion, laceration, and hemorrhage of cerebellum with loss of consciousness greater than 24 hours with return to pre-existing conscious level, initial encounter |
| 851.64 | Cerebellar or brain stem laceration without mention of open intracranial wound, with prolonged [more than 24 hours] loss of consciousness and return to pre-existing conscious level | S06.385A | Contusion, laceration, and hemorrhage of brainstem with loss of consciousness greater than 24 hours with return to pre-existing conscious level, initial encounter |
| 851.65 | Cerebellar or brain stem laceration without mention of open intracranial wound, with prolonged [more than 24 hours] loss of consciousness without return to pre-existing conscious level | S06.376A | Contusion, laceration, and hemorrhage of cerebellum with loss of consciousness greater than 24 hours without return to pre-existing conscious level with patient surviving, initial encounter |
| 851.65 | Cerebellar or brain stem laceration without mention of open intracranial wound, with prolonged [more than 24 hours] loss of consciousness without return to pre-existing conscious level | S06.377A | Contusion, laceration, and hemorrhage of cerebellum with loss of consciousness of any duration with death due to brain injury prior to regaining consciousness, initial encounter |
| 851.65 | Cerebellar or brain stem laceration without mention of open intracranial wound, with prolonged [more than 24 hours] loss of consciousness without return to pre-existing conscious level | S06.378A | Contusion, laceration, and hemorrhage of cerebellum with loss of consciousness of any duration with death due to other cause prior to regaining consciousness, initial encounter |
| 851.65 | Cerebellar or brain stem laceration without mention of open intracranial wound, with prolonged [more than 24 hours] loss of consciousness without return to pre-existing conscious level | S06.386A | Contusion, laceration, and hemorrhage of brainstem with loss of consciousness greater than 24 hours without return to pre-existing conscious level with patient surviving, initial encounter |
| 851.65 | Cerebellar or brain stem laceration without mention of open intracranial wound, with prolonged [more than 24 hours] loss of consciousness without return to pre-existing conscious level | S06.387A | Contusion, laceration, and hemorrhage of brainstem with loss of consciousness of any duration with death due to brain injury prior to regaining consciousness, initial encounter |
| 851.65 | Cerebellar or brain stem laceration without mention of open intracranial wound, with prolonged [more than 24 hours] loss of consciousness without return to pre-existing conscious level | S06.388A | Contusion, laceration, and hemorrhage of brainstem with loss of consciousness of any duration with death due to other cause prior to regaining consciousness, initial encounter |
| 851.66 | Cerebellar or brain stem laceration without mention of open intracranial wound, with loss of consciousness of unspecified duration | S06.379A | Contusion, laceration, and hemorrhage of cerebellum with loss of consciousness of unspecified duration, initial encounter |
| 851.66 | Cerebellar or brain stem laceration without mention of open intracranial wound, with loss of consciousness of unspecified duration | S06.389A | Contusion, laceration, and hemorrhage of brainstem with loss of consciousness of unspecified duration, initial encounter |
| 851.69 | Cerebellar or brain stem laceration without mention of open intracranial wound, with concussion, unspecified | S06.370A | Contusion, laceration, and hemorrhage of cerebellum without loss of consciousness, initial encounter |
| 851.69 | Cerebellar or brain stem laceration without mention of open intracranial wound, with concussion, unspecified | S06.380A | Contusion, laceration, and hemorrhage of brainstem without loss of consciousness, initial encounter |
| 851.70 | Cerebellar or brain stem laceration with open intracranial wound, unspecified state of consciousness | S06.370A | Contusion, laceration, and hemorrhage of cerebellum without loss of consciousness, initial encounter |
| 851.70 | Cerebellar or brain stem laceration with open intracranial wound, unspecified state of consciousness | S06.380A | Contusion, laceration, and hemorrhage of brainstem without loss of consciousness, initial encounter |
| 851.70 | Cerebellar or brain stem laceration with open intracranial wound, unspecified state of consciousness |  |  |
| 851.71 | Cerebellar or brain stem laceration with open intracranial wound, with no loss of consciousness | S06.370A | Contusion, laceration, and hemorrhage of cerebellum without loss of consciousness, initial encounter |
| 851.71 | Cerebellar or brain stem laceration with open intracranial wound, with no loss of consciousness | S06.380A | Contusion, laceration, and hemorrhage of brainstem without loss of consciousness, initial encounter |
| 851.71 | Cerebellar or brain stem laceration with open intracranial wound, with no loss of consciousness |  |  |
| 851.72 | Cerebellar or brain stem laceration with open intracranial wound, with brief [less than one hour] loss of consciousness | S06.371A | Contusion, laceration, and hemorrhage of cerebellum with loss of consciousness of 30 minutes or less, initial encounter |
| 851.72 | Cerebellar or brain stem laceration with open intracranial wound, with brief [less than one hour] loss of consciousness | S06.372A | Contusion, laceration, and hemorrhage of cerebellum with loss of consciousness of 31 minutes to 59 minutes, initial encounter |
| 851.72 | Cerebellar or brain stem laceration with open intracranial wound, with brief [less than one hour] loss of consciousness | S06.381A | Contusion, laceration, and hemorrhage of brainstem with loss of consciousness of 30 minutes or less, initial encounter |
| 851.72 | Cerebellar or brain stem laceration with open intracranial wound, with brief [less than one hour] loss of consciousness | S06.382A | Contusion, laceration, and hemorrhage of brainstem with loss of consciousness of 31 minutes to 59 minutes, initial encounter |
| 851.72 | Cerebellar or brain stem laceration with open intracranial wound, with brief [less than one hour] loss of consciousness |  |  |
| 851.73 | Cerebellar or brain stem laceration with open intracranial wound, with moderate [1-24 hours] loss of consciousness | S06.373A | Contusion, laceration, and hemorrhage of cerebellum with loss of consciousness of 1 hour to 5 hours 59 minutes, initial encounter |
| 851.73 | Cerebellar or brain stem laceration with open intracranial wound, with moderate [1-24 hours] loss of consciousness | S06.374A | Contusion, laceration, and hemorrhage of cerebellum with loss of consciousness of 6 hours to 24 hours, initial encounter |
| 851.73 | Cerebellar or brain stem laceration with open intracranial wound, with moderate [1-24 hours] loss of consciousness | S06.383A | Contusion, laceration, and hemorrhage of brainstem with loss of consciousness of 1 hour to 5 hours 59 minutes, initial encounter |
| 851.73 | Cerebellar or brain stem laceration with open intracranial wound, with moderate [1-24 hours] loss of consciousness | S06.384A | Contusion, laceration, and hemorrhage of brainstem with loss of consciousness of 6 hours to 24 hours, initial encounter |
| 851.73 | Cerebellar or brain stem laceration with open intracranial wound, with moderate [1-24 hours] loss of consciousness |  |  |
| 851.74 | Cerebellar or brain stem laceration with open intracranial wound, with prolonged [more than 24 hours] loss of consciousness and return to pre-existing conscious level | S06.375A | Contusion, laceration, and hemorrhage of cerebellum with loss of consciousness greater than 24 hours with return to pre-existing conscious level, initial encounter |
| 851.74 | Cerebellar or brain stem laceration with open intracranial wound, with prolonged [more than 24 hours] loss of consciousness and return to pre-existing conscious level | S06.385A | Contusion, laceration, and hemorrhage of brainstem with loss of consciousness greater than 24 hours with return to pre-existing conscious level, initial encounter |
| 851.74 | Cerebellar or brain stem laceration with open intracranial wound, with prolonged [more than 24 hours] loss of consciousness and return to pre-existing conscious level |  |  |
| 851.75 | Cerebellar or brain stem laceration with open intracranial wound, with prolonged [more than 24 hours] loss of consciousness without return to pre-existing conscious level | S06.376A | Contusion, laceration, and hemorrhage of cerebellum with loss of consciousness greater than 24 hours without return to pre-existing conscious level with patient surviving, initial encounter |
| 851.75 | Cerebellar or brain stem laceration with open intracranial wound, with prolonged [more than 24 hours] loss of consciousness without return to pre-existing conscious level | S06.377A | Contusion, laceration, and hemorrhage of cerebellum with loss of consciousness of any duration with death due to brain injury prior to regaining consciousness, initial encounter |
| 851.75 | Cerebellar or brain stem laceration with open intracranial wound, with prolonged [more than 24 hours] loss of consciousness without return to pre-existing conscious level | S06.378A | Contusion, laceration, and hemorrhage of cerebellum with loss of consciousness of any duration with death due to other cause prior to regaining consciousness, initial encounter |
| 851.75 | Cerebellar or brain stem laceration with open intracranial wound, with prolonged [more than 24 hours] loss of consciousness without return to pre-existing conscious level | S06.386A | Contusion, laceration, and hemorrhage of brainstem with loss of consciousness greater than 24 hours without return to pre-existing conscious level with patient surviving, initial encounter |
| 851.75 | Cerebellar or brain stem laceration with open intracranial wound, with prolonged [more than 24 hours] loss of consciousness without return to pre-existing conscious level | S06.387A | Contusion, laceration, and hemorrhage of brainstem with loss of consciousness of any duration with death due to brain injury prior to regaining consciousness, initial encounter |
| 851.75 | Cerebellar or brain stem laceration with open intracranial wound, with prolonged [more than 24 hours] loss of consciousness without return to pre-existing conscious level | S06.388A | Contusion, laceration, and hemorrhage of brainstem with loss of consciousness of any duration with death due to other cause prior to regaining consciousness, initial encounter |
| 851.75 | Cerebellar or brain stem laceration with open intracranial wound, with prolonged [more than 24 hours] loss of consciousness without return to pre-existing conscious level |  |  |
| 851.76 | Cerebellar or brain stem laceration with open intracranial wound, with loss of consciousness of unspecified duration | S06.379A | Contusion, laceration, and hemorrhage of cerebellum with loss of consciousness of unspecified duration, initial encounter |
| 851.76 | Cerebellar or brain stem laceration with open intracranial wound, with loss of consciousness of unspecified duration | S06.389A | Contusion, laceration, and hemorrhage of brainstem with loss of consciousness of unspecified duration, initial encounter |
| 851.76 | Cerebellar or brain stem laceration with open intracranial wound, with loss of consciousness of unspecified duration |  |  |
| 851.79 | Cerebellar or brain stem laceration with open intracranial wound, with concussion, unspecified | S06.370A | Contusion, laceration, and hemorrhage of cerebellum without loss of consciousness, initial encounter |
| 851.79 | Cerebellar or brain stem laceration with open intracranial wound, with concussion, unspecified | S06.380A | Contusion, laceration, and hemorrhage of brainstem without loss of consciousness, initial encounter |
| 851.79 | Cerebellar or brain stem laceration with open intracranial wound, with concussion, unspecified |  |  |
| 851.80 | Other and unspecified cerebral laceration and contusion, without mention of open intracranial wound, unspecified state of consciousness | S06.330A | Contusion and laceration of cerebrum, unspecified, without loss of consciousness, initial encounter |
| 851.81 | Other and unspecified cerebral laceration and contusion, without mention of open intracranial wound, with no loss of consciousness | S06.310A | Contusion and laceration of right cerebrum without loss of consciousness, initial encounter |
| 851.81 | Other and unspecified cerebral laceration and contusion, without mention of open intracranial wound, with no loss of consciousness | S06.320A | Contusion and laceration of left cerebrum without loss of consciousness, initial encounter |
| 851.81 | Other and unspecified cerebral laceration and contusion, without mention of open intracranial wound, with no loss of consciousness | S06.330A | Contusion and laceration of cerebrum, unspecified, without loss of consciousness, initial encounter |
| 851.82 | Other and unspecified cerebral laceration and contusion, without mention of open intracranial wound, with brief [less than one hour] loss of consciousness | S06.311A | Contusion and laceration of right cerebrum with loss of consciousness of 30 minutes or less, initial encounter |
| 851.82 | Other and unspecified cerebral laceration and contusion, without mention of open intracranial wound, with brief [less than one hour] loss of consciousness | S06.312A | Contusion and laceration of right cerebrum with loss of consciousness of 31 minutes to 59 minutes, initial encounter |
| 851.82 | Other and unspecified cerebral laceration and contusion, without mention of open intracranial wound, with brief [less than one hour] loss of consciousness | S06.321A | Contusion and laceration of left cerebrum with loss of consciousness of 30 minutes or less, initial encounter |
| 851.82 | Other and unspecified cerebral laceration and contusion, without mention of open intracranial wound, with brief [less than one hour] loss of consciousness | S06.322A | Contusion and laceration of left cerebrum with loss of consciousness of 31 minutes to 59 minutes, initial encounter |
| 851.82 | Other and unspecified cerebral laceration and contusion, without mention of open intracranial wound, with brief [less than one hour] loss of consciousness | S06.331A | Contusion and laceration of cerebrum, unspecified, with loss of consciousness of 30 minutes or less, initial encounter |
| 851.82 | Other and unspecified cerebral laceration and contusion, without mention of open intracranial wound, with brief [less than one hour] loss of consciousness | S06.332A | Contusion and laceration of cerebrum, unspecified, with loss of consciousness of 31 minutes to 59 minutes, initial encounter |
| 851.83 | Other and unspecified cerebral laceration and contusion, without mention of open intracranial wound, with moderate [1-24 hours] loss of consciousness | S06.313A | Contusion and laceration of right cerebrum with loss of consciousness of 1 hour to 5 hours 59 minutes, initial encounter |
| 851.83 | Other and unspecified cerebral laceration and contusion, without mention of open intracranial wound, with moderate [1-24 hours] loss of consciousness | S06.314A | Contusion and laceration of right cerebrum with loss of consciousness of 6 hours to 24 hours, initial encounter |
| 851.83 | Other and unspecified cerebral laceration and contusion, without mention of open intracranial wound, with moderate [1-24 hours] loss of consciousness | S06.323A | Contusion and laceration of left cerebrum with loss of consciousness of 1 hour to 5 hours 59 minutes, initial encounter |
| 851.83 | Other and unspecified cerebral laceration and contusion, without mention of open intracranial wound, with moderate [1-24 hours] loss of consciousness | S06.324A | Contusion and laceration of left cerebrum with loss of consciousness of 6 hours to 24 hours, initial encounter |
| 851.83 | Other and unspecified cerebral laceration and contusion, without mention of open intracranial wound, with moderate [1-24 hours] loss of consciousness | S06.333A | Contusion and laceration of cerebrum, unspecified, with loss of consciousness of 1 hour to 5 hours 59 minutes, initial encounter |
| 851.83 | Other and unspecified cerebral laceration and contusion, without mention of open intracranial wound, with moderate [1-24 hours] loss of consciousness | S06.334A | Contusion and laceration of cerebrum, unspecified, with loss of consciousness of 6 hours to 24 hours, initial encounter |
| 851.84 | Other and unspecified cerebral laceration and contusion, without mention of open intracranial wound, with prolonged [more than 24 hours] loss of consciousness and return to pre- existing conscious level | S06.315A | Contusion and laceration of right cerebrum with loss of consciousness greater than 24 hours with return to pre-existing conscious level, initial encounter |
| 851.84 | Other and unspecified cerebral laceration and contusion, without mention of open intracranial wound, with prolonged [more than 24 hours] loss of consciousness and return to pre- existing conscious level | S06.325A | Contusion and laceration of left cerebrum with loss of consciousness greater than 24 hours with return to pre-existing conscious level, initial encounter |
| 851.84 | Other and unspecified cerebral laceration and contusion, without mention of open intracranial wound, with prolonged [more than 24 hours] loss of consciousness and return to pre- existing conscious level | S06.335A | Contusion and laceration of cerebrum, unspecified, with loss of consciousness greater than 24 hours with return to pre-existing conscious level, initial encounter |
| 851.85 | Other and unspecified cerebral laceration and contusion, without mention of open intracranial wound, with prolonged [more than 24 hours] loss of consciousness without return to pre-existing conscious level | S06.316A | Contusion and laceration of right cerebrum with loss of consciousness greater than 24 hours without return to pre-existing conscious level with patient surviving, initial encounter |
| 851.85 | Other and unspecified cerebral laceration and contusion, without mention of open intracranial wound, with prolonged [more than 24 hours] loss of consciousness without return to pre-existing conscious level | S06.317A | Contusion and laceration of right cerebrum with loss of consciousness of any duration with death due to brain injury prior to regaining consciousness, initial encounter |
| 851.85 | Other and unspecified cerebral laceration and contusion, without mention of open intracranial wound, with prolonged [more than 24 hours] loss of consciousness without return to pre-existing conscious level | S06.318A | Contusion and laceration of right cerebrum with loss of consciousness of any duration with death due to other cause prior to regaining consciousness, initial encounter |
| 851.85 | Other and unspecified cerebral laceration and contusion, without mention of open intracranial wound, with prolonged [more than 24 hours] loss of consciousness without return to pre-existing conscious level | S06.326A | Contusion and laceration of left cerebrum with loss of consciousness greater than 24 hours without return to pre-existing conscious level with patient surviving, initial encounter |
| 851.85 | Other and unspecified cerebral laceration and contusion, without mention of open intracranial wound, with prolonged [more than 24 hours] loss of consciousness without return to pre-existing conscious level | S06.327A | Contusion and laceration of left cerebrum with loss of consciousness of any duration with death due to brain injury prior to regaining consciousness, initial encounter |
| 851.85 | Other and unspecified cerebral laceration and contusion, without mention of open intracranial wound, with prolonged [more than 24 hours] loss of consciousness without return to pre-existing conscious level | S06.328A | Contusion and laceration of left cerebrum with loss of consciousness of any duration with death due to other cause prior to regaining consciousness, initial encounter |
| 851.85 | Other and unspecified cerebral laceration and contusion, without mention of open intracranial wound, with prolonged [more than 24 hours] loss of consciousness without return to pre-existing conscious level | S06.336A | Contusion and laceration of cerebrum, unspecified, with loss of consciousness greater than 24 hours without return to pre-existing conscious level with patient surviving, initial encounter |
| 851.85 | Other and unspecified cerebral laceration and contusion, without mention of open intracranial wound, with prolonged [more than 24 hours] loss of consciousness without return to pre-existing conscious level | S06.337A | Contusion and laceration of cerebrum, unspecified, with loss of consciousness of any duration with death due to brain injury prior to regaining consciousness, initial encounter |
| 851.85 | Other and unspecified cerebral laceration and contusion, without mention of open intracranial wound, with prolonged [more than 24 hours] loss of consciousness without return to pre-existing conscious level | S06.338A | Contusion and laceration of cerebrum, unspecified, with loss of consciousness of any duration with death due to other cause prior to regaining consciousness, initial encounter |
| 851.86 | Other and unspecified cerebral laceration and contusion, without mention of open intracranial wound, with loss of consciousness of unspecified duration | S06.319A | Contusion and laceration of right cerebrum with loss of consciousness of unspecified duration, initial encounter |
| 851.86 | Other and unspecified cerebral laceration and contusion, without mention of open intracranial wound, with loss of consciousness of unspecified duration | S06.329A | Contusion and laceration of left cerebrum with loss of consciousness of unspecified duration, initial encounter |
| 851.86 | Other and unspecified cerebral laceration and contusion, without mention of open intracranial wound, with loss of consciousness of unspecified duration | S06.339A | Contusion and laceration of cerebrum, unspecified, with loss of consciousness of unspecified duration, initial encounter |
| 851.89 | Other and unspecified cerebral laceration and contusion, without mention of open intracranial wound, with concussion, unspecified | S06.330A | Contusion and laceration of cerebrum, unspecified, without loss of consciousness, initial encounter |
| 851.90 | Other and unspecified cerebral laceration and contusion, with open intracranial wound, unspecified state of consciousness | S06.330A | Contusion and laceration of cerebrum, unspecified, without loss of consciousness, initial encounter |
| 851.90 | Other and unspecified cerebral laceration and contusion, with open intracranial wound, unspecified state of consciousness |  |  |
| 851.91 | Other and unspecified cerebral laceration and contusion, with open intracranial wound, with no loss of consciousness | S06.330A | Contusion and laceration of cerebrum, unspecified, without loss of consciousness, initial encounter |
| 851.91 | Other and unspecified cerebral laceration and contusion, with open intracranial wound, with no loss of consciousness |  |  |
| 851.92 | Other and unspecified cerebral laceration and contusion, with open intracranial wound, with brief [less than one hour] loss of consciousness | S06.331A | Contusion and laceration of cerebrum, unspecified, with loss of consciousness of 30 minutes or less, initial encounter |
| 851.92 | Other and unspecified cerebral laceration and contusion, with open intracranial wound, with brief [less than one hour] loss of consciousness | S06.332A | Contusion and laceration of cerebrum, unspecified, with loss of consciousness of 31 minutes to 59 minutes, initial encounter |
| 851.92 | Other and unspecified cerebral laceration and contusion, with open intracranial wound, with brief [less than one hour] loss of consciousness |  |  |
| 851.93 | Other and unspecified cerebral laceration and contusion, with open intracranial wound, with moderate [1-24 hours] loss of consciousness | S06.333A | Contusion and laceration of cerebrum, unspecified, with loss of consciousness of 1 hour to 5 hours 59 minutes, initial encounter |
| 851.93 | Other and unspecified cerebral laceration and contusion, with open intracranial wound, with moderate [1-24 hours] loss of consciousness | S06.334A | Contusion and laceration of cerebrum, unspecified, with loss of consciousness of 6 hours to 24 hours, initial encounter |
| 851.93 | Other and unspecified cerebral laceration and contusion, with open intracranial wound, with moderate [1-24 hours] loss of consciousness |  |  |
| 851.94 | Other and unspecified cerebral laceration and contusion, with open intracranial wound, with prolonged [more than 24 hours] loss of consciousness and return to pre-existing conscious level | S06.335A | Contusion and laceration of cerebrum, unspecified, with loss of consciousness greater than 24 hours with return to pre-existing conscious level, initial encounter |
| 851.94 | Other and unspecified cerebral laceration and contusion, with open intracranial wound, with prolonged [more than 24 hours] loss of consciousness and return to pre-existing conscious level |  |  |
| 851.95 | Other and unspecified cerebral laceration and contusion, with open intracranial wound, with prolonged [more than 24 hours] loss of consciousness without return to pre-existing conscious level | S06.336A | Contusion and laceration of cerebrum, unspecified, with loss of consciousness greater than 24 hours without return to pre-existing conscious level with patient surviving, initial encounter |
| 851.95 | Other and unspecified cerebral laceration and contusion, with open intracranial wound, with prolonged [more than 24 hours] loss of consciousness without return to pre-existing conscious level | S06.337A | Contusion and laceration of cerebrum, unspecified, with loss of consciousness of any duration with death due to brain injury prior to regaining consciousness, initial encounter |
| 851.95 | Other and unspecified cerebral laceration and contusion, with open intracranial wound, with prolonged [more than 24 hours] loss of consciousness without return to pre-existing conscious level | S06.338A | Contusion and laceration of cerebrum, unspecified, with loss of consciousness of any duration with death due to other cause prior to regaining consciousness, initial encounter |
| 851.95 | Other and unspecified cerebral laceration and contusion, with open intracranial wound, with prolonged [more than 24 hours] loss of consciousness without return to pre-existing conscious level |  |  |
| 851.96 | Other and unspecified cerebral laceration and contusion, with open intracranial wound, with loss of consciousness of unspecified duration | S06.339A | Contusion and laceration of cerebrum, unspecified, with loss of consciousness of unspecified duration, initial encounter |
| 851.96 | Other and unspecified cerebral laceration and contusion, with open intracranial wound, with loss of consciousness of unspecified duration |  |  |
| 851.99 | Other and unspecified cerebral laceration and contusion, with open intracranial wound, with concussion, unspecified | S06.330A | Contusion and laceration of cerebrum, unspecified, without loss of consciousness, initial encounter |
| 851.99 | Other and unspecified cerebral laceration and contusion, with open intracranial wound, with concussion, unspecified |  |  |
| 852.00 | Subarachnoid hemorrhage following injury without mention of open intracranial wound, unspecified state of consciousness | S06.6X0A | Traumatic subarachnoid hemorrhage without loss of consciousness, initial encounter |
| 852.01 | Subarachnoid hemorrhage following injury without mention of open intracranial wound, with no loss of consciousness | S06.6X0A | Traumatic subarachnoid hemorrhage without loss of consciousness, initial encounter |
| 852.02 | Subarachnoid hemorrhage following injury without mention of open intracranial wound, with brief [less than one hour] loss of consciousness | S06.6X1A | Traumatic subarachnoid hemorrhage with loss of consciousness of 30 minutes or less, initial encounter |
| 852.02 | Subarachnoid hemorrhage following injury without mention of open intracranial wound, with brief [less than one hour] loss of consciousness | S06.6X2A | Traumatic subarachnoid hemorrhage with loss of consciousness of 31 minutes to 59 minutes, initial encounter |
| 852.03 | Subarachnoid hemorrhage following injury without mention of open intracranial wound, with moderate [1-24 hours] loss of consciousness | S06.6X3A | Traumatic subarachnoid hemorrhage with loss of consciousness of 1 hour to 5 hours 59 minutes, initial encounter |
| 852.03 | Subarachnoid hemorrhage following injury without mention of open intracranial wound, with moderate [1-24 hours] loss of consciousness | S06.6X4A | Traumatic subarachnoid hemorrhage with loss of consciousness of 6 hours to 24 hours, initial encounter |
| 852.04 | Subarachnoid hemorrhage following injury without mention of open intracranial wound, with prolonged [more than 24 hours] loss of consciousness and return to pre-existing conscious level | S06.6X5A | Traumatic subarachnoid hemorrhage with loss of consciousness greater than 24 hours with return to pre-existing conscious level, initial encounter |
| 852.05 | Subarachnoid hemorrhage following injury without mention of open intracranial wound, with prolonged [more than 24 hours] loss of consciousness without return to pre-existing conscious level | S06.6X6A | Traumatic subarachnoid hemorrhage with loss of consciousness greater than 24 hours without return to pre-existing conscious level with patient surviving, initial encounter |
| 852.05 | Subarachnoid hemorrhage following injury without mention of open intracranial wound, with prolonged [more than 24 hours] loss of consciousness without return to pre-existing conscious level | S06.6X7A | Traumatic subarachnoid hemorrhage with loss of consciousness of any duration with death due to brain injury prior to regaining consciousness, initial encounter |
| 852.05 | Subarachnoid hemorrhage following injury without mention of open intracranial wound, with prolonged [more than 24 hours] loss of consciousness without return to pre-existing conscious level | S06.6X8A | Traumatic subarachnoid hemorrhage with loss of consciousness of any duration with death due to other cause prior to regaining consciousness, initial encounter |
| 852.06 | Subarachnoid hemorrhage following injury without mention of open intracranial wound, with loss of consciousness of unspecified duration | S06.6X9A | Traumatic subarachnoid hemorrhage with loss of consciousness of unspecified duration, initial encounter |
| 852.09 | Subarachnoid hemorrhage following injury without mention of open intracranial wound, with concussion, unspecified | S06.6X0A | Traumatic subarachnoid hemorrhage without loss of consciousness, initial encounter |
| 852.09 | Subarachnoid hemorrhage following injury without mention of open intracranial wound, with concussion, unspecified | S06.6X9A | Traumatic subarachnoid hemorrhage with loss of consciousness of unspecified duration, initial encounter |
| 852.10 | Subarachnoid hemorrhage following injury with open intracranial wound, unspecified state of consciousness | S06.6X0A | Traumatic subarachnoid hemorrhage without loss of consciousness, initial encounter |
| 852.11 | Subarachnoid hemorrhage following injury with open intracranial wound, with no loss of consciousness | S06.6X0A | Traumatic subarachnoid hemorrhage without loss of consciousness, initial encounter |
| 852.11 | Subarachnoid hemorrhage following injury with open intracranial wound, with no loss of consciousness |  |  |
| 852.12 | Subarachnoid hemorrhage following injury with open intracranial wound, with brief [less than one hour] loss of consciousness | S06.6X1A | Traumatic subarachnoid hemorrhage with loss of consciousness of 30 minutes or less, initial encounter |
| 852.12 | Subarachnoid hemorrhage following injury with open intracranial wound, with brief [less than one hour] loss of consciousness | S06.6X2A | Traumatic subarachnoid hemorrhage with loss of consciousness of 31 minutes to 59 minutes, initial encounter |
| 852.12 | Subarachnoid hemorrhage following injury with open intracranial wound, with brief [less than one hour] loss of consciousness |  |  |
| 852.13 | Subarachnoid hemorrhage following injury with open intracranial wound, with moderate [1-24 hours] loss of consciousness | S06.6X3A | Traumatic subarachnoid hemorrhage with loss of consciousness of 1 hour to 5 hours 59 minutes, initial encounter |
| 852.13 | Subarachnoid hemorrhage following injury with open intracranial wound, with moderate [1-24 hours] loss of consciousness | S06.6X4A | Traumatic subarachnoid hemorrhage with loss of consciousness of 6 hours to 24 hours, initial encounter |
| 852.13 | Subarachnoid hemorrhage following injury with open intracranial wound, with moderate [1-24 hours] loss of consciousness |  |  |
| 852.14 | Subarachnoid hemorrhage following injury with open intracranial wound, with prolonged [more than 24 hours) loss of consciousness and return to pre-existing conscious level | S06.6X5A | Traumatic subarachnoid hemorrhage with loss of consciousness greater than 24 hours with return to pre-existing conscious level, initial encounter |
| 852.14 | Subarachnoid hemorrhage following injury with open intracranial wound, with prolonged [more than 24 hours) loss of consciousness and return to pre-existing conscious level |  |  |
| 852.15 | Subarachnoid hemorrhage following injury with open intracranial wound, with prolonged [more than 24 hours] loss of consciousness without return to pre-existing conscious level | S06.6X6A | Traumatic subarachnoid hemorrhage with loss of consciousness greater than 24 hours without return to pre-existing conscious level with patient surviving, initial encounter |
| 852.15 | Subarachnoid hemorrhage following injury with open intracranial wound, with prolonged [more than 24 hours] loss of consciousness without return to pre-existing conscious level | S06.6X7A | Traumatic subarachnoid hemorrhage with loss of consciousness of any duration with death due to brain injury prior to regaining consciousness, initial encounter |
| 852.15 | Subarachnoid hemorrhage following injury with open intracranial wound, with prolonged [more than 24 hours] loss of consciousness without return to pre-existing conscious level | S06.6X8A | Traumatic subarachnoid hemorrhage with loss of consciousness of any duration with death due to other cause prior to regaining consciousness, initial encounter |
| 852.15 | Subarachnoid hemorrhage following injury with open intracranial wound, with prolonged [more than 24 hours] loss of consciousness without return to pre-existing conscious level |  |  |
| 852.16 | Subarachnoid hemorrhage following injury with open intracranial wound, with loss of consciousness of unspecified duration | S06.6X9A | Traumatic subarachnoid hemorrhage with loss of consciousness of unspecified duration, initial encounter |
| 852.16 | Subarachnoid hemorrhage following injury with open intracranial wound, with loss of consciousness of unspecified duration |  |  |
| 852.19 | Subarachnoid hemorrhage following injury with open intracranial wound, with concussion, unspecified | S06.6X0A | Traumatic subarachnoid hemorrhage without loss of consciousness, initial encounter |
| 852.19 | Subarachnoid hemorrhage following injury with open intracranial wound, with concussion, unspecified | S06.6X9A | Traumatic subarachnoid hemorrhage with loss of consciousness of unspecified duration, initial encounter |
| 852.19 | Subarachnoid hemorrhage following injury with open intracranial wound, with concussion, unspecified |  |  |
| 852.20 | Subdural hemorrhage following injury without mention of open intracranial wound, unspecified state of consciousness | S06.5X0A | Traumatic subdural hemorrhage without loss of consciousness, initial encounter |
| 852.21 | Subdural hemorrhage following injury without mention of open intracranial wound, with no loss of consciousness | S06.5X0A | Traumatic subdural hemorrhage without loss of consciousness, initial encounter |
| 852.22 | Subdural hemorrhage following injury without mention of open intracranial wound, with brief [less than one hour] loss of consciousness | S06.5X1A | Traumatic subdural hemorrhage with loss of consciousness of 30 minutes or less, initial encounter |
| 852.22 | Subdural hemorrhage following injury without mention of open intracranial wound, with brief [less than one hour] loss of consciousness | S06.5X2A | Traumatic subdural hemorrhage with loss of consciousness of 31 minutes to 59 minutes, initial encounter |
| 852.23 | Subdural hemorrhage following injury without mention of open intracranial wound, with moderate [1-24 hours] loss of consciousness | S06.5X3A | Traumatic subdural hemorrhage with loss of consciousness of 1 hour to 5 hours 59 minutes, initial encounter |
| 852.23 | Subdural hemorrhage following injury without mention of open intracranial wound, with moderate [1-24 hours] loss of consciousness | S06.5X4A | Traumatic subdural hemorrhage with loss of consciousness of 6 hours to 24 hours, initial encounter |
| 852.24 | Subdural hemorrhage following injury without mention of open intracranial wound, with prolonged [more than 24 hours] loss of consciousness and return to pre-existing conscious level | S06.5X5A | Traumatic subdural hemorrhage with loss of consciousness greater than 24 hours with return to pre-existing conscious level, initial encounter |
| 852.25 | Subdural hemorrhage following injury without mention of open intracranial wound, with prolonged [more than 24 hours] loss of consciousness without return to pre-existing conscious level | S06.5X6A | Traumatic subdural hemorrhage with loss of consciousness greater than 24 hours without return to pre-existing conscious level with patient surviving, initial encounter |
| 852.25 | Subdural hemorrhage following injury without mention of open intracranial wound, with prolonged [more than 24 hours] loss of consciousness without return to pre-existing conscious level | S06.5X7A | Traumatic subdural hemorrhage with loss of consciousness of any duration with death due to brain injury before regaining consciousness, initial encounter |
| 852.25 | Subdural hemorrhage following injury without mention of open intracranial wound, with prolonged [more than 24 hours] loss of consciousness without return to pre-existing conscious level | S06.5X8A | Traumatic subdural hemorrhage with loss of consciousness of any duration with death due to other cause before regaining consciousness, initial encounter |
| 852.26 | Subdural hemorrhage following injury without mention of open intracranial wound, with loss of consciousness of unspecified duration | S06.5X9A | Traumatic subdural hemorrhage with loss of consciousness of unspecified duration, initial encounter |
| 852.29 | Subdural hemorrhage following injury without mention of open intracranial wound, with concussion, unspecified | S06.5X0A | Traumatic subdural hemorrhage without loss of consciousness, initial encounter |
| 852.29 | Subdural hemorrhage following injury without mention of open intracranial wound, with concussion, unspecified | S06.5X9A | Traumatic subdural hemorrhage with loss of consciousness of unspecified duration, initial encounter |
| 852.30 | Subdural hemorrhage following injury with open intracranial wound, unspecified state of consciousness | S06.5X0A | Traumatic subdural hemorrhage without loss of consciousness, initial encounter |
| 852.30 | Subdural hemorrhage following injury with open intracranial wound, unspecified state of consciousness |  |  |
| 852.31 | Subdural hemorrhage following injury with open intracranial wound, with no loss of consciousness | S06.5X0A | Traumatic subdural hemorrhage without loss of consciousness, initial encounter |
| 852.31 | Subdural hemorrhage following injury with open intracranial wound, with no loss of consciousness |  |  |
| 852.32 | Subdural hemorrhage following injury with open intracranial wound, with brief [less than one hour] loss of consciousness | S06.5X1A | Traumatic subdural hemorrhage with loss of consciousness of 30 minutes or less, initial encounter |
| 852.32 | Subdural hemorrhage following injury with open intracranial wound, with brief [less than one hour] loss of consciousness | S06.5X2A | Traumatic subdural hemorrhage with loss of consciousness of 31 minutes to 59 minutes, initial encounter |
| 852.32 | Subdural hemorrhage following injury with open intracranial wound, with brief [less than one hour] loss of consciousness |  |  |
| 852.33 | Subdural hemorrhage following injury with open intracranial wound, with moderate [1-24 hours] loss of consciousness | S06.5X3A | Traumatic subdural hemorrhage with loss of consciousness of 1 hour to 5 hours 59 minutes, initial encounter |
| 852.33 | Subdural hemorrhage following injury with open intracranial wound, with moderate [1-24 hours] loss of consciousness | S06.5X4A | Traumatic subdural hemorrhage with loss of consciousness of 6 hours to 24 hours, initial encounter |
| 852.33 | Subdural hemorrhage following injury with open intracranial wound, with moderate [1-24 hours] loss of consciousness |  |  |
| 852.34 | Subdural hemorrhage following injury with open intracranial wound, with prolonged [more than 24 hours] loss of consciousness and return to pre-existing conscious level | S06.5X5A | Traumatic subdural hemorrhage with loss of consciousness greater than 24 hours with return to pre-existing conscious level, initial encounter |
| 852.34 | Subdural hemorrhage following injury with open intracranial wound, with prolonged [more than 24 hours] loss of consciousness and return to pre-existing conscious level |  |  |
| 852.35 | Subdural hemorrhage following injury with open intracranial wound, with prolonged [more than 24 hours] loss of consciousness without return to pre-existing conscious level | S06.5X6A | Traumatic subdural hemorrhage with loss of consciousness greater than 24 hours without return to pre-existing conscious level with patient surviving, initial encounter |
| 852.35 | Subdural hemorrhage following injury with open intracranial wound, with prolonged [more than 24 hours] loss of consciousness without return to pre-existing conscious level | S06.5X7A | Traumatic subdural hemorrhage with loss of consciousness of any duration with death due to brain injury before regaining consciousness, initial encounter |
| 852.35 | Subdural hemorrhage following injury with open intracranial wound, with prolonged [more than 24 hours] loss of consciousness without return to pre-existing conscious level | S06.5X8A | Traumatic subdural hemorrhage with loss of consciousness of any duration with death due to other cause before regaining consciousness, initial encounter |
| 852.35 | Subdural hemorrhage following injury with open intracranial wound, with prolonged [more than 24 hours] loss of consciousness without return to pre-existing conscious level |  |  |
| 852.36 | Subdural hemorrhage following injury with open intracranial wound, with loss of consciousness of unspecified duration | S06.5X9A | Traumatic subdural hemorrhage with loss of consciousness of unspecified duration, initial encounter |
| 852.36 | Subdural hemorrhage following injury with open intracranial wound, with loss of consciousness of unspecified duration |  |  |
| 852.39 | Subdural hemorrhage following injury with open intracranial wound, with concussion, unspecified | S06.5X0A | Traumatic subdural hemorrhage without loss of consciousness, initial encounter |
| 852.39 | Subdural hemorrhage following injury with open intracranial wound, with concussion, unspecified | S06.5X9A | Traumatic subdural hemorrhage with loss of consciousness of unspecified duration, initial encounter |
| 852.39 | Subdural hemorrhage following injury with open intracranial wound, with concussion, unspecified |  |  |
| 852.40 | Extradural hemorrhage following injury without mention of open intracranial wound, unspecified state of consciousness | S06.4X0A | Epidural hemorrhage without loss of consciousness, initial encounter |
| 852.41 | Extradural hemorrhage following injury without mention of open intracranial wound, with no loss of consciousness | S06.4X0A | Epidural hemorrhage without loss of consciousness, initial encounter |
| 852.42 | Extradural hemorrhage following injury without mention of open intracranial wound, with brief [less than 1 hour] loss of consciousness | S06.4X1A | Epidural hemorrhage with loss of consciousness of 30 minutes or less, initial encounter |
| 852.42 | Extradural hemorrhage following injury without mention of open intracranial wound, with brief [less than 1 hour] loss of consciousness | S06.4X2A | Epidural hemorrhage with loss of consciousness of 31 minutes to 59 minutes, initial encounter |
| 852.43 | Extradural hemorrhage following injury without mention of open intracranial wound, with moderate [1-24 hours] loss of consciousness | S06.4X3A | Epidural hemorrhage with loss of consciousness of 1 hour to 5 hours 59 minutes, initial encounter |
| 852.43 | Extradural hemorrhage following injury without mention of open intracranial wound, with moderate [1-24 hours] loss of consciousness | S06.4X4A | Epidural hemorrhage with loss of consciousness of 6 hours to 24 hours, initial encounter |
| 852.44 | Extradural hemorrhage following injury without mention of open intracranial wound, with prolonged [more than 24 hours] loss of consciousness and return to pre-existing conscious level | S06.4X5A | Epidural hemorrhage with loss of consciousness greater than 24 hours with return to pre-existing conscious level, initial encounter |
| 852.45 | Extradural hemorrhage following injury without mention of open intracranial wound, with prolonged [more than 24 hours] loss of consciousness without return to pre-existing conscious level | S06.4X6A | Epidural hemorrhage with loss of consciousness greater than 24 hours without return to pre-existing conscious level with patient surviving, initial encounter |
| 852.45 | Extradural hemorrhage following injury without mention of open intracranial wound, with prolonged [more than 24 hours] loss of consciousness without return to pre-existing conscious level | S06.4X7A | Epidural hemorrhage with loss of consciousness of any duration with death due to brain injury prior to regaining consciousness, initial encounter |
| 852.45 | Extradural hemorrhage following injury without mention of open intracranial wound, with prolonged [more than 24 hours] loss of consciousness without return to pre-existing conscious level | S06.4X8A | Epidural hemorrhage with loss of consciousness of any duration with death due to other causes prior to regaining consciousness, initial encounter |
| 852.46 | Extradural hemorrhage following injury without mention of open intracranial wound, with loss of consciousness of unspecified duration | S06.4X9A | Epidural hemorrhage with loss of consciousness of unspecified duration, initial encounter |
| 852.49 | Extradural hemorrhage following injury without mention of open intracranial wound, with concussion, unspecified | S06.4X0A | Epidural hemorrhage without loss of consciousness, initial encounter |
| 852.49 | Extradural hemorrhage following injury without mention of open intracranial wound, with concussion, unspecified | S06.4X9A | Epidural hemorrhage with loss of consciousness of unspecified duration, initial encounter |
| 852.50 | Extradural hemorrhage following injury with open intracranial wound, unspecified state of consciousness | S06.4X0A | Epidural hemorrhage without loss of consciousness, initial encounter |
| 852.50 | Extradural hemorrhage following injury with open intracranial wound, unspecified state of consciousness |  |  |
| 852.51 | Extradural hemorrhage following injury with open intracranial wound, with no loss of consciousness | S06.4X0A | Epidural hemorrhage without loss of consciousness, initial encounter |
| 852.51 | Extradural hemorrhage following injury with open intracranial wound, with no loss of consciousness |  |  |
| 852.52 | Extradural hemorrhage following injury with open intracranial wound, with brief [less than one hour] loss of consciousness | S06.4X1A | Epidural hemorrhage with loss of consciousness of 30 minutes or less, initial encounter |
| 852.52 | Extradural hemorrhage following injury with open intracranial wound, with brief [less than one hour] loss of consciousness | S06.4X2A | Epidural hemorrhage with loss of consciousness of 31 minutes to 59 minutes, initial encounter |
| 852.52 | Extradural hemorrhage following injury with open intracranial wound, with brief [less than one hour] loss of consciousness |  |  |
| 852.53 | Extradural hemorrhage following injury with open intracranial wound, with moderate [1-24 hours] loss of consciousness | S06.4X3A | Epidural hemorrhage with loss of consciousness of 1 hour to 5 hours 59 minutes, initial encounter |
| 852.53 | Extradural hemorrhage following injury with open intracranial wound, with moderate [1-24 hours] loss of consciousness | S06.4X4A | Epidural hemorrhage with loss of consciousness of 6 hours to 24 hours, initial encounter |
| 852.53 | Extradural hemorrhage following injury with open intracranial wound, with moderate [1-24 hours] loss of consciousness |  |  |
| 852.54 | Extradural hemorrhage following injury with open intracranial wound, with prolonged [more than 24 hours] loss of consciousness and return to pre-existing conscious level | S06.4X5A | Epidural hemorrhage with loss of consciousness greater than 24 hours with return to pre-existing conscious level, initial encounter |
| 852.54 | Extradural hemorrhage following injury with open intracranial wound, with prolonged [more than 24 hours] loss of consciousness and return to pre-existing conscious level |  |  |
| 852.55 | Extradural hemorrhage following injury with open intracranial wound, with prolonged [more than 24 hours] loss of consciousness without return to pre-existing conscious level | S06.4X6A | Epidural hemorrhage with loss of consciousness greater than 24 hours without return to pre-existing conscious level with patient surviving, initial encounter |
| 852.55 | Extradural hemorrhage following injury with open intracranial wound, with prolonged [more than 24 hours] loss of consciousness without return to pre-existing conscious level | S06.4X7A | Epidural hemorrhage with loss of consciousness of any duration with death due to brain injury prior to regaining consciousness, initial encounter |
| 852.55 | Extradural hemorrhage following injury with open intracranial wound, with prolonged [more than 24 hours] loss of consciousness without return to pre-existing conscious level | S06.4X8A | Epidural hemorrhage with loss of consciousness of any duration with death due to other causes prior to regaining consciousness, initial encounter |
| 852.55 | Extradural hemorrhage following injury with open intracranial wound, with prolonged [more than 24 hours] loss of consciousness without return to pre-existing conscious level |  |  |
| 852.56 | Extradural hemorrhage following injury with open intracranial wound, with loss of consciousness of unspecified duration | S06.4X9A | Epidural hemorrhage with loss of consciousness of unspecified duration, initial encounter |
| 852.56 | Extradural hemorrhage following injury with open intracranial wound, with loss of consciousness of unspecified duration |  |  |
| 852.59 | Extradural hemorrhage following injury with open intracranial wound, with concussion, unspecified | S06.4X0A | Epidural hemorrhage without loss of consciousness, initial encounter |
| 852.59 | Extradural hemorrhage following injury with open intracranial wound, with concussion, unspecified | S06.4X9A | Epidural hemorrhage with loss of consciousness of unspecified duration, initial encounter |
| 852.59 | Extradural hemorrhage following injury with open intracranial wound, with concussion, unspecified |  |  |
| 853.00 | Other and unspecified intracranial hemorrhage following injury without mention of open intracranial wound, unspecified state of consciousness | S06.360A | Traumatic hemorrhage of cerebrum, unspecified, without loss of consciousness, initial encounter |
| 853.01 | Other and unspecified intracranial hemorrhage following injury without mention of open intracranial wound, with no loss of consciousness | S06.340A | Traumatic hemorrhage of right cerebrum without loss of consciousness, initial encounter |
| 853.01 | Other and unspecified intracranial hemorrhage following injury without mention of open intracranial wound, with no loss of consciousness | S06.350A | Traumatic hemorrhage of left cerebrum without loss of consciousness, initial encounter |
| 853.01 | Other and unspecified intracranial hemorrhage following injury without mention of open intracranial wound, with no loss of consciousness | S06.360A | Traumatic hemorrhage of cerebrum, unspecified, without loss of consciousness, initial encounter |
| 853.02 | Other and unspecified intracranial hemorrhage following injury without mention of open intracranial wound, with brief [less than one hour] loss of consciousness | S06.341A | Traumatic hemorrhage of right cerebrum with loss of consciousness of 30 minutes or less, initial encounter |
| 853.02 | Other and unspecified intracranial hemorrhage following injury without mention of open intracranial wound, with brief [less than one hour] loss of consciousness | S06.342A | Traumatic hemorrhage of right cerebrum with loss of consciousness of 31 minutes to 59 minutes, initial encounter |
| 853.02 | Other and unspecified intracranial hemorrhage following injury without mention of open intracranial wound, with brief [less than one hour] loss of consciousness | S06.351A | Traumatic hemorrhage of left cerebrum with loss of consciousness of 30 minutes or less, initial encounter |
| 853.02 | Other and unspecified intracranial hemorrhage following injury without mention of open intracranial wound, with brief [less than one hour] loss of consciousness | S06.352A | Traumatic hemorrhage of left cerebrum with loss of consciousness of 31 minutes to 59 minutes, initial encounter |
| 853.02 | Other and unspecified intracranial hemorrhage following injury without mention of open intracranial wound, with brief [less than one hour] loss of consciousness | S06.361A | Traumatic hemorrhage of cerebrum, unspecified, with loss of consciousness of 30 minutes or less, initial encounter |
| 853.02 | Other and unspecified intracranial hemorrhage following injury without mention of open intracranial wound, with brief [less than one hour] loss of consciousness | S06.362A | Traumatic hemorrhage of cerebrum, unspecified, with loss of consciousness of 31 minutes to 59 minutes, initial encounter |
| 853.03 | Other and unspecified intracranial hemorrhage following injury without mention of open intracranial wound, with moderate [1-24 hours] loss of consciousness | S06.343A | Traumatic hemorrhage of right cerebrum with loss of consciousness of 1 hours to 5 hours 59 minutes, initial encounter |
| 853.03 | Other and unspecified intracranial hemorrhage following injury without mention of open intracranial wound, with moderate [1-24 hours] loss of consciousness | S06.344A | Traumatic hemorrhage of right cerebrum with loss of consciousness of 6 hours to 24 hours, initial encounter |
| 853.03 | Other and unspecified intracranial hemorrhage following injury without mention of open intracranial wound, with moderate [1-24 hours] loss of consciousness | S06.353A | Traumatic hemorrhage of left cerebrum with loss of consciousness of 1 hours to 5 hours 59 minutes, initial encounter |
| 853.03 | Other and unspecified intracranial hemorrhage following injury without mention of open intracranial wound, with moderate [1-24 hours] loss of consciousness | S06.354A | Traumatic hemorrhage of left cerebrum with loss of consciousness of 6 hours to 24 hours, initial encounter |
| 853.03 | Other and unspecified intracranial hemorrhage following injury without mention of open intracranial wound, with moderate [1-24 hours] loss of consciousness | S06.363A | Traumatic hemorrhage of cerebrum, unspecified, with loss of consciousness of 1 hours to 5 hours 59 minutes, initial encounter |
| 853.03 | Other and unspecified intracranial hemorrhage following injury without mention of open intracranial wound, with moderate [1-24 hours] loss of consciousness | S06.364A | Traumatic hemorrhage of cerebrum, unspecified, with loss of consciousness of 6 hours to 24 hours, initial encounter |
| 853.04 | Other and unspecified intracranial hemorrhage following injury without mention of open intracranial wound, with prolonged [more than 24 hours] loss of consciousness and return to pre- existing conscious level | S06.345A | Traumatic hemorrhage of right cerebrum with loss of consciousness greater than 24 hours with return to pre-existing conscious level, initial encounter |
| 853.04 | Other and unspecified intracranial hemorrhage following injury without mention of open intracranial wound, with prolonged [more than 24 hours] loss of consciousness and return to pre- existing conscious level | S06.355A | Traumatic hemorrhage of left cerebrum with loss of consciousness greater than 24 hours with return to pre-existing conscious level, initial encounter |
| 853.04 | Other and unspecified intracranial hemorrhage following injury without mention of open intracranial wound, with prolonged [more than 24 hours] loss of consciousness and return to pre- existing conscious level | S06.365A | Traumatic hemorrhage of cerebrum, unspecified, with loss of consciousness greater than 24 hours with return to pre-existing conscious level, initial encounter |
| 853.05 | Other and unspecified intracranial hemorrhage following injury without mention of open intracranial wound, with prolonged [more than 24 hours] loss of consciousness without return to pre-existing conscious level | S06.346A | Traumatic hemorrhage of right cerebrum with loss of consciousness greater than 24 hours without return to pre-existing conscious level with patient surviving, initial encounter |
| 853.05 | Other and unspecified intracranial hemorrhage following injury without mention of open intracranial wound, with prolonged [more than 24 hours] loss of consciousness without return to pre-existing conscious level | S06.347A | Traumatic hemorrhage of right cerebrum with loss of consciousness of any duration with death due to brain injury prior to regaining consciousness, initial encounter |
| 853.05 | Other and unspecified intracranial hemorrhage following injury without mention of open intracranial wound, with prolonged [more than 24 hours] loss of consciousness without return to pre-existing conscious level | S06.348A | Traumatic hemorrhage of right cerebrum with loss of consciousness of any duration with death due to other cause prior to regaining consciousness, initial encounter |
| 853.05 | Other and unspecified intracranial hemorrhage following injury without mention of open intracranial wound, with prolonged [more than 24 hours] loss of consciousness without return to pre-existing conscious level | S06.356A | Traumatic hemorrhage of left cerebrum with loss of consciousness greater than 24 hours without return to pre-existing conscious level with patient surviving, initial encounter |
| 853.05 | Other and unspecified intracranial hemorrhage following injury without mention of open intracranial wound, with prolonged [more than 24 hours] loss of consciousness without return to pre-existing conscious level | S06.357A | Traumatic hemorrhage of left cerebrum with loss of consciousness of any duration with death due to brain injury prior to regaining consciousness, initial encounter |
| 853.05 | Other and unspecified intracranial hemorrhage following injury without mention of open intracranial wound, with prolonged [more than 24 hours] loss of consciousness without return to pre-existing conscious level | S06.358A | Traumatic hemorrhage of left cerebrum with loss of consciousness of any duration with death due to other cause prior to regaining consciousness, initial encounter |
| 853.05 | Other and unspecified intracranial hemorrhage following injury without mention of open intracranial wound, with prolonged [more than 24 hours] loss of consciousness without return to pre-existing conscious level | S06.366A | Traumatic hemorrhage of cerebrum, unspecified, with loss of consciousness greater than 24 hours without return to pre-existing conscious level with patient surviving, initial encounter |
| 853.05 | Other and unspecified intracranial hemorrhage following injury without mention of open intracranial wound, with prolonged [more than 24 hours] loss of consciousness without return to pre-existing conscious level | S06.367A | Traumatic hemorrhage of cerebrum, unspecified, with loss of consciousness of any duration with death due to brain injury prior to regaining consciousness, initial encounter |
| 853.05 | Other and unspecified intracranial hemorrhage following injury without mention of open intracranial wound, with prolonged [more than 24 hours] loss of consciousness without return to pre-existing conscious level | S06.368A | Traumatic hemorrhage of cerebrum, unspecified, with loss of consciousness of any duration with death due to other cause prior to regaining consciousness, initial encounter |
| 853.06 | Other and unspecified intracranial hemorrhage following injury without mention of open intracranial wound, with loss of consciousness of unspecified duration | S06.349A | Traumatic hemorrhage of right cerebrum with loss of consciousness of unspecified duration, initial encounter |
| 853.06 | Other and unspecified intracranial hemorrhage following injury without mention of open intracranial wound, with loss of consciousness of unspecified duration | S06.359A | Traumatic hemorrhage of left cerebrum with loss of consciousness of unspecified duration, initial encounter |
| 853.06 | Other and unspecified intracranial hemorrhage following injury without mention of open intracranial wound, with loss of consciousness of unspecified duration | S06.369A | Traumatic hemorrhage of cerebrum, unspecified, with loss of consciousness of unspecified duration, initial encounter |
| 853.09 | Other and unspecified intracranial hemorrhage following injury without mention of open intracranial wound, with concussion, unspecified | S06.360A | Traumatic hemorrhage of cerebrum, unspecified, without loss of consciousness, initial encounter |
| 853.09 | Other and unspecified intracranial hemorrhage following injury without mention of open intracranial wound, with concussion, unspecified | S06.369A | Traumatic hemorrhage of cerebrum, unspecified, with loss of consciousness of unspecified duration, initial encounter |
| 853.10 | Other and unspecified intracranial hemorrhage following injury with open intracranial wound, unspecified state of consciousness | S06.360A | Traumatic hemorrhage of cerebrum, unspecified, without loss of consciousness, initial encounter |
| 853.10 | Other and unspecified intracranial hemorrhage following injury with open intracranial wound, unspecified state of consciousness |  |  |
| 853.11 | Other and unspecified intracranial hemorrhage following injury with open intracranial wound, with no loss of consciousness | S06.360A | Traumatic hemorrhage of cerebrum, unspecified, without loss of consciousness, initial encounter |
| 853.11 | Other and unspecified intracranial hemorrhage following injury with open intracranial wound, with no loss of consciousness |  |  |
| 853.12 | Other and unspecified intracranial hemorrhage following injury with open intracranial wound, with brief [less than one hour] loss of consciousness | S06.361A | Traumatic hemorrhage of cerebrum, unspecified, with loss of consciousness of 30 minutes or less, initial encounter |
| 853.12 | Other and unspecified intracranial hemorrhage following injury with open intracranial wound, with brief [less than one hour] loss of consciousness | S06.362A | Traumatic hemorrhage of cerebrum, unspecified, with loss of consciousness of 31 minutes to 59 minutes, initial encounter |
| 853.12 | Other and unspecified intracranial hemorrhage following injury with open intracranial wound, with brief [less than one hour] loss of consciousness |  |  |
| 853.13 | Other and unspecified intracranial hemorrhage following injury with open intracranial wound, with moderate [1-24 hours] loss of consciousness | S06.363A | Traumatic hemorrhage of cerebrum, unspecified, with loss of consciousness of 1 hours to 5 hours 59 minutes, initial encounter |
| 853.13 | Other and unspecified intracranial hemorrhage following injury with open intracranial wound, with moderate [1-24 hours] loss of consciousness | S06.364A | Traumatic hemorrhage of cerebrum, unspecified, with loss of consciousness of 6 hours to 24 hours, initial encounter |
| 853.13 | Other and unspecified intracranial hemorrhage following injury with open intracranial wound, with moderate [1-24 hours] loss of consciousness |  |  |
| 853.14 | Other and unspecified intracranial hemorrhage following injury with open intracranial wound, with prolonged [more than 24 hours] loss of consciousness and return to pre-existing conscious level | S06.365A | Traumatic hemorrhage of cerebrum, unspecified, with loss of consciousness greater than 24 hours with return to pre-existing conscious level, initial encounter |
| 853.14 | Other and unspecified intracranial hemorrhage following injury with open intracranial wound, with prolonged [more than 24 hours] loss of consciousness and return to pre-existing conscious level |  |  |
| 853.15 | Other and unspecified intracranial hemorrhage following injury with open intracranial wound, with prolonged [more than 24 hours] loss of consciousness without return to pre-existing conscious level | S06.366A | Traumatic hemorrhage of cerebrum, unspecified, with loss of consciousness greater than 24 hours without return to pre-existing conscious level with patient surviving, initial encounter |
| 853.15 | Other and unspecified intracranial hemorrhage following injury with open intracranial wound, with prolonged [more than 24 hours] loss of consciousness without return to pre-existing conscious level | S06.367A | Traumatic hemorrhage of cerebrum, unspecified, with loss of consciousness of any duration with death due to brain injury prior to regaining consciousness, initial encounter |
| 853.15 | Other and unspecified intracranial hemorrhage following injury with open intracranial wound, with prolonged [more than 24 hours] loss of consciousness without return to pre-existing conscious level | S06.368A | Traumatic hemorrhage of cerebrum, unspecified, with loss of consciousness of any duration with death due to other cause prior to regaining consciousness, initial encounter |
| 853.15 | Other and unspecified intracranial hemorrhage following injury with open intracranial wound, with prolonged [more than 24 hours] loss of consciousness without return to pre-existing conscious level |  |  |
| 853.16 | Other and unspecified intracranial hemorrhage following injury with open intracranial wound, with loss of consciousness of unspecified duration | S06.369A | Traumatic hemorrhage of cerebrum, unspecified, with loss of consciousness of unspecified duration, initial encounter |
| 853.16 | Other and unspecified intracranial hemorrhage following injury with open intracranial wound, with loss of consciousness of unspecified duration |  |  |
| 853.19 | Other and unspecified intracranial hemorrhage following injury with open intracranial wound, with concussion, unspecified | S06.360A | Traumatic hemorrhage of cerebrum, unspecified, without loss of consciousness, initial encounter |
| 853.19 | Other and unspecified intracranial hemorrhage following injury with open intracranial wound, with concussion, unspecified | S06.369A | Traumatic hemorrhage of cerebrum, unspecified, with loss of consciousness of unspecified duration, initial encounter |
| 853.19 | Other and unspecified intracranial hemorrhage following injury with open intracranial wound, with concussion, unspecified |  |  |
| 854.00 | Intracranial injury of other and unspecified nature without mention of open intracranial wound, unspecified state of consciousness | S06.890A | Other specified intracranial injury without loss of consciousness, initial encounter |
| 854.01 | Intracranial injury of other and unspecified nature without mention of open intracranial wound, with no loss of consciousness | S06.1X0A | Traumatic cerebral edema without loss of consciousness, initial encounter |
| 854.01 | Intracranial injury of other and unspecified nature without mention of open intracranial wound, with no loss of consciousness | S06.2X0A | Diffuse traumatic brain injury without loss of consciousness, initial encounter |
| 854.01 | Intracranial injury of other and unspecified nature without mention of open intracranial wound, with no loss of consciousness | S06.300A | Unspecified focal traumatic brain injury without loss of consciousness, initial encounter |
| 854.01 | Intracranial injury of other and unspecified nature without mention of open intracranial wound, with no loss of consciousness | S06.810A | Injury of right internal carotid artery, intracranial portion, not elsewhere classified without loss of consciousness, initial encounter |
| 854.01 | Intracranial injury of other and unspecified nature without mention of open intracranial wound, with no loss of consciousness | S06.820A | Injury of left internal carotid artery, intracranial portion, not elsewhere classified without loss of consciousness, initial encounter |
| 854.01 | Intracranial injury of other and unspecified nature without mention of open intracranial wound, with no loss of consciousness | S06.890A | Other specified intracranial injury without loss of consciousness, initial encounter |
| 854.01 | Intracranial injury of other and unspecified nature without mention of open intracranial wound, with no loss of consciousness | S06.9X0A | Unspecified intracranial injury without loss of consciousness, initial encounter |
| 854.02 | Intracranial injury of other and unspecified nature without mention of open intracranial wound, with brief [less than one hour] loss of consciousness | S06.1X1A | Traumatic cerebral edema with loss of consciousness of 30 minutes or less, initial encounter |
| 854.02 | Intracranial injury of other and unspecified nature without mention of open intracranial wound, with brief [less than one hour] loss of consciousness | S06.1X2A | Traumatic cerebral edema with loss of consciousness of 31 minutes to 59 minutes, initial encounter |
| 854.02 | Intracranial injury of other and unspecified nature without mention of open intracranial wound, with brief [less than one hour] loss of consciousness | S06.2X1A | Diffuse traumatic brain injury with loss of consciousness of 30 minutes or less, initial encounter |
| 854.02 | Intracranial injury of other and unspecified nature without mention of open intracranial wound, with brief [less than one hour] loss of consciousness | S06.2X2A | Diffuse traumatic brain injury with loss of consciousness of 31 minutes to 59 minutes, initial encounter |
| 854.02 | Intracranial injury of other and unspecified nature without mention of open intracranial wound, with brief [less than one hour] loss of consciousness | S06.301A | Unspecified focal traumatic brain injury with loss of consciousness of 30 minutes or less, initial encounter |
| 854.02 | Intracranial injury of other and unspecified nature without mention of open intracranial wound, with brief [less than one hour] loss of consciousness | S06.302A | Unspecified focal traumatic brain injury with loss of consciousness of 31 minutes to 59 minutes, initial encounter |
| 854.02 | Intracranial injury of other and unspecified nature without mention of open intracranial wound, with brief [less than one hour] loss of consciousness | S06.811A | Injury of right internal carotid artery, intracranial portion, not elsewhere classified with loss of consciousness of 30 minutes or less, initial encounter |
| 854.02 | Intracranial injury of other and unspecified nature without mention of open intracranial wound, with brief [less than one hour] loss of consciousness | S06.812A | Injury of right internal carotid artery, intracranial portion, not elsewhere classified with loss of consciousness of 31 minutes to 59 minutes, initial encounter |
| 854.02 | Intracranial injury of other and unspecified nature without mention of open intracranial wound, with brief [less than one hour] loss of consciousness | S06.821A | Injury of left internal carotid artery, intracranial portion, not elsewhere classified with loss of consciousness of 30 minutes or less, initial encounter |
| 854.02 | Intracranial injury of other and unspecified nature without mention of open intracranial wound, with brief [less than one hour] loss of consciousness | S06.822A | Injury of left internal carotid artery, intracranial portion, not elsewhere classified with loss of consciousness of 31 minutes to 59 minutes, initial encounter |
| 854.02 | Intracranial injury of other and unspecified nature without mention of open intracranial wound, with brief [less than one hour] loss of consciousness | S06.891A | Other specified intracranial injury with loss of consciousness of 30 minutes or less, initial encounter |
| 854.02 | Intracranial injury of other and unspecified nature without mention of open intracranial wound, with brief [less than one hour] loss of consciousness | S06.892A | Other specified intracranial injury with loss of consciousness of 31 minutes to 59 minutes, initial encounter |
| 854.02 | Intracranial injury of other and unspecified nature without mention of open intracranial wound, with brief [less than one hour] loss of consciousness | S06.9X1A | Unspecified intracranial injury with loss of consciousness of 30 minutes or less, initial encounter |
| 854.02 | Intracranial injury of other and unspecified nature without mention of open intracranial wound, with brief [less than one hour] loss of consciousness | S06.9X2A | Unspecified intracranial injury with loss of consciousness of 31 minutes to 59 minutes, initial encounter |
| 854.03 | Intracranial injury of other and unspecified nature without mention of open intracranial wound, with moderate [1-24 hours] loss of consciousness | S06.1X3A | Traumatic cerebral edema with loss of consciousness of 1 hour to 5 hours 59 minutes, initial encounter |
| 854.03 | Intracranial injury of other and unspecified nature without mention of open intracranial wound, with moderate [1-24 hours] loss of consciousness | S06.1X4A | Traumatic cerebral edema with loss of consciousness of 6 hours to 24 hours, initial encounter |
| 854.03 | Intracranial injury of other and unspecified nature without mention of open intracranial wound, with moderate [1-24 hours] loss of consciousness | S06.2X3A | Diffuse traumatic brain injury with loss of consciousness of 1 hour to 5 hours 59 minutes, initial encounter |
| 854.03 | Intracranial injury of other and unspecified nature without mention of open intracranial wound, with moderate [1-24 hours] loss of consciousness | S06.2X4A | Diffuse traumatic brain injury with loss of consciousness of 6 hours to 24 hours, initial encounter |
| 854.03 | Intracranial injury of other and unspecified nature without mention of open intracranial wound, with moderate [1-24 hours] loss of consciousness | S06.303A | Unspecified focal traumatic brain injury with loss of consciousness of 1 hour to 5 hours 59 minutes, initial encounter |
| 854.03 | Intracranial injury of other and unspecified nature without mention of open intracranial wound, with moderate [1-24 hours] loss of consciousness | S06.304A | Unspecified focal traumatic brain injury with loss of consciousness of 6 hours to 24 hours, initial encounter |
| 854.03 | Intracranial injury of other and unspecified nature without mention of open intracranial wound, with moderate [1-24 hours] loss of consciousness | S06.813A | Injury of right internal carotid artery, intracranial portion, not elsewhere classified with loss of consciousness of 1 hour to 5 hours 59 minutes, initial encounter |
| 854.03 | Intracranial injury of other and unspecified nature without mention of open intracranial wound, with moderate [1-24 hours] loss of consciousness | S06.814A | Injury of right internal carotid artery, intracranial portion, not elsewhere classified with loss of consciousness of 6 hours to 24 hours, initial encounter |
| 854.03 | Intracranial injury of other and unspecified nature without mention of open intracranial wound, with moderate [1-24 hours] loss of consciousness | S06.823A | Injury of left internal carotid artery, intracranial portion, not elsewhere classified with loss of consciousness of 1 hour to 5 hours 59 minutes, initial encounter |
| 854.03 | Intracranial injury of other and unspecified nature without mention of open intracranial wound, with moderate [1-24 hours] loss of consciousness | S06.824A | Injury of left internal carotid artery, intracranial portion, not elsewhere classified with loss of consciousness of 6 hours to 24 hours, initial encounter |
| 854.03 | Intracranial injury of other and unspecified nature without mention of open intracranial wound, with moderate [1-24 hours] loss of consciousness | S06.893A | Other specified intracranial injury with loss of consciousness of 1 hour to 5 hours 59 minutes, initial encounter |
| 854.03 | Intracranial injury of other and unspecified nature without mention of open intracranial wound, with moderate [1-24 hours] loss of consciousness | S06.894A | Other specified intracranial injury with loss of consciousness of 6 hours to 24 hours, initial encounter |
| 854.03 | Intracranial injury of other and unspecified nature without mention of open intracranial wound, with moderate [1-24 hours] loss of consciousness | S06.9X3A | Unspecified intracranial injury with loss of consciousness of 1 hour to 5 hours 59 minutes, initial encounter |
| 854.03 | Intracranial injury of other and unspecified nature without mention of open intracranial wound, with moderate [1-24 hours] loss of consciousness | S06.9X4A | Unspecified intracranial injury with loss of consciousness of 6 hours to 24 hours, initial encounter |
| 854.04 | Intracranial injury of other and unspecified nature without mention of open intracranial wound, with prolonged [more than 24 hours] loss of consciousness and return to pre-existing conscious level | S06.1X5A | Traumatic cerebral edema with loss of consciousness greater than 24 hours with return to pre-existing conscious level, initial encounter |
| 854.04 | Intracranial injury of other and unspecified nature without mention of open intracranial wound, with prolonged [more than 24 hours] loss of consciousness and return to pre-existing conscious level | S06.2X5A | Diffuse traumatic brain injury with loss of consciousness greater than 24 hours with return to pre-existing conscious levels, initial encounter |
| 854.04 | Intracranial injury of other and unspecified nature without mention of open intracranial wound, with prolonged [more than 24 hours] loss of consciousness and return to pre-existing conscious level | S06.305A | Unspecified focal traumatic brain injury with loss of consciousness greater than 24 hours with return to pre-existing conscious level, initial encounter |
| 854.04 | Intracranial injury of other and unspecified nature without mention of open intracranial wound, with prolonged [more than 24 hours] loss of consciousness and return to pre-existing conscious level | S06.815A | Injury of right internal carotid artery, intracranial portion, not elsewhere classified with loss of consciousness greater than 24 hours with return to pre-existing conscious level, initial encounter |
| 854.04 | Intracranial injury of other and unspecified nature without mention of open intracranial wound, with prolonged [more than 24 hours] loss of consciousness and return to pre-existing conscious level | S06.825A | Injury of left internal carotid artery, intracranial portion, not elsewhere classified with loss of consciousness greater than 24 hours with return to pre-existing conscious level, initial encounter |
| 854.04 | Intracranial injury of other and unspecified nature without mention of open intracranial wound, with prolonged [more than 24 hours] loss of consciousness and return to pre-existing conscious level | S06.895A | Other specified intracranial injury with loss of consciousness greater than 24 hours with return to pre-existing conscious level, initial encounter |
| 854.04 | Intracranial injury of other and unspecified nature without mention of open intracranial wound, with prolonged [more than 24 hours] loss of consciousness and return to pre-existing conscious level | S06.9X5A | Unspecified intracranial injury with loss of consciousness greater than 24 hours with return to pre-existing conscious level, initial encounter |
| 854.05 | Intracranial injury of other and unspecified nature without mention of open intracranial wound, with prolonged [more than 24 hours] loss of consciousness without return to pre-existing conscious level | S06.1X6A | Traumatic cerebral edema with loss of consciousness greater than 24 hours without return to pre-existing conscious level with patient surviving, initial encounter |
| 854.05 | Intracranial injury of other and unspecified nature without mention of open intracranial wound, with prolonged [more than 24 hours] loss of consciousness without return to pre-existing conscious level | S06.1X7A | Traumatic cerebral edema with loss of consciousness of any duration with death due to brain injury prior to regaining consciousness, initial encounter |
| 854.05 | Intracranial injury of other and unspecified nature without mention of open intracranial wound, with prolonged [more than 24 hours] loss of consciousness without return to pre-existing conscious level | S06.1X8A | Traumatic cerebral edema with loss of consciousness of any duration with death due to other cause prior to regaining consciousness, initial encounter |
| 854.05 | Intracranial injury of other and unspecified nature without mention of open intracranial wound, with prolonged [more than 24 hours] loss of consciousness without return to pre-existing conscious level | S06.2X6A | Diffuse traumatic brain injury with loss of consciousness greater than 24 hours without return to pre-existing conscious level with patient surviving, initial encounter |
| 854.05 | Intracranial injury of other and unspecified nature without mention of open intracranial wound, with prolonged [more than 24 hours] loss of consciousness without return to pre-existing conscious level | S06.2X7A | Diffuse traumatic brain injury with loss of consciousness of any duration with death due to brain injury prior to regaining consciousness, initial encounter |
| 854.05 | Intracranial injury of other and unspecified nature without mention of open intracranial wound, with prolonged [more than 24 hours] loss of consciousness without return to pre-existing conscious level | S06.2X8A | Diffuse traumatic brain injury with loss of consciousness of any duration with death due to other cause prior to regaining consciousness, initial encounter |
| 854.05 | Intracranial injury of other and unspecified nature without mention of open intracranial wound, with prolonged [more than 24 hours] loss of consciousness without return to pre-existing conscious level | S06.306A | Unspecified focal traumatic brain injury with loss of consciousness greater than 24 hours without return to pre-existing conscious level with patient surviving, initial encounter |
| 854.05 | Intracranial injury of other and unspecified nature without mention of open intracranial wound, with prolonged [more than 24 hours] loss of consciousness without return to pre-existing conscious level | S06.307A | Unspecified focal traumatic brain injury with loss of consciousness of any duration with death due to brain injury prior to regaining consciousness, initial encounter |
| 854.05 | Intracranial injury of other and unspecified nature without mention of open intracranial wound, with prolonged [more than 24 hours] loss of consciousness without return to pre-existing conscious level | S06.308A | Unspecified focal traumatic brain injury with loss of consciousness of any duration with death due to other cause prior to regaining consciousness, initial encounter |
| 854.05 | Intracranial injury of other and unspecified nature without mention of open intracranial wound, with prolonged [more than 24 hours] loss of consciousness without return to pre-existing conscious level | S06.816A | Injury of right internal carotid artery, intracranial portion, not elsewhere classified with loss of consciousness greater than 24 hours without return to pre-existing conscious level with patient surviving, initial encounter |
| 854.05 | Intracranial injury of other and unspecified nature without mention of open intracranial wound, with prolonged [more than 24 hours] loss of consciousness without return to pre-existing conscious level | S06.817A | Injury of right internal carotid artery, intracranial portion, not elsewhere classified with loss of consciousness of any duration with death due to brain injury prior to regaining consciousness, initial encounter |
| 854.05 | Intracranial injury of other and unspecified nature without mention of open intracranial wound, with prolonged [more than 24 hours] loss of consciousness without return to pre-existing conscious level | S06.818A | Injury of right internal carotid artery, intracranial portion, not elsewhere classified with loss of consciousness of any duration with death due to other cause prior to regaining consciousness, initial encounter |
| 854.05 | Intracranial injury of other and unspecified nature without mention of open intracranial wound, with prolonged [more than 24 hours] loss of consciousness without return to pre-existing conscious level | S06.826A | Injury of left internal carotid artery, intracranial portion, not elsewhere classified with loss of consciousness greater than 24 hours without return to pre-existing conscious level with patient surviving, initial encounter |
| 854.05 | Intracranial injury of other and unspecified nature without mention of open intracranial wound, with prolonged [more than 24 hours] loss of consciousness without return to pre-existing conscious level | S06.827A | Injury of left internal carotid artery, intracranial portion, not elsewhere classified with loss of consciousness of any duration with death due to brain injury prior to regaining consciousness, initial encounter |
| 854.05 | Intracranial injury of other and unspecified nature without mention of open intracranial wound, with prolonged [more than 24 hours] loss of consciousness without return to pre-existing conscious level | S06.828A | Injury of left internal carotid artery, intracranial portion, not elsewhere classified with loss of consciousness of any duration with death due to other cause prior to regaining consciousness, initial encounter |
| 854.05 | Intracranial injury of other and unspecified nature without mention of open intracranial wound, with prolonged [more than 24 hours] loss of consciousness without return to pre-existing conscious level | S06.896A | Other specified intracranial injury with loss of consciousness greater than 24 hours without return to pre-existing conscious level with patient surviving, initial encounter |
| 854.05 | Intracranial injury of other and unspecified nature without mention of open intracranial wound, with prolonged [more than 24 hours] loss of consciousness without return to pre-existing conscious level | S06.897A | Other specified intracranial injury with loss of consciousness of any duration with death due to brain injury prior to regaining consciousness, initial encounter |
| 854.05 | Intracranial injury of other and unspecified nature without mention of open intracranial wound, with prolonged [more than 24 hours] loss of consciousness without return to pre-existing conscious level | S06.898A | Other specified intracranial injury with loss of consciousness of any duration with death due to other cause prior to regaining consciousness, initial encounter |
| 854.05 | Intracranial injury of other and unspecified nature without mention of open intracranial wound, with prolonged [more than 24 hours] loss of consciousness without return to pre-existing conscious level | S06.9X6A | Unspecified intracranial injury with loss of consciousness greater than 24 hours without return to pre-existing conscious level with patient surviving, initial encounter |
| 854.05 | Intracranial injury of other and unspecified nature without mention of open intracranial wound, with prolonged [more than 24 hours] loss of consciousness without return to pre-existing conscious level | S06.9X7A | Unspecified intracranial injury with loss of consciousness of any duration with death due to brain injury prior to regaining consciousness, initial encounter |
| 854.05 | Intracranial injury of other and unspecified nature without mention of open intracranial wound, with prolonged [more than 24 hours] loss of consciousness without return to pre-existing conscious level | S06.9X8A | Unspecified intracranial injury with loss of consciousness of any duration with death due to other cause prior to regaining consciousness, initial encounter |
| 854.06 | Intracranial injury of other and unspecified nature without mention of open intracranial wound, with loss of consciousness of unspecified duration | S06.1X9A | Traumatic cerebral edema with loss of consciousness of unspecified duration, initial encounter |
| 854.06 | Intracranial injury of other and unspecified nature without mention of open intracranial wound, with loss of consciousness of unspecified duration | S06.2X9A | Diffuse traumatic brain injury with loss of consciousness of unspecified duration, initial encounter |
| 854.06 | Intracranial injury of other and unspecified nature without mention of open intracranial wound, with loss of consciousness of unspecified duration | S06.309A | Unspecified focal traumatic brain injury with loss of consciousness of unspecified duration, initial encounter |
| 854.06 | Intracranial injury of other and unspecified nature without mention of open intracranial wound, with loss of consciousness of unspecified duration | S06.819A | Injury of right internal carotid artery, intracranial portion, not elsewhere classified with loss of consciousness of unspecified duration, initial encounter |
| 854.06 | Intracranial injury of other and unspecified nature without mention of open intracranial wound, with loss of consciousness of unspecified duration | S06.829A | Injury of left internal carotid artery, intracranial portion, not elsewhere classified with loss of consciousness of unspecified duration, initial encounter |
| 854.06 | Intracranial injury of other and unspecified nature without mention of open intracranial wound, with loss of consciousness of unspecified duration | S06.899A | Other specified intracranial injury with loss of consciousness of unspecified duration, initial encounter |
| 854.06 | Intracranial injury of other and unspecified nature without mention of open intracranial wound, with loss of consciousness of unspecified duration | S06.9X9A | Unspecified intracranial injury with loss of consciousness of unspecified duration, initial encounter |
| 854.09 | Intracranial injury of other and unspecified nature without mention of open intracranial wound, with concussion, unspecified | S06.890A | Other specified intracranial injury without loss of consciousness, initial encounter |
| 854.09 | Intracranial injury of other and unspecified nature without mention of open intracranial wound, with concussion, unspecified |  |  |
| 854.10 | Intracranial injury of other and unspecified nature with open intracranial wound, unspecified state of consciousness | S06.890A | Other specified intracranial injury without loss of consciousness, initial encounter |
| 854.10 | Intracranial injury of other and unspecified nature with open intracranial wound, unspecified state of consciousness |  |  |
| 854.11 | Intracranial injury of other and unspecified nature with open intracranial wound, with no loss of consciousness | S06.890A | Other specified intracranial injury without loss of consciousness, initial encounter |
| 854.11 | Intracranial injury of other and unspecified nature with open intracranial wound, with no loss of consciousness |  |  |
| 854.12 | Intracranial injury of other and unspecified nature with open intracranial wound, with brief [less than one hour] loss of consciousness | S06.891A | Other specified intracranial injury with loss of consciousness of 30 minutes or less, initial encounter |
| 854.12 | Intracranial injury of other and unspecified nature with open intracranial wound, with brief [less than one hour] loss of consciousness | S06.892A | Other specified intracranial injury with loss of consciousness of 31 minutes to 59 minutes, initial encounter |
| 854.12 | Intracranial injury of other and unspecified nature with open intracranial wound, with brief [less than one hour] loss of consciousness |  |  |
| 854.13 | Intracranial injury of other and unspecified nature with open intracranial wound, with moderate [1-24 hours] loss of consciousness | S06.893A | Other specified intracranial injury with loss of consciousness of 1 hour to 5 hours 59 minutes, initial encounter |
| 854.13 | Intracranial injury of other and unspecified nature with open intracranial wound, with moderate [1-24 hours] loss of consciousness | S06.894A | Other specified intracranial injury with loss of consciousness of 6 hours to 24 hours, initial encounter |
| 854.13 | Intracranial injury of other and unspecified nature with open intracranial wound, with moderate [1-24 hours] loss of consciousness |  |  |
| 854.14 | Intracranial injury of other and unspecified nature with open intracranial wound, with prolonged [more than 24 hours] loss of consciousness and return to pre-existing conscious level | S06.895A | Other specified intracranial injury with loss of consciousness greater than 24 hours with return to pre-existing conscious level, initial encounter |
| 854.14 | Intracranial injury of other and unspecified nature with open intracranial wound, with prolonged [more than 24 hours] loss of consciousness and return to pre-existing conscious level |  |  |
| 854.15 | Intracranial injury of other and unspecified nature with open intracranial wound, with prolonged [more than 24 hours] loss of consciousness without return to pre-existing conscious level | S06.896A | Other specified intracranial injury with loss of consciousness greater than 24 hours without return to pre-existing conscious level with patient surviving, initial encounter |
| 854.15 | Intracranial injury of other and unspecified nature with open intracranial wound, with prolonged [more than 24 hours] loss of consciousness without return to pre-existing conscious level | S06.897A | Other specified intracranial injury with loss of consciousness of any duration with death due to brain injury prior to regaining consciousness, initial encounter |
| 854.15 | Intracranial injury of other and unspecified nature with open intracranial wound, with prolonged [more than 24 hours] loss of consciousness without return to pre-existing conscious level | S06.898A | Other specified intracranial injury with loss of consciousness of any duration with death due to other cause prior to regaining consciousness, initial encounter |
| 854.15 | Intracranial injury of other and unspecified nature with open intracranial wound, with prolonged [more than 24 hours] loss of consciousness without return to pre-existing conscious level |  |  |
| 854.16 | Intracranial injury of other and unspecified nature with open intracranial wound, with loss of consciousness of unspecified duration | S06.899A | Other specified intracranial injury with loss of consciousness of unspecified duration, initial encounter |
| 854.16 | Intracranial injury of other and unspecified nature with open intracranial wound, with loss of consciousness of unspecified duration |  |  |
| 854.19 | Intracranial injury of other and unspecified nature with open intracranial wound, with concussion, unspecified | S06.890A | Other specified intracranial injury without loss of consciousness, initial encounter |
| 854.19 | Intracranial injury of other and unspecified nature with open intracranial wound, with concussion, unspecified | S06.899A | Other specified intracranial injury with loss of consciousness of unspecified duration, initial encounter |
| 854.19 | Intracranial injury of other and unspecified nature with open intracranial wound, with concussion, unspecified |  |  |
| 950.0 | Optic nerve injury | S04.011A | Injury of optic nerve, right eye, initial encounter |
| 950.0 | Optic nerve injury | S04.012A | Injury of optic nerve, left eye, initial encounter |
| 950.0 | Optic nerve injury | S04.019A | Injury of optic nerve, unspecified eye, initial encounter |
| 950.2 | Injury to optic pathways | S04.031A | Injury of optic tract and pathways, right side, initial encounter |
| 950.2 | Injury to optic pathways | S04.032A | Injury of optic tract and pathways, left side, initial encounter |
| 950.2 | Injury to optic pathways | S04.039A | Injury of optic tract and pathways, unspecified side, initial encounter |
| 950.3 | Injury to visual cortex | S04.041A | Injury of visual cortex, right side, initial encounter |
| 950.3 | Injury to visual cortex | S04.042A | Injury of visual cortex, left side, initial encounter |
| 950.3 | Injury to visual cortex | S04.049A | Injury of visual cortex, unspecified side, initial encounter |
| 950.9 | Injury to unspecified optic nerve and pathways | S04.019A | Injury of optic nerve, unspecified eye, initial encounter |
| 997.02 | Iatrogenic cerebrovascular infarction or hemorrhage | G97.31 | Intraoperative hemorrhage and hematoma of a nervous system organ or structure complicating a nervous system procedure |
| 997.02 | Iatrogenic cerebrovascular infarction or hemorrhage | G97.32 | Intraoperative hemorrhage and hematoma of a nervous system organ or structure complicating other procedure |
| 997.02 | Iatrogenic cerebrovascular infarction or hemorrhage | I97.810 | Intraoperative cerebrovascular infarction during cardiac surgery |
| 997.02 | Iatrogenic cerebrovascular infarction or hemorrhage | I97.811 | Intraoperative cerebrovascular infarction during other surgery |
| 997.02 | Iatrogenic cerebrovascular infarction or hemorrhage | I97.820 | Postprocedural cerebrovascular infarction following cardiac surgery |
| 997.02 | Iatrogenic cerebrovascular infarction or hemorrhage | I97.821 | Postprocedural cerebrovascular infarction following other surgery |
| 998.11 | Hemorrhage complicating a procedure | G97.31 | Intraoperative hemorrhage and hematoma of a nervous system organ or structure complicating a nervous system procedure |
| 998.11 | Hemorrhage complicating a procedure | G97.32 | Intraoperative hemorrhage and hematoma of a nervous system organ or structure complicating other procedure |
| 998.11 | Hemorrhage complicating a procedure | G97.51 | Postprocedural hemorrhage of a nervous system organ or structure following a nervous system procedure |
| 998.11 | Hemorrhage complicating a procedure | G97.52 | Postprocedural hemorrhage of a nervous system organ or structure following other procedure |
| 998.12 | Hematoma complicating a procedure | G97.31 | Intraoperative hemorrhage and hematoma of a nervous system organ or structure complicating a nervous system procedure |
| 998.12 | Hematoma complicating a procedure | G97.32 | Intraoperative hemorrhage and hematoma of a nervous system organ or structure complicating other procedure |
| 998.12 | Hematoma complicating a procedure | G97.61 | Postprocedural hematoma of a nervous system organ or structure following a nervous system procedure |
| 998.12 | Hematoma complicating a procedure | G97.62 | Postprocedural hematoma of a nervous system organ or structure following other procedure |
| V12.54 | Personal history of transient ischemic attack (TIA), and cerebral infarction without residual deficits | Z86.73 | Personal history of transient ischemic attack (TIA), and cerebral infarction without residual deficits |
| V15.52 | Personal history of traumatic brain injury | Z87.820 | Personal history of traumatic brain injury |
| V41.6 | Problems with swallowing and mastication | R13.10 | Dysphagia, unspecified |

**ICD/CPT procedure codes or procedure-related diagnosis codes**

**Neurovascular Procedure**

| **Procedure Code** | **Description** |
| --- | --- |
| 39.51 | Clipping of aneurysm |
| 39.52 | Other repair of aneurysm |
| 39.53 | Repair of arteriovenous fistula |
| 39.72 | Endovascular (total) embolization or occlusion of head and neck vessels |
| 39.75 | Endovascular embolization or occlusion of vessel(s) of head or neck using bare coils |
| 39.76 | Endovascular embolization or occlusion of vessel(s) of head or neck using bioactive coils |
| 39.79 | Other endovascular procedures on other vessels |
| 03LG3DZ | Occlusion of Intracranial Artery with Intraluminal Device, Percutaneous Approach |
| 03LK3DZ | Occlusion of Right Internal Carotid Artery with Intraluminal Device, Percutaneous Approach |
| 03LL3DZ | Occlusion of Left Internal Carotid Artery with Intraluminal Device, Percutaneous Approach |
| 03LP3DZ | Occlusion of Right Vertebral Artery with Intraluminal Device, Percutaneous Approach |
| 03LQ3DZ | Occlusion of Left Vertebral Artery with Intraluminal Device, Percutaneous Approach |
| 03VG0BZ | Restriction of Intracranial Artery with Bioactive Intraluminal Device, Open Approach |
| 03VG0CZ | Restriction of Intracranial Artery with Extraluminal Device, Open Approach |
| 03VG0DZ | Restriction of Intracranial Artery with Intraluminal Device, Open Approach |
| 03VG0ZZ | Restriction of Intracranial Artery, Open Approach |
| 03VG3BZ | Restriction of Intracranial Artery with Bioactive Intraluminal Device, Percutaneous Approach |
| 03VG3CZ | Restriction of Intracranial Artery with Extraluminal Device, Percutaneous Approach |
| 03VG3DZ | Restriction of Intracranial Artery with Intraluminal Device, Percutaneous Approach |
| 03VG3ZZ | Restriction of Intracranial Artery, Percutaneous Approach |
| 03VG4BZ | Restriction of Intracranial Artery with Bioactive Intraluminal Device, Percutaneous Endoscopic Approach |
| 03VG4CZ | Restriction of Intracranial Artery with Extraluminal Device, Percutaneous Endoscopic Approach |
| 03VG4DZ | Restriction of Intracranial Artery with Intraluminal Device, Percutaneous Endoscopic Approach |
| 03VG4ZZ | Restriction of Intracranial Artery, Percutaneous Endoscopic Approach |
| 03VH0BZ | Restriction of Right Common Carotid Artery with Bioactive Intraluminal Device, Open Approach |
| 03VH0CZ | Restriction of Right Common Carotid Artery with Extraluminal Device, Open Approach |
| 03VH0DZ | Restriction of Right Common Carotid Artery with Intraluminal Device, Open Approach |
| 03VH0ZZ | Restriction of Right Common Carotid Artery, Open Approach |
| 03VH3BZ | Restriction of Right Common Carotid Artery with Bioactive Intraluminal Device, Percutaneous Approach |
| 03VH3CZ | Restriction of Right Common Carotid Artery with Extraluminal Device, Percutaneous Approach |
| 03VH3DZ | Restriction of Right Common Carotid Artery with Intraluminal Device, Percutaneous Approach |
| 03VH3ZZ | Restriction of Right Common Carotid Artery, Percutaneous Approach |
| 03VH4BZ | Restriction of Right Common Carotid Artery with Bioactive Intraluminal Device, Percutaneous Endoscopic Approach |
| 03VH4CZ | Restriction of Right Common Carotid Artery with Extraluminal Device, Percutaneous Endoscopic Approach |
| 03VH4DZ | Restriction of Right Common Carotid Artery with Intraluminal Device, Percutaneous Endoscopic Approach |
| 03VH4ZZ | Restriction of Right Common Carotid Artery, Percutaneous Endoscopic Approach |
| 03VJ0BZ | Restriction of Left Common Carotid Artery with Bioactive Intraluminal Device, Open Approach |
| 03VJ0CZ | Restriction of Left Common Carotid Artery with Extraluminal Device, Open Approach |
| 03VJ0DZ | Restriction of Left Common Carotid Artery with Intraluminal Device, Open Approach |
| 03VJ0ZZ | Restriction of Left Common Carotid Artery, Open Approach |
| 03VJ3BZ | Restriction of Left Common Carotid Artery with Bioactive Intraluminal Device, Percutaneous Approach |
| 03VJ3CZ | Restriction of Left Common Carotid Artery with Extraluminal Device, Percutaneous Approach |
| 03VJ3DZ | Restriction of Left Common Carotid Artery with Intraluminal Device, Percutaneous Approach |
| 03VJ3ZZ | Restriction of Left Common Carotid Artery, Percutaneous Approach |
| 03VJ4BZ | Restriction of Left Common Carotid Artery with Bioactive Intraluminal Device, Percutaneous Endoscopic Approach |
| 03VJ4CZ | Restriction of Left Common Carotid Artery with Extraluminal Device, Percutaneous Endoscopic Approach |
| 03VJ4DZ | Restriction of Left Common Carotid Artery with Intraluminal Device, Percutaneous Endoscopic Approach |
| 03VJ4ZZ | Restriction of Left Common Carotid Artery, Percutaneous Endoscopic Approach |
| 03VK0BZ | Restriction of Right Internal Carotid Artery with Bioactive Intraluminal Device, Open Approach |
| 03VK0CZ | Restriction of Right Internal Carotid Artery with Extraluminal Device, Open Approach |
| 03VK0DZ | Restriction of Right Internal Carotid Artery with Intraluminal Device, Open Approach |
| 03VK0ZZ | Restriction of Right Internal Carotid Artery, Open Approach |
| 03VK3BZ | Restriction of Right Internal Carotid Artery with Bioactive Intraluminal Device, Percutaneous Approach |
| 03VK3CZ | Restriction of Right Internal Carotid Artery with Extraluminal Device, Percutaneous Approach |
| 03VK3DZ | Restriction of Right Internal Carotid Artery with Intraluminal Device, Percutaneous Approach |
| 03VK3ZZ | Restriction of Right Internal Carotid Artery, Percutaneous Approach |
| 03VK4BZ | Restriction of Right Internal Carotid Artery with Bioactive Intraluminal Device, Percutaneous Endoscopic Approach |
| 03VK4CZ | Restriction of Right Internal Carotid Artery with Extraluminal Device, Percutaneous Endoscopic Approach |
| 03VK4DZ | Restriction of Right Internal Carotid Artery with Intraluminal Device, Percutaneous Endoscopic Approach |
| 03VK4ZZ | Restriction of Right Internal Carotid Artery, Percutaneous Endoscopic Approach |
| 03VL0BZ | Restriction of Left Internal Carotid Artery with Bioactive Intraluminal Device, Open Approach |
| 03VL0CZ | Restriction of Left Internal Carotid Artery with Extraluminal Device, Open Approach |
| 03VL0DZ | Restriction of Left Internal Carotid Artery with Intraluminal Device, Open Approach |
| 03VL0ZZ | Restriction of Left Internal Carotid Artery, Open Approach |
| 03VL3BZ | Restriction of Left Internal Carotid Artery with Bioactive Intraluminal Device, Percutaneous Approach |
| 03VL3CZ | Restriction of Left Internal Carotid Artery with Extraluminal Device, Percutaneous Approach |
| 03VL3DZ | Restriction of Left Internal Carotid Artery with Intraluminal Device, Percutaneous Approach |
| 03VL3ZZ | Restriction of Left Internal Carotid Artery, Percutaneous Approach |
| 03VL4BZ | Restriction of Left Internal Carotid Artery with Bioactive Intraluminal Device, Percutaneous Endoscopic Approach |
| 03VL4CZ | Restriction of Left Internal Carotid Artery with Extraluminal Device, Percutaneous Endoscopic Approach |
| 03VL4DZ | Restriction of Left Internal Carotid Artery with Intraluminal Device, Percutaneous Endoscopic Approach |
| 03VL4ZZ | Restriction of Left Internal Carotid Artery, Percutaneous Endoscopic Approach |
| 03VM0BZ | Restriction of Right External Carotid Artery with Bioactive Intraluminal Device, Open Approach |
| 03VM0CZ | Restriction of Right External Carotid Artery with Extraluminal Device, Open Approach |
| 03VM0DZ | Restriction of Right External Carotid Artery with Intraluminal Device, Open Approach |
| 03VM0ZZ | Restriction of Right External Carotid Artery, Open Approach |
| 03VM3BZ | Restriction of Right External Carotid Artery with Bioactive Intraluminal Device, Percutaneous Approach |
| 03VM3CZ | Restriction of Right External Carotid Artery with Extraluminal Device, Percutaneous Approach |
| 03VM3DZ | Restriction of Right External Carotid Artery with Intraluminal Device, Percutaneous Approach |
| 03VM3ZZ | Restriction of Right External Carotid Artery, Percutaneous Approach |
| 03VM4BZ | Restriction of Right External Carotid Artery with Bioactive Intraluminal Device, Percutaneous Endoscopic Approach |
| 03VM4CZ | Restriction of Right External Carotid Artery with Extraluminal Device, Percutaneous Endoscopic Approach |
| 03VM4DZ | Restriction of Right External Carotid Artery with Intraluminal Device, Percutaneous Endoscopic Approach |
| 03VM4ZZ | Restriction of Right External Carotid Artery, Percutaneous Endoscopic Approach |
| 03VN0BZ | Restriction of Left External Carotid Artery with Bioactive Intraluminal Device, Open Approach |
| 03VN0CZ | Restriction of Left External Carotid Artery with Extraluminal Device, Open Approach |
| 03VN0DZ | Restriction of Left External Carotid Artery with Intraluminal Device, Open Approach |
| 03VN0ZZ | Restriction of Left External Carotid Artery, Open Approach |
| 03VN3BZ | Restriction of Left External Carotid Artery with Bioactive Intraluminal Device, Percutaneous Approach |
| 03VN3CZ | Restriction of Left External Carotid Artery with Extraluminal Device, Percutaneous Approach |
| 03VN3DZ | Restriction of Left External Carotid Artery with Intraluminal Device, Percutaneous Approach |
| 03VN3ZZ | Restriction of Left External Carotid Artery, Percutaneous Approach |
| 03VN4BZ | Restriction of Left External Carotid Artery with Bioactive Intraluminal Device, Percutaneous Endoscopic Approach |
| 03VN4CZ | Restriction of Left External Carotid Artery with Extraluminal Device, Percutaneous Endoscopic Approach |
| 03VN4DZ | Restriction of Left External Carotid Artery with Intraluminal Device, Percutaneous Endoscopic Approach |
| 03VN4ZZ | Restriction of Left External Carotid Artery, Percutaneous Endoscopic Approach |
| 03VP0BZ | Restriction of Right Vertebral Artery with Bioactive Intraluminal Device, Open Approach |
| 03VP0CZ | Restriction of Right Vertebral Artery with Extraluminal Device, Open Approach |
| 03VP0DZ | Restriction of Right Vertebral Artery with Intraluminal Device, Open Approach |
| 03VP0ZZ | Restriction of Right Vertebral Artery, Open Approach |
| 03VP3BZ | Restriction of Right Vertebral Artery with Bioactive Intraluminal Device, Percutaneous Approach |
| 03VP3CZ | Restriction of Right Vertebral Artery with Extraluminal Device, Percutaneous Approach |
| 03VP3DZ | Restriction of Right Vertebral Artery with Intraluminal Device, Percutaneous Approach |
| 03VP3ZZ | Restriction of Right Vertebral Artery, Percutaneous Approach |
| 03VP4BZ | Restriction of Right Vertebral Artery with Bioactive Intraluminal Device, Percutaneous Endoscopic Approach |
| 03VP4CZ | Restriction of Right Vertebral Artery with Extraluminal Device, Percutaneous Endoscopic Approach |
| 03VP4DZ | Restriction of Right Vertebral Artery with Intraluminal Device, Percutaneous Endoscopic Approach |
| 03VP4ZZ | Restriction of Right Vertebral Artery, Percutaneous Endoscopic Approach |
| 03VQ0BZ | Restriction of Left Vertebral Artery with Bioactive Intraluminal Device, Open Approach |
| 03VQ0CZ | Restriction of Left Vertebral Artery with Extraluminal Device, Open Approach |
| 03VQ0DZ | Restriction of Left Vertebral Artery with Intraluminal Device, Open Approach |
| 03VQ0ZZ | Restriction of Left Vertebral Artery, Open Approach |
| 03VQ3BZ | Restriction of Left Vertebral Artery with Bioactive Intraluminal Device, Percutaneous Approach |
| 03VQ3CZ | Restriction of Left Vertebral Artery with Extraluminal Device, Percutaneous Approach |
| 03VQ3DZ | Restriction of Left Vertebral Artery with Intraluminal Device, Percutaneous Approach |
| 03VQ3ZZ | Restriction of Left Vertebral Artery, Percutaneous Approach |
| 03VQ4BZ | Restriction of Left Vertebral Artery with Bioactive Intraluminal Device, Percutaneous Endoscopic Approach |
| 03VQ4CZ | Restriction of Left Vertebral Artery with Extraluminal Device, Percutaneous Endoscopic Approach |
| 03VQ4DZ | Restriction of Left Vertebral Artery with Intraluminal Device, Percutaneous Endoscopic Approach |
| 03VQ4ZZ | Restriction of Left Vertebral Artery, Percutaneous Endoscopic Approach |

**Valve Procedure**

**Open Valve replacement**

| **Code** | **Description** |
| --- | --- |
| **Mitral** |  |
| 35.24 | Open and other replacement of mitral valve |
| 02RG0JZ | Replacement of Mitral Valve with Synth Sub, Open Approach |
| 02RG3JZ | Replacement of Mitral Valve with Synth Sub, Perc Approach |
| 02RG4JZ | Replace of Mitral Valve with Synth Sub, Perc Endo Approach |
| 35.23 | Open and other replacement of mitral valve with tissue graft |
| 02RG07Z | Replacement of Mitral Valve with Autol Sub, Open Approach |
| 02RG08Z | Replacement of Mitral Valve with Zooplastic, Open Approach |
| 02RG0KZ | Replacement of Mitral Valve with Nonaut Sub, Open Approach |
| 33430 | Replacement, mitral valve, with cardiopulmonary bypass |
| **Aortic** |  |
| 33405 | Replacement, aortic valve, with cardiopulmonary bypass; with prosthetic valve other than homograft or stentless valve |
| 33406 | Replacement, aortic valve, with cardiopulmonary bypass; with allograft valve (freehand) |
| 33410 | Replacement, aortic valve, with cardiopulmonary bypass; with stentless tissue valve |
| 33411 | Replacement, aortic valve; with aortic annulus enlargement, noncoronary sinus |
| 33412 | Replacement, aortic valve; with transventricular aortic annulus enlargement (Konno procedure) |
| 33413 | Replacement, aortic valve; by translocation of autologous pulmonary valve with allograft replacement of pulmonary valve (Ross procedure) |
| 35.21 | Open and other replacement of aortic valve with tissue graft |
| 02RF07Z | Replacement of Aortic Valve with Autol Sub, Open Approach |
| 02RF08Z | Replacement of Aortic Valve with Zooplastic, Open Approach |
| 02RF0KZ | Replacement of Aortic Valve with Nonaut Sub, Open Approach |
| 35.22 | Open and other replacement of aortic valve |
| 02RF0JZ | Replacement of Aortic Valve with Synth Sub, Open Approach |
| 02RF4JZ | Replace of Aortic Valve with Synth Sub, Perc Endo Approach |
| **Other** |  |
| 33465 | Replacement, tricuspid valve, with cardiopulmonary bypass |
| 33475 | Replacement, pulmonary valve |
| 35.25 | Open and other replacement of pulmonary valve with tissue graft |
| 02RH07Z | Replacement of Pulmonary Valve with Autol Sub, Open Approach |
| 02RH08Z | Replacement of Pulm Valve with Zooplastic, Open Approach |
| 02RH0KZ | Replacement of Pulm Valve with Nonaut Sub, Open Approach |
| 35.27 | Open and other replacement of tricuspid valve with tissue graft |
| 02RJ07Z | Replacement of Tricuspid Valve with Autol Sub, Open Approach |
| 02RJ08Z | Replacement of Tricusp Valve with Zooplastic, Open Approach |
| 02RJ0KZ | Replacement of Tricuspid Valve with Nonautologous Tissue Substitute, Open Approach |
| 35.26 | Open and other replacement of pulmonary valve |
| 02RH0JZ | Replacement of Pulmonary Valve with Synth Sub, Open Approach |
| 02RH4JZ | Replacement of Pulm Valve with Synth Sub, Perc Endo Approach |
| 35.28 | Open and other replacement of tricuspid valve |
| 02RJ0JZ | Replacement of Tricuspid Valve with Synth Sub, Open Approach |
| 02RJ4JZ | Replace of Tricusp Valve with Synth Sub, Perc Endo Approach |
| **Indicating valve replacement** | |
| 33496 | Repair of non-structural prosthetic valve dysfunction with cardiopulmonary bypass (separate procedure) |
| **ICD9 dx** |  |
| 996.02 | Mechanical complication due to heart valve prosthesis |
| 996.61 | Infection and inflammatory reaction due to cardiac device, implant, and graft |
| V43.3 | Heart valve replaced by other means |
| V42.2 | Heart valve replaced by transplant |
| 996.71 | Other complications due to heart valve prosthesis |
| **ICD10 dx** |  |
| T82.0x | Mechanical complication of heart valve prosthesis |
| T82.22x | Mechanical complication of biological heart valve graft |
| T82.6x | Infection and inflammatory reaction due to cardiac valve prosthesis |
| Z95.2 | Presence of prosthetic heart valve |
| Z95.3 | Presence of xenogenic heart valve |

**TAVR**

| 33361 | Transcatheter aortic valve replacement (TAVR/TAVI) with prosthetic valve; percutaneous femoral artery approach |
| --- | --- |
| 33362 | Transcatheter aortic valve replacement (TAVR/TAVI) with prosthetic valve; open femoral artery approach |
| 33363 | Transcatheter aortic valve replacement (TAVR/TAVI) with prosthetic valve; open axillary artery approach |
| 33364 | Transcatheter aortic valve replacement (TAVR/TAVI) with prosthetic valve; open iliac artery approach |
| 33365 | Transcatheter aortic valve replacement (TAVR/TAVI) with prosthetic valve; transaortic approach (eg, median sternotomy, mediastinotomy) |
| 33366 | Transcatheter aortic valve replacement (TAVR/TAVI) with prosthetic valve; transapical exposure (eg, left thoracotomy) |
| 33367 | Transcatheter aortic valve replacement (TAVR/TAVI) with prosthetic valve; cardiopulmonary bypass support with percutaneous peripheral arterial and venous cannulation (eg, femoral vessels) (List separately in addition to code for primary procedure) |
| 33368 | Transcatheter aortic valve replacement (TAVR/TAVI) with prosthetic valve; cardiopulmonary bypass support with open peripheral arterial and venous cannulation (eg, femoral, iliac, axillary vessels) (List separately in addition to code for primary procedure) |
| 33369 | Transcatheter aortic valve replacement (TAVR/TAVI) with prosthetic valve; cardiopulmonary bypass support with central arterial and venous cannulation (eg, aorta, right atrium, pulmonary artery) (List separately in addition to code for primary procedure) |
| 0257T | Implantation of catheter-delivered prosthetic aortic heart valve; open thoracic approach (eg, transapical, transventricular) |
| 0258T | Transthoracic cardiac exposure (eg, sternotomy, thoracotomy, subxiphoid) for catheter-delivered aortic valve replacement; without cardiopulmonary bypass |
| 0259T | Transthoracic cardiac exposure (eg, sternotomy, thoracotomy, subxiphoid) for catheter-delivered aortic valve replacement; with cardiopulmonary bypass |
| 0262T | implantation of catheter-delivered prosthetic pulmonary valve, endovascular approach |
| 0318T | Implantation of catheter-delivered prosthetic aortic heart valve, open thoracic approach, (eg, transapical, other than transaortic) |
| 35.05 | Endovascular replacement of aortic valve |
| 02RF37Z | Replacement of Aortic Valve with Autol Sub, Perc Approach |
| 02RF38Z | Replacement of Aortic Valve with Zooplastic, Perc Approach |
| 02RF3JZ | Replacement of Aortic Valve with Synth Sub, Perc Approach |
| 02RF3KZ | Replacement of Aortic Valve with Nonaut Sub, Perc Approach |
| X2RF332 | Replace Aort Valve w Zoopl Rapid Depl, Perc, New Tech 2 |
| 02RF47Z | Replace of Aortic Valve with Autol Sub, Perc Endo Approach |
| 02RF48Z | Replace of Aortic Valve with Zooplastic, Perc Endo Approach |
| 02RF4KZ | Replacement of Aortic Valve with Nonautologous Tissue Substitute, Percutaneous Endoscopic Approach |
| 35.06 | Transapical replacement of aortic valve |
| 02RF37H | Replace Aortic Valve w Autol Sub, Transap, Perc |
| 02RF38H | Replace Aortic Valve w Zooplastic, Transap, Perc |
| 02RF3JH | Replace Aortic Valve w Synth Sub, Transap, Perc |
| 02RF3KH | Replace Aortic Valve w Nonaut Sub, Transap, Perc |
| 35.07 | Endovascular replacement of pulmonary valve |
| 02RH37Z | Replacement of Pulmonary Valve with Autol Sub, Perc Approach |
| 02RH38Z | Replacement of Pulm Valve with Zooplastic, Perc Approach |
| 02RH3JZ | Replacement of Pulmonary Valve with Synth Sub, Perc Approach |
| 02RH3KZ | Replacement of Pulm Valve with Nonaut Sub, Perc Approach |
| 35.08 | Transapical replacement of pulmonary valve |
| 02RH37H | Replace of Pulm Valve with Autol Sub, Transap, Perc Approach |
| 02RH38H | Replace Pulm Valve w Zooplastic, Transap, Perc |
| 02RH3JH | Replace of Pulm Valve with Synth Sub, Transap, Perc Approach |
| 02RH3KH | Replace Pulm Valve w Nonaut Sub, Transap, Perc |
| 02RH47Z | Replacement of Pulm Valve with Autol Sub, Perc Endo Approach |
| 02RH48Z | Replace of Pulm Valve with Zooplastic, Perc Endo Approach |
| 02RH4KZ | Replace of Pulm Valve with Nonaut Sub, Perc Endo Approach |
| 35.09 | Endovascular replacement of unspecified heart valve |
| 02RG37H | Replace Mitral Valve w Autol Sub, Transap, Perc |
| 02RG37Z | Replacement of Mitral Valve with Autologous Tissue Substitute, Percutaneous Approach |
| 02RG38H | Replace Mitral Valve w Zooplastic, Transap, Perc |
| 02RG38Z | Replacement of Mitral Valve with Zooplastic Tissue, Percutaneous Approach |
| 02RG3JH | Replace Mitral Valve w Synth Sub, Transap, Perc |
| 02RG3KH | Replace Mitral Valve w Nonaut Sub, Transap, Perc |
| 02RG3KZ | Replacement of Mitral Valve with Nonaut Sub, Perc Approach |
| 02RJ47Z | Replace of Tricusp Valve with Autol Sub, Perc Endo Approach |
| 02RJ48Z | Replace of Tricusp Valve with Zooplastic, Perc Endo Approach |
| 02RJ4KZ | Replace of Tricusp Valve with Nonaut Sub, Perc Endo Approach |
| 02RG4KZ | Replace of Mitral Valve with Nonaut Sub, Perc Endo Approach |
| 02RG47Z | Replacement of Mitral Valve with Autologous Tissue Substitute, Percutaneous Endoscopic Approach |
| 02RG48Z | Replacement of Mitral Valve with Zooplastic Tissue, Percutaneous Endoscopic Approach |
| 33418 | Transcatheter mitral valve repair, percutaneous approach, including transsepptal puncture when performed; initial prosthesis |
| 33419 | Transcatheter mitral valve repair, percutaneous approach, including transsepptal puncture when performed; additional prosthesis(es) during same session (List separately in additional to code for primary procedure) |

**Open heart repair**

| **Code** | **Description** |
| --- | --- |
| **Mitral valve** |  |
| 33422 | Valvotomy, mitral valve; open heart, with cardiopulmonary bypass |
| 33425 | Valvuloplasty, mitral valve, with cardiopulmonary bypass; |
| 33426 | Valvuloplasty, mitral valve, with cardiopulmonary bypass; with prosthetic ring |
| 33427 | Valvuloplasty, mitral valve, with cardiopulmonary bypass; radical reconstruction, with or without ring |
| 35.12 | Open heart valvuloplasty of mitral valve without replacement |
| 027G04Z | Dilate Mitral Valve w Drug-elut Intralum, Open |
| 027G0DZ | Dilation of Mitral Valve with Intralum Dev, Open Approach |
| 027G0ZZ | Dilation of Mitral Valve, Open Approach |
| 02NG0ZZ | Release Mitral Valve, Open Approach |
| 02QG0ZZ | Repair Mitral Valve, Open Approach |
| 02QG3ZZ | Repair Mitral Valve, Percutaneous Approach |
| 02QG4ZZ | Repair Mitral Valve, Percutaneous Endoscopic Approach |
| 02CG0ZZ | Extirpation of Matter from Mitral Valve, Open Approach |
| 02UG0JZ | Supplement Mitral Valve with Synthetic Substitute, Open Approach |
| **Aortic valve** |  |
| 35.11 | Open heart valvuloplasty of aortic valve without replacement |
| 027F0ZZ | Dilation of Aortic Valve, Open Approach |
| 02NF0ZZ | Release Aortic Valve, Open Approach |
| 02QF0ZZ | Repair Aortic Valve, Open Approach |
| 02UF0JZ | Supplement Aortic Valve with Synthetic Substitute, Open Approach |
| 33400 | Valvuloplasty, aortic valve; open, with cardiopulmonary bypass |
| 33401 | Valvuloplasty, aortic valve; open, with inflow occlusion |
| 33403 | Valvuloplasty, aortic valve; using transventricular dilation, with cardiopulmonary bypass |
| 33864 | Ascending aorta graft, with cardiopulmonary bypass with valve suspension, with coronary reconstruction and valve-sparing aortic root remodeling (eg, David Procedure, Yacoub Procedure) |
| **Pulmonary or tricuspid valve, or unspecified** | |
| 35.13 | Open heart valvuloplasty of pulmonary valve without replacement |
| 027H0ZZ | Dilation of Pulmonary Valve, Open Approach |
| 02NH0ZZ | Release Pulmonary Valve, Open Approach |
| 02QH0ZZ | Repair Pulmonary Valve, Open Approach |
| 35.14 | Open heart valvuloplasty of tricuspid valve without replacement |
| 027J0ZZ | Dilation of Tricuspid Valve, Open Approach |
| 02NJ0ZZ | Release Tricuspid Valve, Open Approach |
| 02QJ0ZZ | Repair Tricuspid Valve, Open Approach |
| 35.33 | Annuloplasty |
| 02UH0JZ | Supplement Pulmonary Valve with Synthetic Substitute, Open Approach |
| 02UJ0JZ | Supplement Tricuspid Valve with Synthetic Substitute, Open Approach |
| 02R94KZ | Replace Chordae Tendineae w Nonaut Sub, Perc Endo |
| 33460 | Valvectomy, tricuspid valve, with cardiopulmonary bypass |
| 33463 | Valvuloplasty, tricuspid valve; without ring insertion |
| 33464 | Valvuloplasty, tricuspid valve; with ring insertion |
| 33474 | Valvotomy, pulmonary valve, open heart, with cardiopulmonary bypass |
| 33417 | Aortoplasty (gusset) for supravalvular stenosis |
| 33468 | Tricuspid valve repositioning and plication for Ebstein anomaly |
| 33496 | Repair of non-structural prosthetic valve dysfunction with cardiopulmonary bypass (separate procedure) |
| 33600 | Closure of atrioventricular valve (mitral or tricuspid) by suture or patch |
| 33602 | Closure of semilunar valve (aortic or pulmonary) by suture or patch |

**Percutaneous heart repair**

| **Code** | **Description** |
| --- | --- |
| **Mitral valve** |  |
| 33420 | Valvotomy, mitral valve; closed heart |
| 92987 | Percutaneous balloon valvuloplasty; mitral valve |
| 35.02 | Closed heart valvotomy, mitral valve |
| 02NG3ZZ | Release Mitral Valve, Percutaneous Approach |
| 02CG3ZZ | Extirpation of Matter from Mitral Valve, Perc Approach |
| 02CG4ZZ | Extirpation of Matter from Mitral Valve, Perc Endo Approach |
| 02NG4ZZ | Release Mitral Valve, Percutaneous Endoscopic Approach |
| 35.97 | Percutaneous mitral valve repair with implant |
| 02UG3JZ | Supplement Mitral Valve with Synthetic Substitute, Percutaneous Approach |
| 027G3ZZ | Dilation of Mitral Valve, Percutaneous Approach |
| **Aortic valve** |  |
| 35.01 | Closed heart valvotomy, aortic valve |
| 02NF4ZZ | Release Aortic Valve, Percutaneous Endoscopic Approach |
| 02CF4ZZ | Extirpation of Matter from Aortic Valve, Perc Endo Approach |
| 02CF3ZZ | Extirpation of Matter from Aortic Valve, Perc Approach |
| 02NF3ZZ | Release Aortic Valve, Percutaneous Approach |
| 027F3ZZ | Dilation of Aortic Valve, Percutaneous Approach |
| **Pulmonary or tricuspid valve** | |
| 35.00 | Closed heart valvotomy, unspecified valve |
| 35.03 | Closed heart valvotomy, pulmonary valve |
| 02CH3ZZ | Extirpation of Matter from Pulmonary Valve, Perc Approach |
| 02NH3ZZ | Release Pulmonary Valve, Percutaneous Approach |
| 02NH4ZZ | Release Pulmonary Valve, Percutaneous Endoscopic Approach |
| 02CH4ZZ | Extirpation of Matter from Pulm Valve, Perc Endo Approach |
| 35.04 | Closed heart valvotomy, tricuspid valve |
| 02CJ3ZZ | Extirpation of Matter from Tricuspid Valve, Perc Approach |
| 02CJ4ZZ | Extirpation of Matter from Tricusp Valve, Perc Endo Approach |
| 02NJ3ZZ | Release Tricuspid Valve, Percutaneous Approach |
| 02NJ4ZZ | Release Tricuspid Valve, Percutaneous Endoscopic Approach |
| 33470 | Valvotomy, pulmonary valve, closed heart; transventricular |
| 33471 | Valvotomy, pulmonary valve, closed heart; via pulmonary artery |
| 33477 | Transcatheter pulmonary valve implantation, percutaneous approach, including pre-stenting of the valve delivery site, when performed |
| 35.96 | Percutaneous balloon valvuloplasty |
| 027H3ZZ | Dilation of Pulmonary Valve, Percutaneous Approach |
| 027J3ZZ | Dilation of Tricuspid Valve, Percutaneous Approach |

**Cardiovascular Procedure**

| **Code** | **Description** |
| --- | --- |
| 37.34 | atrial fibrillation ablation |
| 0210093 | CABG |
| 0210099 | CABG |
| 021009W | CABG |
| 02100A8 | CABG |
| 02100A9 | CABG |
| 02100AW | CABG |
| 02100KW | CABG |
| 02100Z3 | CABG |
| 02100Z9 | CABG |
| 0211093 | CABG |
| 0211099 | CABG |
| 021109W | CABG |
| 02110AW | CABG |
| 02110Z9 | CABG |
| 02111ZZ | CABG |
| 0212099 | CABG |
| 021209W | CABG |
| 02120A3 | CABG |
| 0213093 | CABG |
| 021309W | CABG |
| 02132 | CABG |
| 0213T | CABG |
| 36.1 | CABG |
| 36.10 | CABG |
| 36.11 | CABG |
| 36.12 | CABG |
| 36.13 | CABG |
| 36.14 | CABG |
| 36.15 | CABG |
| 36.16 | CABG |
| 36.19 | CABG |
| 36.22 | CABG |
| 36.23 | CABG |
| 01925 | Carotid revascularization |
| 031H0KK | Carotid revascularization |
| 031J0KK | Carotid revascularization |
| 031J0ZK | Carotid revascularization |
| 037K3DZ | Carotid revascularization |
| 33889 | Carotid revascularization |
| 33891 | Carotid revascularization |
| 34001 | Carotid revascularization |
| 35301 | Carotid revascularization |
| 35390 | Carotid revascularization |
| 35509 | Carotid revascularization |
| 35601 | Carotid revascularization |
| 35606 | Carotid revascularization |
| 35626 | Carotid revascularization |
| 35693 | Carotid revascularization |
| 35694 | Carotid revascularization |
| 35695 | Carotid revascularization |
| 37205 | Carotid revascularization |
| 37206 | Carotid revascularization |
| 37207 | Carotid revascularization |
| 37208 | Carotid revascularization |
| 37215 | Carotid revascularization |
| 37216 | Carotid revascularization |
| 37236 | Carotid revascularization |
| 37237 | Carotid revascularization |
| 37600 | Carotid revascularization |
| 61613 | Carotid revascularization |
| 61700 | Carotid revascularization |
| 61703 | Carotid revascularization |
| 61705 | Carotid revascularization |
| 61710 | Carotid revascularization |
| G8348 | Carotid revascularization |
| 025T3ZZ | Destruction of Left Pulmonary Vein, Percutaneous Approach |
| 025S3ZZ | Destruction of Right Pulmonary Vein, Percutaneous Approach |
| 38.12 | Endarterectomy, other vessels of head and neck |
| 88.57 | Other and unspecified coronary arteriography |
| 0270346 | PCI |
| 027034Z | PCI |
| 0270356 | PCI |
| 027035Z | PCI |
| 027036Z | PCI |
| 027037Z | PCI |
| 02703DZ | PCI |
| 02703EZ | PCI |
| 02703GZ | PCI |
| 02703Z6 | PCI |
| 02703ZZ | PCI |
| 027044Z | PCI |
| 02704ZZ | PCI |
| 0271346 | PCI |
| 027134Z | PCI |
| 027135Z | PCI |
| 027136Z | PCI |
| 027137Z | PCI |
| 02713DZ | PCI |
| 02713ZZ | PCI |
| 027234Z | PCI |
| 027236Z | PCI |
| 0272376 | PCI |
| 027334Z | PCI |
| 02733ZZ | PCI |
| 36.0 | PCI |
| 36.01 | PCI |
| 36.02 | PCI |
| 36.03 | PCI |
| 36.04 | PCI |
| 36.05 | PCI |
| 36.06 | PCI |
| 36.07 | PCI |
| 36.09 | PCI |
| 00.63 | Percutaneous insertion of carotid artery stent |
| 00.66 | PTCA |
| 35.52 | Repair of atrial septal defect with prosthesis, closed technique |
| 93580 | Under Repair Procedures of Structural Heart Defect |

#### **Supplement A2. ICD-9 and ICD-10 codes and medications to define dementia.**

**ICD codes used to define dementia**

| **ICD-9 codes and Descriptions** | |
| --- | --- |
| 290.0 | Senile dementia, uncomplicated |
| 290.10 | Presenile dementia, uncomplicated |
| 290.11 | Presenile dementia with delirium |
| 290.12 | Presenile dementia with delusional features |
| 290.13 | Presenile dementia with depressive features |
| 290.20 | Senile dementia with delusional features |
| 290.21 | Senile dementia with depressive features |
| 290.3 | Senile dementia with delirium |
| 290.40 | Vascular dementia, uncomplicated |
| 290.41 | Vascular dementia, with delirium |
| 290.42 | Vascular dementia, with delusions |
| 290.43 | Vascular dementia, with depressed mood |
| 294.20 | Dementia, unspecified, without behavioral disturbance |
| 331.0 | Alzheimer's disease |
| **ICD-10 Codes and Descriptions** | |
| F03.90 | Unspecified dementia without behavioral disturbance |
| F01.50 | Vascular dementia without behavioral disturbance |
| F01.51 | Vascular dementia with behavioral disturbance |
| G30.0 | Alzheimer's disease with early onset |
| G30.1 | Alzheimer's disease with late onset |
| G30.8 | Other Alzheimer's disease |
| G30.9 | Alzheimer's disease, unspecified |

**Cognitive enhancement medications to define dementia**

| **Generic** | **Brand** |
| --- | --- |
| Donepezil | Aricept |
| Galantamine | Razadyne |
| Memantine | Namenda |
| Rivastigmine | Exelon |
| Memantine + Donepezil | Namzaric |

#### **Supplement A3. Development of the natural language processing algorithm to classify patients into different WMD severity categories.**

The NLP algorithms for extracting WMD grading and CBI location were developed based on our prior system under the ESPRESSO project (1). The infrastructure of the NLP system was an open-source NLP pipeline MedTaggerIE (2), which was developed using an open-source unstructured information management architecture–based information extraction framework (3). The pipeline separates task-specific NLP knowledge engineering (i.e., CBI location) from the generic routine NLP, which enables words and phrases containing clinical information to be directly defined by subject matter experts (e.g., neurologists and radiologists). To enhance research reproducibility and transparency, we released the NLP system under the Open Health Natural Language Processing Consortium, which can be accessed through <https://github.com/OHNLP/MedTagger>.

The gold standard corpus for the evaluation of the NLP system was established through a standard corpus annotation process (4). Two trained radiology residents (AOR, KN) manually reviewed 932 radiology reports randomly stratified by the presence of CBIs and WMDs from the Mayo, TMC and Kaiser EHRs. The annotation was conducted using a previously validated annotation guideline and scheme (4, 5). The gold standard dataset was equally split into two subsets including 466 training and 466 test. We followed an iterative training and refining process (6) to develop the WMD grading and CBI location algorithms. Briefly, a prototype system was applied to the training data. Error cases were manually reviewed by a neurologist (LYL) at TMC. Keywords were manually curated through an iteratively refining process until all issues were resolved. The evaluation of the final system on the blinded test data yielded 0.953 for CBI location and 0.884 for WMD grading in f1-score, respectively. F1-score is a well-established metric in the information retrieval and machine learning community that measures both positive predictive value (precision) and sensitivity (recall) of the imbalanced test object (7). The detailed breakdown of each class can be found in Table 1 and Table 2. False cases on the test data were further refined and corrected prior to the final system deployment.

**Table 1.** 4 × 4 Contingency Table of CBI Location

|  |  | **NLP** | | | | |
| --- | --- | --- | --- | --- | --- | --- |
|  |  | **Lacunar Subcortical** | **Cortical Juxtacortical** | **Both** | **No mention** | **Total** |
| **Annotation** | **Lacunar Subcortical** | 40 | 0 | 0 | 4 | 44 |
|  | **Cortical Juxtacortical** | 2 | 8 | 0 | 7 | 17 |
|  | **Both** | 1 | 0 | 0 | 0 | 1 |
|  | **No mention** | 8 | 0 | 0 | 396 | 404 |
|  | **Total** | 51 | 8 | 0 | 407 | 466 |

**Table 2.** 4 x 4 Contingency Table of WMD Grading

|  |  | **NLP** | | | | |
| --- | --- | --- | --- | --- | --- | --- |
|  |  | **Mild** | **Moderate** | **Severe** | **No mention** | **Total** |
| **Annotation** | **Mild** | 265 | 3 | 3 | 14 | 285 |
|  | **Moderate** | 4 | 67 | 4 | 7 | 82 |
|  | **Severe** | 1 | 0 | 17 | 0 | 18 |
|  | **No mention** | 13 | 3 | 2 | 63 | 81 |
|  | **Total** | 283 | 73 | 26 | 84 | 466 |

7.	Van Rijsbergen CJ. The geometry of information retrieval: Cambridge University Press; 2004.

### **Supplement B: Supplementary results**

#### **eFigure 1. Cohort Diagram**

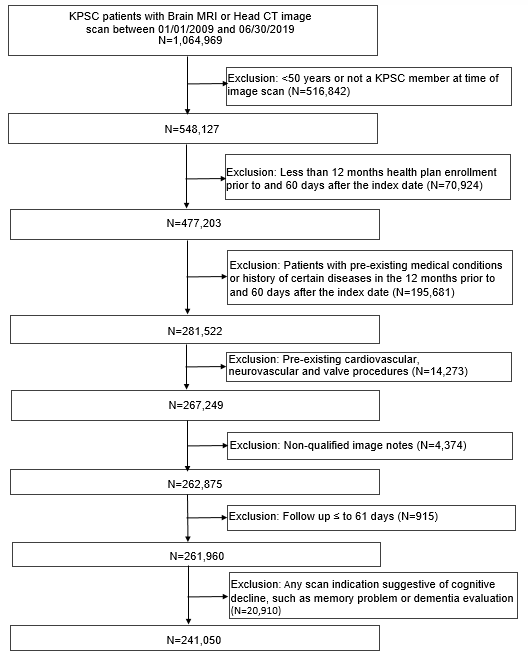

#### **eTable 1. Crude and adjusted effects on dementia risk for other risk factors in model**

|  | **HR** | **95% CI** |
| --- | --- | --- |
| ***Crude*** |  |  |
| Age group (reference <70 years) | 14.99 | (14.15, 15.69) |
| Sex (reference=Female) | 0.93 | (0.89, 0.97) |
| Race/ ethnicity (reference=Non-Hispanic) white) |  |  |
| Asian and Pacific Islander | 0.55 | (0.52, 0.59) |
| African American | 0.97 | (0.92, 1.02) |
| Hispanic | 0.61 | (0.58, 0.64) |
| Multiple/Other/Unknown | 0.46 | (0.38, 0.57) |
| Atrial fibrillation (AF) | 3.13 | (2.97, 3.30) |
| Carotid atherosclerosis | 2.56 | (2.26, 2.91) |
| Congestive heart failure (CHF) | 3.11 | (2.93, 3.30) |
| Coronary artery disease (CAD) | 2.37 | (2.27, 2.48) |
| Diabetes mellitus (DM) | 1.59 | (1.53, 1.65) |
| Hypercholesterolemia (HC) | 1.58 | (1.51, 1.64) |
| Hypertension (HTN) | 3.34 | (3.18, 3.51) |
| Peripheral arterial disease (PAD) | 2.96 | (2.79, 3.13) |
| Tobacco use (ever) | 1.27 | (1.22, 1.31) |
| Systolic blood pressure | 1.02 | (1.02, 1.02) |
| Statin | 1.67 | (1.61, 1.73) |
| Antiplatelet | 1.60 | (1.49, 1.72) |
| Depression | 1.28 | (1.22, 1.33) |
| Exercise (hour/per week) | 0.89 | (0.88, 0.90) |
| BMI (kg/m^2^) | 0.92 | (0.92, 0.92) |
| MRI (reference=CT) | 0.51 | (0.49, 0.54) |
| Covert brain infarction | 2.91 | (2.75, 3.07) |
| White matter disease | 3.72 | (3.58, 3.86) |
| ***Adjusted*** |  |  |
| Sex (reference=Female) | 1.07 | (1.03, 1.11) |
| race/ ethnicity (reference=Non-Hispanic) white) |  |  |
| Asian and Pacific Islander | 0.60 | (0.56, 0.65) |
| African American | 1.10 | (1.04, 1.16) |
| Hispanic | 0.86 | (0.82, 0.90) |
| Multiple/Other/Unknown | 0.88 | (0.71, 1.09) |
| Atrial fibrillation (AF) | 1.25 | (1.18, 1.32) |
| Carotid atherosclerosis | 1.01 | (0.89, 1.15) |
| Congestive heart failure (CHF) | 1.26 | (1.18, 1.35) |
| Coronary artery disease (CAD) | 1.10 | (1.05, 1.15) |
| Diabetes mellitus (DM) | 1.23 | (1.18, 1.29) |
| Hypercholesterolemia (HC) | 0.96 | (0.91, 1.01) |
| Hypertension (HTN) | 1.40 | (1.33, 1.48) |
| Peripheral arterial disease (PAD) | 1.24 | (1.16, 1.31) |
| Tobacco use (ever) | 0.99 | (0.95, 1.03) |
| Systolic blood pressure | 1.08 | (1.05, 1.11) |
| Statin | 1.01 | (0.96, 1.05) |
| Antiplatelet | 1.00 | (0.92, 1.07) |
| Depression | 1.41 | (1.36, 1.49) |
| Exercise (hour/per week) | 0.81 | (0.79, 0.83) |
| BMI (kg/m^2^) | 0.61 | (0.60, 0.63) |
| Covert brain infarction |  |  |
| Age<70 years | 2.02 | (1.70, 2.41) |
| Age≥70 years | 1.22 | (1.15, 1.30) |
| White matter disease |  |  |
| MRI, Age<70 | 2.28 | (1.99, 2.62) |
| MRI, Age≥70 | 1.48 | (1.32, 1.66) |
| CT, Age<70 | 2.87 | (2.58, 3.19) |
| CT, Age≥70 | 1.87 | (1.79, 1.95) |

#### **eTable 2. Dementia^a^ incidence rate defined by one ICD diagnose code, overall and in subgroups (n= 241,050)**

| **Patient Characteristics** | **Patients with covert brain infarction (CBI) (n=11,328)** | | | **Patient with white matter disease (WMD) (n=69,931)** | | | **Patients with both CBI and WMD disease (n=6,284)** | | | **Patients without CBI or WMD**  **(n=166,075)** | | |
| --- | --- | --- | --- | --- | --- | --- | --- | --- | --- | --- | --- | --- |
|  | **Avg f/u, yrs** | **No. events** | **Incidence rate**  **(95% CI)** | **Avg f/u, yrs** | **No. events** | **Incidence rate**  **(95% CI)** | **Avg f/u, yrs** | **No. events** | **Incidence rate**  **(95% CI)** | **Avg f/u, yrs** | **No. events** | **Incidence rate**  **(95% CI)** |
| All | 4.26 | 1,696 | 35.2 (33.5,36.9) | 4.06 | 7,937 | 27.9 (27.3,28.6) | 3.87 | 1,202 | 49.4 (46.7,52.3) | 4.45 | 5,411 | 7.3 (7.1,7.5) |
| Age group (yrs) |  |  |  |  |  |  |  |  |  |  |  |  |
| 50-59 | 4.61 | 31 | 3.9 (2.7,5.5) | 4.40 | 85 | 1.5 (1.2,1.8) | 4.40 | 12 | 5.2 (2.8,8.8) | 4.42 | 262 | 0.7 (0.7,0.8) |
| 60-69 | 4.67 | 149 | 10.7 (9.1,12.5) | 4.33 | 673 | 7.7 (7.1,8.3) | 4.28 | 91 | 15.5 (12.6,19.0) | 4.54 | 1,067 | 4.4 (4.2,4.7) |
| 70-79 | 4.47 | 589 | 35.8 (33.0,38.8) | 4.14 | 2,869 | 32.0 (30.9,33.2) | 4.14 | 397 | 42.9 (38.8,47.3) | 4.55 | 2,227 | 19.3 (18.5,20.1) |
| 80+ | 3.35 | 927 | 93.8 (87.9,100.0) | 3.29 | 4,310 | 87.3 (84.7,90.0) | 3.21 | 702 | 101.8 (94.4,109.5) | 3.83 | 1,855 | 59.7 (57.0,62.4) |
| Sex |  |  |  |  |  |  |  |  |  |  |  |  |
| Female | 4.46 | 1,022 | 35.5 (33.4,37.7) | 4.17 | 5,114 | 28.8 (28.0,29.6) | 4.05 | 719 | 50.0 (46.4,53.7) | 4.55 | 3,455 | 7.4 (7.2,7.7) |
| Male | 3.98 | 674 | 34.7 (32.1,37.4) | 3.90 | 2,823 | 26.6 (25.6,27.6) | 3.64 | 483 | 48.6 (44.4,53.1) | 4.29 | 1,956 | 7.1 (6.8,7.5) |
| Race/ethnicity |  |  |  |  |  |  |  |  |  |  |  |  |
| Non-Hisp. White | 4.17 | 890 | 39.3 (36.8,41.9) | 4.07 | 4,477 | 30.8 (29.9,31.7) | 3.82 | 667 | 54.1 (50.2,58.4) | 4.53 | 2,636 | 8.9 (8.6,9.3) |
| Asian/PI | 4.58 | 135 | 23.5 (19.8,27.7) | 4.26 | 648 | 19.0 (17.5,20.5) | 4.09 | 88 | 31.3 (25.3,38.4) | 4.69 | 462 | 5.0 (4.5,5.5) |
| African Am. | 4.36 | 304 | 42.5 (37.9,47.5) | 4.10 | 1,129 | 35.2 (33.2,37.3) | 3.86 | 206 | 59.0 (51.4,67.5) | 4.72 | 783 | 8.9 (8.3,9.5) |
| Hispanic | 4.26 | 355 | 29.1 (26.2,32.2) | 3.98 | 1,629 | 23.5 (22.4,24.6) | 3.95 | 235 | 42.7 (37.5,48.4) | 4.25 | 1,487 | 5.9 (5.6,6.2) |
| Other^b^ | 3.20 | 12 | 28.0 (15.3,47.4) | 3.30 | 54 | 16.5 (12.5,21.4) | 2.66 | 6 | 29.6 (10.9,64.5) | 3.54 | 43 | 4.1 (3.0,5.5) |
| Imaging Modality |  |  |  |  |  |  |  |  |  |  |  |  |
| CT | 4.22 | 1,494 | 39.1 (37.2,41.2) | 3.79 | 6,285 | 44.9 (43.8,46.0) | 3.73 | 1,029 | 61.4 (57.7,65.2) | 4.45 | 4,949 | 8.1 (7.9,8.3) |
| MRI | 4.39 | 202 | 20.1 (17.5,23.1) | 4.36 | 1,652 | 11.5 (10.9,12.0) | 4.23 | 173 | 22.8 (19.6,26.4) | 4.46 | 462 | 3.6 (3.3,3.9) |

PI: Pacific islander.

^a^ Dementia was defined by two ICD diagnostic codes or one diagnostic code and one medication dispensing.

^b^ Multiple, other or unknown race/ethnicity.

#### **eTable 3. Dementia^a^ incidence rate in patients without antithrombotic medications/indications^b^, overall and in subgroups (n= 187,533)**

| **Patient Characteristics** | **Patients with covert brain infarction (CBI) (n=7,241)** | | | **Patient with white matter disease (WMD) (n=48,977)** | | | **Patients with both CBI and WMD disease (n=3,757)** | | | **Patients without CBI or WMD**  **(n= 135,072)** | | |
| --- | --- | --- | --- | --- | --- | --- | --- | --- | --- | --- | --- | --- |
|  | **Avg f/u, yrs** | **No. events** | **Incidence rate**  **(95% CI)** | **Avg f/u, yrs** | **No. events** | **Incidence rate**  **(95% CI)** | **Avg f/u, yrs** | **No. events** | **Incidence rate**  **(95% CI)** | **Avg f/u, yrs** | **No. events** | **Incidence rate**  **(95% CI)** |
| All | 4.49 | 772 | 23.8 (22.1,25.5) | 4.22 | 3,804 | 18.4 (17.8,19.0) | 4.08 | 529 | 34.5 (31.7,37.6) | 4.48 | 2,803 | 4.6 (4.5,4.8) |
| Age group (yrs) |  |  |  |  |  |  |  |  |  |  |  |  |
| 50-59 | 4.60 | 19 | 2.9 (1.8,4.5) | 4.43 | 47 | 0.9 (0.7,1.2) | 4.39 | 8 | 4.4 (1.9,8.6) | 4.43 | 142 | 0.5 (0.4,0.5) |
| 60-69 | 4.78 | 76 | 7.4 (5.8,9.1) | 4.38 | 392 | 5.7 (5.2,6.3) | 4.34 | 48 | 11.3 (8.4,14.9) | 4.57 | 625 | 3.2 (3.0,3.5) |
| 70-79 | 4.64 | 283 | 27.8 (24.7,31.2) | 4.27 | 1,483 | 24.7 (23.4,25.9) | 4.23 | 183 | 33.5 (28.9,38.6) | 4.62 | 1,200 | 14.9 (14.0,15.7) |
| 80+ | 3.72 | 394 | 71.3 (64.6,78.7) | 3.51 | 1,882 | 68.8 (65.7,71.9) | 3.54 | 290 | 76.4 (67.9,85.5) | 4.04 | 836 | 45.7 (42.7,48.9) |
| Sex |  |  |  |  |  |  |  |  |  |  |  |  |
| Female | 4.67 | 499 | 24.7 (22.6,27.0) | 4.30 | 2,592 | 19.5 (18.8,20.3) | 4.24 | 340 | 35.9 (32.3,39.9) | 4.56 | 1,863 | 4.8 (4.6,5.0) |
| Male | 4.21 | 273 | 22.2 (19.7,25.0) | 4.09 | 1,212 | 16.4 (15.5,17.3) | 3.85 | 189 | 32.2 (27.9,37.1) | 4.35 | 940 | 4.3 (4.0,4.6) |
| Race/ethnicity |  |  |  |  |  |  |  |  |  |  |  |  |
| Non-Hisp. white | 4.49 | 376 | 25.5 (23.0,28.2) | 4.28 | 2,021 | 20.0 (19.1,20.8) | 4.11 | 286 | 38.0 (33.8,42.6) | 4.59 | 1,293 | 5.6 (5.3,5.9) |
| Asian/PI | 4.73 | 67 | 16.1 (12.6,20.3) | 4.35 | 371 | 13.7 (12.4,15.2) | 4.23 | 42 | 21.1 (15.4,28.2) | 4.72 | 261 | 3.3 (2.9,3.7) |
| African Am. | 4.64 | 141 | 30.6 (25.9,36.0) | 4.30 | 533 | 23.7 (21.8,25.8) | 4.12 | 90 | 42.7 (34.6,52.3) | 4.78 | 386 | 5.5 (5.0,6.1) |
| Hispanic | 4.37 | 179 | 20.7 (17.8,23.9) | 4.08 | 847 | 15.8 (14.8,16.9) | 4.03 | 106 | 29.8 (24.6,35.9) | 4.27 | 833 | 3.9 (3.6,4.1) |
| Other^c^ | 3.01 | 9 | 27.6 (12.6,52.5) | 3.26 | 32 | 12.0 (8.4,16.8) | 2.31 | 5 | 35.5 (11.5,82.9) | 3.51 | 30 | 3.3 (2.3,4.6) |
| Imaging Modality |  |  |  |  |  |  |  |  |  |  |  |  |
| CT | 4.49 | 669 | 26.6 (24.6,28.7) | 3.99 | 2,892 | 31.8 (30.7,33.0) | 3.95 | 441 | 44.7 (40.7,49.0) | 4.49 | 2,521 | 5.1 (4.9,5.3) |
| MRI | 4.48 | 103 | 14.1 (11.5,17.0) | 4.42 | 912 | 7.9 (7.4,8.4) | 4.33 | 88 | 16.1 (13.0,19.8) | 4.47 | 282 | 2.5 (2.2,2.8) |

PI: Pacific islander.

^a^ Dementia was defined by two ICD diagnostic codes or one diagnostic code and one medication dispensing.

^b^ Excluded patients diagnosed with (1) any history of atrial fibrillation, moderate to severe valvopathy, MI, CAD and CHF, (2) DVT/PE within one year, or (3) received oral anticoagulation medication or antiplatelets within one year.

^c^ Multiple, other or unknown race/ethnicity.

#### **eTable 4. Dementia^a^ incidence rate in patients whose follow-up was longer than 1 year, overall and in subgroups (n= 209,780)**

| **Patient characteristics** | **Patients with covert brain infarction (CBI) (n=9,705)** | | | **Patient with white matter disease (WMD) (n=59,794)** | | | **Patients with both CBI and WMD disease (n=5,281)** | | | **Patients without CBI or WMD**  **(n= 145,562)** | | |
| --- | --- | --- | --- | --- | --- | --- | --- | --- | --- | --- | --- | --- |
|  | **Avg f/u, yrs** | **No. events** | **Incidence rate**  **(95% CI)** | **Avg f/u, yrs** | **No. events** | **Incidence rate**  **(95% CI)** | **Avg f/u, yrs** | **No. events** | **Incidence rate**  **(95% CI)** | **Avg f/u, yrs** | **No. events** | **Incidence rate**  **(95% CI)** |
| All | 4.08 | 1,177 | 29.7 (28.0,31.4) | 3.86 | 5,510 | 23.9 (23.3,24.5) | 3.72 | 836 | 42.6 (39.8,45.5) | 4.19 | 3,930 | 6.4 (6.3,6.7) |
| Age group (yrs) |  |  |  |  |  |  |  |  |  |  |  |  |
| 50-59 | 4.53 | 22 | 3.3 (2.1,4.9) | 4.17 | 50 | 1.1 (0.8,1.4) | 4.42 | 10 | 5.2 (2.5,9.6) | 4.18 | 145 | 0.5 (0.4,0.6) |
| 60-69 | 4.44 | 98 | 8.4 (6.9,10.2) | 4.11 | 486 | 6.8 (6.2,7.4) | 4.09 | 57 | 11.9 (9.1,15.2) | 4.26 | 772 | 3.9 (3.6,4.1) |
| 70-79 | 4.26 | 414 | 30.3 (27.5,33.4) | 3.90 | 2,096 | 28.7 (27.5,29.9) | 3.96 | 275 | 36.4 (32.3,40.9) | 4.23 | 1,695 | 17.7 (16.9,18.6) |
| 80+ | 3.19 | 643 | 83.0 (76.8,89.6) | 3.15 | 2,878 | 74.8 (72.1,77.5) | 3.04 | 494 | 92.4 (84.5,100.8) | 3.62 | 1,318 | 52.6 (49.8,55.5) |
| Sex |  |  |  |  |  |  |  |  |  |  |  |  |
| Female | 4.24 | 731 | 30.6 (28.5,32.9) | 3.94 | 3,577 | 24.6 (23.8,25.5) | 3.88 | 512 | 43.6 (40.0,47.6) | 4.27 | 2,522 | 6.5 (6.3,6.8) |
| Male | 3.86 | 446 | 28.3 (25.8,31.0) | 3.72 | 1,933 | 22.6 (21.6,23.6) | 3.50 | 324 | 41.0 (36.7,45.6) | 4.04 | 1,408 | 6.3 (6.0,6.6) |
| Race/ethnicity |  |  |  |  |  |  |  |  |  |  |  |  |
| Non-Hisp. white | 4.02 | 626 | 33.7 (31.2,36.4) | 3.87 | 3,110 | 26.3 (25.4,27.3) | 3.67 | 474 | 47.8 (43.7,52.3) | 4.25 | 1,920 | 7.8 (7.5,8.2) |
| Asian/PI | 4.40 | 93 | 19.4 (15.8,23.7) | 3.96 | 453 | 16.2 (14.8,17.8) | 3.91 | 60 | 26.2 (20.2,33.5) | 4.35 | 342 | 4.4 (4.0,4.9) |
| African Am. | 4.20 | 205 | 34.6 (30.1,39.5) | 3.95 | 765 | 29.2 (27.2,31.3) | 3.77 | 137 | 48.4 (40.8,57.0) | 4.46 | 568 | 7.7 (7.1,8.4) |
| Hispanic | 4.04 | 244 | 24.3 (21.4,27.5) | 3.76 | 1,147 | 20.5 (19.3,21.7) | 3.76 | 161 | 36.1 (30.9,42.0) | 4.00 | 1,074 | 5.2 (4.9,5.5) |
| Other^b^ | 3.09 | 9 | 27.5 (12.6,52.1) | 3.32 | 35 | 13.8 (9.8,19.0) | 2.63 | 4 | 27.2 (7.4,69.6) | 3.47 | 26 | 3.2 (2.1,4.6) |
| Imaging Modality |  |  |  |  |  |  |  |  |  |  |  |  |
| CT | 4.04 | 1,033 | 33.0 (31.0,35.0) | 3.60 | 4,278 | 38.1 (37.0,39.3) | 3.56 | 708 | 52.7 (49.0,56.7) | 4.18 | 3,586 | 7.1 (6.9,7.4) |
| MRI | 4.25 | 144 | 17.4 (14.7,20.4) | 4.13 | 1,232 | 10.4 (9.8,11.0) | 4.10 | 128 | 20.6 (17.3,24.4) | 4.21 | 344 | 3.2 (2.9,3.6) |

PI: Pacific islander.

^a^ Dementia was defined by two ICD diagnostic codes or one diagnostic code and one medication dispensing.

^b^ Multiple, other or unknown race/ethnicity.

#### **eTable 5. Crude and adjusted hazard ratios (HR) for dementia by covert brain infarction (CBI) and white matter disease (WMD) for patients with follow up longer than 1 year (n=209,780)**

|  | **HR** | **95% CI** |
| --- | --- | --- |
| ***Crude*** |  |  |
| CBI | 2.83 | (2.66, 3.01) |
| WMD | 3.53 | (3.39, 3.68) |
| ***Adjusted (varied by age and modality)^a^*** | | |
| CBI |  |  |
| Age<70 years | 1.97 | (1.64, 2.38) |
| Age≥70 years | 1.22 | (1.14, 1.30) |
| WMD |  |  |
| MRI, age<70 | 2.27 | (1.96, 2.63) |
| MRI, age≥70 | 1.50 | (1.33, 1.69) |
| CT, age<70 | 2.71 | (2.42, 3.04) |
| CT, age≥70 | 1.79 | (1.71, 1.88) |

^a^ The multivariable model included age, sex, race/ethnicity, atrial fibrillation, carotid atherosclerosis, congestive heart failure, coronary artery disease, diabetes mellitus, hypercholesterolemia, hypertension, peripheral arterial disease, ever tobacco use, mean systolic blood pressure in one year, antiplatelet use, stain use, depression, exercise (hour/per week), BMI (kg/m^2^), modality, age*CBI, age*WMD, and modality*WMD. Crude and adjusted HRs of all other risk factors included in the model are not shown.

#### **eTable 6. Incidence rate and crude and adjusted hazard ratios (HR) of white matter disease (WMD) and risk of dementia by severity and modality (n=241,050)**

| Modality and WMD Severity | Number of patients | Average f/u, years | Number of events | Incidence rate/1,000 person years (95% CI) | Crude HR  (95% CI) | Adjusted HR^a^  (95% CI) |
| --- | --- | --- | --- | --- | --- | --- |
| MRI |  |  |  |  |  |  |
| none (ref) | 29,451 | 4.48 | 403 | 3.1 (2.8, 3.4) | 1.00 | 1.00 |
| mild WMD | 20,610 | 4.31 | 621 | 7.0 (6.5, 7.6) | 2.30 (2.03, 2.60) | 1.41 (1.25, 1.60) |
| moderate WMD | 5,568 | 4.41 | 368 | 15.0 (13.5, 16.6) | 4.91 (4.26, 5.66) | 2.03 (1.76, 2.34) |
| severe WMD | 1,768 | 4.72 | 179 | 21.5 (18.5, 24.8) | 6.98 (5.85, 8.32) | 2.32 (1.94, 2.76) |
| not specified | 5,082 | 4.48 | 222 | 9.7 (8.5, 11.1) | 3.19 (2.70, 3.75) | 1.67 (1.41, 1.96) |
| CT |  |  |  |  |  |  |
| none | 141,668 | 4.47 | 4,454 | 7.0 (6.8, 7.2) | 2.31 (2.08, 2.55) | 1.58 (1.42, 1.75) |
| mild | 20,162 | 3.98 | 2,304 | 28.7 (27.5, 29.9) | 9.51 (8.56, 10.57) | 2.62 (2.36, 2.92) |
| moderate | 3,724 | 3.46 | 791 | 61.4 (57.2, 65.7) | 20.59 (18.26, 23.21) | 4.16 (3.68, 4.70) |
| severe | 1,959 | 3.46 | 435 | 64.1 (58.3, 70.4) | 21.49 (18.77, 24.61) | 4.11 (3.58, 4.72) |
| not specified | 11,058 | 3.69 | 1,777 | 43.6 (41.6, 45.6) | 14.51 (13.02, 16.17) | 3.40 (3.04, 3.79) |

^a^ Adjusting for age group, sex, race/ethnicity, atrial fibrillation, carotid atherosclerosis, congestive heart failure, coronary artery disease, diabetes mellitus, hypercholesterolemia, hypertension, peripheral arterial disease, ever tobacco use, mean systolic blood pressure in one year, antiplatelet use, stain use, depression, exercise (hour/per week), BMI (kg/m^2^), and CBI.
